# Characterizing Population-level Changes in Human Behavior during the COVID-19 Pandemic in the United States

**DOI:** 10.1101/2024.12.20.24319446

**Authors:** Tamanna Urmi, Binod Pant, George Dewey, Alexi Quintana-Mathé, Iris Lang, James N. Druckman, Katherine Ognyanova, Matthew Baum, Roy H. Perlis, Christoph Riedl, David Lazer, Mauricio Santillana

## Abstract

The transmission of communicable diseases in human populations is known to be modulated by behavioral patterns. However, detailed characterizations of how population-level behaviors change over time during multiple disease outbreaks and spatial resolutions are still not widely available. We used data from 431,211 survey responses collected in the United States, between April 2020 and June 2022, to provide a description of how human behaviors fluctuated during the first two years of the COVID-19 pandemic. Our analysis suggests that at the national and state levels, people’s adherence to recommendations to avoid contact with others (a preventive behavior) was highest early in the pandemic but gradually—and linearly—decreased over time. Importantly, during periods of intense COVID-19 mortality, adherence to preventive behaviors increased—despite the overall temporal decrease. These spatial-temporal characterizations help improve our understanding of the bidirectional feedback loop between outbreak severity and human behavior. Our findings should benefit both computational modeling teams developing methodologies to predict the dynamics of future epidemics and policymakers designing strategies to mitigate the effects of future disease outbreaks.

## 1 Introduction

Changes in human behavior at the population level have been known to impact the spread of communicable diseases. It is well understood that frequent and close human contact, such as shaking hands, hugging, or attending crowded events, can increase the risk of transmission of respiratory diseases such as seasonal influenza or the common cold [1]. In contrast, regular hand washing and the use of hand sanitizers are also known to significantly reduce pathogen transmission [2]. In fact, Semmelweis’s work in the mid 1800s in Vienna, Austria, linking the absence of hand washing to high mortality in maternity wards began a revolution that eventually led to the enshrinement of hand washing as a preventive health behavior in Western societies [3]. Historically, behaviors such as the intermittent migration of people from the countryside to densely packed overpopulated cities contributed to the spread of plague in Europe and Asia from the Roman Empire to the 19^th^ century [4]. More recently, the rise in the adoption of preventive behaviors such as adhering to stay-at-home recommendations and/or wearing facial masks substantially slowed the spread of infections during the COVID-19 pandemic before vaccines or other treatments were widely available [5, 6, 7, 8, 9]. Thus, having a clear characterization of how populations behave and how they change their behaviors can in turn help better understand how communicable disease transmission may unfold during an epidemic outbreak.

Human behavior at the population level can change in response to the emergence of major crises such as wars, famines, or pandemic events. Changes in economic activity and weather patterns, as well as local beliefs and political identity, may trigger and/or modulate behavior changes across geographies. In the context of epidemiological outbreaks, people can change their behavior patterns to reduce their risk of infection and potential death in times of high disease transmission [10]. For example, during periods of increased mortality caused by the West African Ebola outbreaks of the mid-2010s, people changed their behavior by abandoning the cultural practice of touching the bodies of deceased relatives when they learned that the deadly disease could be transmitted by contact with bodily fluids from corpses [11, 12]. Similarly, during the COVID-19 pandemic, people reduced their visits to crowded places, such as music concerts, movie theaters, and museums, during times when the number of deaths attributable to COVID-19 was high, as will be shown in this study.

In the same way that human behavior changes will influence the temporal evolution of the spread of infection during an outbreak, awareness of the risks that an infection may bring to individuals (such as their potential death) may lead to changes in population-level human behavior, modulated perhaps by local societal practices and/or ideologies. This bidirectional feedback between human behavior patterns and epidemiological dynamics has not consistently been characterized across diseases and outbreaks, in part due to poor epidemiological surveillance and the limited availability (and reliability) of estimates of human behavior indicators.

Accurate and timely epidemiological surveillance is challenging [13]. It would be impractical, in terms of economic resources and people’s consent, to test everyone in a population to fully identify the number of infected individuals at all points in time. Instead, the epidemiological community generally estimates the severity of a disease outbreak using various *correlates* or *proxies* such as the number of reported infections, hospitalizations, reported deaths, and the amount of viral RNA in wastewater [14, 15, 16]. Although these proxies are imperfect for fully characterizing the spread of communicable diseases due to limiting factors such as testing availability, test accuracy, reporting delays, heterogeneous ascertainment rates, and under-reporting, they can, in some cases, provide us with meaningful estimates of the time evolution of the severity of an ongoing outbreak, especially when multiple proxies are used in conjunction with each other [13]. However, in the context of COVID-19, spatially and temporally heterogeneous access to tests, asymptomatic infections, and reporting delays, among other reasons, made it difficult to track the number of infections in multiple locations in a timely manner [17, 18, 19].

Characterizing human behavior (e.g., risk-averting or risk-exposing actions) at the population level in the context of disease transmission is also a challenge. Multiple novel data sources have been used to monitor changes in human behavior during an epidemic outbreak. For example, Nsoesie et al. [20] used aerial images to monitor the number of cars in hospital parking lots to estimate the population’s need for medical attention, thus providing an indicator of the incidence of respiratory viruses. Commercial airline traffic data has been used to estimate inter-regional human movement around the world, thus allowing the estimation of potential risks of importation of pathogens [21, 10, 22, 23]. Human mobility estimates obtained from aggregated and anonymized location-enabled mobile phones have also been used to estimate changes in human contact patterns that have sometimes been linked to spatial and temporal changes in disease transmission [24, 25, 26, 27, 28, 29]. Although existing survey data have been helpful in characterizing patterns of behavior in the population during the COVID-19 pandemic [30, 31, 32, 33], survey approaches are frequently limited due to low sample sizes and inconsistent (and frequently short) time periods of deployment. Additional efforts have also been made to infer human behavior through the Oxford Stringency Index [34, 35, 36, 37], an index that attempts to quantify the stringency of government mandates to mitigate disease transmission [38, 39]. However, as will be shown in this study, government mandates may not reflect how individuals in a community choose to behave.

Having incomplete and poor characterizations of the interplay (feedback) between changes in human behavior and the evolving dynamics of disease outbreaks frequently leads public health officials and decision-makers to design mitigation strategies in the face of imminent disease outbreaks, based on intuition rather than observed evidence. Furthermore, researchers designing and implementing mathematical models of infectious disease transmission to predict upcoming disease events often do not include this important feedback in their formulations, leading to discrepancies between model predictions and eventually observed epidemic trajectories [40, 41].

In this study, we used data from a large-scale national representative survey conducted in the United States during the COVID-19 pandemic to characterize temporal changes in *risk-averting* (for example, avoiding contact with others) and *risk-exposing* behaviors (e.g., going to visit a friend). We analyzed the temporal evolution of 15 behaviors at the state and national levels in the United States, aggregated from survey respondents, and evaluated how their temporal trends changed as COVID-19 mortality fluctuated over the first two years of the pandemic, at the state and national levels.

We hypothesized (1) that people’s choice to adopt *risk-averting* behaviors (e.g., avoiding contact with others) would be highest during the earlier months of the pandemic, when uncertainty about the biology and consequences of COVID-19 infections was highest, and that the adoption of protective behaviors would decrease over time, due perhaps to personal fatigue, the availability of successful treatments and vaccines, or the eventual perception of proportions of the population that their infection risk was low; (2) that people’s perception of risk would increase in times when COVID-19 mortality was high, and that such perception would trigger a higher proportion of people to adopt preventive or *risk-averting* behaviors (conversely, in times of low mortality, we hypothesized that people would be more prone to engage in *risk-exposing* behaviors); (3) that changes of behavior such as the increase of *risk-exposing* behaviors (*e.g.*, going to visit a friend) would eventually lead to increases in detected COVID-19 cases, hospitalization, and deaths; (4) and that population-level changes of behaviors due to hypotheses 1, 2, and 3 would vary across states based on differences in political leaning. Finally, we hypothesized (5) that state-level adherence to government-directed policies would demonstrate substantial variation even in states with similar levels of non-pharmaceutical intervention stringency as measured by the Oxford Stringency Index.

## 2 Results

The key results of our national and state-level analysis can be summarized as follows: first, people were more willing to adhere to protective behaviors during the earlier stages of the pandemic, and the proportion of people adhering to protective behaviors decreased *linearly* and systematically over time (hypothesis 1); second, we found that higher proportions of people engaged in behaviors such as social distancing and mask wearing during intense disease transmission periods, and during low disease transmission periods, higher proportions of people engaged in *risk-exposing* behaviors (hypothesis 2); third, we found that increases in the adoption of *risk-exposing* behaviors were followed (or synchronously accompanied) by increases in detected COVID-19 cases, hospitalization, and deaths (hypothesis 3). Fourth, we found substantial variation in the way the above-listed findings manifested at the state level (hypothesis 4). Specifically, we found that Democratic-leaning states had higher proportions of people adhering to preventive behaviors compared to Swing and Republican leaning states. Finally, we found that community adherence to government-led state-level non-pharmaceutical recommendations was substantially different even in states with similar levels of mandate stringency as recorded by the Oxford Stringency Index (hypothesis 5). We detail the quantitative aspects of all of these findings below.

### 2.1 Survey characteristics

We analyzed survey data from 431,211 responses to 19 survey waves of the COVID States Project (https://www.covidstates.org) collected between April 2020 and June 2022 from respondents in the United States. Each survey wave contained about 20,000 responses and lasted two to four weeks, providing both national and state-level samples. We focus on responses regarding risk-averting and risk-exposing behavior. The survey is weighted to be representative of the U.S. population, and separate state-level weights are used for the state-level analysis. Given the large sample sizes and the use of weights, we are confident that the trajectories of each behavior we observe are meaningful and representative of behavioral trends at the national level.

We use three different questions from the survey to obtain 15 different variables describing the levels of adherence to behaviors mitigating or exacerbating COVID-19 spread. The temporal resolution of the survey data was interpolated to ensure that we had monthly behavior estimates that would correspond to each data point from our COVID-19 severity observations (i.e., mortality, hospitalizations, and cases). Interpolation is used to generate monthly data from April 2020 to May 2022, using survey-collected behavior data from April 2020 to June 2022. Hence, the study period is from April 2020 to May 2022. See **Methods** for further details.

### 2.2 Protective behaviors were more prevalent when mortality was high and exhibited temporal linear decreases

We analyzed the national-level temporal trends of risk-averting and risk-exposure behaviors by decomposing them into a linear decay (or increase) component and an oscillatory component **(Fig. 1)**. This decomposition revealed several findings. First, participation in risk-averting behaviors was highest at the beginning of the pandemic (in April 2020), with about 70% of respondents reporting *washing hands frequently* and/or *avoiding contact with others*. In contrast, participation in risk-exposing behaviors, such as *visiting a friend*, was lowest in April 2020, with approximately only 8% of the individuals participating in these activities (hypothesis 1).

**Figure 1:**
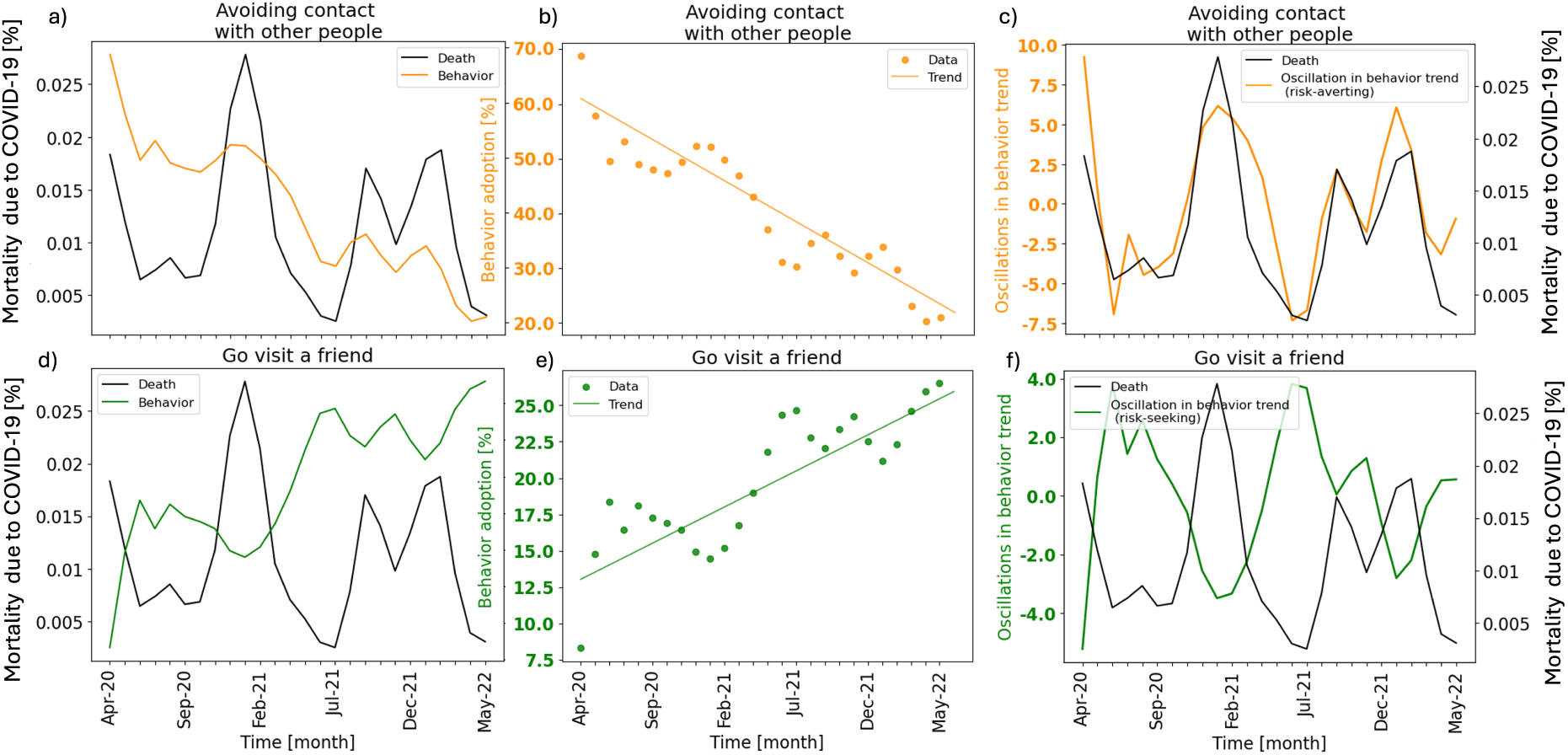
Decomposition of behavior trends into a linear decay component and an oscillatory component. **a, d**: The relationship between risk-averting behaviors (orange) or risk-exposing behaviors (green) and disease severity (mortality data, black) is not readily apparent. **b, e**: The linear decay components capture how the prevalence of risk-averting behaviors decreased over time, while the prevalence of risk-exposing behaviors increased over time. **c, f**: The oscillatory components are synchronized with trends in risk-averting behaviors and oppose the risk-exposing behavior trends.

Adherence to risk-averting behaviors decayed linearly over time (**Fig. 1b**), with a decrease of people *avoiding contact with others* of about 50 percentage points over two years, a three-fold decrease from roughly 70% of respondents social distancing in April 2020 to about 20% in May 2022. Conversely, people willing to participate in risk-exposing behaviors such as *going to visit a friend* increased linearly over time (**Fig. 1e**), more than tripling in the same two-year period from about 8% to 28% (hypothesis 1).

Second, the oscillatory components indicated that higher proportions of the population would practice social distancing (and other risk-averting behaviors) when COVID-19 transmission was most severe (as measured by real-time mortality indicators). Conversely, when COVID-19 severity was low, higher proportions of the population would engage in risk-exposing behaviors (**Fig. 1c**, **Fig. 1f**). We confirmed this using a lag correlation analysis (**Fig. 2**) that showed that the correlations between the oscillatory components of risk-averting behaviors and the reported deaths from COVID-19 were highest at lag 0 (synchronous) (hypothesis 2). The correlations between COVID-19 hospitalizations or cases and the behavior trends demonstrated a different pattern, with similarly high correlations between those two severity metrics and behavior at lag 0 and with behavior values shifted one month into the past. This suggests that behavior changes may influence future severity metrics (hypothesis 3).

**Figure 2:**
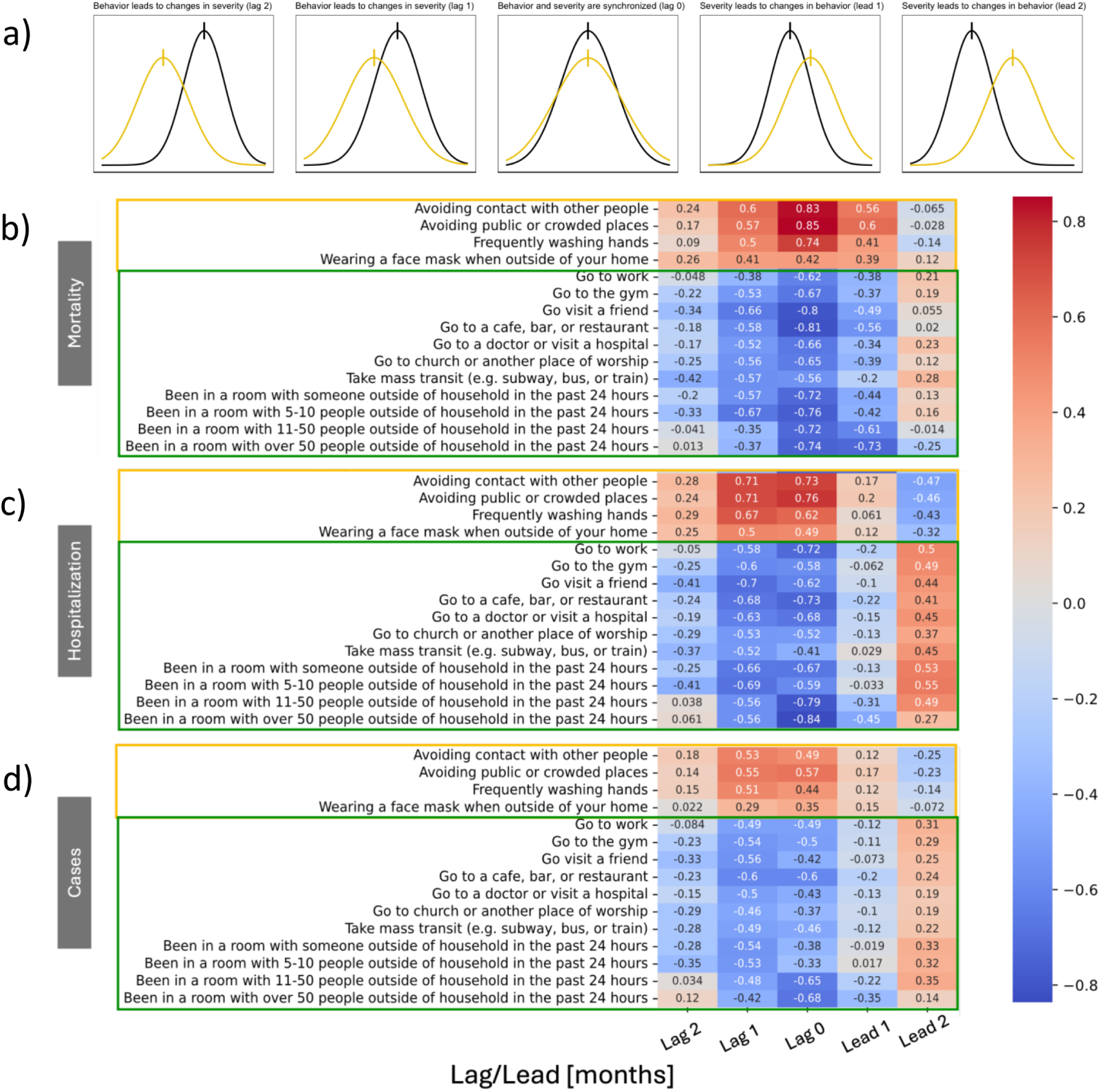
Correlations between oscillations in behavior trends and mortality are strongest at lag zero, while behavior trends anticipate cases and hospitalizations. Risk-averting behaviors (orange outline) are strongly positively correlated with mortality (**b**) at lag zero. Risk-exposing behaviors (green outline) are similarly strongly negatively correlated with mortality at lag zero. Correlations comparing current behavior to cases and hospitalizations in the future are stronger than current or past cases and hospitalizations. Red boxes indicate positive correlations, while blue boxes indicate negative correlations. Darker hues represent strong correlations, while lighter hues represent weaker correlations.

For clarity and communication purposes, **Fig. 1** shows the time evolution of only two behaviors: *Avoiding contact with other people* and *Go visit a friend*. In **Supplementary Fig. S1** we show the time evolution, decomposition, and synchronicity analysis for all behaviors at the national level. Visually, the qualitative results described in the previous paragraph hold for each risk-averting and risk-exposing behavior. To further quantify this finding, we analyzed the extent to which the time evolution of these behaviors was co-linear with each other over the two-year time period of this study using Principal Component Analysis (PCA). The PCA (See **Methods** for details) allowed us to identify that about 88% of the variance of all 15 behaviors could be captured by the first principal component (*eigen-behavior*). This finding suggests that survey participants respond to survey questions with consistency*, i.e.*, a respondent who chooses to avoid contact with others in times of high mortality, very likely chooses not to go to the gym and washes their hands frequently. This is also supported by the fact that, on average, the 15 behaviors considered in this study are highly correlated (see **Supplementary Fig. S13**). Mathematically speaking, the decomposition and synchronicity analyses conducted for the two behaviors in **Fig. 1** yield very similar results across behaviors as shown in **Supplementary Fig. S1** and Supplementary Materials.

### 2.3 Geographic and temporal differences of behavior trends and their relationship with mortality

Because population-level behavioral responses to the severity of outbreaks varied significantly across different geographic regions, we also performed decomposition and synchronicity analyses of behaviors at the state level (**Supplementary Fig. S1, Supplementary Table S1, Table 2**). We conducted two sets of state-level synchronicity analyses: 1) we compared state-level behavior trends to the same national-level severity measures used in the previous section and 2) we compared state-level behavior trends to state-level severity measures (*e.g.,* correlating behavior data from Massachusetts to the count of deaths over time in Massachusetts only). We used states as the unit of analysis in an ecological manner to reflect the jurisdictional nature of public health in the United States, as epidemiological data collection is handled by individual state health authorities and is predominantly operationalized for use only for individual states. We conducted these analyses only for states that had sample sizes that would yield meaningful insights over the two years of our study period. Specifically, we excluded 10 states/regions that had smaller sample sizes and relatively larger margins of error (see Supplementary Section 5 for details on the exclusion criteria). The regions that were not included in this analysis were Alaska, Wyoming, North Dakota, South Dakota, New Mexico, Vermont, Rhode Island, Montana, Hawaii, and the District of Columbia.

Overall, we observed that the linear components of behaviors in the 41 states analyzed displayed patterns similar to those observed at the national level, *i.e.*, risk-averting behaviors linearly decreased over time and risk-exposing behaviors linearly increased over time (Hypotheses 2 and 3). For example, in April 2020, the median adherence to avoiding contact with others was 66.9% (95% percentile range [PR]: 56.4%-76.9%), falling to a median adherence of 20.0% (95% PR: 13.7%-27.3%) in May 2022 (**Supplementary Fig. S2A**). Examples of states with low adherence at the start of the study period include Utah and Missouri, while states with high adherence include California and New Jersey. Conversely, in April 2020, the state-level median for going to visit a friend was 8.1% (95% PR: 4.2%-14.8%), increasing to a median of 27.5% (95% PR: 21.1%-32.7%) in May 2022 (**Supplementary Fig. S2B**). Examples of states with high participation in this behavior at the beginning of the study period were Oklahoma and Mississippi, while Michigan and New York had low participation.

We found that oscillations of a risk-averting behavior (specifically, avoiding contact with other people) at the state level were synchronous with national COVID-19 mortality, as 40/41 (97.6%) of the evaluated states had the highest correlation between the behavior trend and mortality at lag 0 **(Table 1)**. Only Delaware had a different correlation pattern, with the highest correlation between behavior and mortality at lag 1. Comparing state-level behavior trends to state-level COVID-19 mortality, we found that their correlations in 24 of the 41 states analyzed shared the pattern we observed at the national level (highest correlation with mortality at lag 0) **(Table 1)**. 14 states had the highest correlations between state-level behaviors and state-level mortality shifted 1 month to the past (*i.e.*, the highest correlation was at lag 1). The remaining 3 states had correlations between behaviors and mortality shifted 1 month into the future (*i.e.*, the highest correlation was at lead 1). We additionally observe that all but one state in the “lag 1” group are categorized as Swing or Republican-leaning states, while all but two of the Democratic-leaning states were in the “lag 0” group which mirrored the correlation pattern we observed at the national level.

**Table 1:** Temporal relationships between state-level behavior and national-level mortality are synchronously unified but relationships between state-level behavior and state-level mortality vary. All but 1 state (Delaware) had the highest correlation between state-level behavior and national mortality at lag 0. When comparing state-level behavior to state-specific mortality, a majority of states (24/41, 58.5%) had behavior trends that were most correlated with mortality at lag 0. The second most common pattern (14/41, 34.1%) shifted the highest correlation between risk-averting behavior oscillations and mortality to 1 month in the past.

| Pattern | National Mortality | State Mortality |
| --- | --- | --- |
| Highest Correlation with Mortality at Lag 0 | Alabama, Arizona, California, Connecticut, Florida, Illinois, Louisiana, Maine, Maryland, Massachusetts, Michigan, Mississippi, Nebraska, New Hampshire, New Jersey, New York, North Carolina, Oregon, Pennsylvania, South Carolina, Texas, Washington, West Virginia, Arizona, Colorado, Idaho, Indiana, Iowa, Kansas, Minnesota, Missouri, Nevada, Ohio, Oklahoma, Tennessee, Utah, Wisconsin, Georgia, Kentucky, Virginia | Alabama, Arizona, California, Connecticut, Delaware, Florida, Illinois, Louisiana, Maine, Maryland, Massachusetts, Michigan, Mississippi, Nebraska, New Hampshire, New Jersey, New York, North Carolina, Oregon, Pennsylvania, South Carolina, Texas, Washington, West Virginia |
| Highest Correlation with Mortality at Lag 1 | Delaware | Arizona, Colorado, Idaho, Indiana, Iowa, Kansas, Minnesota, Missouri, Nevada, Ohio, Oklahoma, Tennessee, Utah, Wisconsin |
| Highest Correlation with Mortality at Lead 1 | None | Georgia, Kentucky, Virginia |

**Table 2:** Democratic states reported higher baseline adherence to risk-averting behaviors and lower baseline adherence to risk-exposing behaviors compared to Swing and Republican states. The mean y-intercept for the linear components of behavior trends in Democratic states was substantially higher than the y-intercepts in Swing and Republican states. Conversely, the y-intercepts for risk-exposing behaviors were substantially lower in Democratic states compared to Swing or Republican states. The similarity of slopes across political leanings suggests that Democratic, Swing, and Republican states experienced simliar rates of decreases (or increases) in adherence even though the baseline levels of adherence differed. SD: Standard deviation.

| Behavior type | Avg. slope (SD) | Avg. y-intercept (SD) |
| --- | --- | --- |
| **NATIONAL** |  |  |
| Risk-averting | -1.300 (0.405) | 72.627 (8.814) |
| Risk-exposing | 0.366 (0.232) | 10.943 (11.960) |
| **DEMOCRATIC** |  |  |
| Risk-averting | -1.310 (0.435) | 74.629 (9.132) |
| Risk-exposing | 0.373 (0.256) | 10.189 (11.025) |
| **SWING** |  |  |
| Risk-averting | -1.318 (0.419) | 70.364 (8.616) |
| Risk-exposing | 0.366 (0.234) | 10.995 (11.634) |
| **REPUBLICAN** |  |  |
| Risk-averting | -1.231 (0.353) | 66.014 (10.461) |
| Risk-exposing | 0.307 (0.218) | 12.660 (13.207) |

In summary, state-level risk-averting behaviors were synchronous with the national mortality trend but temporally varied when correlated with state-level mortality trends. While a substantial proportion of states displayed a synchronous relationship between state-level behavior and state-level mortality, comparisons from some states suggest that behavior trends preceded mortality (*i.e.*, increases in risk-exposing behavior were followed by increases in mortality in the following month) (hypothesis 2). We also observed that the synchronicity between behavior and mortality corresponded to both increased adoption of risk-averting behaviors and reduction in risk-exposing behaviors following increases in mortality (hypothesis 3).

### 2.4 State-level policies and political leaning are associated with differences in adherence to preventive health behaviors

Next, we examined how political leaning (as defined in Methods) and differences in recommendations or policies implemented by different states contributed to differences in behavior trends.

We found that on average, people in Democratic states were more likely to adhere to protective behaviors at the start of the pandemic compared to those in swing or Republican states (**Fig. 3a**, **3d**) (hypothesis 4). A complete comparison of political leaning and the 15 behaviors we analyzied can be found in **Fig. S4**. These results concur with those from the decomposition analysis mentioned previously, as shown by the higher y-intercepts among linear components of each behavior trend among Democratic states compared to those for swing and Republican-leaning states (**Supplementary Table 2**). The slopes of the linear components were not substantially different across political leanings, suggesting that changes in behavioral patterns across the nation were fairly uniform.

**Figure 3:**
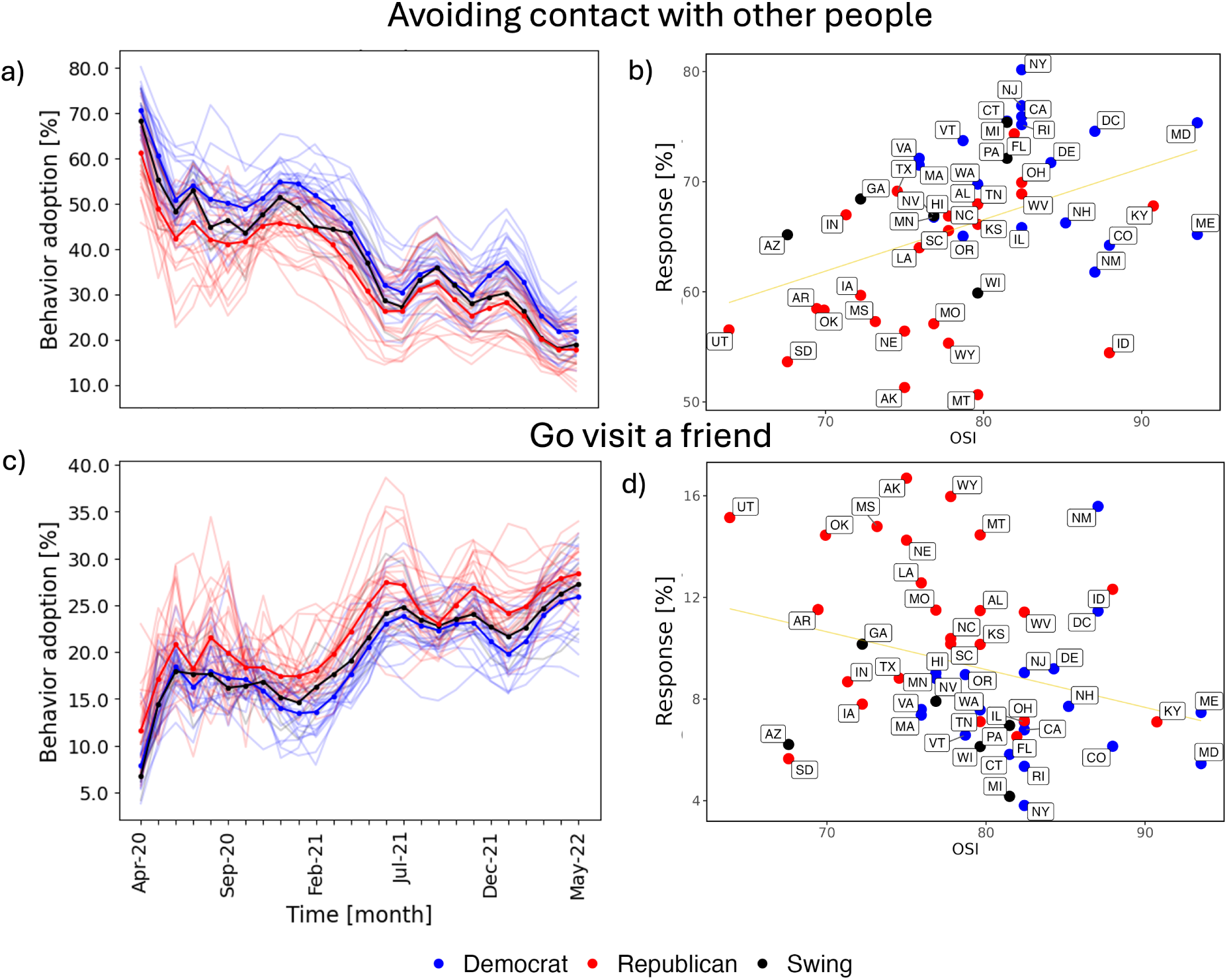
State political leaning influences both trends in behavior and responses to differing levels of stringency of public health recommendations. **a, c**: Survey respondents in Democratic states (blue) were more likely to adhere to preventive health behavior recommendations and less likely to take part in risk-exposing behaviors than those in Republican states (red), even as the adoption of preventive behaviors decreased and participation in risk-exposing behaviors increased over time. **b, d**: For a fixed level of the Oxford Stringency Index (OSI), survey participants from Democratic states were more likely to be strongly adherent to risk-averting recommendations and less likely to take part in risk-exposing behaviors compared to their Republican counterparts. Lines in panels **a** and **c** represent trajectories of behavior participation. Each dot in panels **b** and **d** represents an individual US state.

We also compared participation in both risk-averting and risk-exposing behaviors at the state level at the start of the pandemic to values of the Oxford Stringency Index (OSI) [42], a measure of the strictness of government policies in response to COVID-19, for a single risk-averting and risk-seeking behavior in (**Fig. 3b**, **3d**). We find that for a single value of OSI, participation in behaviors varied across states. Republican states on average exhibited lower participation in risk-averting behaviors and higher participation in risk-exposing behaviors compared to Democratic states, implying that there were discrepancies between the rate of adherence to preventive behaviors as recommended by health authorities and the actual rate of participation as reported by respondents to our survey. For example, for an OSI value of 75 (indicating relatively high stringency of government recommendations), a greater proportion of respondents in the two Democratic states (Massachusetts (MA) and Virginia (VA)) adhered to risk-averting behaviors compared to respondents in the three Republican states (Louisiana (LA), Nebraska (NE) and Alaska (AK)) (hypothesis 5).

For these five states, we also compared the level of adherence to the risk-averting behavior “avoiding contact with other people” to the OSI value for the states across our study period (**Supplementary Fig. S6**). We observed that the behavior curve fits closer to the OSI curve in Democratic states over time compared to in Republican states, even though all five states started with very similar OSI values. We repeated this analysis for all states for the “avoiding contact with other people” behavior (**Supplementary Fig. S6**) and similarly observed higher adherence and closer trajectories between OSI and adherence among Democratic states versus swing and Republican States. Collectively, these analyses show a discrepancy in how the government would like individuals to behave (based on suggested mandates) and how individuals claim they are behaving (as inferred through the survey data). Similar figures to **Fig. 3a**, **3c** and **Fig. 3b**, **3d** for the remaining behaviors are respectively provided in **Supplementary Fig. S4** and **Supplementary Fig. S6**.

## 3 Discussion

In this study, we used data from a representative, non-probability survey to characterize temporal trends of both risk-averting and risk-exposing behaviors, at the national and state levels, during the first two years of the COVID-19 pandemic in the United States. We found that adherence to social distancing (a risk-averting behavior) recommendations was highest at the start of the pandemic and that this adherence waned over time; conversely, participation in risk-exposing behaviors, such as going to visit a friend, steadily increased over time. We showed that these trends could be decomposed into a linear component and an oscillatory component and that the oscillations tracked disease severity metrics, such as COVID-19-attributable mortality and hospitalizations. This synchronicity between population-level behaviors and disease transmission severity indicated that people were more likely to behave cautiously (following risk-averting recommendations) when the risk of COVID-19 was highest and that people were most likely to engage in risk-exposing behaviors such as going to the gym, when the mortality and hospitalization rates were low. Additionally, we identified that at the state level, adherence to risk-averting behaviors and participation in risk-exposing behaviors varied based on state political leaning and that this variation is not comprehensively captured by single-item stringency metrics like the Oxford Stringency Index [43].

Our national-level results have several important implications. First, we found that adherence to the four risk-averting behaviors we analyzed (avoiding contact with other people, avoiding public or crowded places, frequently washing hands, and wearing a face mask outside of your home) was high at the start of our study period in April 2020, with between 60% to 80% of survey respondents reporting adherence to each of these behaviors. These behaviors were all recommended by public health authorities as ways for people to reduce their risk of COVID-19 infection; their high adherence is a testament to successful messaging of these non-pharmaceutical interventions at the start of the pandemic. Second, the linear, consistent decreases in adherence to the risk-averting behaviors over the course of the pandemic are evidence of a combination of factors that wore away at people’s willingness to reduce their risk of infection, such as pandemic fatigue [44, 45], getting infected with SARS-CoV-2, individually not belonging to a high-risk group, or the growing availability of treatments and vaccines. We observed that, for comparable levels of COVID-19-related mortality, in different points in time, the proportion of survey respondents who reported that they were “avoiding contact with other people” declined from 68% in April 2020 to 50% in September 2021 and further to 30% in February 2022, suggesting a population-level desensitization to the effects of the pandemic over time. As our study includes the time period where COVID-19 vaccinations began to be released to portions of the population, we would expect and in fact, observed waning adherence to non-pharmaceutical protective behaviors; however, COVID-19 mortality did not wane to the same degree across this time period, possibly due to delayed uptake associated with different roll-out phases for portions of the population (for example, healthcare workers and the elderly could receive the COVID-19 vaccine earlier than the rest of the population). Third, the correlation of the oscillations of the behavior trends with COVID-19 severity suggests that, despite the steady decay of engagement in cautious behaviors, high numbers of individuals paid attention to the changes in COVID-19 mortality, hospitalizations, or case counts and synchronized their behaviors to the trajectory of the pandemic.

It is noteworthy that wearing a face mask in public demonstrated a different trajectory at the start of the pandemic compared to the other risk-averting behaviors, experiencing an increase in prevalence prior to declining (see fourth row of **Supplementary Fig. S1**). This is reflected mathematically in the PCA as mask-wearing behavior makes the most significant contribution to the second eigen-behavior that accounts about a 10.44% of the variance across behaviors (see **Supplementary Fig. S11** and **Supplementary Fig. S12**), suggesting that mask-wearing was differed from the other behaviors. However, changes in mask-wearing behavior may have been influenced by the limited availability of masks and a lack of clear communication from the Centers for Disease Control and Prevention (CDC) and other government agencies about the benefit of masking in the early phase of the pandemic [46]. For instance, on February 29, 2020, the U.S. Surgeon General advised against buying masks because “they are NOT effective” in preventing the general public from being infected with COVID-19 (although he argued that masks were much needed among the healthcare worker community) [46].

Differences in the temporal relationship between behaviors and COVID-19 mortality compared to the relationship between behaviors and the counts of COVID-19 hospitalizations or cases also have implications for messaging and policy-making. The synchronization of the national-level behavior oscillations and national mortality data at the monthly level (*i.e.*, the strongest correlations at lag 0) suggests a bidirectional feedback loop: information about deaths contributed to changes in behavior just as changes in behavior resulted in shifts in the number of deaths. However, we found that correlations between hospitalizations or cases were relatively strong when the behavior trend was shifted one month forward in addition to at lag 0, indicating that changes in hospitalizations or case counts were followed by changes in behavior. This implies that people’s awareness of changes of the pandemic’s severity could trigger large-scale changes in peoples’ behavior, supporting hypothesis 2. For example, news reports of high case counts or hospitals at full capacity could lead to a reduction in the number of people choosing to travel for a holiday weekend. With these relationships in mind, we can hypothesize further that people may consider hospitalizations or cases (measures of severity that are more likely for any individual) a stronger stimulus for immediate behavior change that is realized over the coming weeks but that deaths (a serious, more concerning endpoint for a disease) are a stronger marker for the overall state of the pandemic.

Conducting the analyses at the state level expanded the context of the national-level results. First, synchronicity analyses comparing state-level behavior to national-level mortality determined that risk-averting behaviors in all but one state (Delaware) were synchronous with trends in national mortality, implying that people across the country were in tune with information about national-level COVID-19 deaths. Second, comparisons between state-level behavior and state-level mortality identified two temporal patterns: one that matched the pattern we found at the national level and one where the highest correlation between mortality and behavior oscillations was shifted one month into the past. A majority of states mirrored the correlation patterns between behavior oscillations of risk-averting behaviors and disease severity found at the national level; this majority included most of the Democratic-leaning states (only Colorado, Minnesota, and Virginia were not in this category). The second pattern implies that past behavior was more correlated with present COVID-19 mortality and therefore that changes in behavior potentially lead to changes in mortality (hypothesis 3); however, the interpretation of this second pattern is limited by our temporal data aggregation method.

Our finding that trajectories of behavior change were similar for almost all states but that there was noticeably higher adherence to risk-averting behaviors in Democratic states compared to Swing or Republican states both at the start of the pandemic and over time lends credence to calls for differing levels of support for public health programs in states of different political backgrounds. This finding correlates with our observation that OSI values (reflecting the estimated stringency of non-pharmaceutical interventions in individual regions) more accurately reflected the adherence levels of respondents in Democratic states compared to those in Swing states or Republican states (because those respondents were more likely to be adherent in general). These findings, combined with evidence highlighting increased excess mortality due to COVID-19 among Republican leaning counties compared to Democratic ones [47, 48, 49, 50] and differences in COVID-19 testing consistency [51] and reporting based on the political leaning of state governors [52] suggests that baseline differences in attitudes toward public health and pandemic preparedness could be potential targets for improvement as opposed to addressing policies governing the deployment of emergency services during a future pandemic.

With these policy considerations in mind, our data and our analyses aim to fill a gap in the availability of high-quality epidemiological data related to the severity of an epidemic and human behavior. Past research has shown that accounting for behavior change improves epidemiological models’ ability to capture the trajectory of a disease and make predictions compared to models that do not explicitly account for behavior change [53, 54, 55]. However, in the absence of data like the survey data we present, these models often rely on disease severity metrics to serve as proxies of human behavior. For example, the transmission rate parameter of a susceptible-exposed-infectious-recovered (SEIR) model can be written as a function of the prevalence of the disease, such that when cases increase, transmission decreases and vice versa. Our analyses shows that behavior data were more correlated with mortality data than reported hospitalization or case data during our study period. This suggests that it may be more accurate to induce behavior change in COVID-19 models as a response to the number of deaths than to the number of cases or even hospitalizations. Additionally, the observed linear trends of behaviors suggest that there is a need to incorporate multiple mechanisms of behavior change into a model. Specifically, only incorporating feedback from mortality to behavior may not be enough, particularly when models are used to fit data over a long period of time.

Furthermore, it is known that human behavior can vary by region, potentially requiring region-specific behavior-related parameters to be included in any epidemiological model. The time series data of 15 risk-averting and risk-seeking behaviors across all states may help modelers parameterize their models specifically for their region of interest. For example, modelers looking to capture granular data (such as changes in patterns in visits made to a workplace, restaurant, or a hospital) through a complex agent-based model (ABM) may benefit from our time series state-level data on these behaviors. Furthermore, although numerous models have incorporated masking behavior, very few, if any, have included time series masking behavior data into their model. The inclusion of this time series masking data (which, for the reasons discussed above, might differ from intuitive expectation) may not only add realism to the model but also help accurately assess the impact of masking. Overall, incorporating region-specific time series risk-averting and risk-exposing behaviors into models would add realism to the modeling effort.

Such region-specific time series can be further complemented or augmented with cellphone-based location data. However, evidence from the pandemic suggests that the specificity of location data is necessary to draw insightful conclusions from such data. For example, Kogan and colleagues [56] identified that a large state-level drop in human mobility (measured using Cuebiq and Apple mobility data) led to an eventual decrease (4 weeks later) in deaths during the first COVID-19 wave; however, this prominent relationship between large changes in human mobility (obtained from aggregated mobile phone records) and changes in disease transmission was not consistently observed in subsequent COVID-19 outbreaks [57]. Perofsky and colleagues evaluated the correlation between COVID-19 disease metrics (such as the effective reproduction number) and aggregated mobility data (such as visits to restaurants, religious organizations, groceries, and pharmacies) during the first two years of the pandemic and found that “mobility is most predictive of respiratory virus transmission during periods of dramatic behavioral change and at the beginning of epidemic waves” [58]. They found that the effective reproduction number was not significantly associated with mobility. In this study, we show that the survey data, even when aggregated at the national and state levels, continued to have a strong relationship with mortality during our 2-year study period.

We highlight that beyond the value of our survey data in monitoring changes in human behavior, as shown in this study, our research team has also shown that our survey data have been able to closely track COVID-19 infection rates, at the national level, during the first two years of the pandemic. Moreover, we have shown that our survey data may have better captured COVID-19 infections in the population after the mass distribution of rapid at-home tests (in the absence of a centralizing government-led system to report positive infection results from at-home tests) in January 2022 [18]. In the context of vaccine resistance, our survey data has also been successful at uncovering relationships between an individual’s habits to obtain their news from conservative media outlets (e.g. Fox news) and social media platforms, most notably Facebook, and their willingness to get a COVID-19 vaccine [59]. Multiple other studies using our data have uncovered undesirable outcomes of lockdown conditions during the COVID-19 on individuals’ mental health [60].

Our study has several limitations. First, the survey data we used in this study were collected in waves that varied in duration and frequency, thus requiring us to use interpolation methods to ensure we had data for each month of the study period. This coarse temporal resolution (monthly) prevented us from being able to fully characterize the bi-directional feedback between behaviors and severity that most likely takes place in daily or weekly time intervals. Second, our analysis was not restricted to responses to (a) yes/no binary questions, which could lead to different population-level behavior outcomes depending on the choice of responses that are chosen to define the *adherent* vs *non-adherent* threshold; and (b) included responses from participants who participated multiple times (about 15%). In both cases, i.e. (a) when choosing different *adherent* vs *non-adherent* threshold for non-binary questions, and (b) removing repeat participants, our results do not change in any meaningful way and remain consistent with what was summarized in this manuscript. Sensitivity analyses are shown in **Supplementary Fig. S8** and **Supplementary Fig. S9**. Third, although this study was able to show differences in behavior across Democratic, Swing, and Republican states, further analysis of our data should pursue to identify the extent to which Republican (or Democratic) individuals in Republican leaning states behaved similarly to Republican (or Democratic) individuals in Democratic leaning states. Individual-level analysis would also allow us to better understand the role of socioeconomic and sociodemographic factors, as well as the role of the location-specific environmental variables (such as local mandates) on adherence to different behaviors. Finally, we recognize that a continuous, monotonic linear increase or decrease for the adherence rate to any of the behaviors we studied could not extend infinitely forward in time (as adherence rates cannot be negative or exceed 100%); our observations of such trends during the study period encourage future work that evaluates when asymptotes or plateaus of these trends occurred later in the pandemic.

In conclusion, we expect that the available survey data will be useful in the parameterization of epidemiological models that integrate behavior into their processes by providing multiple behavior change metrics over the course of an extended outbreak. Such data should be especially useful for long-term prediction of disease trajectories or the building of policies that seek to provide effective prevention tools before the onset of disease outbreaks.

## 4 Methods

### 4.1 Survey data

We used survey data from the COVID States Project [61], a non-probability survey started on April 16, 2020 in the United States. Previous research has shown that this survey provided accurate estimates of health indicators like COVID-19 vaccination and infection rates in the U.S. [62, 59, 18] and depression [63]. In addition, it matches administrative data or other surveys in other domains, including vote share in the 2020 elections [64], gun purchases [65] or participation in the BLM protests [66]. Following the American Association for Public Opinion Research (AAPOR) reporting guidelines [67], we provide details on the recruitment, weighting, and survey content of the survey data. Most respondents are recruited through PureSpectrum, a survey platform that aggregates survey respondents from different online survey vendors. An additional small percentage of survey responses (4.6%) are recruited through Facebook ads. The recruitment through PureSpectrum does not specify the focus of the survey on COVID-19 related topics, reducing the risk of selection bias. Survey respondents are 18 years or older and reside in the United States.

The data consist of several survey waves, defined as the distinct periods of time in which responses are gathered from survey respondents, that were fielded approximately every 6 weeks. Each survey wave contains about 20,000 survey responses, with viable sample sizes for most U.S. states in most waves (see **Section 5** for details on the states with smaller samples). For this study, we used data from 19 survey waves, collected between April 2020 and June 2022. A detailed description of the fielding period of each survey wave and its total sample size can be found in **Supplementary Table S3**. We also provide the average number of respondents per wave for each state in **Supplementary Table S4**. Survey respondents were permitted to participate in more than one survey wave; in total, we used 431,211 survey responses from 307,771 different respondents. We test for the robustness of our results to the removal of responses from repeat participants in **Supplementary Section 5**.

We use quotas and post-stratification weights to ensure the representativeness of the COVID States samples, nationally and at the state level. State-level quotas for age, gender and race/ethnicity were used at the sampling stage to approximate the demographic composition of each state in the survey sample. Weights based on interlocking race/ethnicity-gender-age subgroups as well as education, rurality, and region were used at the national level. A separate set of weights was used at the state level, matching the gender, race, age, education, and rurality composition of the survey samples with the composition of the state from which responses were collected. The top and bottom 1% of the weights are trimmed to reduce the influence of a small set of respondents. Population benchmarks are taken from the US Census Bureau. The surveys also include multiple closed and open-ended attention checks, with the goal of filtering out inattentive respondents, addressing a common issue associated with online opt-in surveys [68, 69]. Around 25% of respondents are filtered due to failed attention checks, and the attrition rate is roughly 15%-20%. The surveys include questions on health, social, informational, and political issues related to the COVID-19 pandemic. The questions analyzed in this study were included in blocks of questions related to preventive behaviors (such as vaccinations or avoiding infection risks) and social behaviors, typically included around the middle of the survey. We provide the full survey text for each survey wave upon request. The study design was determined to be exempt by the institutional review boards of Harvard University and Northeastern University. Moreover, survey participants accepted an electronic informed consent online before having access to the survey.

### 4.2 Survey collected behavior data

The behavior data was obtained from survey questions encompassing both risk-averting and risk-exposing behaviors. For risk-averting behaviors, we used survey answers to the question “In the last week, how closely did you personally follow the health recommendations listed below?”, that was followed-up by four behaviors: “Avoiding contact with other people”, “Avoiding public or crowded places”, “Frequently washing hands”, and “Wearing a face mask when outside of your home”. Possible responses to these questions ranged from 1 to 4, where 1 represented “Not at all closely”, 2 represented “Not very closely”, 3 represented “Somewhat closely”, and 4 represented “Very closely”. We defined adherence or participation in risk-averting behaviors as answering “Very closely”, but also include a sensitivity analysis in **Supplementary Fig. S9** where we defined adherence as the proportion of participants that answered “Somewhat closely” or “Very closely.”

For risk-exposing behavior, we asked, “In the last 24 hours, did you or any members of your household do any of the following activities outside of your home?”, which included possible responses such as “Go visit a friend” and “Go to a cafe, bar, or restaurant”, and “In the last 24 hours, have you been in a room (or another enclosed space) with people who were not members of your household?”, with answers options ranging from “No, I have not” to “Yes, with over 100 other people”. We aggregate the answers to this question in four different groups for our main analysis and test two additional groupings in **Supplementary Fig. S9**. Details of the questions, answers, and aggregation procedures as well as the missing data for each question can be found in **Supplementary Materials Section 5.** We calculate the 95% confidence intervals corresponding to each survey estimate, at the national level and for each state, using the standard error provided by the *survey* package in R, that uses a design effect to take into account the influence of survey weights on errors. A summary of the states with larger error margins on average can be found at **Supplementary Materials Section 5** while **Supplementary Materials Section 4** provides sample size at a national level and **Supplementary Table S4** provides sample size per survey wave at a state level. Results presented at the national level include data from all 50 states and the District of Columbia; however, state-level results related to behavior data include only the 40 states that had less than 200 responses for each behavior question for no more than one survey wave (the excluded regions were Alaska, Wyoming, North Dakota, South Dakota, New Mexico, Vermont, Rhode Island, Montana, Hawaii, and the District of Columbia).

### 4.3 COVID-19 severity data

We obtained COVID-19 mortality data from the Johns Hopkins University COVID-19 Data Repository [70] and COVID-19 hospitalization and detected case data from Our World in Data [71]. We used the 2021 United States Census estimates [72] to scale survey response levels to the population of each state. State political leaning was determined by aggregating publicly available data from CNN on the results of the 2016 and 2020 United States presidential elections [73]. We obtained Oxford Stringency Index (OSI) values from the University of Oxford Coronavirus Government Response Tracker GitHub page [74].

### 4.4 Data preparation

To account for differences in the length of each survey wave and the time interval between individual waves, we used polynomial interpolation (specifically, second-order interpolation) to generate estimates of behavioral data for each month of the study period. For simplicity, we refer to the data generated through interpolation as *behavior data*.

Since the behavior data in some form is compared with disease severity data (namely, mortality, hospitalization, and case data) monthly time series disease severity data is generated. Specifically, COVID-19 death data is obtained from the Johns Hopkins University COVID-19 Data Repository [70] while COVID-19 hospitalizations and detected case data are obtained from Our World in Data [71].

### 4.5 Decomposition of behavior data

We decomposed behavior data into temporal linear and oscillatory components. The linear trends were obtained by fitting a linear model to each of the 15 risk-averting and risk-exposing behaviors. The oscillatory component of the behavior data is the deviation of the behavior data from the linear model. In other words, the oscillatory component is the residual of the linear model.

### 4.6 Demographic data

To assess the influence of political leaning on each individual behavioral trend, we categorized each state as either a Democratic, Republican, or swing state based on the voting results in the past two United States presidential elections (2016 and 2020; see Table S2 in Supplementary Materials for more information). States were classified as swing states if the state did not vote to elect a presidential candidate from the same political party in consecutive elections (for example, in 2016, Wisconsin voted to elect Donald Trump, the Republican candidate, but voted in 2020 to elect Joe Biden, the Democratic candidate).

We also evaluated if a simple stringency measure could serve as a proxy for our behavioral time-series data by comparing the prevalence of each behavior during survey wave 1 for fixed levels of stringency. We chose the Oxford Stringency Index (OSI) [43, 75, 42] for our comparison, which is a composite index that reflects the average level of restriction in a geographic sub-region. OSI is based on nine mitigation policies, including cancellation of public events, school closures, gathering restrictions, workplace closures, border closures, internal movement restrictions, public transport closure, recommendations to stay at home, and stay-at-home orders [76]. The OSI ranges from 0 to 100, where 100 is the most stringent level of restriction. We hypothesized that our time-varying prevalence estimates would be useful if we observed differences in prevalence across different values of the OSI.

### 4.7 Correlation analysis

Lagged correlation analysis is conducted by temporally shifting the oscillatory component of risk-averting and risk-exposing behaviors with respect to the trends in disease severity metrics, namely COVID-19 mortality, hospitalizations, and detected cases. A lag of zero indicates no temporal shift in the oscillatory component of behavior, that is disease severity metrics are being correlated with this month’s behavior. Meanwhile, a positive lag indicates that disease severity metrics are correlated with future months’ behavior, and a negative lag indicates that disease severity metrics are correlated with past months’ behavior.

### 4.8 Principal component analysis

Principal component analysis (PCA) is used to transform a large set of variables (here, fifteen risk-exposing and risk-averting behavior data) into a smaller set that contains most of the information of the original variables. This is done by creating new variables, called principal components (referred to in this study as the *eigen-behaviors*), which are linear combinations of the original variables. The first eigen-behavior explains the largest variance of the behavior data. The subsequent eigen-behaviors explain the next largest amount of variance while remaining orthogonal to all previous eigen-behaviors (i.e., the subsequent eigen-behaviors capture variance that is not captured by the earlier eigen-behaviors). The contribution of each variable towards the eigen-behaviors can be assessed from principal component coefficients (loadings). A positive or negative loading respectively indicates a positive or negative correlation towards the eigen-behaviors.

## Data Availability

All data produced in the present study are available upon reasonable request to the authors

## Data availability

Data and analysis code used in the study can be found in the following Github Repository: https://github.com/tam-urmi/behavior_covid_states.

## Acknowledgments

Dr. Santillana acknowledges that the study was supported in part by the cooperative agreement CDC-RFA-FT-23-0069 from the Centers for Disease Control and Prevention’s Center for Forecasting and Outbreak Analytics. Dr. Druckman acknowledges the support the National Science Foundation (NSF) under grants SES-2029292, SES-2029792, SES-2116465, SES-2116189, SES-2116458, SES-211663, SES-2241884, SES-2241885, SES-2241886, and SES-2241887 as well as the John S. and James L. Knight Foundation, Amazon Web Services, and the Peter G. Peterson Foundation. Dr. Ognyanova acknowledges the support of NSF under grant SES 2241886. Dr. Baum acknowledges the support of Harvard’s Shorenstein Center on Media and Public Policy. Dr. Perlis acknowledges support in part by the National Institute of Mental Health (R01MH123804 and U01MH136059).

