## Supplementary materials for "Characterizing Population-level Changes in Human Behavior during the COVID-19 Pandemic in the United States"

#### Supplementary Sections

**1 National-level behavior trends** 3

**2 Lag between COVID-19 cases, hospitalization and mortality** 6

**3 State political leaning** 7

**4 Survey waves** 12

**5 Survey questions, missing data, and state sample sizes** 13

**6 Principal component analysis (PCA)** 18

**7 Correlation between behaviors** 19

**7 State-level behavioral trends and their relationships with epidemic severity** 20

**8 State-level correlations between oscillations in behavior trends and disease severity metrics** 72

**9 Correlations between state-level oscillations in behavior trends and national disease severity metrics** 98

*These authors contributed equally to this study

### 1 National-level behavior trends

| **Behaviors** | **Slope (% change/month)** | **Y-intercept (% adherence)** | **Adherence % at the beginning of the study period** | **Adherence % at the end of the study period** | **Absolute difference in adherence from beginning to end** | **Absolute percentage change in adherence from beginning to end** |
|---|---|---|---|---|---|---|
| Avoiding contact with other people | -1.506 | 61.032 | 68.77 | 20.94 | 47.82 | 69.54 |
| Avoiding public or crowded places | -1.617 | 70.703 | 75.23 | 27.02 | 48.20 | 64.08 |
| Frequently washing hands | -0.713 | 78.058 | 80.36 | 58.85 | 21.51 | 26.77 |
| Wearing a face mask when outside of your home | -1.364 | 80.716 | 55.96 | 33.74 | 22.23 | 39.72 |
| Go to work | 0.31 | 31.884 | 25.71 | 39.94 | 14.26 | 55.38 |
| Go to the gym | 0.376 | 3.627 | 1.36 | 12.65 | 11.28 | 827.80 |
| Go visit a friend | 0.497 | 13.027 | 8.31 | 26.52 | 18.20 | 219 |
| Go to a cafe, bar, or restaurant | 0.689 | 10.063 | 5.78 | 28.47 | 22.69 | 392.65 |
| Go to a doctor or visit a hospital | 0.162 | 9.899 | 5.54 | 14.29 | 8.75 | 157.73 |
| Go to church or another place of worship | 0.297 | 3.336 | 1.29 | 10.23 | 8.94 | 692.78 |
| Take mass transit (e.g. subway, bus, or train) | 0.121 | 2.286 | 1.82 | 5.40 | 3.58 | 196.86 |
| Been in a room with someone outside of household in the past 24 hours | 0.864 | 35.535 | 26.26 | 26.77 | 30.51 | 116.19 |
| Been in a room with 5-10 people outside of household in the past 24 hours | 0.261 | 8.097 | 5.15 | 14.79 | 9.65 | 187.26 |
| Been in a room with 11-50 people outside of household in the past 24 hours | 0.271 | 2.015 | 1.42 | 9.54 | 8.12 | 571.98 |
| Been in a room with over 50 people outside of household in the past 24 hours | 0.172 | 0.601 | 0.83 | 5.36 | 4.54 | 549.24 |

Table S1: **Adherence metrics across different behaviors at a national level.** The slopes and y-intercept of lines fitted to the fifteen risk-averting and risk-exposing behavior trends between April 2020 and May 2022 as shown in Fig. S1. The percentage of adherence at the beginning (April 2020) and end (May 2022) of the study period.

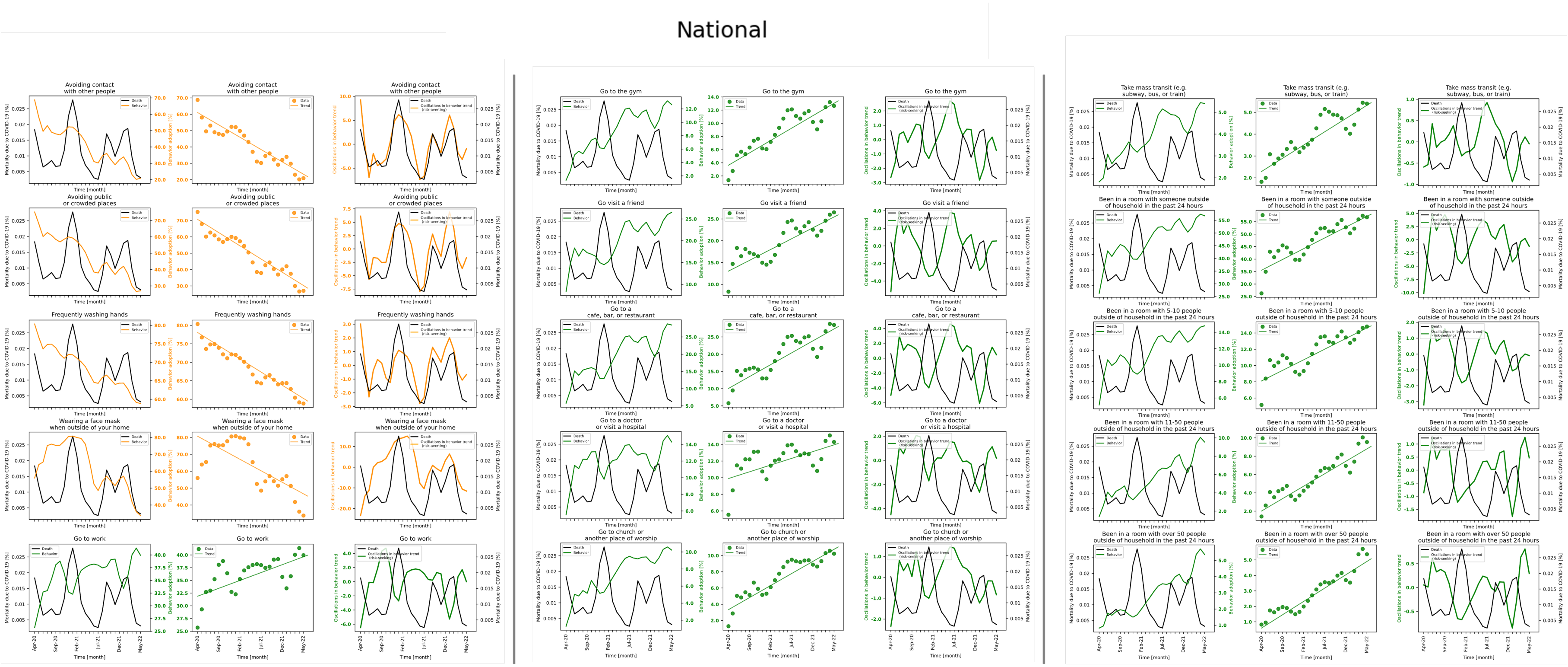

Figure S1: **Decomposition of national-level behavior trends into linear and oscillatory components reveals synchronicity with COVID-19 mortality**. The decomposition was performed for both risk-averting (orange) and risk-exposing (green) behaviors.

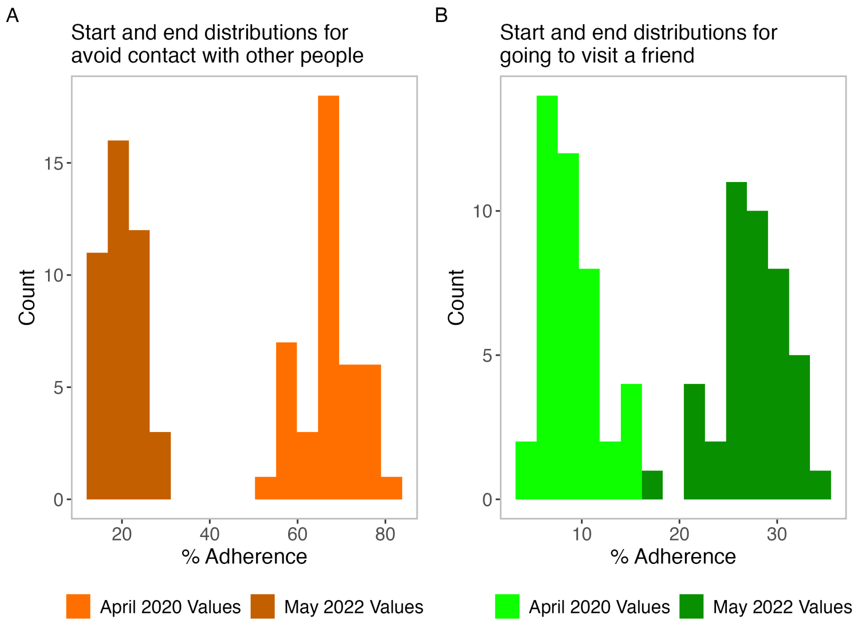

Figure S2: **State-level differences for risk-averting and risk-exposing behaviors between the start and end of the evaluation period mirror trends at the national level.** For risk-averting behaviors (orange), states experienced linear decays from the start to the end of the study period, while for risk-exposing behaviors (green), states experienced linear increases. Lighter shades represent rates of behavior adherence or participation in April 2020 (the start of the evaluation period) while darker shades represent rates of behavior in May 2022 (the end of the evaluation period).

### 2 Lag between COVID-19 cases, hospitalization and mortality

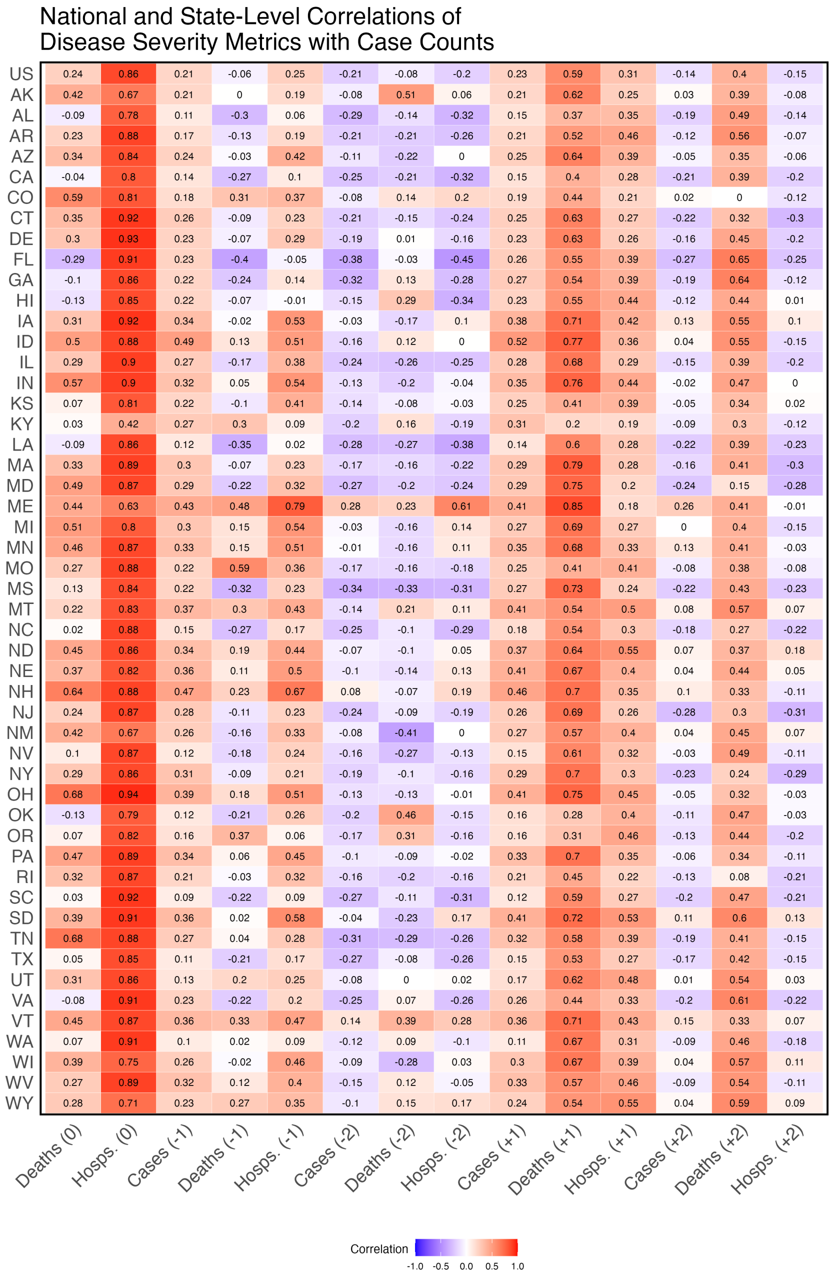

Figure S3: **National and state-level correlation of COVID-19 hospitalization and mortality with cases.**

### 3 State political leaning

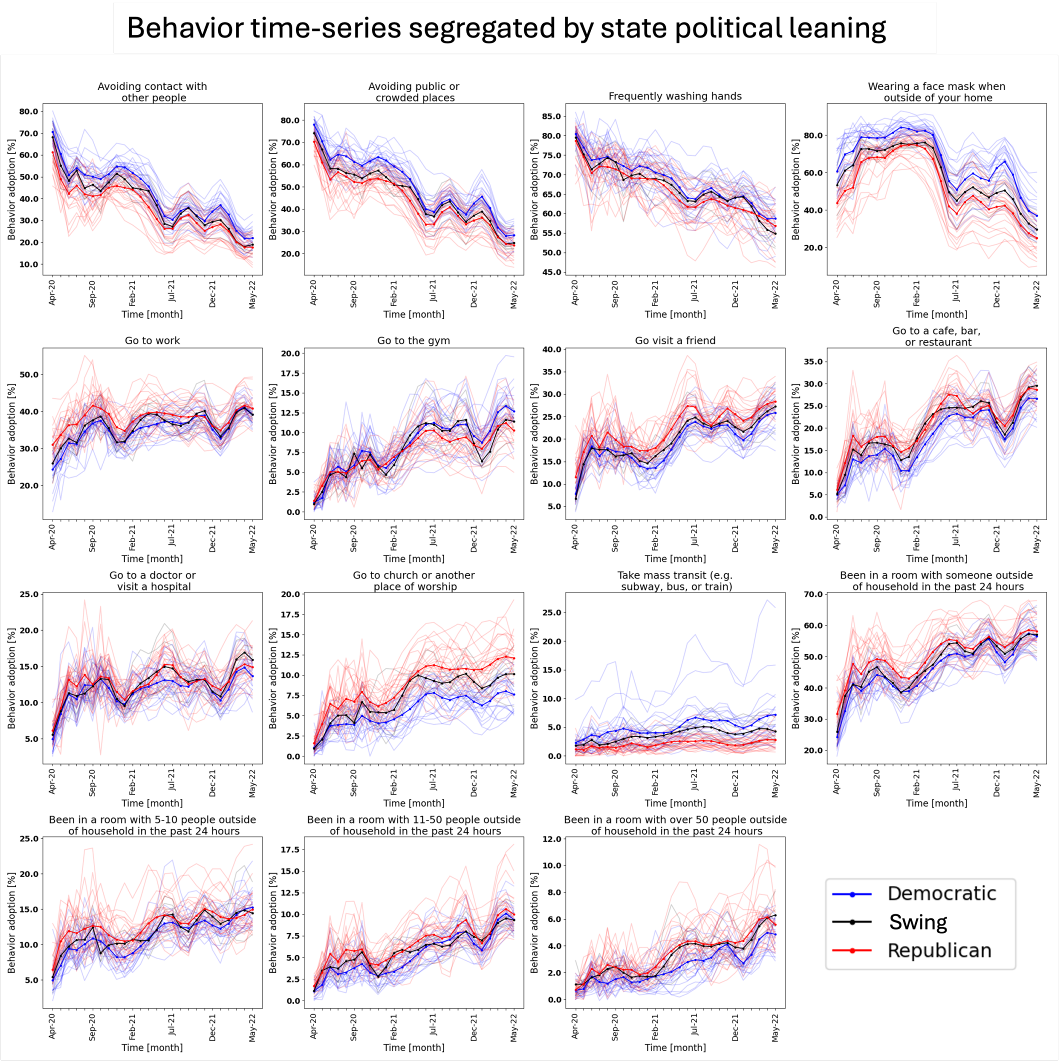

Figure S4: **Trends in risk-averting and risk-exposing behaviors differed based on state political leaning.** Each line represents the trend of an individual behavior based on survey responses from participants from each state.

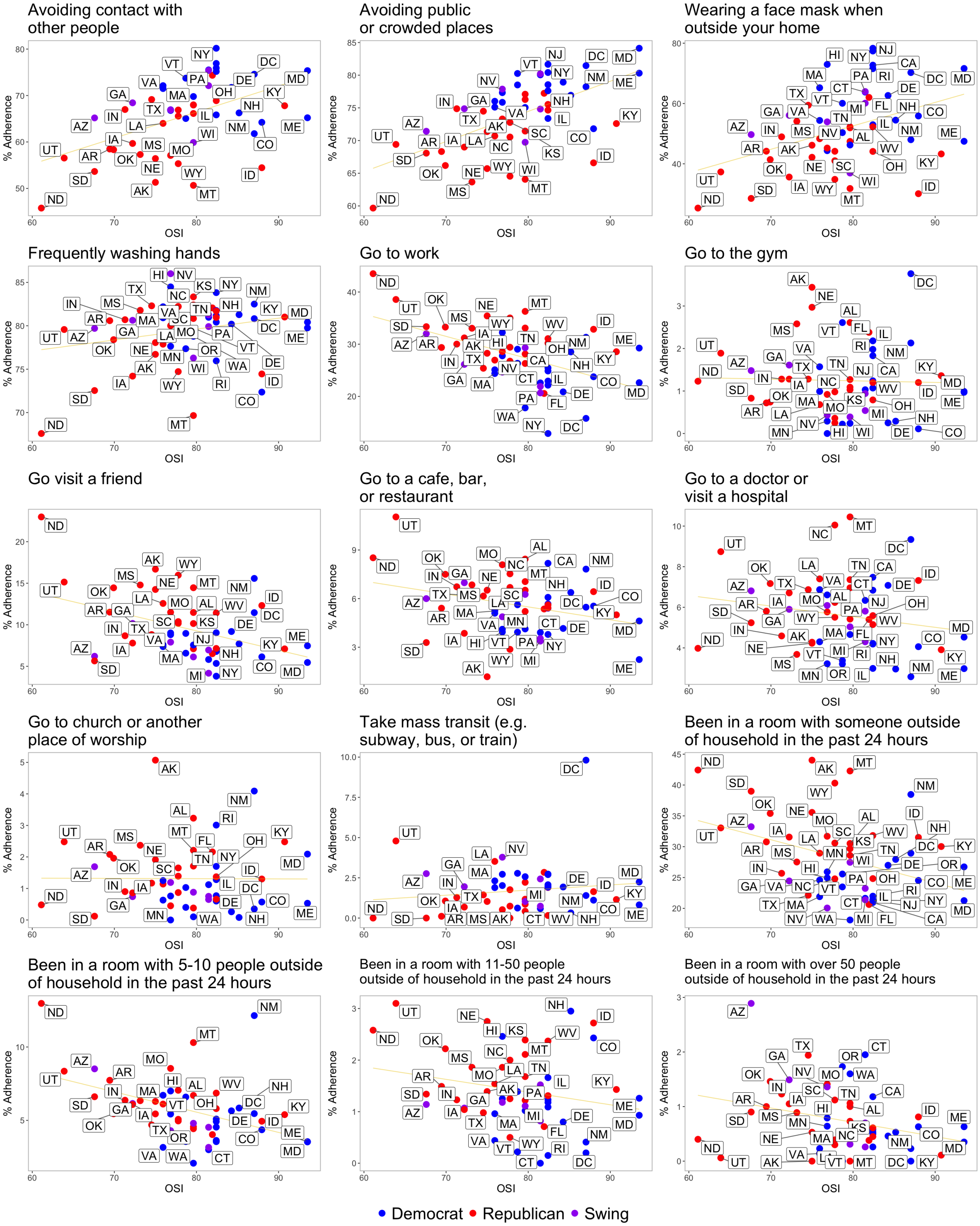

Figure S5: **Scatterplot of Oxford Stringency Index (OSI) and percentage adherence to risk-averting and risk-exposing behaviors for each state during Wave 1 of the survey.** The OSI values correspond to the OSI as calculated on the midpoint date of Survey Wave 1 (04/24/2020).

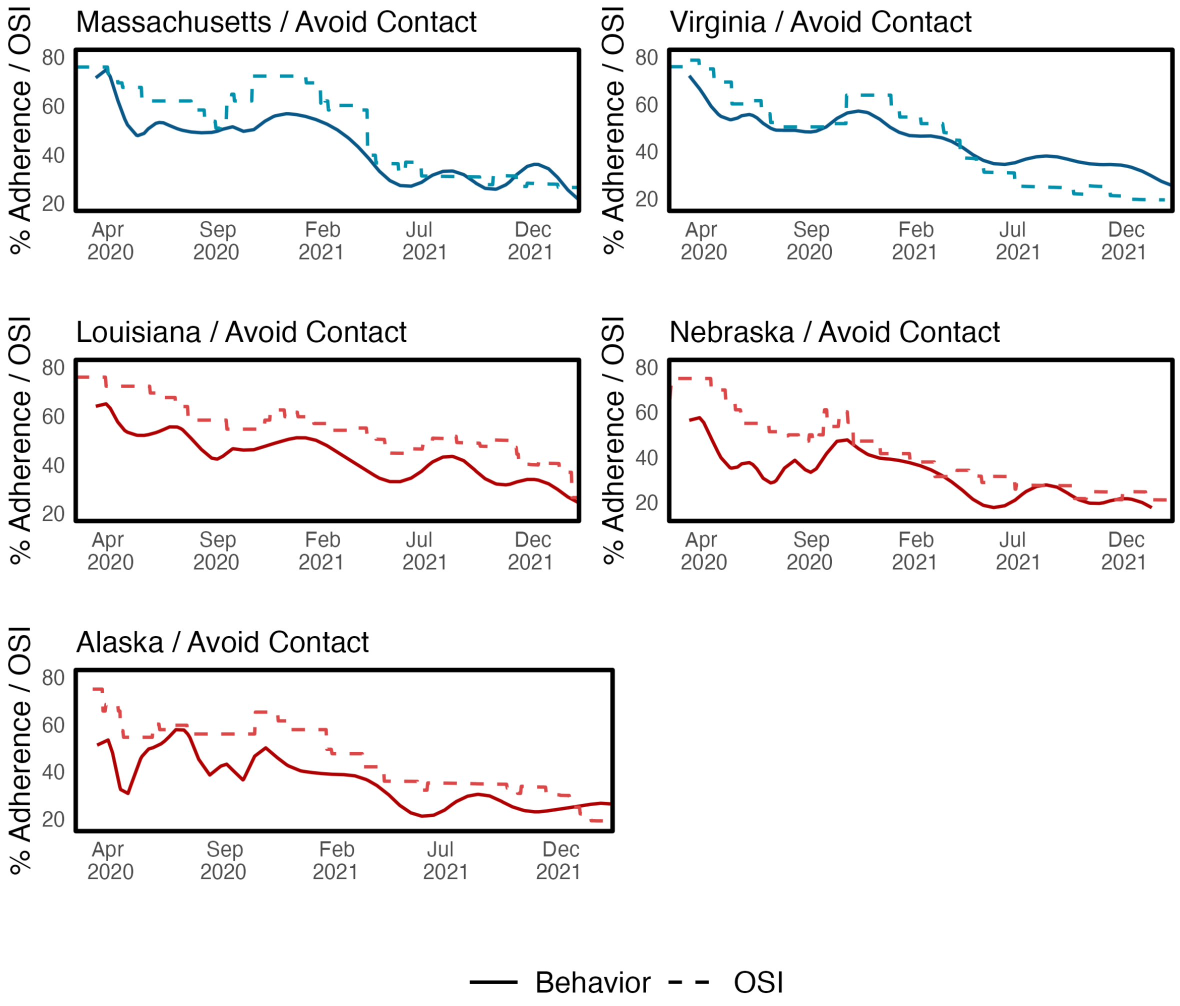

Figure S6: **Adherence to a risk-averting behavior (avoiding contact with other people) fits closer to Oxford Stringency Index (OSI) values in Democratic states versus Republican states**. The five states shown start at the same OSI value at the start of the study period but changed their recommendations for preventive measures differently over time. Blue/red lines indicate OSI values, orange lines indicate reported adherence to avoiding contact with other people.

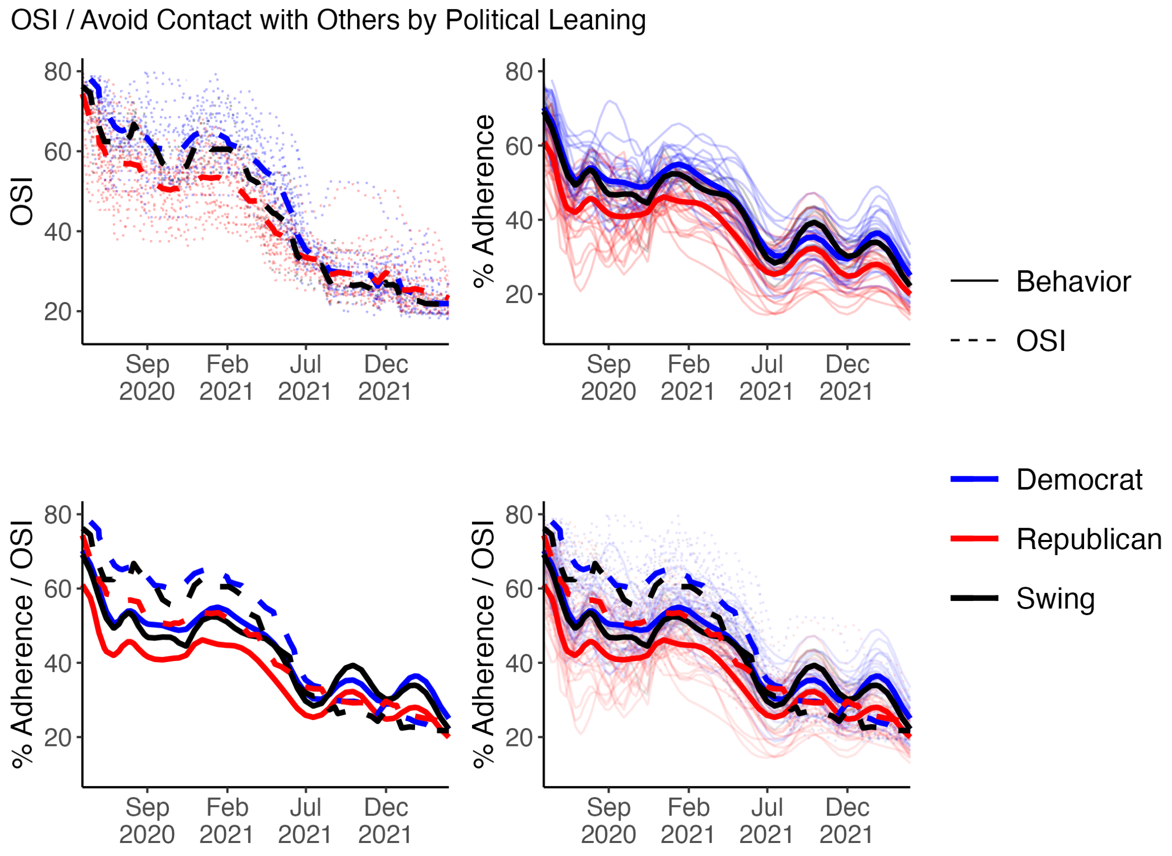

Figure S7: **Oxford Stringency Index (OSI) values overestimate adherence to a risk-averting behavior ("avoiding contact with other people") in Democrat, Republican, and Swing states.** The deviation between the OSI curves and the behavior curves reflect the overestimation of behavior prevalence if OSI levels are considered to reflect individuals' participation in preventive behaviors. Dashed lines indicate OSI values, while solid lines indicate level of adherence to the selected behavior. The bolded lines reflect the average OSI or adherence level by political leaning across states. Individual state time series for either OSI or behavior are lighter lines in the figure background.

Table S2: **U.S. states and the District of Columbia and their political classification (Democratic, Republican, or Swing).**

| State | Code | Political Affiliation |
|---|---|---|
| Alabama | AL | Republican |
| Alaska | AK | Republican |
| Arizona | AZ | Swing |
| Arkansas | AR | Republican |
| California | CA | Democrat |
| Colorado | CO | Democrat |
| Connecticut | CT | Democrat |
| Delaware | DE | Democrat |
| District of Columbia | DC | Democrat |
| Florida | FL | Republican |
| Georgia | GA | Swing |
| Hawaii | HI | Democrat |
| Idaho | ID | Republican |
| Illinois | IL | Democrat |
| Indiana | IN | Republican |
| Iowa | IA | Republican |
| Kansas | KS | Republican |
| Kentucky | KY | Republican |
| Louisiana | LA | Republican |
| Maine | ME | Democrat |
| Maryland | MD | Democrat |
| Massachusetts | MA | Democrat |
| Michigan | MI | Swing |
| Minnesota | MN | Democrat |
| Mississippi | MS | Republican |
| Missouri | MO | Republican |
| Montana | MT | Republican |
| Nebraska | NE | Republican |
| Nevada | NV | Democrat |
| New Hampshire | NH | Democrat |
| New Jersey | NJ | Democrat |
| New Mexico | NM | Democrat |
| New York | NY | Democrat |
| North Carolina | NC | Republican |
| North Dakota | ND | Republican |
| Ohio | OH | Republican |
| Oklahoma | OK | Republican |
| Oregon | OR | Democrat |
| Pennsylvania | PA | Swing |
| Rhode Island | RI | Democrat |
| South Carolina | SC | Republican |
| South Dakota | SD | Republican |
| Tennessee | TN | Republican |
| Texas | TX | Republican |
| Utah | UT | Republican |
| Vermont | VT | Democrat |
| Virginia | VA | Democrat |
| Washington | WA | Democrat |
| West Virginia | WV | Republican |
| Wisconsin | WI | Swing |
| Wyoming | WY | Republican |

### 4 Survey waves

Table S3: **Survey wave periods and the gaps between waves.** Waves 4, 6, 8, and 15 have small sample sizes with only national-level data and are omitted from the dataset. The average number of respondents (N) for the 19 survey waves considered in this study is over 20,000.

| Wave | Start Date | End Date | Length [days] | N |
|---|---|---|---|---|
| 1 | 2020-04-16 | 2020-04-30 | 14 | 21405 |
| 2 | 2020-05-01 | 2020-05-21 | 20 | 23814 |
| 3 | 2020-05-16 | 2020-06-01 | 16 | 20391 |
| gap | 2020-06-02 | 2020-06-11 | 9 | NA |
| 5 | 2020-06-12 | 2020-06-28 | 16 | 22905 |
| gap | 2020-06-29 | 2020-07-09 | 10 | NA |
| 7 | 2020-07-10 | 2020-07-26 | 16 | 19437 |
| gap | 2020-07-27 | 2020-08-06 | 10 | NA |
| 9 | 2020-08-07 | 2020-08-26 | 19 | 21496 |
| gap | 2020-08-27 | 2020-09-03 | 7 | NA |
| 10 | 2020-09-04 | 2020-09-30 | 26 | 23050 |
| 11 | 2020-10-02 | 2020-10-23 | 21 | 19570 |
| 12 | 2020-10-23 | 2020-11-14 | 22 | 12127 |
| 13 | 2020-11-03 | 2020-11-30 | 27 | 26642 |
| gap | 2020-12-01 | 2020-12-15 | 14 | NA |
| 14 | 2020-12-16 | 2021-01-12 | 27 | 26113 |
| gap | 2021-01-13 | 2021-02-04 | 22 | NA |
| 16 | 2021-02-05 | 2021-02-28 | 23 | 23348 |
| gap | 2021-03-01 | 2021-03-31 | 30 | NA |
| 17 | 2021-04-01 | 2021-05-03 | 32 | 23718 |
| gap | 2021-05-04 | 2021-06-08 | 35 | NA |
| 18 | 2021-06-09 | 2021-07-15 | 36 | 22275 |
| gap | 2021-07-16 | 2021-08-25 | 40 | NA |
| 19 | 2021-08-26 | 2021-09-28 | 33 | 23938 |
| gap | 2021-09-29 | 2021-11-02 | 34 | NA |
| 20 | 2021-11-03 | 2021-12-02 | 29 | 24623 |
| gap | 2021-12-03 | 2021-12-21 | 18 | NA |
| 21 | 2021-12-22 | 2022-01-25 | 34 | 25353 |
| gap | 2022-01-25 | 2022-03-01 | 35 | NA |
| 22 | 2022-03-02 | 2022-04-09 | 38 | 23376 |
| gap | 2022-04-10 | 2022-06-07 | 58 | NA |
| 23 | 2022-06-08 | 2022-07-06 | 28 | 24625 |

### 5 Survey questions, missing data, and state sample sizes

We include in this section the full survey questions and possible answers corresponding to the data we use. We also explain how the survey answers are aggregated to generate the percentages we use in our analysis, and provide details on the missing data corresponding to each question. Finally, we discuss the sample sizes and error margins in each state.

We use the following three survey questions:

- In the last 24 hours, did you or any members of your household do any of the following activities outside of your home? (Please select all that apply)

  Go to work, Go to the gym, Go to the store, Go visit a friend, Go out for a walk or a run, Go to a cafe, bar, or restaurant, Go to a doctor or visit a hospital, Go to church or another place of worship, Go somewhere else outside of your home, Take mass transit (e.g. subway, bus, or train)

  Each behavior is coded as 1 if the respondent selected the corresponding behavior, or 0 if they did not. The default response is 0, therefore, all respondents who receive the question while filling the survey but do not select a given behavior are coded as 0. This question is asked to all respondents in all waves except waves 11, 12, and 13, in which only a fraction of the respondents in a set of low-population states were asked the question. In total, 1663, 2412 and 3362 respondents were not asked the question in waves 11, 12, and 13, respectively. We provide in Supplementary Tab. S4 the average number of respondents per wave per state.

- In the last 24 hours, have you been in a room (or another enclosed space) with people who were not members of your household? This might have been at a social gathering, a work meeting, or another type of event.

  1.“No, I have not”, 2.“Yes, with 1-2 other people”, 3.“Yes, with 3-4 other people”, 4.“Yes, with 5-6 other people”, 5.“Yes, with 7-8 other people”, 6.“Yes, with 9-10 other people”, 7.“Yes, with 11-50 other people”, 8.“Yes, with 51-100 other people”, 9.“Yes, with over 100 other people”

  For the main text analysis, we calculate the percentage of respondents who selected answers 2 to 9 (“Been in a room with someone outside of household in the past 24 hours”), answers 4, 5, or 6 (“Been in a room with 5-10 people outside of household in the past 24 hours”), answers 7 and 8 (“Been in a room with 11-50 people outside of household in the past 24 hours”), and answer 9 (“Been in a room with over 50 people outside of household in the past 24 hours”). These aggregations summarize the answer options to the question while providing information about the percentage of respondents who did not fully isolate, and also those who were with large groups of people in the same room. In **Fig. S9** we provide a sensitivity analysis with two alternative groupings of the question: answers 4 to 9, and answers 7 to 9. Only around 0.1% of the respondents per wave do not provide an answer to the question.

- In the last week, how closely did you personally follow the health recommendations listed below?

  Avoiding contact with other people, Avoiding public or crowded places, Frequently washing hands, Wearing a face mask when outside of your home. Response options for each: 1.“Not at all closely”, 2.“Not very closely”, 3.“Somewhat closely”, 4.“Very closely”. For the main text analysis, we calculate the percentage of respondents that selected option 4. We also provide a sensitivity analysis with the percentage of respondents who answer options 3 and 4 (**Fig. S9**). Less than 1% of respondents per wave do not provide an answer to the question.

As mentioned in the main text, the Covid States survey has viable samples for most U.S. states in most waves. However, some states tend to have small sample sizes, and our results for these states may be unreliable. The average number of respondents per wave per state is provided in Tab. S4. In addition, we plot in (**Fig. S10**) the average sample size across the 19 survey waves for each state, against the average 95% error margin across the 19 waves of the estimates of the 15 behaviors we use in the main text. This plot helps identify the states that typically generate the most problematic estimates. In particular, the results for a set of 6 states (Alaska, Wyoming, North and South Dakota, New Mexico, and Vermont) plus the District of Columbia, should be taken with caution. While less problematic, the states of Rhode Island, Montana, and Hawaii, also have relatively large error margins or sample sizes, and we also exclude them from the state-level analysis.

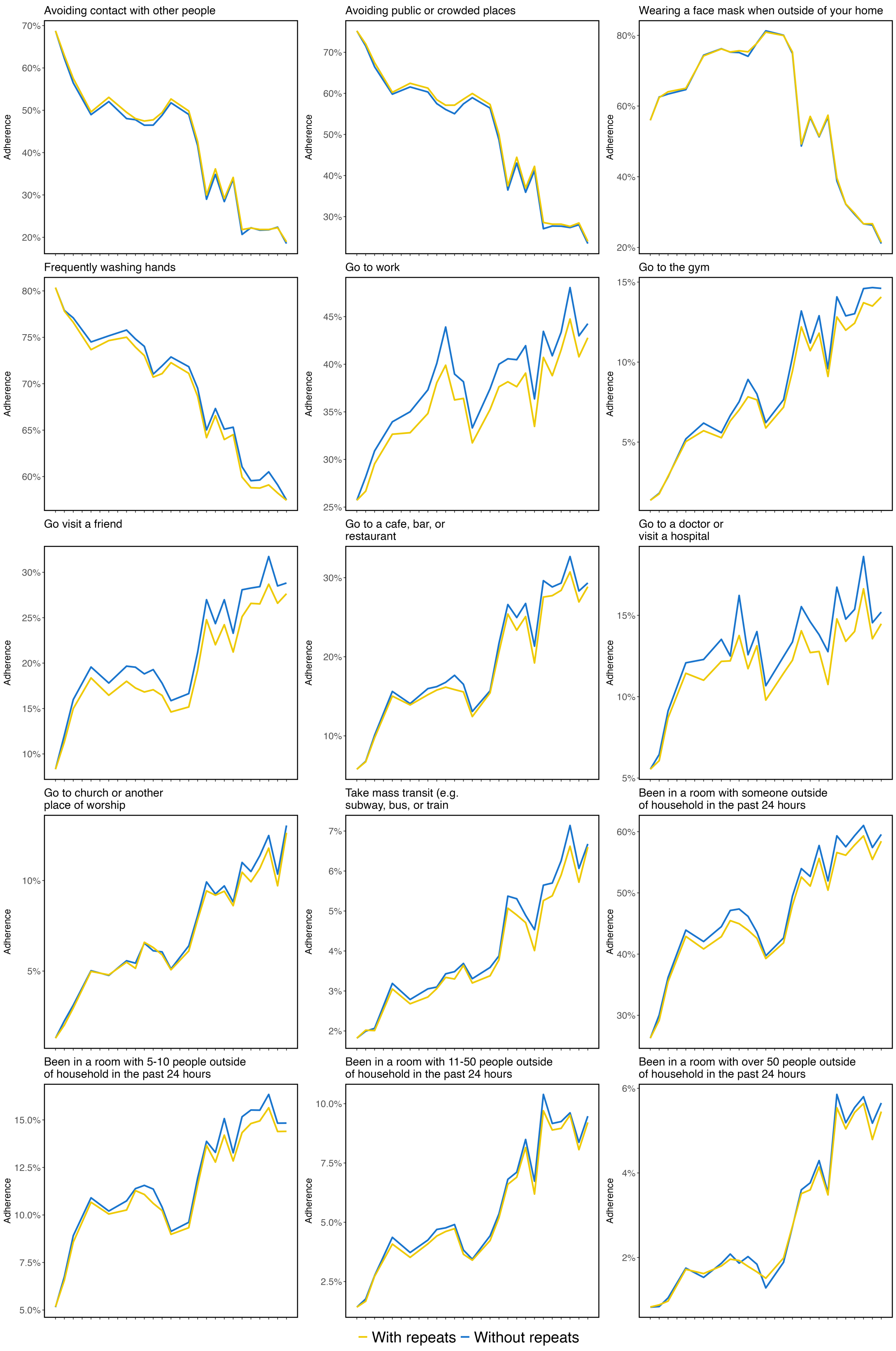

Figure S8: **Responses from repeat participants do not change the trajectory of behavior trends over time**. The 15 risk-averting and risk-exposing as a function of time when repeated participants of the survey are included (gold) and excluded (blue). The main analyses in the study were performed with responses from repeat participants included in the dataset. See Section S5 for details.

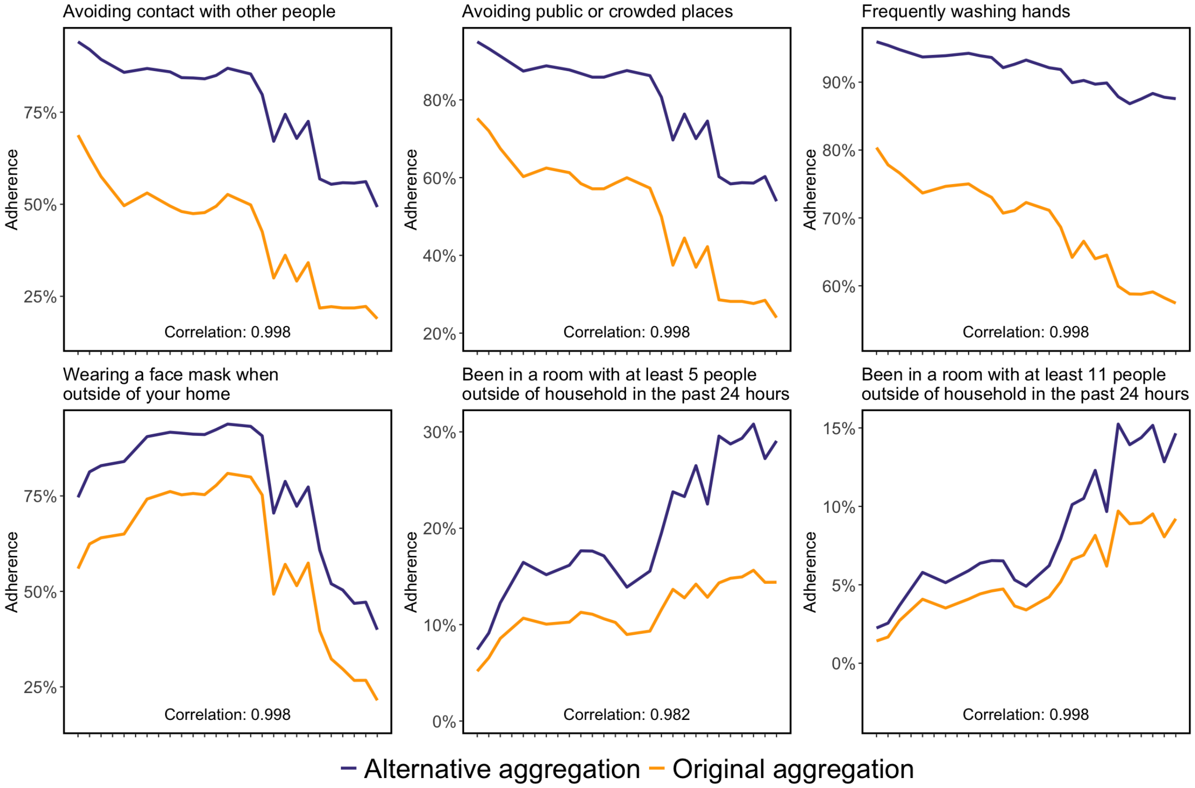

Figure S9: **Changing the aggregation rules for risk-averting and risk-exposing behaviors does not affect the trajectories of the trends.** For the shown risk-averting behaviors, survey responses of "somewhat likely" and "very likely" were included (purple) while the original aggregation only contained "very likely" responses (yellow). For the shown risk-exposing behaviors, the alternative aggregation includes who have been in a room with "at least 5" or "at least 11" people outside of one's household in the past 24 hours while the original aggregation used the categories "5-11 people" and "11-50 people," respectively. The parallel trajectories of the trend lines and the high Pearson correlations between the trends suggest that different aggregations would not substantially influence the trend composition. See Section S5 for details.

| State | W1 | W2 | W3 | W5 | W7 | W9 | W10 | W11 | W12 | W13 | W14 | W16 | W17 | W18 | W19 | W20 | W21 | W22 | W23 |
|---|---|---|---|---|---|---|---|---|---|---|---|---|---|---|---|---|---|---|---|
| AK | 103 | 172 | 128 | 168 | 129 | 126 | 187 | 115 | 138 | 226 | 419 | 313 | 225 | 239 | 247 | 257 | 332 | 289 | 242 |
| AL | 464 | 435 | 394 | 415 | 439 | 523 | 456 | 365 | 242 | 461 | 464 | 397 | 461 | 416 | 438 | 411 | 425 | 441 | 464 |
| AR | 458 | 406 | 332 | 393 | 393 | 390 | 290 | 214 | 319 | 487 | 489 | 519 | 482 | 398 | 509 | 349 | 409 | 378 | 411 |
| AZ | 482 | 379 | 525 | 613 | 351 | 466 | 522 | 339 | 265 | 528 | 477 | 419 | 433 | 455 | 514 | 517 | 468 | 493 | 547 |
| CA | 809 | 974 | 992 | 870 | 557 | 816 | 1178 | 725 | 380 | 787 | 608 | 582 | 807 | 771 | 1038 | 1345 | 1105 | 944 | 1026 |
| CO | 469 | 442 | 431 | 495 | 376 | 479 | 419 | 222 | 339 | 494 | 528 | 505 | 479 | 463 | 457 | 522 | 509 | 465 | 473 |
| CT | 348 | 399 | 262 | 423 | 405 | 415 | 433 | 261 | 280 | 429 | 505 | 436 | 423 | 427 | 369 | 340 | 405 | 426 | 462 |
| DC | 125 | 207 | 148 | 306 | 282 | 226 | 269 | 135 | 160 | 250 | 433 | 347 | 354 | 223 | 298 | 278 | 348 | 248 | 338 |
| DE | 335 | 360 | 416 | 355 | 284 | 284 | 321 | 241 | 238 | 311 | 549 | 411 | 353 | 372 | 392 | 372 | 465 | 416 | 380 |
| FL | 615 | 851 | 697 | 860 | 497 | 594 | 871 | 694 | 312 | 726 | 558 | 564 | 806 | 704 | 682 | 940 | 699 | 578 | 609 |
| GA | 513 | 618 | 479 | 507 | 440 | 607 | 565 | 409 | 248 | 606 | 492 | 491 | 527 | 454 | 518 | 666 | 602 | 500 | 554 |
| HI | 277 | 245 | 317 | 497 | 294 | 260 | 347 | 212 | 222 | 277 | 618 | 389 | 405 | 329 | 402 | 389 | 481 | 431 | 400 |
| IA | 466 | 445 | 263 | 413 | 410 | 256 | 326 | 323 | 337 | 495 | 442 | 429 | 395 | 423 | 407 | 356 | 429 | 414 | 437 |
| ID | 348 | 397 | 402 | 362 | 357 | 272 | 372 | 288 | 252 | 484 | 594 | 494 | 394 | 400 | 466 | 415 | 460 | 416 | 428 |
| IL | 517 | 656 | 580 | 554 | 473 | 509 | 619 | 544 | 280 | 573 | 541 | 506 | 557 | 545 | 610 | 666 | 640 | 598 | 659 |
| IN | 490 | 505 | 416 | 496 | 321 | 466 | 454 | 460 | 231 | 444 | 485 | 439 | 454 | 412 | 418 | 454 | 493 | 471 | 498 |
| KS | 473 | 516 | 259 | 377 | 367 | 441 | 426 | 308 | 211 | 421 | 490 | 434 | 406 | 422 | 380 | 365 | 407 | 382 | 420 |
| KY | 457 | 440 | 359 | 440 | 394 | 517 | 348 | 456 | 322 | 559 | 446 | 433 | 393 | 433 | 427 | 428 | 448 | 448 | 476 |
| LA | 467 | 444 | 409 | 404 | 411 | 445 | 330 | 266 | 315 | 499 | 490 | 409 | 384 | 465 | 351 | 360 | 413 | 380 | 430 |
| MA | 487 | 542 | 483 | 478 | 381 | 605 | 484 | 339 | 290 | 469 | 516 | 485 | 474 | 385 | 512 | 475 | 496 | 497 | 498 |
| MD | 403 | 473 | 438 | 469 | 471 | 544 | 520 | 397 | 257 | 382 | 514 | 454 | 438 | 443 | 478 | 451 | 481 | 449 | 530 |
| ME | 340 | 396 | 410 | 380 | 316 | 285 | 314 | 173 | 287 | 484 | 626 | 509 | 518 | 430 | 470 | 429 | 445 | 432 | 407 |
| MI | 524 | 622 | 514 | 469 | 371 | 584 | 428 | 448 | 319 | 592 | 507 | 502 | 481 | 465 | 562 | 559 | 575 | 585 | 622 |
| MN | 474 | 477 | 371 | 497 | 388 | 535 | 364 | 380 | 371 | 596 | 525 | 450 | 435 | 464 | 452 | 437 | 475 | 475 | 520 |
| MO | 461 | 528 | 495 | 485 | 357 | 511 | 539 | 502 | 284 | 537 | 494 | 439 | 456 | 479 | 488 | 481 | 522 | 492 | 508 |
| MS | 436 | 424 | 251 | 349 | 388 | 403 | 388 | 436 | 311 | 567 | 517 | 465 | 463 | 367 | 403 | 356 | 369 | 406 | 404 |
| MT | 230 | 283 | 232 | 295 | 291 | 211 | 279 | 158 | 225 | 319 | 562 | 469 | 401 | 355 | 379 | 364 | 428 | 386 | 317 |
| NC | 511 | 606 | 557 | 562 | 446 | 516 | 559 | 561 | 288 | 569 | 533 | 513 | 526 | 495 | 514 | 590 | 619 | 525 | 623 |
| ND | 176 | 236 | 236 | 199 | 214 | 194 | 253 | 155 | 128 | 250 | 517 | 373 | 336 | 290 | 342 | 356 | 420 | 334 | 358 |
| NE | 361 | 298 | 177 | 384 | 336 | 306 | 322 | 358 | 274 | 497 | 601 | 508 | 529 | 380 | 444 | 337 | 398 | 411 | 421 |
| NH | 374 | 397 | 377 | 375 | 317 | 204 | 336 | 298 | 236 | 468 | 574 | 400 | 535 | 385 | 396 | 317 | 341 | 379 | 386 |
| NJ | 539 | 637 | 590 | 505 | 395 | 522 | 510 | 464 | 254 | 473 | 460 | 468 | 487 | 435 | 468 | 605 | 533 | 489 | 525 |
| NM | 135 | 236 | 172 | 324 | 352 | 319 | 386 | 282 | 274 | 450 | 493 | 349 | 478 | 371 | 431 | 360 | 447 | 383 | 448 |
| NV | 472 | 459 | 300 | 394 | 433 | 497 | 333 | 309 | 340 | 585 | 497 | 422 | 362 | 483 | 445 | 372 | 446 | 406 | 445 |
| NY | 638 | 839 | 734 | 658 | 474 | 627 | 804 | 656 | 339 | 749 | 539 | 628 | 763 | 693 | 914 | 1045 | 1044 | 775 | 951 |
| OH | 502 | 620 | 528 | 584 | 484 | 522 | 603 | 602 | 297 | 567 | 511 | 559 | 567 | 564 | 579 | 669 | 561 | 540 | 560 |
| OK | 458 | 392 | 327 | 367 | 391 | 497 | 376 | 309 | 315 | 571 | 518 | 455 | 412 | 465 | 445 | 379 | 444 | 414 | 435 |
| OR | 474 | 460 | 451 | 458 | 403 | 477 | 405 | 318 | 291 | 513 | 577 | 477 | 391 | 480 | 494 | 403 | 455 | 470 | 489 |
| PA | 523 | 649 | 593 | 604 | 478 | 519 | 677 | 789 | 290 | 578 | 498 | 533 | 617 | 489 | 537 | 678 | 615 | 538 | 586 |
| RI | 313 | 342 | 195 | 329 | 349 | 256 | 317 | 151 | 178 | 345 | 550 | 399 | 376 | 294 | 328 | 336 | 435 | 397 | 364 |
| SC | 477 | 486 | 393 | 475 | 427 | 456 | 381 | 443 | 368 | 572 | 447 | 421 | 430 | 490 | 412 | 430 | 444 | 411 | 470 |
| SD | 192 | 257 | 197 | 311 | 270 | 191 | 326 | 214 | 163 | 271 | 509 | 463 | 355 | 314 | 323 | 281 | 419 | 380 | 278 |
| TN | 471 | 519 | 428 | 486 | 478 | 514 | 557 | 478 | 235 | 460 | 521 | 485 | 460 | 476 | 461 | 479 | 493 | 505 | 510 |
| TX | 611 | 751 | 721 | 881 | 490 | 646 | 1117 | 621 | 425 | 732 | 563 | 603 | 839 | 769 | 754 | 1038 | 791 | 633 | 652 |
| UT | 436 | 354 | 314 | 357 | 455 | 278 | 434 | 182 | 298 | 406 | 490 | 432 | 418 | 381 | 436 | 378 | 411 | 401 | 430 |
| VA | 522 | 621 | 510 | 570 | 409 | 540 | 522 | 398 | 277 | 487 | 464 | 465 | 456 | 464 | 526 | 623 | 522 | 507 | 557 |
| VT | 188 | 273 | 181 | 240 | 215 | 136 | 179 | 134 | 117 | 188 | 429 | 336 | 301 | 246 | 246 | 285 | 355 | 306 | 272 |
| WA | 397 | 526 | 551 | 449 | 454 | 496 | 553 | 465 | 246 | 535 | 555 | 489 | 442 | 493 | 549 | 531 | 521 | 498 | 558 |
| WI | 473 | 620 | 460 | 486 | 447 | 449 | 482 | 434 | 259 | 739 | 516 | 480 | 478 | 480 | 498 | 463 | 465 | 495 | 518 |
| WV | 369 | 368 | 217 | 332 | 363 | 371 | 335 | 403 | 225 | 437 | 451 | 419 | 409 | 342 | 420 | 353 | 402 | 388 | 407 |
| WY | 165 | 167 | 115 | 148 | 155 | 143 | 178 | 115 | 121 | 179 | 355 | 330 | 299 | 205 | 251 | 275 | 390 | 294 | 270 |

Table S4: **The average number of responses per wave per state.** In the first column, the states and District of Columbia (DC) are given while the waves considered in this study are listed as W1, W2, etc.

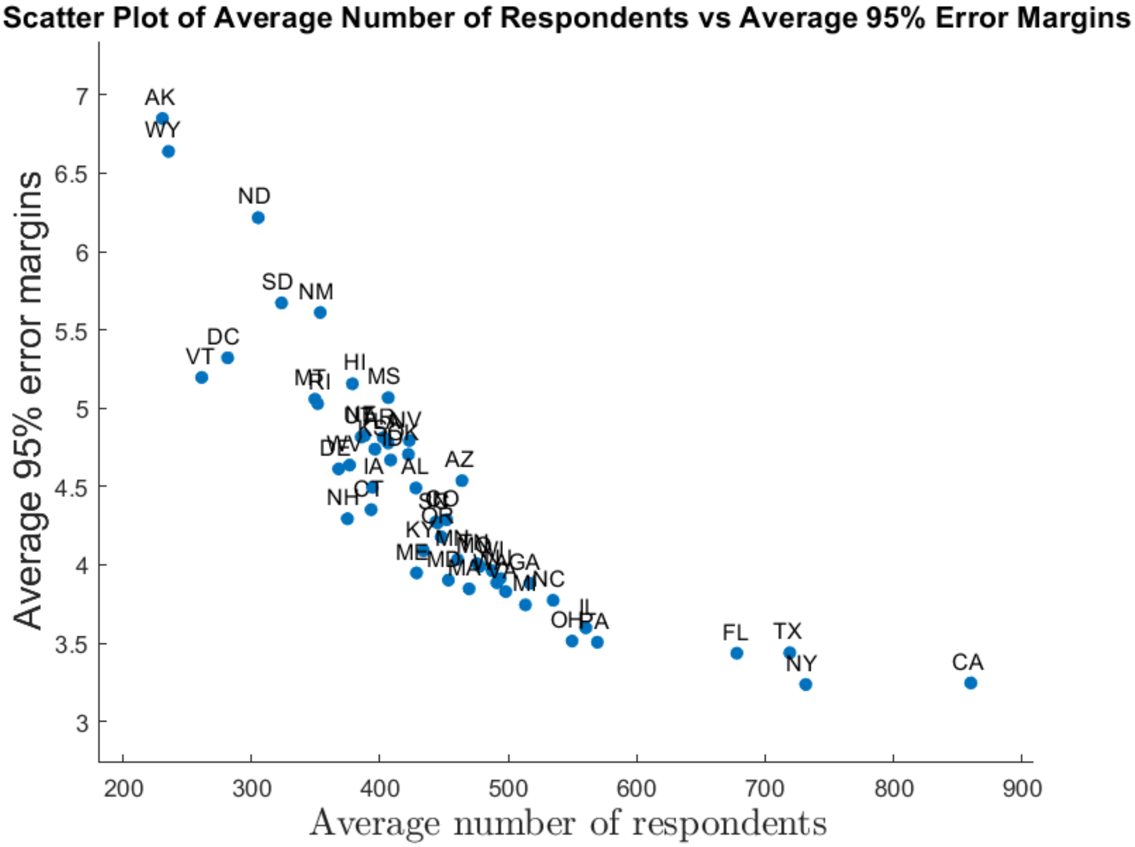

Figure S10: **Scatter plot of the average number of respondents vs average 95% error margins per state.**

### 6 Principal component analysis (PCA)

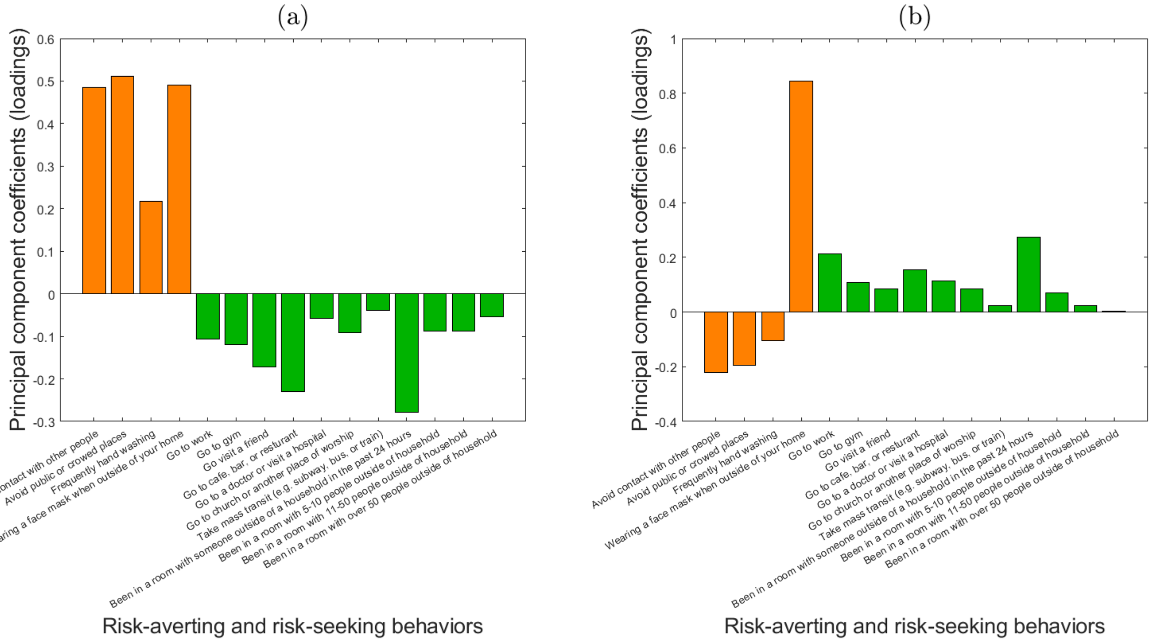

Figure S11: **The principal component coefficients (loadings) of each risk-exposing and risk-averting behavior towards the (a) first and (b) second principal components obtained using principal component analysis (PCA).** The value of loadings associated with risk-averting and risk-exposing behaviors are depicted with orange and green bars, respectively.

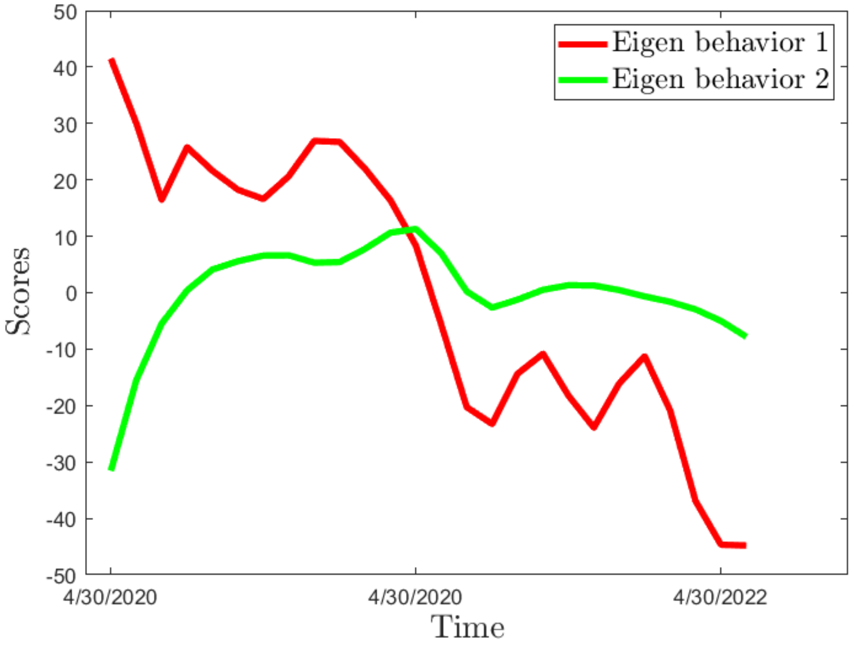

Figure S12: **The first and second eigen behaviors as a function of time.**

### 7 Correlation between behaviors

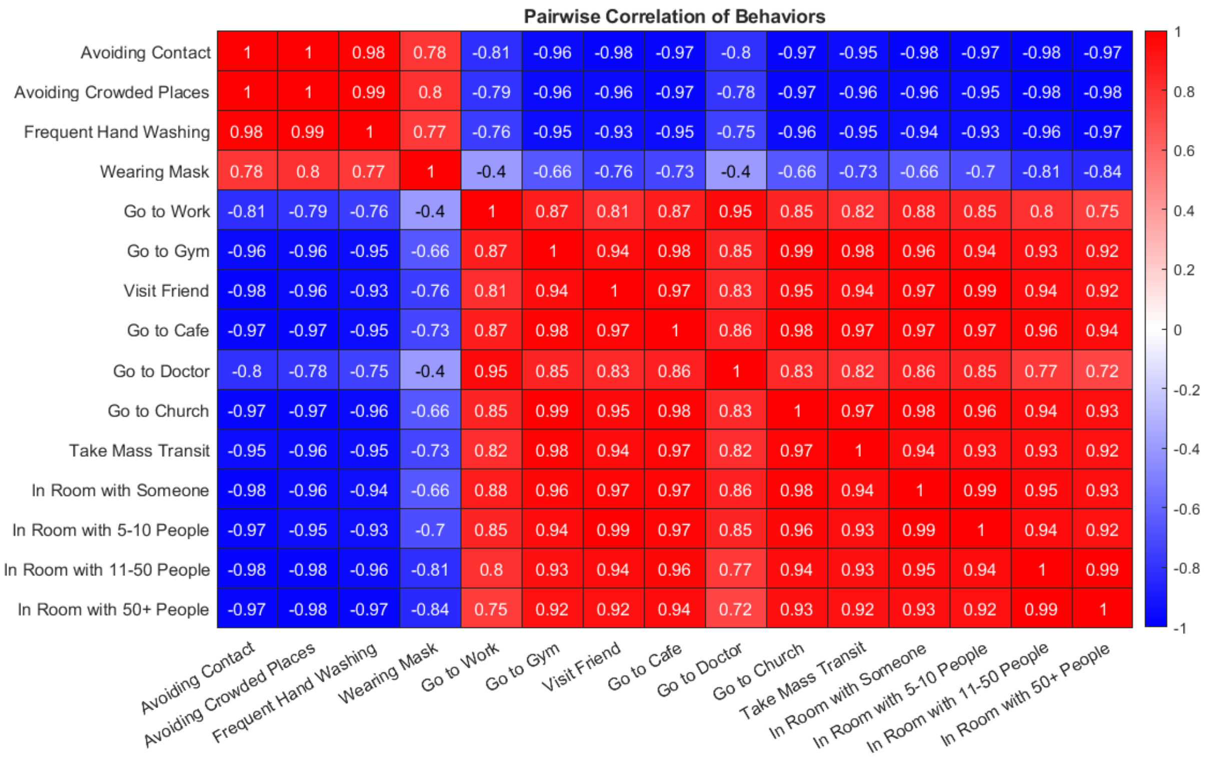

Figure S13: **The 15 risk-averting and risk-exposing behaviors are, on average, highly correlated.** The pairwise correlation of all behaviors considered in this study. Highly positively and highly negatively correlated pairs are colored denoted with dark red and dark blue colors, respectively. The behaviors with the highest and lowest average correlation was "avoiding contact" and "wearing a mask", respectively (with a respective 0.94 and 0.69 average correlation). The overall average correlation across all behaviors was 0.89. The absolute value of the correlation was used in the calculation of average correlation and self-correlation of behavior was not included.

### 7 State-level behavioral trends and their relationships with epidemic severity

**State name: Alabama**

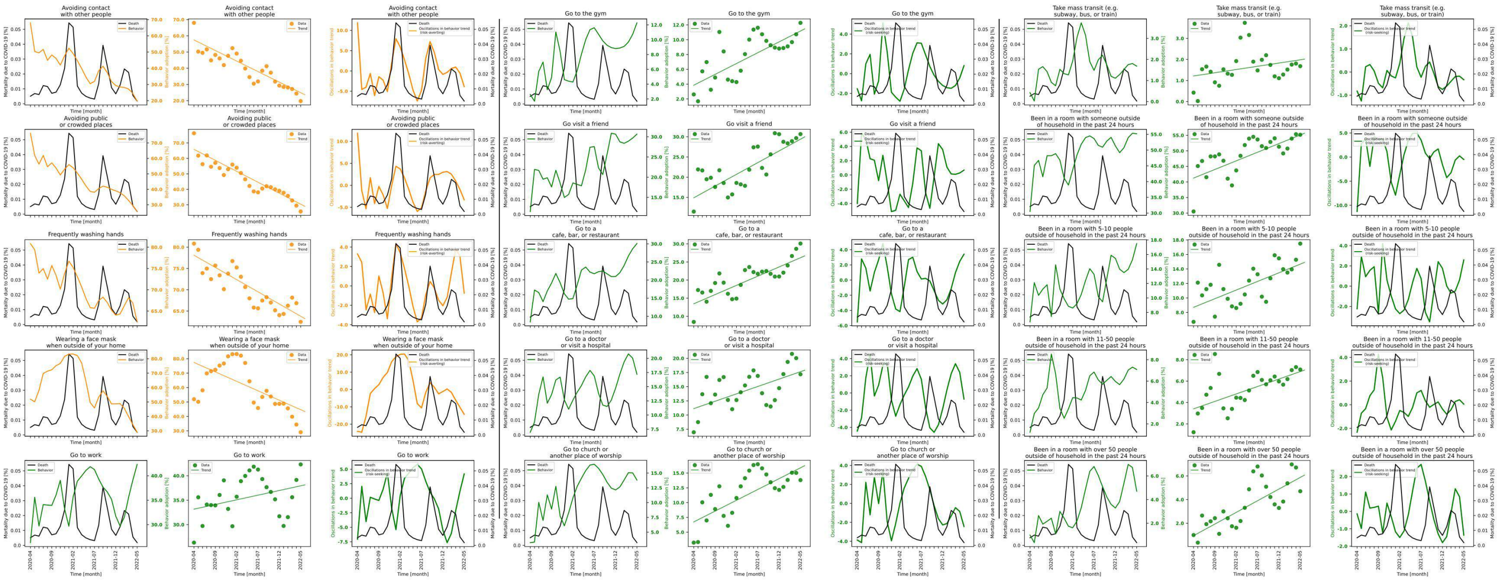

**Figure S7.1 shows behavior and mortality on 1st, 4th and 7th column, behavior trends in 2nd, 5th and 8th column, and behavior oscillatory components juxtaposed with mortality rates on 3rd, 6th and 9th columns for Alabama.**

| Behaviors | Slopes | Y-intercepts | Start %$(S)$ | End %$(E)$ | $\|E-S\|$ | $\frac{\|E-S\|*100}{S}$ |
|---|---|---|---|---|---|---|
| Avoiding contact with other people | -1.3 | 57.439 | 67.994 | 19.773 | 48.221 | 70.919 |
| Avoiding public or crowded places | -1.418 | 65.849 | 76.275 | 25.678 | 50.597 | 66.335 |
| Frequently washing hands | -0.573 | 78.113 | 80.838 | 62.472 | 18.366 | 22.72 |
| Wearing a face mask when outside of your home | -1.311 | 77.421 | 51.939 | 29.031 | 22.908 | 44.106 |
| Go to work | 0.191 | 33.165 | 26.314 | 42.322 | 16.008 | 60.835 |
| Go to the gym | 0.293 | 3.857 | 2.611 | 12.278 | 9.667 | 370.241 |
| Go visit a friend | 0.58 | 14.899 | 11.484 | 30.663 | 19.179 | 167.006 |
| Go to a cafe, bar, or restaurant | 0.509 | 13.452 | 8.442 | 30.088 | 21.646 | 256.408 |
| Go to a doctor or visit a hospital | 0.26 | 11.131 | 6.961 | 17.185 | 10.224 | 146.875 |
| Go to church or another place of worship | 0.366 | 6.734 | 3.23 | 13.81 | 10.58 | 327.554 |
| Take mass transit (e.g. subway, bus, or train) | 0.031 | 1.214 | 0.422 | 1.681 | 1.259 | 298.341 |
| Been in a room with someone outside of household in the past 24 hours | 0.554 | 41.249 | 30.52 | 55.169 | 24.649 | 80.763 |
| Been in a room with 5-10 people outside of household in the past 24 hours | 0.238 | 8.706 | 6.704 | 17.492 | 10.788 | 160.919 |
| Been in a room with 11-50 people outside of household in the past 24 hours | 0.139 | 3.407 | 1.22 | 7.082 | 5.862 | 480.492 |
| Been in a room with over 50 people outside of household in the past 24 hours | 0.197 | 0.946 | 0.999 | 4.707 | 3.708 | 371.171 |

**Table S7.1 shows the slopes, y-intercept, adherence % at the beginning of the study period, and at the end, the absolute difference between the beginning and the end, and the absolute difference as a percentage of the adherence % at the beginning of the study period of each behavior's trend in Alabama.**

### State name: Alaska

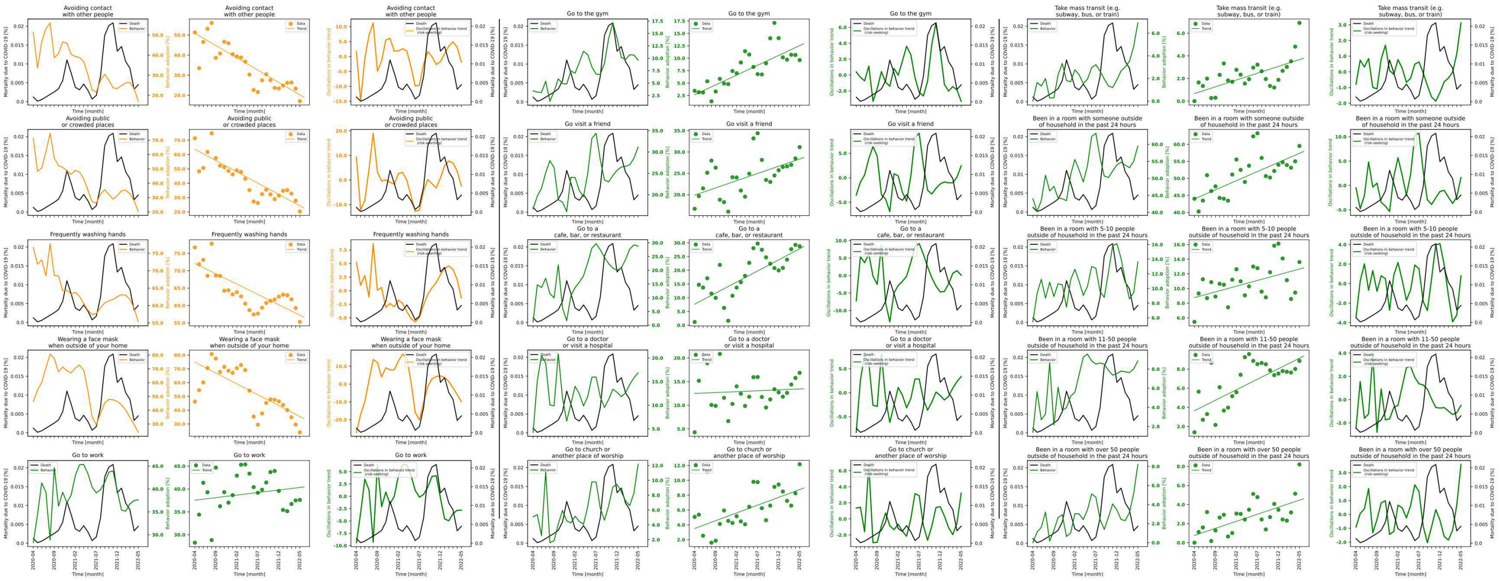

**Figure S7.2 shows behavior and mortality on 1st, 4th and 7th column, behavior trends in 2nd, 5th and 8th column, and behavior oscillatory components juxtaposed with mortality rates on 3rd, 6th and 9th columns for Alaska.**

| Behaviors | Slopes | Y-intercepts | Start %($S$) | End %($E$) | $\lvert E-S \rvert$ | $\frac{\lvert E-S \rvert *100}{S}$ |
|---|---|---|---|---|---|---|
| Avoiding contact with other people | -1.227 | 50.899 | 51.308 | 17.116 | 34.192 | 66.641 |
| Avoiding public or crowded places | -1.576 | 63.686 | 71.374 | 20.307 | 51.067 | 71.548 |
| Frequently washing hands | -0.594 | 72.089 | 76.707 | 55.281 | 21.426 | 27.932 |
| Wearing a face mask when outside of your home | -1.574 | 74.801 | 46.175 | 23.952 | 22.223 | 48.128 |
| Go to work | 0.111 | 37.55 | 28.259 | 37.61 | 9.351 | 33.09 |
| Go to the gym | 0.395 | 2.656 | 3.439 | 9.628 | 6.189 | 179.965 |
| Go visit a friend | 0.338 | 19.895 | 16.688 | 31.142 | 14.454 | 86.613 |
| Go to a cafe, bar, or restaurant | 0.808 | 7.644 | 1.177 | 28.764 | 27.587 | 2343.84 |
| Go to a doctor or visit a hospital | 0.038 | 12.497 | 4.275 | 16.854 | 12.579 | 294.246 |
| Go to church or another place of worship | 0.21 | 3.509 | 5.073 | 12.165 | 7.092 | 139.799 |
| Take mass transit (e.g. subway, bus, or train) | 0.12 | 0.664 | 0.0 | 6.911 | 6.911 | inf |
| Been in a room with someone outside of household in the past 24 hours | 0.536 | 43.97 | 44.028 | 59.523 | 15.495 | 35.194 |
| Been in a room with 5-10 people outside of household in the past 24 hours | 0.156 | 8.763 | 5.446 | 13.614 | 8.168 | 149.982 |
| Been in a room with 11-50 people outside of household in the past 24 hours | 0.22 | 3.647 | 1.391 | 8.861 | 7.47 | 537.024 |
| Been in a room with over 50 people outside of household in the past 24 hours | 0.148 | 0.762 | 0.0 | 8.207 | 8.207 | inf |

**Table S7.2 shows the slopes, y-intercept, adherence % at the beginning of the study period, and at the end, the absolute difference between the beginning and the end, and the absolute difference as a percentage of the adherence % at the beginning of the study period of each behavior's trend in Alaska.**

### State name: Arizona

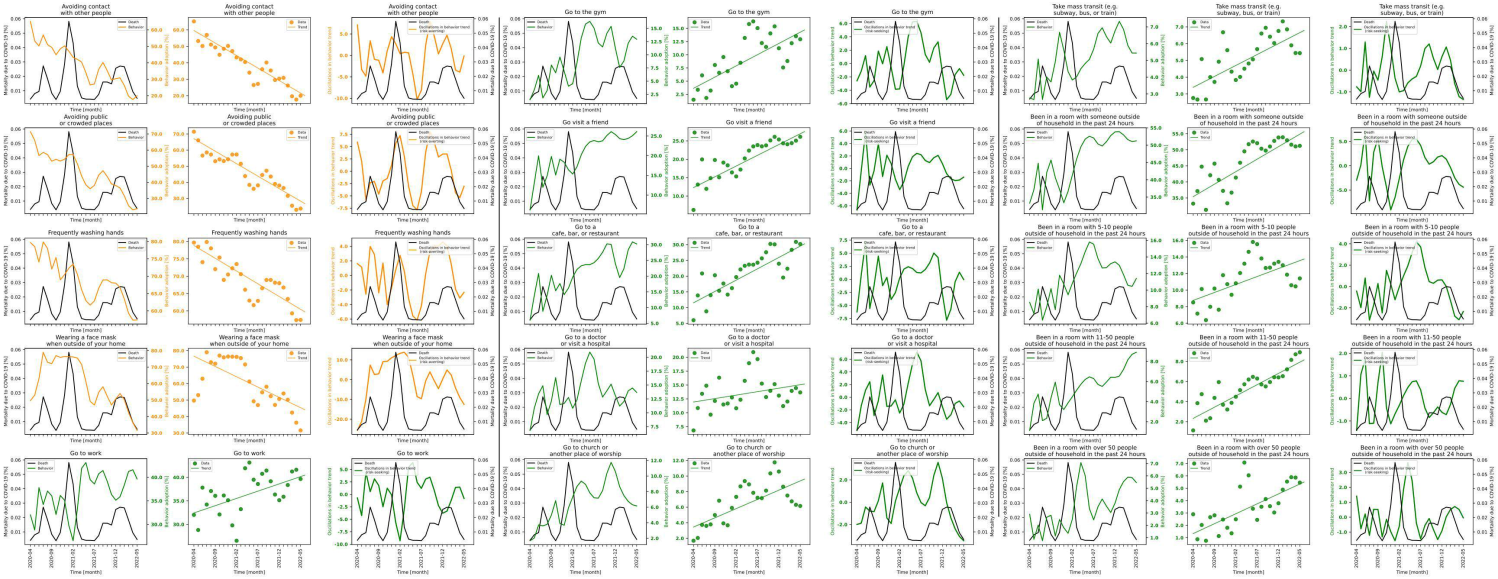

**Figure S7.3 shows behavior and mortality on 1st, 4th and 7th column, behavior trends in 2nd, 5th and 8th column, and behavior oscillatory components juxtaposed with mortality rates on 3rd, 6th and 9th columns for Arizona.**

| Behaviors | Slopes | Y-intercepts | Start %($S$) | End %($E$) | $\|E-S\|$ | $\frac{\|E-S\|*100}{S}$ |
|---|---|---|---|---|---|---|
| Avoiding contact with other people | -1.521 | 59.615 | 65.181 | 19.955 | 45.226 | 69.385 |
| Avoiding public or crowded places | -1.551 | 67.059 | 71.422 | 23.691 | 47.731 | 66.83 |
| Frequently washing hands | -0.735 | 78.837 | 79.703 | 57.404 | 22.299 | 27.978 |
| Wearing a face mask when outside of your home | -1.251 | 76.641 | 49.698 | 31.58 | 18.118 | 36.456 |
| Go to work | 0.309 | 32.438 | 32.025 | 39.65 | 7.625 | 23.81 |
| Go to the gym | 0.426 | 3.638 | 1.477 | 12.948 | 11.471 | 776.642 |
| Go visit a friend | 0.585 | 12.303 | 6.215 | 26.111 | 19.896 | 320.129 |
| Go to a cafe, bar, or restaurant | 0.721 | 11.529 | 5.997 | 30.214 | 24.217 | 403.819 |
| Go to a doctor or visit a hospital | 0.125 | 11.937 | 6.815 | 13.673 | 6.858 | 100.631 |
| Go to church or another place of worship | 0.237 | 3.448 | 1.693 | 6.16 | 4.467 | 263.851 |
| Take mass transit (e.g. subway, bus, or train) | 0.131 | 3.396 | 2.759 | 5.438 | 2.679 | 97.1 |
| Been in a room with someone outside of household in the past 24 hours | 0.778 | 35.413 | 33.247 | 51.25 | 18.003 | 54.149 |
| Been in a room with 5-10 people outside of household in the past 24 hours | 0.192 | 8.754 | 8.512 | 11.41 | 2.898 | 34.046 |
| Been in a room with 11-50 people outside of household in the past 24 hours | 0.224 | 2.333 | 1.139 | 8.914 | 7.775 | 682.616 |
| Been in a room with over 50 people outside of household in the past 24 hours | 0.16 | 1.334 | 2.899 | 5.461 | 2.562 | 88.375 |

**Table S7.3 shows the slopes, y-intercept, adherence % at the beginning of the study period, and at the end, the absolute difference between the beginning and the end, and the absolute difference as a percentage of the adherence % at the beginning of the study period of each behavior's trend in Arizona.**

### State name: Arkansas

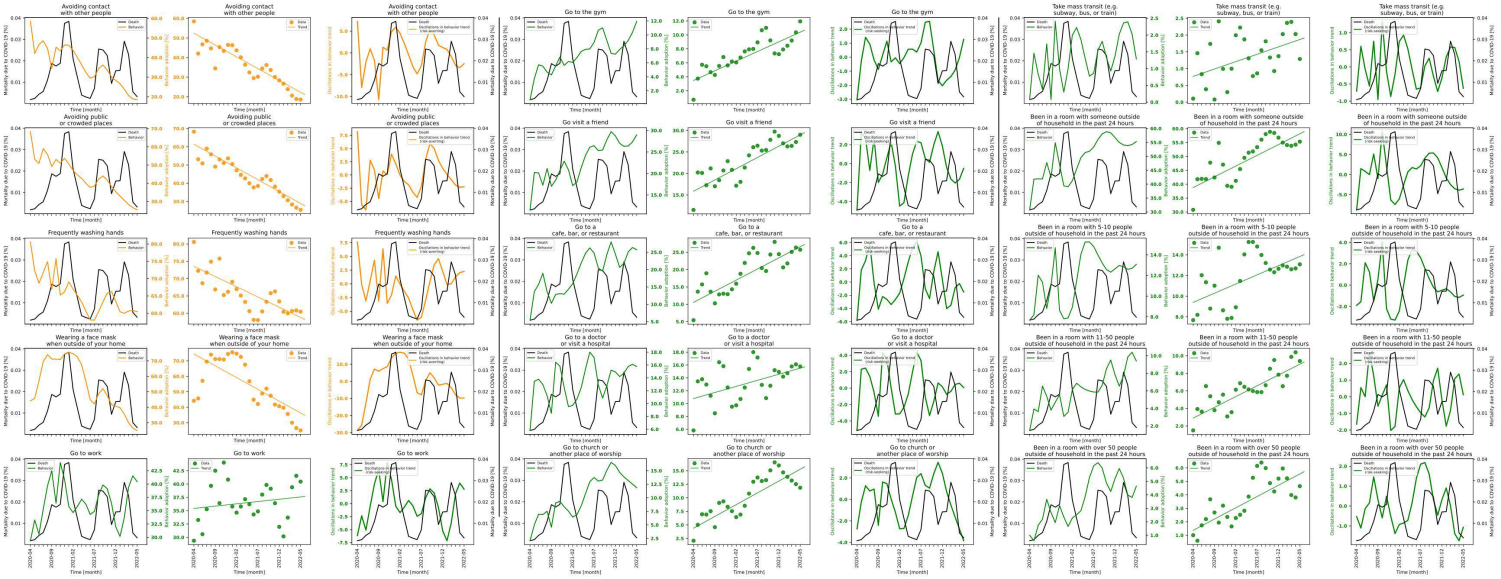

**Figure S7.4 shows behavior and mortality on 1st, 4th and 7th column, behavior trends in 2nd, 5th and 8th column, and behavior oscillatory components juxtaposed with mortality rates on 3rd, 6th and 9th columns for Arkansas.**

| Behaviors | Slopes | Y-intercepts | Start %$(S)$ | End %$(E)$ | $\lvert E-S \rvert$ | $\frac{\lvert E-S \rvert * 100}{S}$ |
|---|---|---|---|---|---|---|
| Avoiding contact with other people | -1.207 | 52.451 | 58.483 | 18.584 | 39.899 | 68.223 |
| Avoiding public or crowded places | -1.299 | 61.426 | 68.324 | 25.421 | 42.903 | 62.793 |
| Frequently washing hands | -0.591 | 73.557 | 80.59 | 60.445 | 20.145 | 24.997 |
| Wearing a face mask when outside of your home | -1.525 | 74.44 | 44.193 | 25.01 | 19.183 | 43.407 |
| Go to work | 0.086 | 35.425 | 29.37 | 40.448 | 11.078 | 37.719 |
| Go to the gym | 0.276 | 3.479 | 0.721 | 11.936 | 11.215 | 1555.479 |
| Go visit a friend | 0.524 | 15.875 | 11.516 | 29.003 | 17.487 | 151.85 |
| Go to a cafe, bar, or restaurant | 0.642 | 10.552 | 5.407 | 25.733 | 20.326 | 375.92 |
| Go to a doctor or visit a hospital | 0.189 | 10.781 | 5.814 | 15.775 | 9.961 | 171.328 |
| Go to church or another place of worship | 0.435 | 4.384 | 2.082 | 11.856 | 9.774 | 469.452 |
| Take mass transit (e.g. subway, bus, or train) | 0.044 | 0.769 | 0.106 | 1.286 | 1.18 | 1113.208 |
| Been in a room with someone outside of household in the past 24 hours | 0.774 | 38.793 | 30.78 | 55.322 | 24.542 | 79.734 |
| Been in a room with 5-10 people outside of household in the past 24 hours | 0.178 | 9.399 | 7.685 | 13.079 | 5.394 | 70.189 |
| Been in a room with 11-50 people outside of household in the past 24 hours | 0.246 | 2.892 | 1.491 | 9.397 | 7.906 | 530.248 |
| Been in a room with over 50 people outside of household in the past 24 hours | 0.167 | 1.368 | 1.003 | 4.646 | 3.643 | 363.21 |

**Table S7.4 shows the slopes, y-intercept, adherence % at the beginning of the study period, and at the end, the absolute difference between the beginning and the end, and the absolute difference as a percentage of the adherence % at the beginning of the study period of each behavior's trend in Arkansas.**

#### State name: California

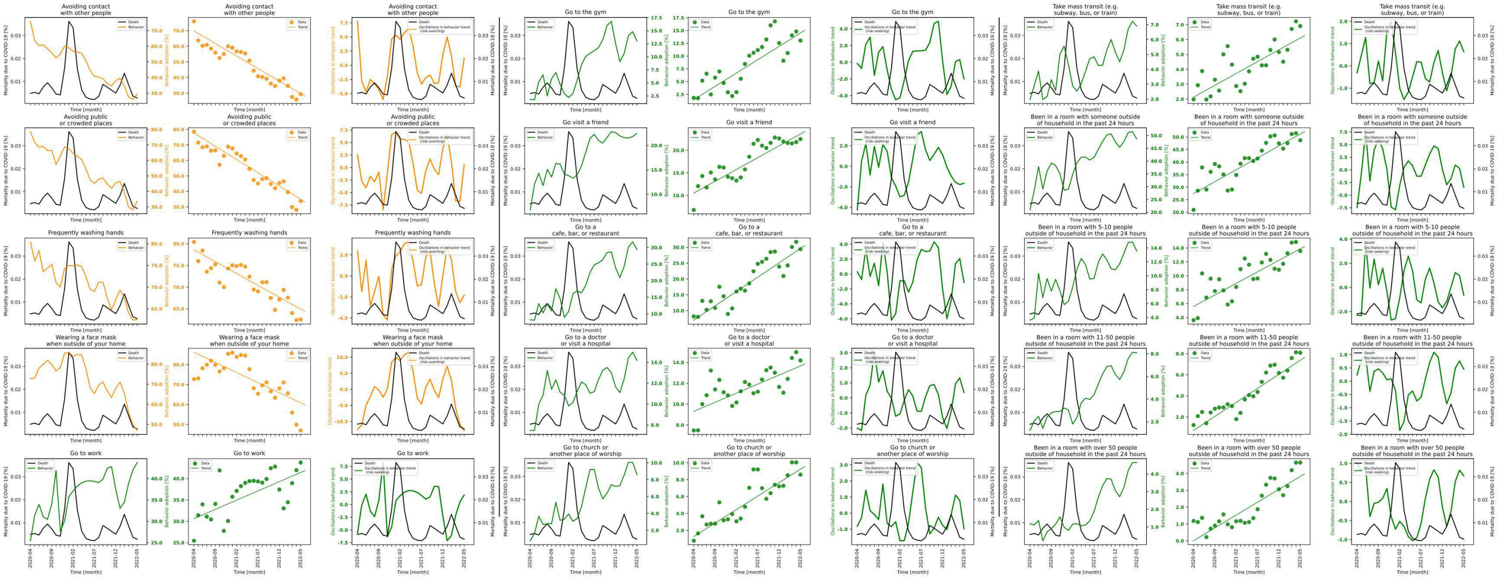

**Figure S7.5 shows behavior and mortality on 1st, 4th and 7th column, behavior trends in 2nd, 5th and 8th column, and behavior oscillatory components juxtaposed with mortality rates on 3rd, 6th and 9th columns for California.**

| Behaviors | Slopes | Y-intercepts | Start %$(S)$ | End %$(E)$ | $\lvert E-S \rvert$ | $\frac{\lvert E-S \rvert * 100}{S}$ |
|---|---|---|---|---|---|---|
| Avoiding contact with other people | -1.602 | 69.768 | 75.907 | 29.387 | 46.52 | 61.286 |
| Avoiding public or crowded places | -1.713 | 77.694 | 78.553 | 33.726 | 44.827 | 57.066 |
| Frequently washing hands | -0.546 | 78.587 | 80.416 | 62.56 | 17.856 | 22.205 |
| Wearing a face mask when outside of your home | -1.015 | 86.196 | 72.726 | 46.997 | 25.729 | 35.378 |
| Go to work | 0.431 | 30.645 | 25.46 | 43.748 | 18.288 | 71.83 |
| Go to the gym | 0.522 | 1.459 | 1.825 | 13.031 | 11.206 | 614.027 |
| Go visit a friend | 0.523 | 10.526 | 6.79 | 22.466 | 15.676 | 230.869 |
| Go to a cafe, bar, or restaurant | 0.906 | 6.982 | 8.177 | 29.407 | 21.23 | 259.631 |
| Go to a doctor or visit a hospital | 0.174 | 9.307 | 7.477 | 14.182 | 6.705 | 89.675 |
| Go to church or another place of worship | 0.319 | 1.269 | 0.769 | 8.596 | 7.827 | 1017.815 |
| Take mass transit (e.g. subway, bus, or train) | 0.159 | 2.129 | 1.98 | 6.91 | 4.93 | 248.99 |
| Been in a room with someone outside of household in the past 24 hours | 0.949 | 27.447 | 20.953 | 48.694 | 27.741 | 132.396 |
| Been in a room with 5-10 people outside of household in the past 24 hours | 0.332 | 5.582 | 3.621 | 13.559 | 9.938 | 274.455 |
| Been in a room with 11-50 people outside of household in the past 24 hours | 0.265 | 0.731 | 1.234 | 8.087 | 6.853 | 555.348 |
| Been in a room with over 50 people outside of household in the past 24 hours | 0.153 | 0.006 | 1.181 | 4.648 | 3.467 | 293.565 |

**Table S7.5 shows the slopes, y-intercept, adherence % at the beginning of the study period, and at the end, the absolute difference between the beginning and the end, and the absolute difference as a percentage of the adherence % at the beginning of the study period of each behavior's trend in California.**

### State name: Colorado

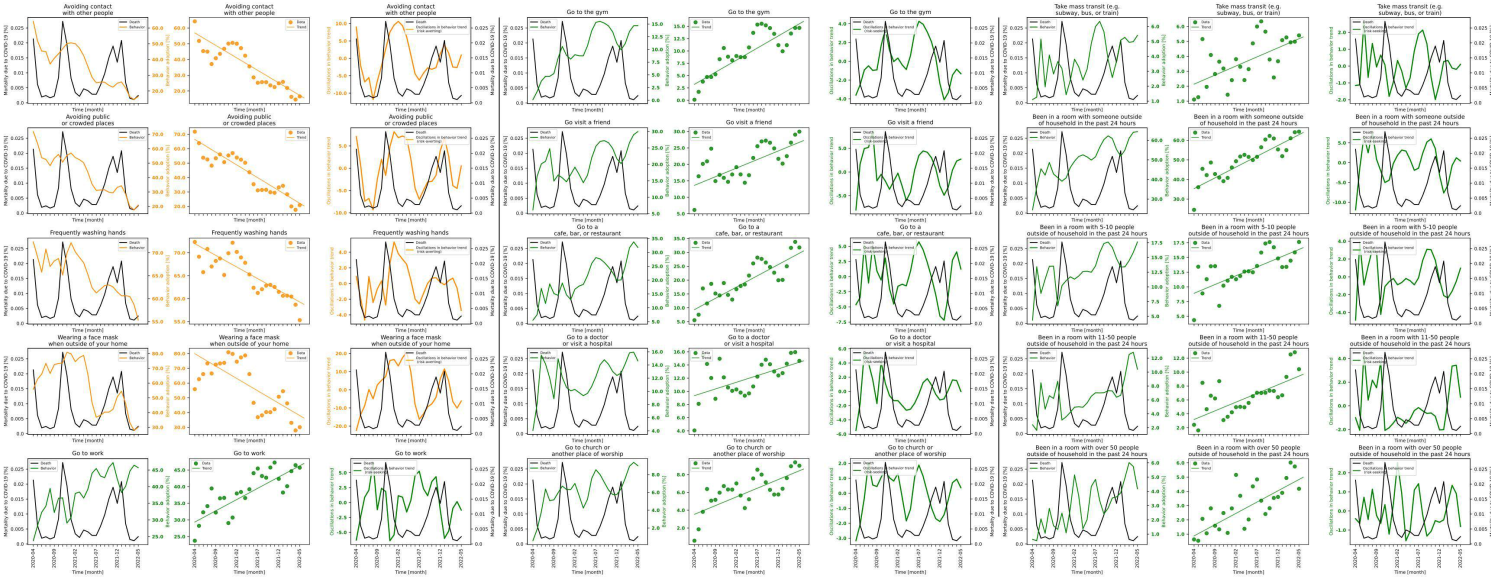

**Figure S7.6 shows behavior and mortality on 1st, 4th and 7th column, behavior trends in 2nd, 5th and 8th column, and behavior oscillatory components juxtaposed with mortality rates on 3rd, 6th and 9th columns for Colorado.**

| Behaviors | Slopes | Y-intercepts | Start %$(S)$ | End %$(E)$ | $\lvert E-S\rvert$ | $\frac{\lvert E-S\rvert*100}{S}$ |
|---|---|---|---|---|---|---|
| Avoiding contact with other people | -1.587 | 56.892 | 64.233 | 16.548 | 47.685 | 74.238 |
| Avoiding public or crowded places | -1.772 | 66.465 | 71.814 | 20.906 | 50.908 | 70.889 |
| Frequently washing hands | -0.508 | 71.982 | 72.363 | 55.337 | 17.026 | 23.529 |
| Wearing a face mask when outside of your home | -1.677 | 79.912 | 55.865 | 30.097 | 25.768 | 46.125 |
| Go to work | 0.676 | 29.373 | 23.761 | 45.7 | 21.939 | 92.332 |
| Go to the gym | 0.491 | 3.229 | 0.113 | 14.673 | 14.56 | 12884.956 |
| Go visit a friend | 0.523 | 13.629 | 6.143 | 29.952 | 23.809 | 387.579 |
| Go to a cafe, bar, or restaurant | 0.814 | 9.349 | 5.537 | 31.772 | 26.235 | 473.813 |
| Go to a doctor or visit a hospital | 0.203 | 9.347 | 4.061 | 14.628 | 10.567 | 260.207 |
| Go to church or another place of worship | 0.196 | 3.539 | 0.571 | 9.003 | 8.432 | 1476.708 |
| Take mass transit (e.g. subway, bus, or train) | 0.122 | 2.138 | 1.108 | 5.401 | 4.293 | 387.455 |
| Been in a room with someone outside of household in the past 24 hours | 1.101 | 35.32 | 24.523 | 64.368 | 39.845 | 162.48 |
| Been in a room with 5-10 people outside of household in the past 24 hours | 0.298 | 8.914 | 4.355 | 17.631 | 13.276 | 304.845 |
| Been in a room with 11-50 people outside of household in the past 24 hours | 0.25 | 3.188 | 2.426 | 10.415 | 7.989 | 329.308 |
| Been in a room with over 50 people outside of household in the past 24 hours | 0.158 | 0.875 | 0.627 | 4.18 | 3.553 | 566.667 |

**Table S7.6 shows the slopes, y-intercept, adherence % at the beginning of the study period, and at the end, the absolute difference between the beginning and the end, and the absolute difference as a percentage of the adherence % at the beginning of the study period of each behavior's trend in Colorado.**

#### State name: Connecticut

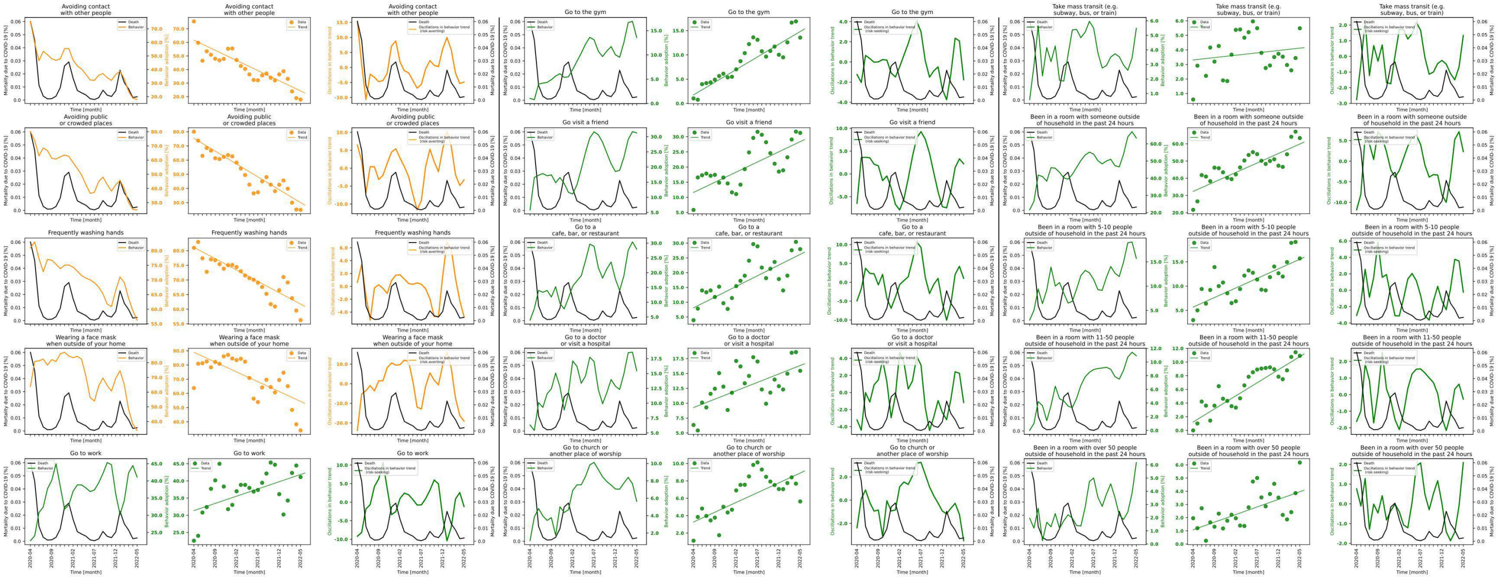

**Figure S7.7 shows behavior and mortality on 1st, 4th and 7th column, behavior trends in 2nd, 5th and 8th column, and behavior oscillatory components juxtaposed with mortality rates on 3rd, 6th and 9th columns for Connecticut.**

| Behaviors | Slopes | Y-intercepts | Start %$(S)$ | End %$(E)$ | $\|E-S\|$ | $\frac{\|E-S\|*100}{S}$ |
|---|---|---|---|---|---|---|
| Avoiding contact with other people | -1.497 | 61.668 | 75.514 | 17.881 | 57.633 | 76.321 |
| Avoiding public or crowded places | -1.815 | 75.391 | 80.054 | 24.961 | 55.093 | 68.82 |
| Frequently washing hands | -0.775 | 81.146 | 80.983 | 56.283 | 24.7 | 30.5 |
| Wearing a face mask when outside of your home | -1.384 | 88.975 | 63.793 | 33.936 | 29.857 | 46.803 |
| Go to work | 0.421 | 31.351 | 22.594 | 41.114 | 18.52 | 81.969 |
| Go to the gym | 0.522 | 1.753 | 1.028 | 13.565 | 12.537 | 1219.553 |
| Go visit a friend | 0.67 | 11.616 | 5.824 | 31.268 | 25.444 | 436.882 |
| Go to a cafe, bar, or restaurant | 0.724 | 8.105 | 3.909 | 27.97 | 24.061 | 615.528 |
| Go to a doctor or visit a hospital | 0.283 | 9.272 | 6.339 | 15.456 | 9.117 | 143.824 |
| Go to church or another place of worship | 0.225 | 3.269 | 1.12 | 5.634 | 4.514 | 403.036 |
| Take mass transit (e.g. subway, bus, or train) | 0.033 | 3.307 | 0.586 | 5.487 | 4.901 | 836.348 |
| Been in a room with someone outside of household in the past 24 hours | 1.102 | 32.347 | 21.641 | 63.361 | 41.72 | 192.782 |
| Been in a room with 5-10 people outside of household in the past 24 hours | 0.395 | 5.667 | 2.987 | 15.706 | 12.719 | 425.812 |
| Been in a room with 11-50 people outside of household in the past 24 hours | 0.383 | 1.231 | 0.0 | 10.917 | 10.917 | inf |
| Been in a room with over 50 people outside of household in the past 24 hours | 0.115 | 1.08 | 1.956 | 6.175 | 4.219 | 215.695 |

**Table S7.7 shows the slopes, y-intercept, adherence % at the beginning of the study period, and at the end, the absolute difference between the beginning and the end, and the absolute difference as a percentage of the adherence % at the beginning of the study period of each behavior's trend in Connecticut.**

### State name: Delaware

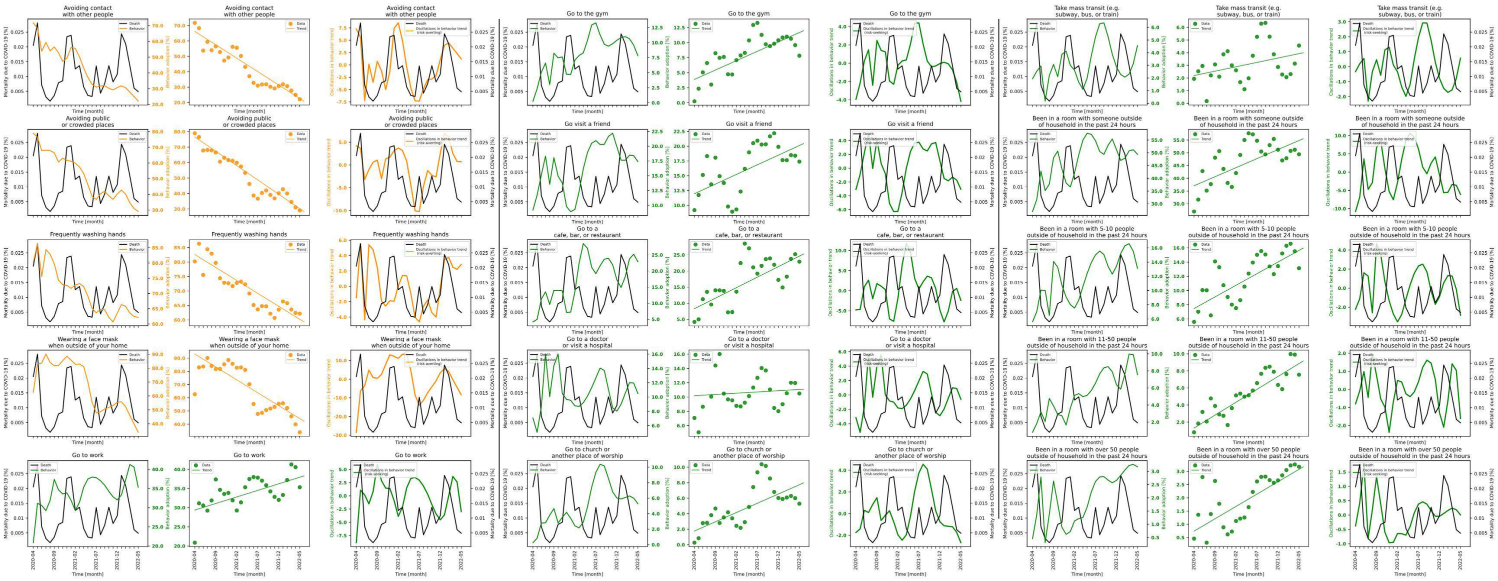

**Figure S7.8 shows behavior and mortality on 1st, 4th and 7th column, behavior trends in 2nd, 5th and 8th column, and behavior oscillatory components juxtaposed with mortality rates on 3rd, 6th and 9th columns for Delaware.**

| Behaviors | Slopes | Y-intercepts | Start %($S$) | End %($E$) | $\|E-S\|$ | $\frac{\|E-S\|*100}{S}$ |
|---|---|---|---|---|---|---|
| Avoiding contact with other people | -1.742 | 66.209 | 71.74 | 22.113 | 49.627 | 69.176 |
| Avoiding public or crowded places | -1.854 | 76.579 | 78.927 | 29.052 | 49.875 | 63.191 |
| Frequently washing hands | -0.895 | 82.582 | 80.194 | 62.171 | 18.023 | 22.474 |
| Wearing a face mask when outside of your home | -1.954 | 93.089 | 62.671 | 33.856 | 28.815 | 45.978 |
| Go to work | 0.342 | 29.379 | 20.868 | 35.33 | 14.462 | 69.302 |
| Go to the gym | 0.312 | 3.884 | 0.246 | 7.823 | 7.577 | 3080.081 |
| Go visit a friend | 0.326 | 11.962 | 9.186 | 17.394 | 8.208 | 89.353 |
| Go to a cafe, bar, or restaurant | 0.654 | 8.29 | 4.156 | 22.909 | 18.753 | 451.227 |
| Go to a doctor or visit a hospital | 0.035 | 10.19 | 7.077 | 10.497 | 3.42 | 48.326 |
| Go to church or another place of worship | 0.239 | 1.765 | 0.261 | 5.29 | 5.029 | 1926.82 |
| Take mass transit (e.g. subway, bus, or train) | 0.069 | 2.195 | 1.933 | 4.562 | 2.629 | 136.006 |
| Been in a room with someone outside of household in the past 24 hours | 0.709 | 37.074 | 26.988 | 49.41 | 22.422 | 83.081 |
| Been in a room with 5-10 people outside of household in the past 24 hours | 0.327 | 7.472 | 5.592 | 13.14 | 7.548 | 134.979 |
| Been in a room with 11-50 people outside of household in the past 24 hours | 0.302 | 1.306 | 0.804 | 7.558 | 6.754 | 840.05 |
| Been in a room with over 50 people outside of household in the past 24 hours | 0.093 | 0.741 | 0.457 | 3.176 | 2.719 | 594.967 |

**Table S7.8 shows the slopes, y-intercept, adherence % at the beginning of the study period, and at the end, the absolute difference between the beginning and the end, and the absolute difference as a percentage of the adherence % at the beginning of the study period of each behavior's trend in Delaware.**

### State name: District of Columbia

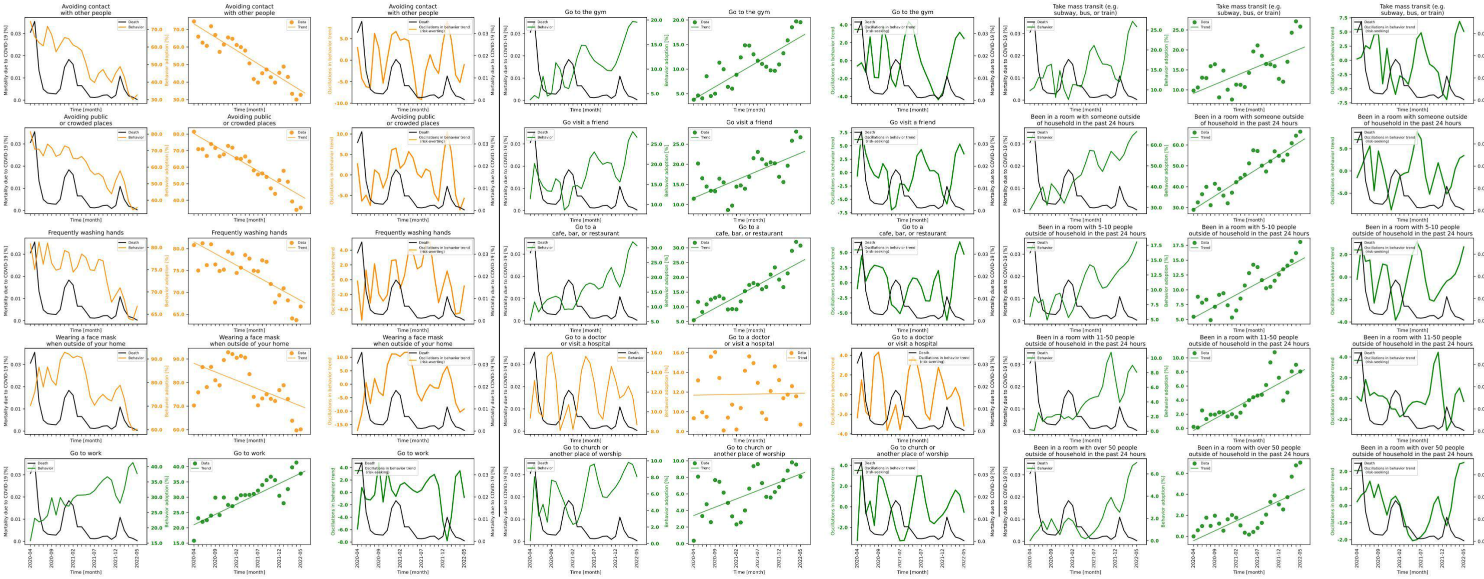

**Figure S7.9 shows behavior and mortality on 1st, 4th and 7th column, behavior trends in 2nd, 5th and 8th column, and behavior oscillatory components juxtaposed with mortality rates on 3rd, 6th and 9th columns for District of Columbia.**

| Behaviors | Slopes | Y-intercepts | Start %($S$) | End %($E$) | $\lvert E-S \rvert$ | $\frac{\lvert E-S \rvert*100}{S}$ |
|---|---|---|---|---|---|---|
| Avoiding contact with other people | -1.516 | 73.156 | 74.583 | 32.705 | 41.878 | 56.15 |
| Avoiding public or crowded places | -1.502 | 80.263 | 81.462 | 35.516 | 45.946 | 56.402 |
| Frequently washing hands | -0.535 | 81.539 | 80.825 | 66.77 | 14.055 | 17.389 |
| Wearing a face mask when outside of your home | -0.723 | 88.143 | 70.27 | 60.153 | 10.117 | 14.397 |
| Go to work | 0.674 | 21.014 | 15.791 | 37.729 | 21.938 | 138.927 |
| Go to the gym | 0.508 | 3.852 | 3.764 | 19.551 | 15.787 | 419.421 |
| Go visit a friend | 0.445 | 11.653 | 11.458 | 26.714 | 15.256 | 133.147 |
| Go to a cafe, bar, or restaurant | 0.784 | 5.647 | 5.474 | 30.759 | 25.285 | 461.911 |
| Go to a doctor or visit a hospital | 0.008 | 11.689 | 9.341 | 8.693 | 0.648 | 6.937 |
| Go to church or another place of worship | 0.2 | 3.398 | 0.352 | 8.081 | 7.729 | 2195.739 |
| Take mass transit (e.g. subway, bus, or train) | 0.447 | 9.121 | 9.809 | 25.846 | 16.037 | 163.493 |
| Been in a room with someone outside of household in the past 24 hours | 1.3 | 29.121 | 28.954 | 66.38 | 37.426 | 129.26 |
| Been in a room with 5-10 people outside of household in the past 24 hours | 0.389 | 5.279 | 5.467 | 18.063 | 12.596 | 230.401 |
| Been in a room with 11-50 people outside of household in the past 24 hours | 0.332 | -0.307 | 0.202 | 8.06 | 7.858 | 3890.099 |
| Been in a room with over 50 people outside of household in the past 24 hours | 0.189 | -0.414 | 0.0 | 7.014 | 7.014 | inf |

**Table S7.9 shows the slopes, y-intercept, adherence % at the beginning of the study period, and at the end, the absolute difference between the beginning and the end, and the absolute difference as a percentage of the adherence % at the beginning of the study period of each behavior's trend in District of Columbia.**

### State name: Florida

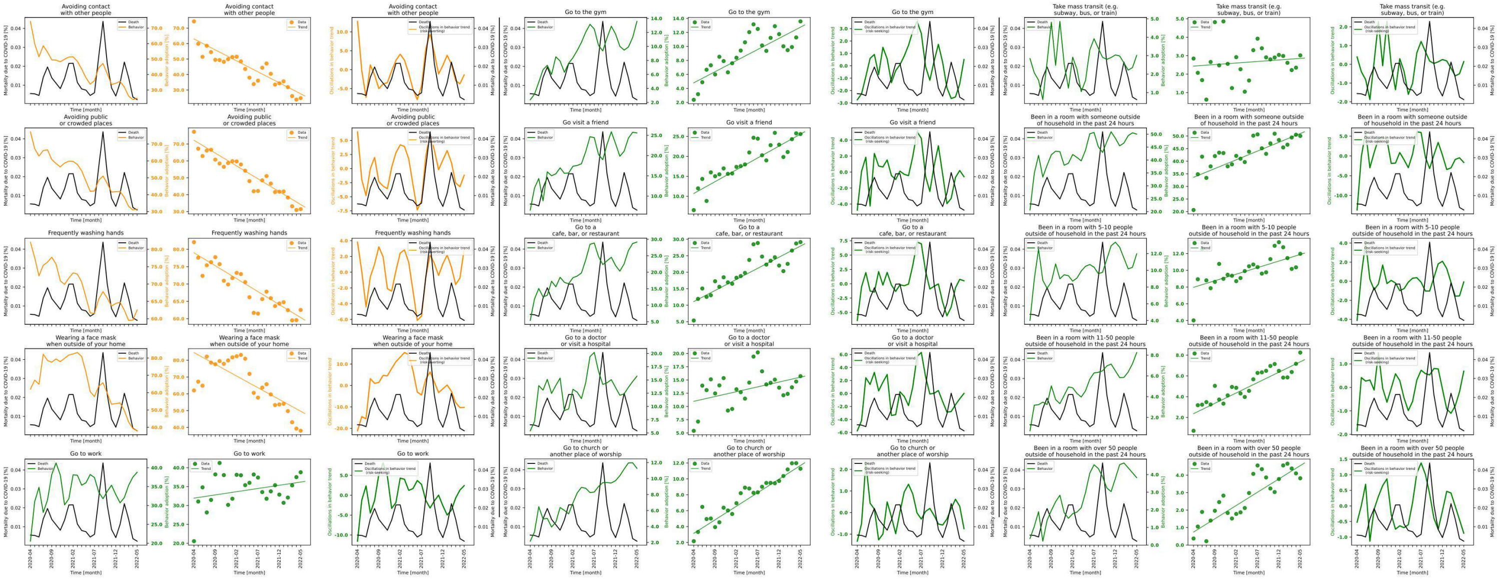

**Figure S7.10 shows behavior and mortality on 1st, 4th and 7th column, behavior trends in 2nd, 5th and 8th column, and behavior oscillatory components juxtaposed with mortality rates on 3rd, 6th and 9th columns for Florida.**

| Behaviors | Slopes | Y-intercepts | Start %($S$) | End %($E$) | $\lvert E-S\rvert$ | $\frac{\lvert E-S\rvert*100}{S}$ |
|---|---|---|---|---|---|---|
| Avoiding contact with other people | -1.42 | 63.009 | 74.349 | 24.723 | 49.626 | 66.747 |
| Avoiding public or crowded places | -1.523 | 72.222 | 77.227 | 31.447 | 45.78 | 59.28 |
| Frequently washing hands | -0.747 | 79.001 | 82.064 | 62.517 | 19.547 | 23.819 |
| Wearing a face mask when outside of your home | -1.397 | 84.557 | 62.027 | 38.002 | 24.025 | 38.733 |
| Go to work | 0.17 | 31.947 | 20.555 | 38.789 | 18.234 | 88.708 |
| Go to the gym | 0.318 | 4.804 | 2.375 | 13.578 | 11.203 | 471.705 |
| Go visit a friend | 0.595 | 10.691 | 6.523 | 25.588 | 19.065 | 292.273 |
| Go to a cafe, bar, or restaurant | 0.674 | 11.094 | 5.365 | 29.147 | 23.782 | 443.281 |
| Go to a doctor or visit a hospital | 0.182 | 10.966 | 5.401 | 15.715 | 10.314 | 190.965 |
| Go to church or another place of worship | 0.342 | 3.146 | 2.161 | 11.28 | 9.119 | 421.981 |
| Take mass transit (e.g. subway, bus, or train) | 0.017 | 2.435 | 2.844 | 3.017 | 0.173 | 6.083 |
| Been in a room with someone outside of household in the past 24 hours | 0.695 | 33.607 | 20.627 | 50.033 | 29.406 | 142.561 |
| Been in a room with 5-10 people outside of household in the past 24 hours | 0.157 | 7.968 | 4.017 | 11.969 | 7.952 | 197.959 |
| Been in a room with 11-50 people outside of household in the past 24 hours | 0.2 | 2.363 | 0.715 | 8.25 | 7.535 | 1053.846 |
| Been in a room with over 50 people outside of household in the past 24 hours | 0.152 | 0.741 | 0.372 | 3.824 | 3.452 | 927.957 |

**Table S7.10 shows the slopes, y-intercept, adherence % at the beginning of the study period, and at the end, the absolute difference between the beginning and the end, and the absolute difference as a percentage of the adherence % at the beginning of the study period of each behavior's trend in Florida.**

### State name: Georgia

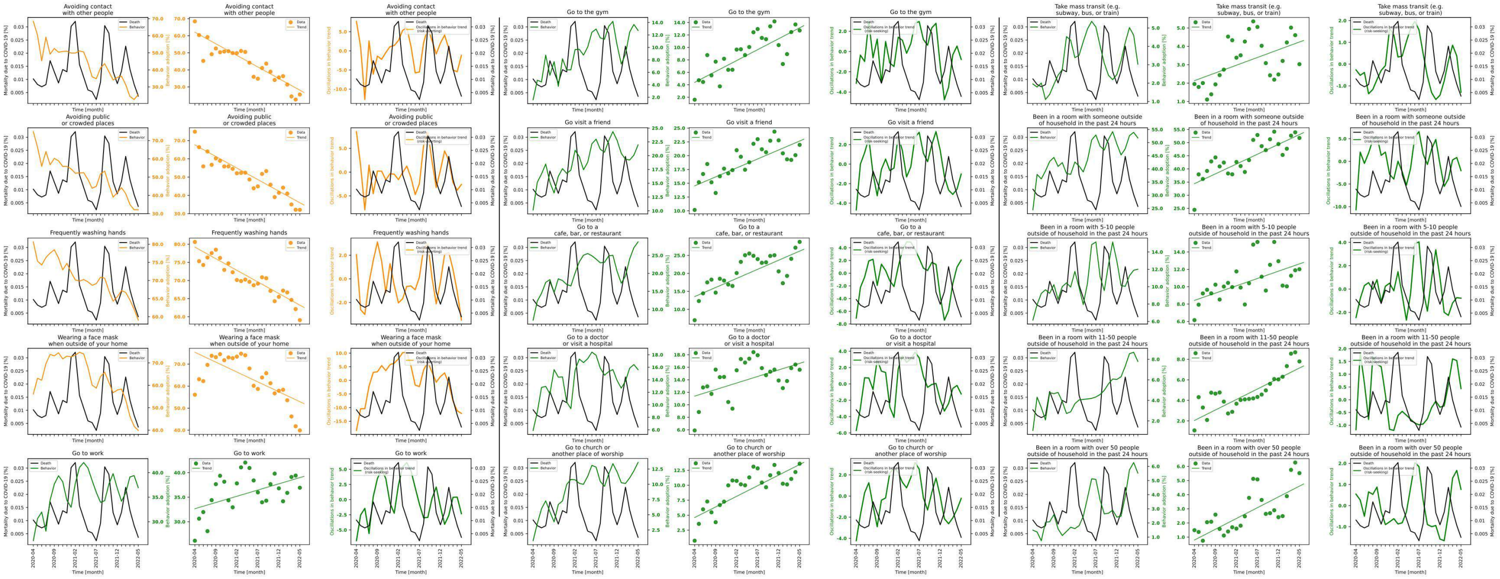

**Figure S7.11 shows behavior and mortality on 1st, 4th and 7th column, behavior trends in 2nd, 5th and 8th column, and behavior oscillatory components juxtaposed with mortality rates on 3rd, 6th and 9th columns for Georgia.**

| Behaviors | Slopes | Y-intercepts | Start %($S$) | End %($E$) | $\|E-S\|$ | $\frac{\|E-S\|*100}{S}$ |
|---|---|---|---|---|---|---|
| Avoiding contact with other people | -1.359 | 62.078 | 68.428 | 25.568 | 42.86 | 62.635 |
| Avoiding public or crowded places | -1.266 | 67.616 | 74.866 | 32.103 | 42.763 | 57.119 |
| Frequently washing hands | -0.641 | 79.211 | 80.63 | 59.066 | 21.564 | 26.744 |
| Wearing a face mask when outside of your home | -0.898 | 75.373 | 56.092 | 39.889 | 16.203 | 28.886 |
| Go to work | 0.254 | 32.656 | 26.132 | 36.929 | 10.797 | 41.317 |
| Go to the gym | 0.35 | 4.388 | 1.607 | 12.72 | 11.113 | 691.537 |
| Go visit a friend | 0.325 | 14.53 | 10.158 | 21.961 | 11.803 | 116.194 |
| Go to a cafe, bar, or restaurant | 0.506 | 13.553 | 6.985 | 28.762 | 21.777 | 311.768 |
| Go to a doctor or visit a hospital | 0.209 | 11.388 | 5.9 | 15.603 | 9.703 | 164.458 |
| Go to church or another place of worship | 0.354 | 4.612 | 0.742 | 13.598 | 12.856 | 1732.615 |
| Take mass transit (e.g. subway, bus, or train) | 0.084 | 2.134 | 1.951 | 3.03 | 1.079 | 55.305 |
| Been in a room with someone outside of household in the past 24 hours | 0.747 | 34.395 | 24.447 | 51.769 | 27.322 | 111.76 |
| Been in a room with 5-10 people outside of household in the past 24 hours | 0.168 | 8.422 | 6.151 | 12.013 | 5.862 | 95.302 |
| Been in a room with 11-50 people outside of household in the past 24 hours | 0.204 | 2.044 | 1.073 | 7.791 | 6.718 | 626.095 |
| Been in a room with over 50 people outside of household in the past 24 hours | 0.152 | 0.793 | 1.487 | 5.524 | 4.037 | 271.486 |

**Table S7.11 shows the slopes, y-intercept, adherence % at the beginning of the study period, and at the end, the absolute difference between the beginning and the end, and the absolute difference as a percentage of the adherence % at the beginning of the study period of each behavior's trend in Georgia.**

#### State name: Hawaii

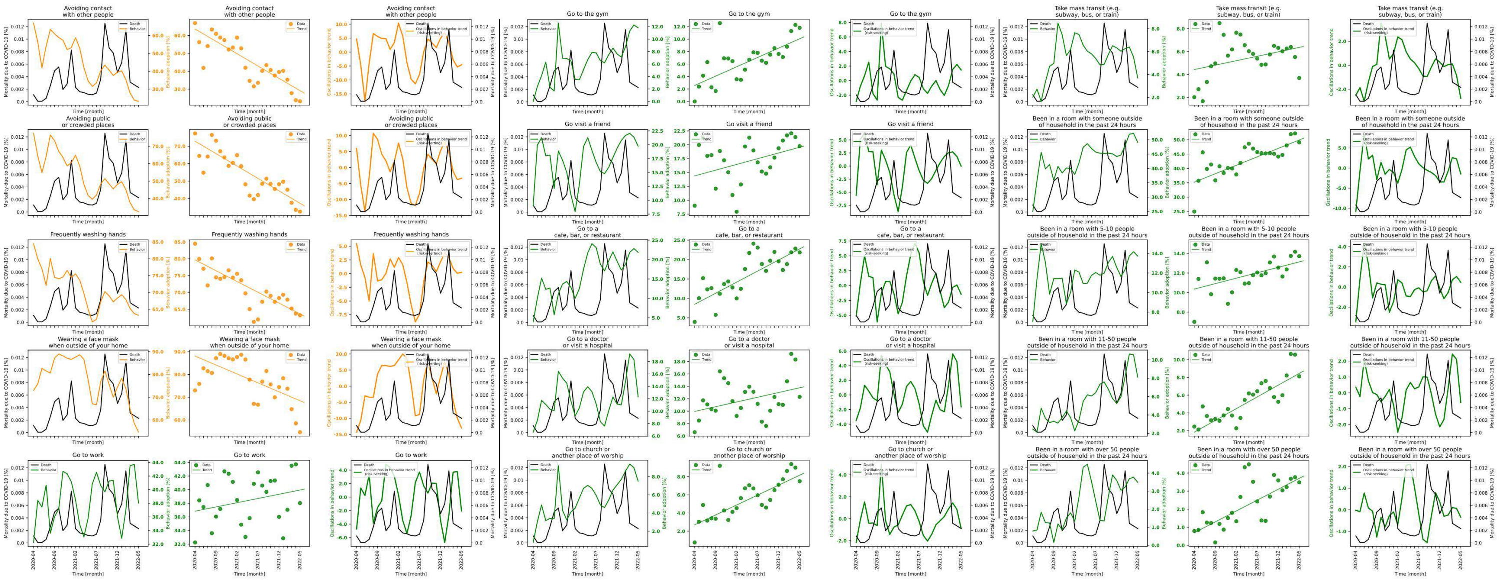

**Figure S7.12 shows behavior and mortality on 1st, 4th and 7th column, behavior trends in 2nd, 5th and 8th column, and behavior oscillatory components juxtaposed with mortality rates on 3rd, 6th and 9th columns for Hawaii.**

| Behaviors | Slopes | Y-intercepts | Start %($S$) | End %($E$) | $\lvert E-S \rvert$ | $\frac{\lvert E-S \rvert * 100}{S}$ |
|---|---|---|---|---|---|---|
| Avoiding contact with other people | -1.38 | 63.629 | 66.927 | 23.189 | 43.738 | 65.352 |
| Avoiding public or crowded places | -1.423 | 72.865 | 77.334 | 32.526 | 44.808 | 57.941 |
| Frequently washing hands | -0.642 | 79.666 | 84.479 | 63.201 | 21.278 | 25.187 |
| Wearing a face mask when outside of your home | -0.785 | 88.071 | 72.974 | 54.622 | 18.352 | 25.149 |
| Go to work | 0.124 | 36.839 | 32.243 | 38.027 | 5.784 | 17.939 |
| Go to the gym | 0.3 | 2.565 | 0.0 | 11.815 | 11.815 | inf |
| Go visit a friend | 0.207 | 14.39 | 9.016 | 19.742 | 10.726 | 118.966 |
| Go to a cafe, bar, or restaurant | 0.57 | 8.486 | 3.937 | 21.849 | 17.912 | 454.966 |
| Go to a doctor or visit a hospital | 0.153 | 9.958 | 6.609 | 12.305 | 5.696 | 86.186 |
| Go to church or another place of worship | 0.219 | 2.697 | 0.757 | 7.509 | 6.752 | 891.942 |
| Take mass transit (e.g. subway, bus, or train) | 0.077 | 4.432 | 2.018 | 3.695 | 1.677 | 83.102 |
| Been in a room with someone outside of household in the past 24 hours | 0.589 | 35.202 | 24.952 | 49.066 | 24.114 | 96.642 |
| Been in a room with 5-10 people outside of household in the past 24 hours | 0.111 | 10.357 | 7.004 | 13.723 | 6.719 | 95.931 |
| Been in a room with 11-50 people outside of household in the past 24 hours | 0.265 | 1.831 | 2.458 | 8.12 | 5.662 | 230.35 |
| Been in a room with over 50 people outside of household in the past 24 hours | 0.12 | 0.694 | 0.79 | 3.485 | 2.695 | 341.139 |

**Table S7.12 shows the slopes, y-intercept, adherence % at the beginning of the study period, and at the end, the absolute difference between the beginning and the end, and the absolute difference as a percentage of the adherence % at the beginning of the study period of each behavior's trend in Hawaii.**

### State name: Idaho

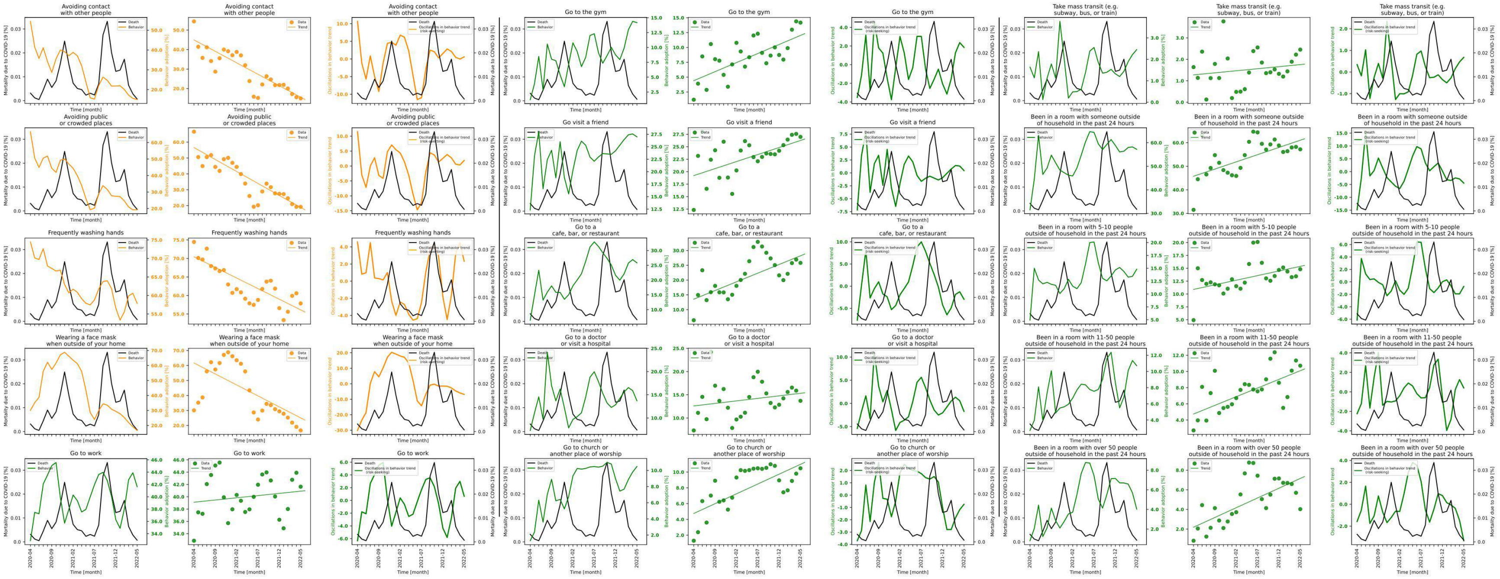

**Figure S7.13 shows behavior and mortality on 1st, 4th and 7th column, behavior trends in 2nd, 5th and 8th column, and behavior oscillatory components juxtaposed with mortality rates on 3rd, 6th and 9th columns for Idaho.**

| Behaviors | Slopes | Y-intercepts | Start %($S$) | End %($E$) | $\lvert E-S \rvert$ | $\frac{\lvert E-S \rvert * 100}{S}$ |
|---|---|---|---|---|---|---|
| Avoiding contact with other people | -1.176 | 44.971 | 54.457 | 14.976 | 39.481 | 72.499 |
| Avoiding public or crowded places | -1.509 | 56.59 | 66.591 | 19.216 | 47.375 | 71.143 |
| Frequently washing hands | -0.574 | 70.454 | 74.454 | 57.859 | 16.595 | 22.289 |
| Wearing a face mask when outside of your home | -1.465 | 61.675 | 30.088 | 16.706 | 13.382 | 44.476 |
| Go to work | 0.072 | 39.116 | 32.86 | 41.634 | 8.774 | 26.701 |
| Go to the gym | 0.305 | 4.404 | 1.199 | 14.182 | 12.983 | 1082.819 |
| Go visit a friend | 0.282 | 19.215 | 12.325 | 26.984 | 14.659 | 118.937 |
| Go to a cafe, bar, or restaurant | 0.587 | 13.538 | 6.433 | 25.827 | 19.394 | 301.477 |
| Go to a doctor or visit a hospital | 0.109 | 12.597 | 7.315 | 13.706 | 6.391 | 87.368 |
| Go to church or another place of worship | 0.249 | 4.75 | 1.297 | 10.481 | 9.184 | 708.096 |
| Take mass transit (e.g. subway, bus, or train) | 0.019 | 1.273 | 1.637 | 2.452 | 0.815 | 49.786 |
| Been in a room with someone outside of household in the past 24 hours | 0.612 | 45.757 | 31.503 | 57.199 | 25.696 | 81.567 |
| Been in a room with 5-10 people outside of household in the past 24 hours | 0.179 | 10.893 | 4.928 | 14.821 | 9.893 | 200.751 |
| Been in a room with 11-50 people outside of household in the past 24 hours | 0.214 | 4.741 | 2.715 | 10.716 | 8.001 | 294.696 |
| Been in a room with over 50 people outside of household in the past 24 hours | 0.199 | 2.187 | 0.812 | 4.027 | 3.215 | 395.936 |

**Table S7.13 shows the slopes, y-intercept, adherence % at the beginning of the study period, and at the end, the absolute difference between the beginning and the end, and the absolute difference as a percentage of the adherence % at the beginning of the study period of each behavior's trend in Idaho.**

#### State name: Illinois

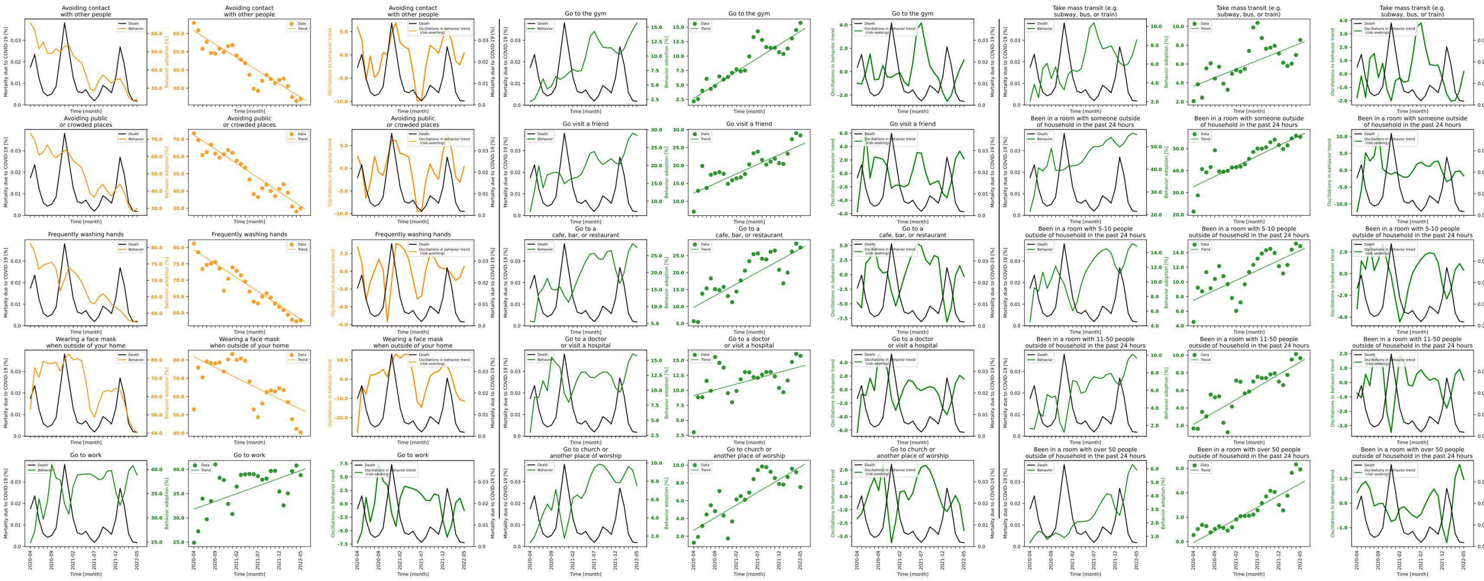

**Figure S7.14 shows behavior and mortality on 1st, 4th and 7th column, behavior trends in 2nd, 5th and 8th column, and behavior oscillatory components juxtaposed with mortality rates on 3rd, 6th and 9th columns for Illinois.**

| Behaviors | Slopes | Y-intercepts | Start %($S$) | End %($E$) | $\|E-S\|$ | $\frac{\|E-S\|*100}{S}$ |
|---|---|---|---|---|---|---|
| Avoiding contact with other people | -1.454 | 61.402 | 65.833 | 24.021 | 41.812 | 63.512 |
| Avoiding public or crowded places | -1.646 | 72.372 | 73.379 | 29.923 | 43.456 | 59.221 |
| Frequently washing hands | -0.848 | 79.258 | 81.168 | 57.734 | 23.434 | 28.871 |
| Wearing a face mask when outside of your home | -1.15 | 81.707 | 52.974 | 40.425 | 12.549 | 23.689 |
| Go to work | 0.318 | 31.8 | 24.871 | 38.8 | 13.929 | 56.005 |
| Go to the gym | 0.464 | 2.69 | 2.177 | 15.732 | 13.555 | 622.646 |
| Go visit a friend | 0.537 | 12.272 | 7.104 | 28.399 | 21.295 | 299.761 |
| Go to a cafe, bar, or restaurant | 0.691 | 9.758 | 5.667 | 27.416 | 21.749 | 383.783 |
| Go to a doctor or visit a hospital | 0.194 | 9.11 | 2.993 | 15.741 | 12.748 | 425.927 |
| Go to church or another place of worship | 0.286 | 2.662 | 1.276 | 7.527 | 6.251 | 489.89 |
| Take mass transit (e.g. subway, bus, or train) | 0.182 | 3.643 | 2.066 | 8.55 | 6.484 | 313.843 |
| Been in a room with someone outside of household in the past 24 hours | 0.933 | 32.692 | 21.413 | 55.314 | 33.901 | 158.32 |
| Been in a room with 5-10 people outside of household in the past 24 hours | 0.273 | 7.486 | 4.532 | 14.885 | 10.353 | 228.442 |
| Been in a room with 11-50 people outside of household in the past 24 hours | 0.284 | 2.124 | 1.663 | 9.668 | 8.005 | 481.359 |
| Been in a room with over 50 people outside of household in the past 24 hours | 0.193 | -0.091 | 0.531 | 5.905 | 5.374 | 1012.053 |

**Table S7.14 shows the slopes, y-intercept, adherence % at the beginning of the study period, and at the end, the absolute difference between the beginning and the end, and the absolute difference as a percentage of the adherence % at the beginning of the study period of each behavior's trend in Illinois.**

### State name: Indiana

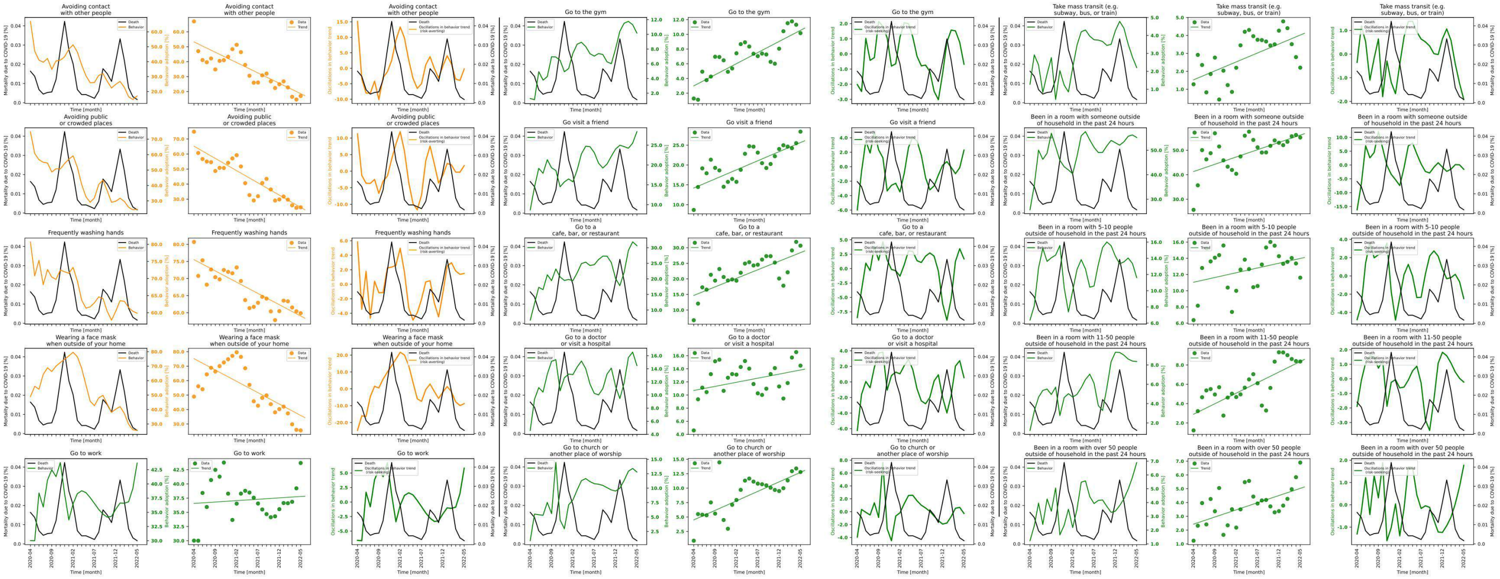

**Figure S7.15 shows behavior and mortality on 1st, 4th and 7th column, behavior trends in 2nd, 5th and 8th column, and behavior oscillatory components juxtaposed with mortality rates on 3rd, 6th and 9th columns for Indiana.**

| Behaviors | Slopes | Y-intercepts | Start %$(S)$ | End %$(E)$ | $\lvert E-S\rvert$ | $\frac{\lvert E-S\rvert*100}{S}$ |
|---|---|---|---|---|---|---|
| Avoiding contact with other people | -1.385 | 53.231 | 66.992 | 17.0 | 49.992 | 74.624 |
| Avoiding public or crowded places | -1.627 | 65.214 | 74.872 | 24.581 | 50.291 | 67.169 |
| Frequently washing hands | -0.66 | 75.523 | 80.7 | 59.865 | 20.835 | 25.818 |
| Wearing a face mask when outside of your home | -1.571 | 75.191 | 48.947 | 25.556 | 23.391 | 47.788 |
| Go to work | 0.049 | 36.568 | 30.019 | 43.681 | 13.662 | 45.511 |
| Go to the gym | 0.302 | 2.995 | 1.276 | 10.166 | 8.89 | 696.708 |
| Go visit a friend | 0.458 | 14.201 | 8.675 | 28.415 | 19.74 | 227.55 |
| Go to a cafe, bar, or restaurant | 0.549 | 14.688 | 6.733 | 30.662 | 23.929 | 355.399 |
| Go to a doctor or visit a hospital | 0.125 | 10.694 | 4.589 | 14.487 | 9.898 | 215.69 |
| Go to church or another place of worship | 0.329 | 4.503 | 0.899 | 12.789 | 11.89 | 1322.581 |
| Take mass transit (e.g. subway, bus, or train) | 0.1 | 1.525 | 1.276 | 2.205 | 0.929 | 72.806 |
| Been in a room with someone outside of household in the past 24 hours | 0.594 | 41.306 | 25.69 | 55.2 | 29.51 | 114.87 |
| Been in a room with 5-10 people outside of household in the past 24 hours | 0.117 | 11.046 | 6.372 | 11.611 | 5.239 | 82.219 |
| Been in a room with 11-50 people outside of household in the past 24 hours | 0.219 | 2.899 | 1.227 | 8.36 | 7.133 | 581.337 |
| Been in a room with over 50 people outside of household in the past 24 hours | 0.104 | 2.433 | 1.231 | 6.9 | 5.669 | 460.52 |

**Table S7.15 shows the slopes, y-intercept, adherence % at the beginning of the study period, and at the end, the absolute difference between the beginning and the end, and the absolute difference as a percentage of the adherence % at the beginning of the study period of each behavior's trend in Indiana.**

### State name: Iowa

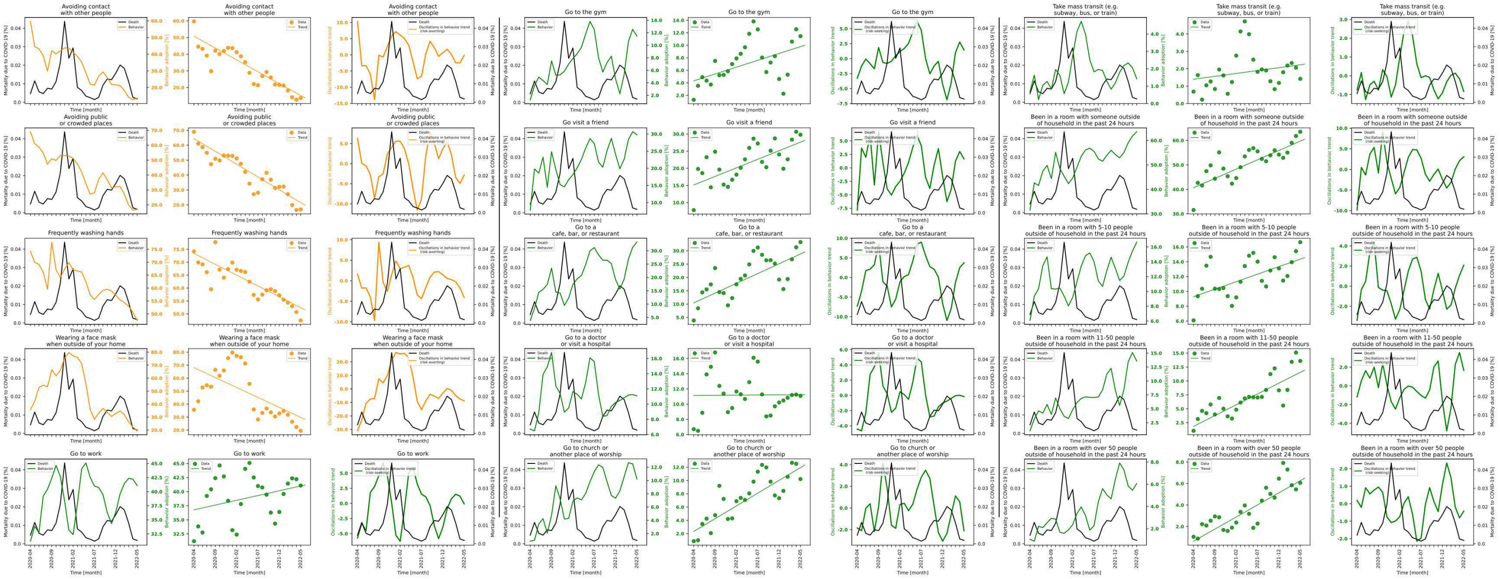

**Figure S7.16 shows behavior and mortality on 1st, 4th and 7th column, behavior trends in 2nd, 5th and 8th column, and behavior oscillatory components juxtaposed with mortality rates on 3rd, 6th and 9th columns for Iowa.**

| Behaviors | Slopes | Y-intercepts | Start %($S$) | End %($E$) | $\|E-S\|$ | $\frac{\|E-S\|*100}{S}$ |
|---|---|---|---|---|---|---|
| Avoiding contact with other people | -1.422 | 50.792 | 59.682 | 13.667 | 46.015 | 77.1 |
| Avoiding public or crowded places | -1.716 | 64.427 | 68.999 | 16.966 | 52.033 | 75.411 |
| Frequently washing hands | -0.84 | 73.41 | 74.2 | 47.484 | 26.716 | 36.005 |
| Wearing a face mask when outside of your home | -1.532 | 68.126 | 35.611 | 19.474 | 16.137 | 45.315 |
| Go to work | 0.17 | 36.769 | 31.208 | 41.09 | 9.882 | 31.665 |
| Go to the gym | 0.219 | 4.317 | 1.283 | 11.454 | 10.171 | 792.751 |
| Go visit a friend | 0.5 | 15.137 | 7.797 | 29.789 | 21.992 | 282.057 |
| Go to a cafe, bar, or restaurant | 0.733 | 10.451 | 3.858 | 33.25 | 29.392 | 761.846 |
| Go to a doctor or visit a hospital | 0.004 | 11.133 | 6.705 | 11.083 | 4.378 | 65.295 |
| Go to church or another place of worship | 0.384 | 2.326 | 0.862 | 10.213 | 9.351 | 1084.803 |
| Take mass transit (e.g. subway, bus, or train) | 0.033 | 1.397 | 0.684 | 1.419 | 0.735 | 107.456 |
| Been in a room with someone outside of household in the past 24 hours | 0.761 | 40.671 | 31.571 | 63.529 | 31.958 | 101.226 |
| Been in a room with 5-10 people outside of household in the past 24 hours | 0.203 | 9.282 | 6.087 | 16.646 | 10.559 | 173.468 |
| Been in a room with 11-50 people outside of household in the past 24 hours | 0.388 | 1.837 | 1.043 | 13.59 | 12.547 | 1202.972 |
| Been in a room with over 50 people outside of household in the past 24 hours | 0.225 | 0.636 | 1.047 | 6.063 | 5.016 | 479.083 |

**Table S7.16 shows the slopes, y-intercept, adherence % at the beginning of the study period, and at the end, the absolute difference between the beginning and the end, and the absolute difference as a percentage of the adherence % at the beginning of the study period of each behavior's trend in Iowa.**

### State name: Kansas

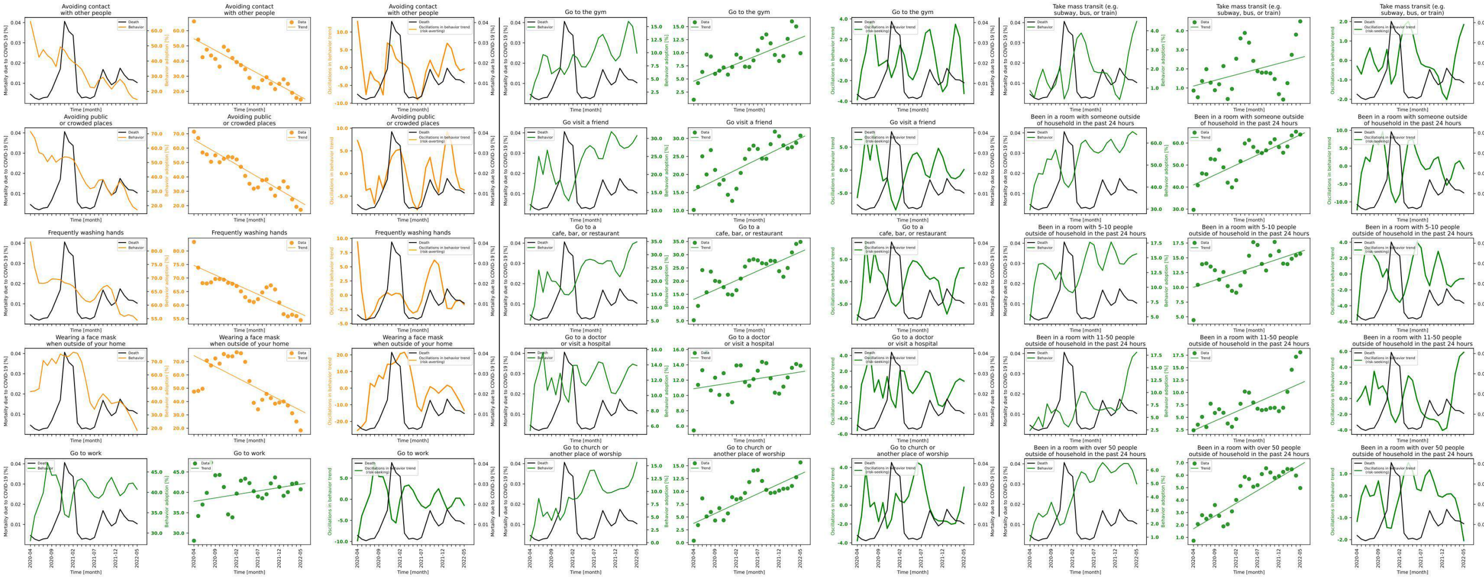

**Figure S7.17 shows behavior and mortality on 1st, 4th and 7th column, behavior trends in 2nd, 5th and 8th column, and behavior oscillatory components juxtaposed with mortality rates on 3rd, 6th and 9th columns for Kansas.**

| Behaviors | Slopes | Y-intercepts | Start %($S$) | End %($E$) | $\lvert E-S \rvert$ | $\frac{\lvert E-S \rvert * 100}{S}$ |
|---|---|---|---|---|---|---|
| Avoiding contact with other people | -1.524 | 54.654 | 66.125 | 14.651 | 51.474 | 77.843 |
| Avoiding public or crowded places | -1.738 | 65.912 | 71.45 | 17.111 | 54.339 | 76.052 |
| Frequently washing hands | -0.713 | 74.669 | 83.328 | 54.451 | 28.877 | 34.655 |
| Wearing a face mask when outside of your home | -1.647 | 74.47 | 47.417 | 18.32 | 29.097 | 61.364 |
| Go to work | 0.17 | 37.775 | 28.13 | 40.749 | 12.619 | 44.86 |
| Go to the gym | 0.332 | 4.557 | 1.032 | 9.921 | 8.889 | 861.337 |
| Go visit a friend | 0.582 | 15.508 | 10.153 | 30.778 | 20.625 | 203.142 |
| Go to a cafe, bar, or restaurant | 0.717 | 13.097 | 5.205 | 34.836 | 29.631 | 569.28 |
| Go to a doctor or visit a hospital | 0.088 | 10.883 | 5.424 | 13.894 | 8.47 | 156.158 |
| Go to church or another place of worship | 0.384 | 3.769 | 0.382 | 15.666 | 15.284 | 4001.047 |
| Take mass transit (e.g. subway, bus, or train) | 0.059 | 1.102 | 0.864 | 4.491 | 3.627 | 419.792 |
| Been in a room with someone outside of household in the past 24 hours | 0.909 | 40.981 | 29.677 | 63.87 | 34.193 | 115.217 |
| Been in a room with 5-10 people outside of household in the past 24 hours | 0.241 | 10.008 | 4.44 | 15.678 | 11.238 | 253.108 |
| Been in a room with 11-50 people outside of household in the past 24 hours | 0.378 | 2.371 | 2.392 | 18.091 | 15.699 | 656.313 |
| Been in a room with over 50 people outside of household in the past 24 hours | 0.205 | 1.697 | 0.734 | 4.969 | 4.235 | 576.975 |

**Table S7.17 shows the slopes, y-intercept, adherence % at the beginning of the study period, and at the end, the absolute difference between the beginning and the end, and the absolute difference as a percentage of the adherence % at the beginning of the study period of each behavior's trend in Kansas.**

#### State name: Kentucky

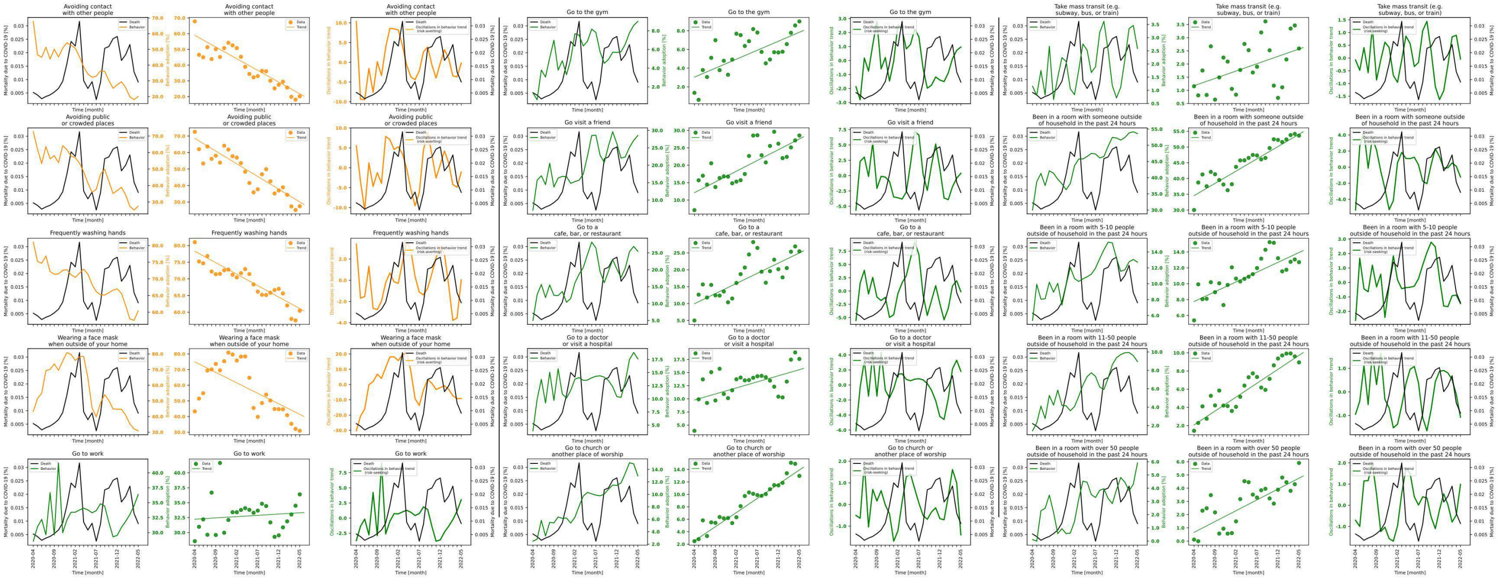

**Figure S7.18 shows behavior and mortality on 1st, 4th and 7th column, behavior trends in 2nd, 5th and 8th column, and behavior oscillatory components juxtaposed with mortality rates on 3rd, 6th and 9th columns for Kentucky.**

| Behaviors | Slopes | Y-intercepts | Start %($S$) | End %($E$) | $\|E-S\|$ | $\frac{\|E-S\|*100}{S}$ |
|---|---|---|---|---|---|---|
| Avoiding contact with other people | -1.485 | 58.908 | 67.803 | 20.14 | 47.663 | 70.296 |
| Avoiding public or crowded places | -1.542 | 68.595 | 72.597 | 27.455 | 45.142 | 62.182 |
| Frequently washing hands | -0.688 | 78.28 | 81.009 | 60.442 | 20.567 | 25.389 |
| Wearing a face mask when outside of your home | -1.342 | 74.923 | 43.282 | 30.894 | 12.388 | 28.622 |
| Go to work | 0.043 | 32.236 | 28.556 | 36.396 | 7.84 | 27.455 |
| Go to the gym | 0.192 | 3.038 | 1.361 | 9.003 | 7.642 | 561.499 |
| Go visit a friend | 0.617 | 12.114 | 7.101 | 28.536 | 21.435 | 301.859 |
| Go to a cafe, bar, or restaurant | 0.601 | 9.954 | 5.003 | 25.444 | 20.441 | 408.575 |
| Go to a doctor or visit a hospital | 0.231 | 9.78 | 3.905 | 17.601 | 13.696 | 350.73 |
| Go to church or another place of worship | 0.456 | 2.527 | 2.478 | 12.968 | 10.49 | 423.325 |
| Take mass transit (e.g. subway, bus, or train) | 0.056 | 1.175 | 1.153 | 2.589 | 1.436 | 124.545 |
| Been in a room with someone outside of household in the past 24 hours | 0.779 | 34.568 | 30.018 | 53.604 | 23.586 | 78.573 |
| Been in a room with 5-10 people outside of household in the past 24 hours | 0.247 | 7.759 | 5.382 | 12.712 | 7.33 | 136.195 |
| Been in a room with 11-50 people outside of household in the past 24 hours | 0.306 | 2.072 | 1.429 | 8.952 | 7.523 | 526.452 |
| Been in a room with over 50 people outside of household in the past 24 hours | 0.164 | 0.661 | 0.113 | 5.908 | 5.795 | 5128.319 |

**Table S7.18 shows the slopes, y-intercept, adherence % at the beginning of the study period, and at the end, the absolute difference between the beginning and the end, and the absolute difference as a percentage of the adherence % at the beginning of the study period of each behavior's trend in Kentucky.**

### State name: Louisiana

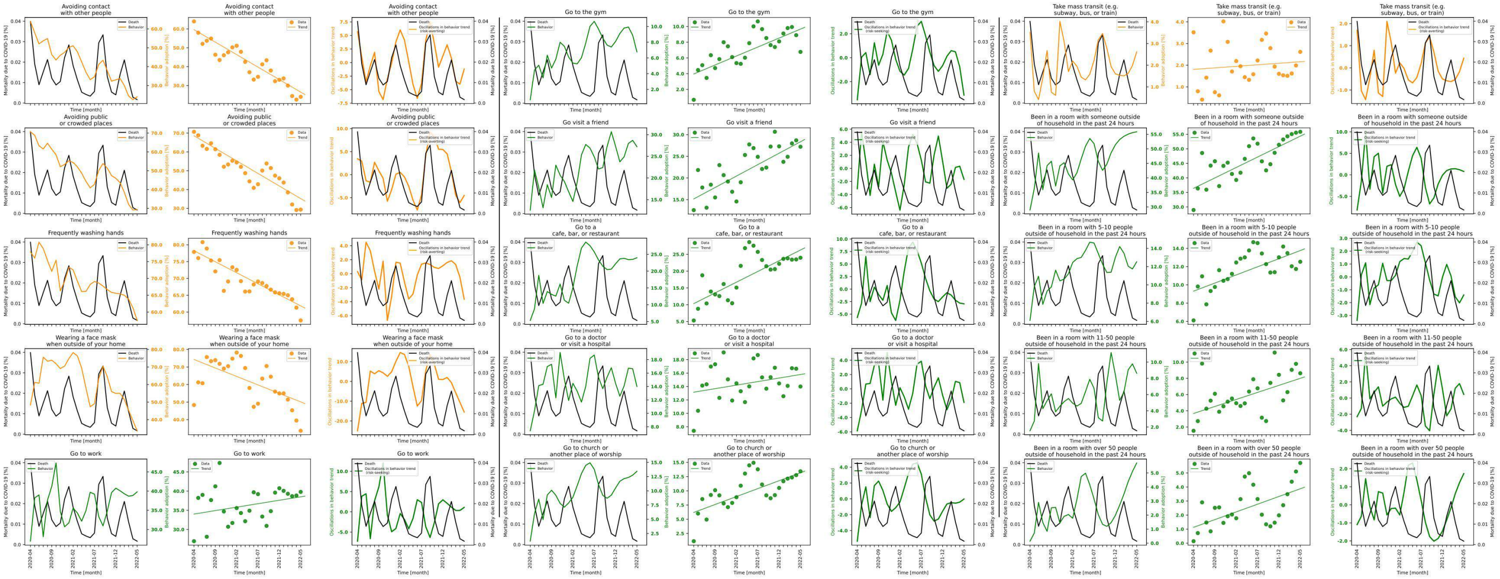

**Figure S7.19 shows behavior and mortality on 1st, 4th and 7th column, behavior trends in 2nd, 5th and 8th column, and behavior oscillatory components juxtaposed with mortality rates on 3rd, 6th and 9th columns for Louisiana.**

| Behaviors | Slopes | Y-intercepts | Start %($S$) | End %($E$) | $\|E-S\|$ | $\frac{\|E-S\|*100}{S}$ |
|---|---|---|---|---|---|---|
| Avoiding contact with other people | -1.325 | 59.63 | 64.003 | 23.984 | 40.019 | 62.527 |
| Avoiding public or crowded places | -1.337 | 68.588 | 70.699 | 29.327 | 41.372 | 58.519 |
| Frequently washing hands | -0.656 | 78.245 | 77.885 | 57.521 | 20.364 | 26.146 |
| Wearing a face mask when outside of your home | -0.966 | 74.268 | 48.325 | 33.638 | 14.687 | 30.392 |
| Go to work | 0.18 | 34.017 | 26.949 | 39.795 | 12.846 | 47.668 |
| Go to the gym | 0.228 | 3.964 | 0.676 | 6.767 | 6.091 | 901.036 |
| Go visit a friend | 0.528 | 15.164 | 12.574 | 27.21 | 14.636 | 116.399 |
| Go to a cafe, bar, or restaurant | 0.638 | 10.332 | 5.305 | 23.989 | 18.684 | 352.196 |
| Go to a doctor or visit a hospital | 0.105 | 13.15 | 7.397 | 13.999 | 6.602 | 89.252 |
| Go to church or another place of worship | 0.279 | 6.201 | 1.144 | 13.438 | 12.294 | 1074.65 |
| Take mass transit (e.g. subway, bus, or train) | 0.014 | 1.806 | 3.516 | 2.615 | 0.901 | 25.626 |
| Been in a room with someone outside of household in the past 24 hours | 0.718 | 36.398 | 28.951 | 55.852 | 26.901 | 92.919 |
| Been in a room with 5-10 people outside of household in the past 24 hours | 0.178 | 9.311 | 6.107 | 12.531 | 6.424 | 105.191 |
| Been in a room with 11-50 people outside of household in the past 24 hours | 0.173 | 3.671 | 1.538 | 8.626 | 7.088 | 460.858 |
| Been in a room with over 50 people outside of household in the past 24 hours | 0.11 | 1.129 | 0.141 | 5.712 | 5.571 | 3951.064 |

**Table S7.19 shows the slopes, y-intercept, adherence % at the beginning of the study period, and at the end, the absolute difference between the beginning and the end, and the absolute difference as a percentage of the adherence % at the beginning of the study period of each behavior's trend in Louisiana.**

### State name: Maine

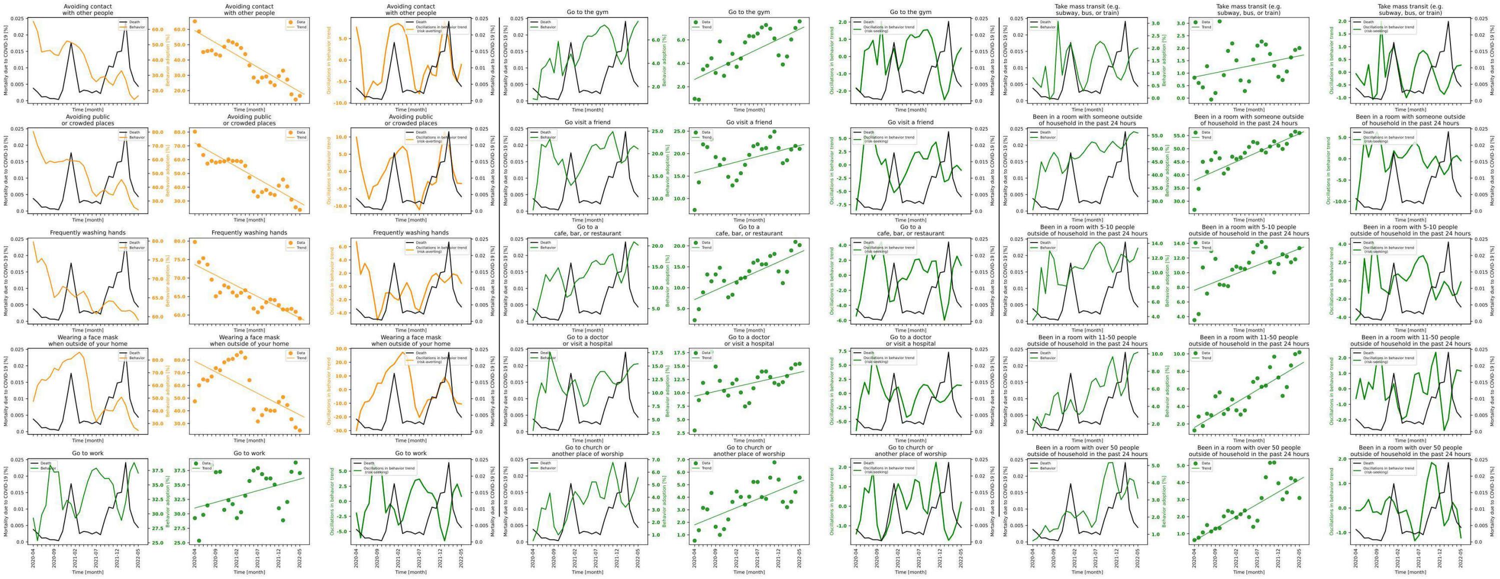

**Figure S7.20 shows behavior and mortality on 1st, 4th and 7th column, behavior trends in 2nd, 5th and 8th column, and behavior oscillatory components juxtaposed with mortality rates on 3rd, 6th and 9th columns for Maine.**

| Behaviors | Slopes | Y-intercepts | Start %($S$) | End %($E$) | $\|E-S\|$ | $\frac{\|E-S\|*100}{S}$ |
|---|---|---|---|---|---|---|
| Avoiding contact with other people | -1.598 | 59.173 | 65.214 | 16.6 | 48.614 | 74.545 |
| Avoiding public or crowded places | -1.73 | 72.076 | 80.307 | 23.617 | 56.69 | 70.592 |
| Frequently washing hands | -0.577 | 73.617 | 79.759 | 59.096 | 20.663 | 25.907 |
| Wearing a face mask when outside of your home | -1.691 | 79.037 | 47.469 | 24.573 | 22.896 | 48.234 |
| Go to work | 0.2 | 31.01 | 29.263 | 37.054 | 7.791 | 26.624 |
| Go to the gym | 0.172 | 2.629 | 0.969 | 7.581 | 6.612 | 682.353 |
| Go visit a friend | 0.243 | 15.712 | 7.467 | 21.052 | 13.585 | 181.934 |
| Go to a cafe, bar, or restaurant | 0.45 | 7.177 | 2.235 | 20.183 | 17.948 | 803.043 |
| Go to a doctor or visit a hospital | 0.178 | 9.38 | 2.976 | 15.431 | 12.455 | 418.515 |
| Go to church or another place of worship | 0.137 | 1.803 | 0.532 | 5.566 | 5.034 | 946.241 |
| Take mass transit (e.g. subway, bus, or train) | 0.034 | 0.844 | 0.817 | 2.006 | 1.189 | 145.532 |
| Been in a room with someone outside of household in the past 24 hours | 0.711 | 37.973 | 26.713 | 55.993 | 29.28 | 109.61 |
| Been in a room with 5-10 people outside of household in the past 24 hours | 0.227 | 7.603 | 3.518 | 13.352 | 9.834 | 279.534 |
| Been in a room with 11-50 people outside of household in the past 24 hours | 0.286 | 1.599 | 1.263 | 10.181 | 8.918 | 706.097 |
| Been in a room with over 50 people outside of household in the past 24 hours | 0.143 | 0.589 | 0.626 | 3.095 | 2.469 | 394.409 |

**Table S7.20 shows the slopes, y-intercept, adherence % at the beginning of the study period, and at the end, the absolute difference between the beginning and the end, and the absolute difference as a percentage of the adherence % at the beginning of the study period of each behavior's trend in Maine.**

### State name: Maryland

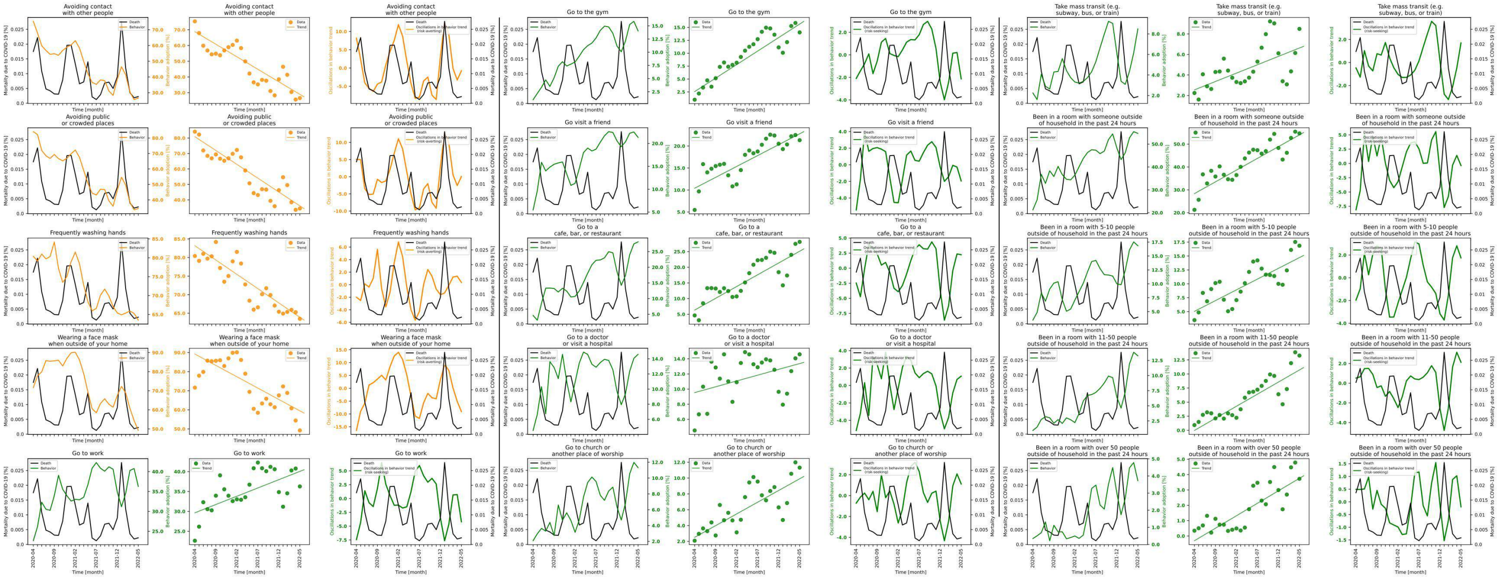

**Figure S7.21 shows behavior and mortality on 1st, 4th and 7th column, behavior trends in 2nd, 5th and 8th column, and behavior oscillatory components juxtaposed with mortality rates on 3rd, 6th and 9th columns for Maryland.**

| Behaviors | Slopes | Y-intercepts | Start %($S$) | End %($E$) | $\|E-S\|$ | $\frac{\|E-S\|*100}{S}$ |
|---|---|---|---|---|---|---|
| Avoiding contact with other people | -1.593 | 68.764 | 75.354 | 26.579 | 48.775 | 64.728 |
| Avoiding public or crowded places | -1.789 | 80.868 | 84.124 | 34.419 | 49.705 | 59.085 |
| Frequently washing hands | -0.77 | 83.181 | 80.412 | 63.506 | 16.906 | 21.024 |
| Wearing a face mask when outside of your home | -1.19 | 89.223 | 71.634 | 49.214 | 22.42 | 31.298 |
| Go to work | 0.416 | 29.646 | 22.63 | 36.271 | 13.641 | 60.278 |
| Go to the gym | 0.523 | 2.56 | 0.99 | 14.037 | 13.047 | 1317.879 |
| Go visit a friend | 0.495 | 10.406 | 5.463 | 21.33 | 15.867 | 290.445 |
| Go to a cafe, bar, or restaurant | 0.751 | 6.336 | 4.621 | 28.101 | 23.48 | 508.115 |
| Go to a doctor or visit a hospital | 0.154 | 9.536 | 4.53 | 14.597 | 10.067 | 222.23 |
| Go to church or another place of worship | 0.296 | 2.511 | 2.085 | 11.319 | 9.234 | 442.878 |
| Take mass transit (e.g. subway, bus, or train) | 0.163 | 2.551 | 2.244 | 8.489 | 6.245 | 278.298 |
| Been in a room with someone outside of household in the past 24 hours | 1.027 | 28.353 | 21.264 | 54.686 | 33.422 | 157.176 |
| Been in a room with 5-10 people outside of household in the past 24 hours | 0.387 | 5.047 | 3.471 | 16.855 | 13.384 | 385.595 |
| Been in a room with 11-50 people outside of household in the past 24 hours | 0.429 | -0.02 | 0.928 | 13.266 | 12.338 | 1329.526 |
| Been in a room with over 50 people outside of household in the past 24 hours | 0.163 | -0.308 | 0.346 | 3.724 | 3.378 | 976.301 |

**Table S7.21 shows the slopes, y-intercept, adherence % at the beginning of the study period, and at the end, the absolute difference between the beginning and the end, and the absolute difference as a percentage of the adherence % at the beginning of the study period of each behavior's trend in Maryland.**

#### State name: Massachusetts

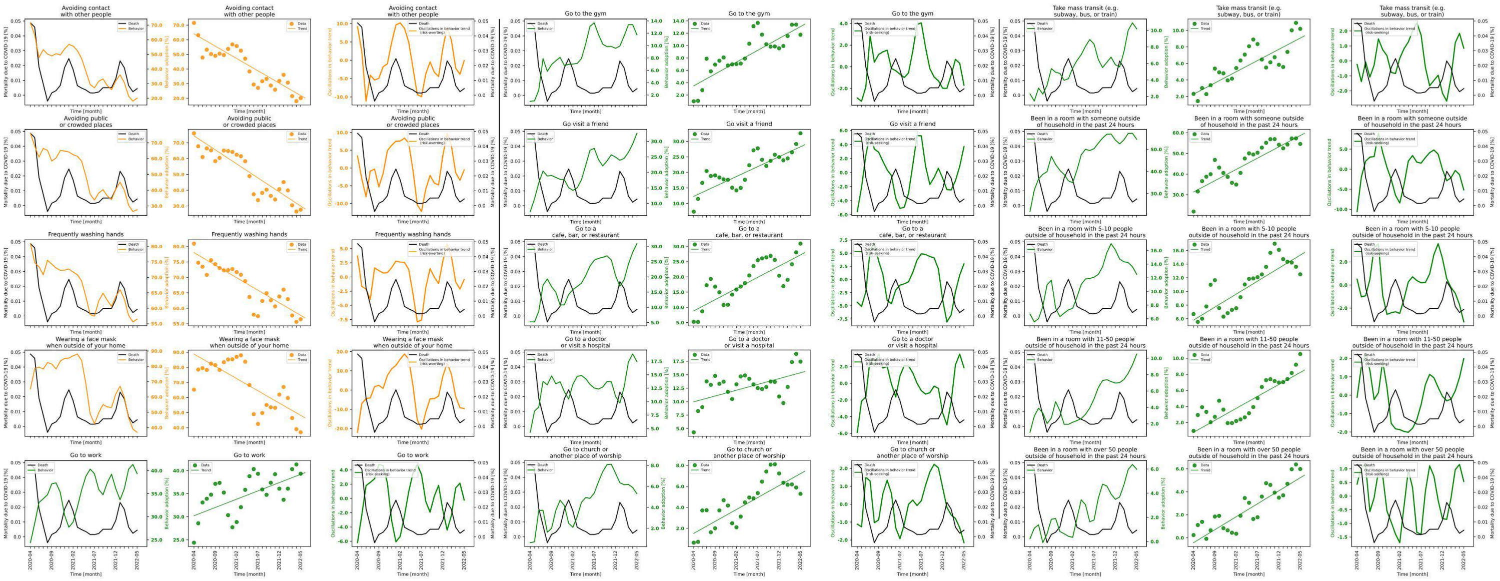

**Figure S7.22 shows behavior and mortality on 1st, 4th and 7th column, behavior trends in 2nd, 5th and 8th column, and behavior oscillatory components juxtaposed with mortality rates on 3rd, 6th and 9th columns for Massachusetts.**

| Behaviors | Slopes | Y-intercepts | Start %($S$) | End %($E$) | $\lvert E-S\rvert$ | $\frac{\lvert E-S\rvert*100}{S}$ |
|---|---|---|---|---|---|---|
| Avoiding contact with other people | -1.685 | 64.038 | 71.504 | 20.132 | 51.372 | 71.845 |
| Avoiding public or crowded places | -1.777 | 74.403 | 76.008 | 27.658 | 48.35 | 63.612 |
| Frequently washing hands | -0.813 | 78.024 | 80.916 | 56.457 | 24.459 | 30.228 |
| Wearing a face mask when outside of your home | -1.625 | 88.799 | 65.227 | 36.936 | 28.291 | 43.373 |
| Go to work | 0.359 | 30.222 | 24.38 | 39.339 | 14.959 | 61.358 |
| Go to the gym | 0.384 | 3.486 | 0.986 | 11.752 | 10.766 | 1091.886 |
| Go visit a friend | 0.639 | 12.29 | 7.366 | 32.617 | 25.251 | 342.805 |
| Go to a cafe, bar, or restaurant | 0.736 | 8.83 | 5.235 | 30.951 | 25.716 | 491.232 |
| Go to a doctor or visit a hospital | 0.216 | 9.965 | 4.302 | 17.438 | 13.136 | 305.346 |
| Go to church or another place of worship | 0.228 | 1.521 | 0.634 | 5.289 | 4.655 | 734.227 |
| Take mass transit (e.g. subway, bus, or train) | 0.272 | 2.277 | 2.312 | 10.194 | 7.882 | 340.917 |
| Been in a room with someone outside of household in the past 24 hours | 1.092 | 31.36 | 21.904 | 54.582 | 32.678 | 149.187 |
| Been in a room with 5-10 people outside of household in the past 24 hours | 0.381 | 5.801 | 6.687 | 12.508 | 5.821 | 87.049 |
| Been in a room with 11-50 people outside of household in the past 24 hours | 0.298 | 0.779 | 0.958 | 10.517 | 9.559 | 997.808 |
| Been in a room with over 50 people outside of household in the past 24 hours | 0.226 | -0.431 | 0.233 | 5.98 | 5.747 | 2466.524 |

**Table S7.22 shows the slopes, y-intercept, adherence % at the beginning of the study period, and at the end, the absolute difference between the beginning and the end, and the absolute difference as a percentage of the adherence % at the beginning of the study period of each behavior's trend in Massachusetts.**

#### State name: Michigan

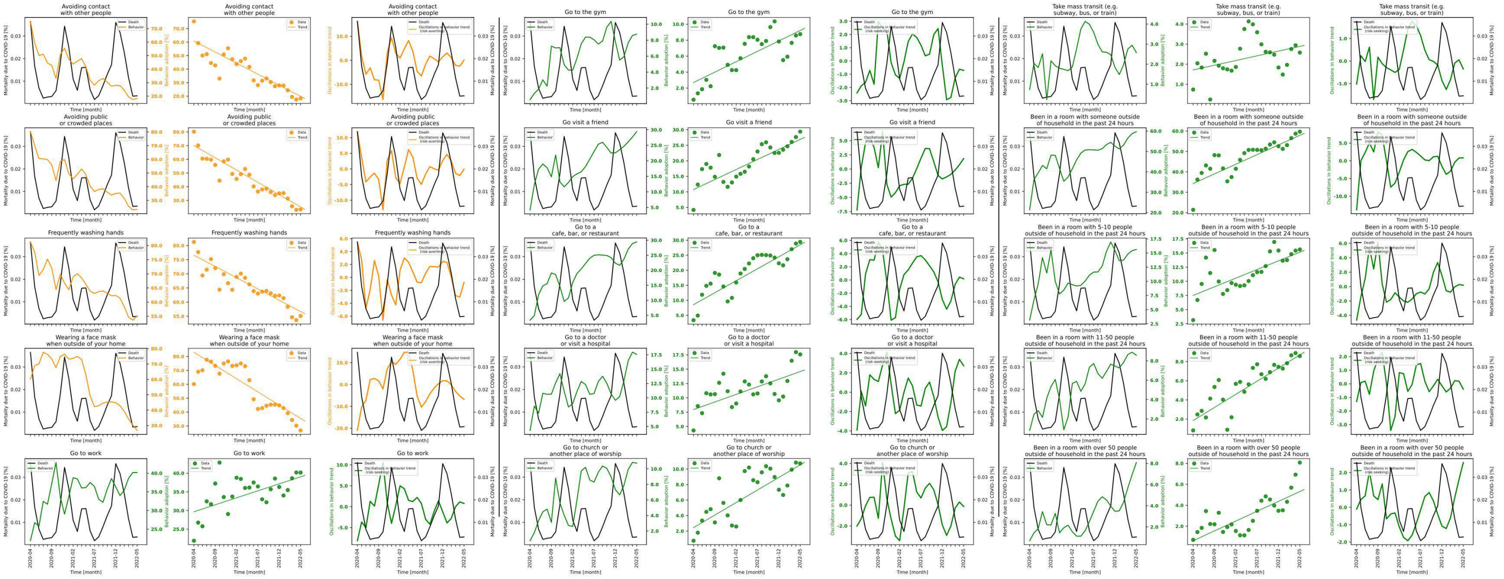

**Figure S7.23 shows behavior and mortality on 1st, 4th and 7th column, behavior trends in 2nd, 5th and 8th column, and behavior oscillatory components juxtaposed with mortality rates on 3rd, 6th and 9th columns for Michigan.**

| Behaviors | Slopes | Y-intercepts | Start %$(S)$ | End %$(E)$ | $\lvert E-S \rvert$ | $\frac{\lvert E-S \rvert *100}{S}$ |
|---|---|---|---|---|---|---|
| Avoiding contact with other people | -1.637 | 60.81 | 75.393 | 18.32 | 57.073 | 75.701 |
| Avoiding public or crowded places | -1.794 | 70.081 | 80.207 | 23.385 | 56.822 | 70.844 |
| Frequently washing hands | -0.796 | 76.569 | 81.259 | 55.114 | 26.145 | 32.175 |
| Wearing a face mask when outside of your home | -1.901 | 83.014 | 60.121 | 26.821 | 33.3 | 55.388 |
| Go to work | 0.374 | 29.669 | 21.913 | 40.168 | 18.255 | 83.307 |
| Go to the gym | 0.26 | 2.694 | 0.543 | 8.753 | 8.21 | 1511.971 |
| Go visit a friend | 0.646 | 10.738 | 4.171 | 29.394 | 25.223 | 604.723 |
| Go to a cafe, bar, or restaurant | 0.797 | 8.559 | 3.383 | 29.448 | 26.065 | 770.47 |
| Go to a doctor or visit a hospital | 0.265 | 7.948 | 4.304 | 17.575 | 13.271 | 308.341 |
| Go to church or another place of worship | 0.324 | 2.444 | 0.753 | 10.73 | 9.977 | 1324.967 |
| Take mass transit (e.g. subway, bus, or train) | 0.047 | 1.713 | 0.738 | 2.579 | 1.841 | 249.458 |
| Been in a room with someone outside of household in the past 24 hours | 0.945 | 34.278 | 21.383 | 59.692 | 38.309 | 179.156 |
| Been in a room with 5-10 people outside of household in the past 24 hours | 0.305 | 7.508 | 3.152 | 15.63 | 12.478 | 395.876 |
| Been in a room with 11-50 people outside of household in the past 24 hours | 0.274 | 1.843 | 0.815 | 8.575 | 7.76 | 952.147 |
| Been in a room with over 50 people outside of household in the past 24 hours | 0.188 | 0.615 | 0.701 | 8.076 | 7.375 | 1052.068 |

**Table S7.23 shows the slopes, y-intercept, adherence % at the beginning of the study period, and at the end, the absolute difference between the beginning and the end, and the absolute difference as a percentage of the adherence % at the beginning of the study period of each behavior's trend in Michigan.**

### State name: Minnesota

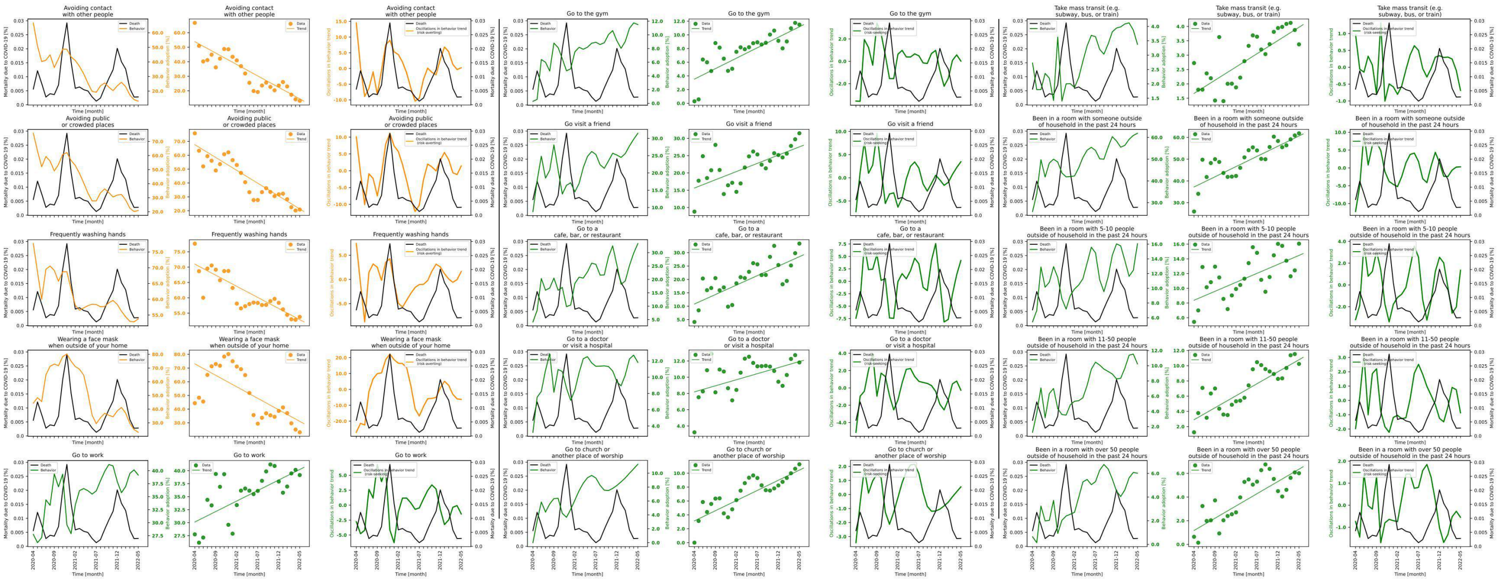

**Figure S7.24 shows behavior and mortality on 1st, 4th and 7th column, behavior trends in 2nd, 5th and 8th column, and behavior oscillatory components juxtaposed with mortality rates on 3rd, 6th and 9th columns for Minnesota.**

| Behaviors | Slopes | Y-intercepts | Start %($S$) | End %($E$) | $\|E-S\|$ | $\frac{\|E-S\|*100}{S}$ |
|---|---|---|---|---|---|---|
| Avoiding contact with other people | -1.575 | 53.838 | 66.776 | 13.114 | 53.662 | 80.361 |
| Avoiding public or crowded places | -1.853 | 67.641 | 75.855 | 20.885 | 54.97 | 72.467 |
| Frequently washing hands | -0.724 | 70.973 | 77.639 | 53.782 | 23.857 | 30.728 |
| Wearing a face mask when outside of your home | -1.679 | 73.043 | 44.358 | 23.094 | 21.264 | 47.937 |
| Go to work | 0.404 | 30.137 | 27.766 | 39.169 | 11.403 | 41.068 |
| Go to the gym | 0.306 | 3.534 | 0.29 | 11.501 | 11.211 | 3865.862 |
| Go visit a friend | 0.476 | 15.643 | 8.811 | 31.481 | 22.67 | 257.292 |
| Go to a cafe, bar, or restaurant | 0.707 | 10.821 | 4.106 | 33.384 | 29.278 | 713.054 |
| Go to a doctor or visit a hospital | 0.149 | 8.197 | 3.219 | 11.817 | 8.598 | 267.102 |
| Go to church or another place of worship | 0.292 | 3.14 | 0.0 | 11.285 | 11.285 | inf |
| Take mass transit (e.g. subway, bus, or train) | 0.092 | 1.668 | 2.725 | 3.376 | 0.651 | 23.89 |
| Been in a room with someone outside of household in the past 24 hours | 0.938 | 37.219 | 25.93 | 61.902 | 35.972 | 138.727 |
| Been in a room with 5-10 people outside of household in the past 24 hours | 0.244 | 8.375 | 5.434 | 16.064 | 10.63 | 195.62 |
| Been in a room with 11-50 people outside of household in the past 24 hours | 0.315 | 2.901 | 1.249 | 10.239 | 8.99 | 719.776 |
| Been in a room with over 50 people outside of household in the past 24 hours | 0.207 | 1.174 | 0.64 | 6.015 | 5.375 | 839.844 |

**Table S7.24 shows the slopes, y-intercept, adherence % at the beginning of the study period, and at the end, the absolute difference between the beginning and the end, and the absolute difference as a percentage of the adherence % at the beginning of the study period of each behavior's trend in Minnesota.**

### State name: Mississippi

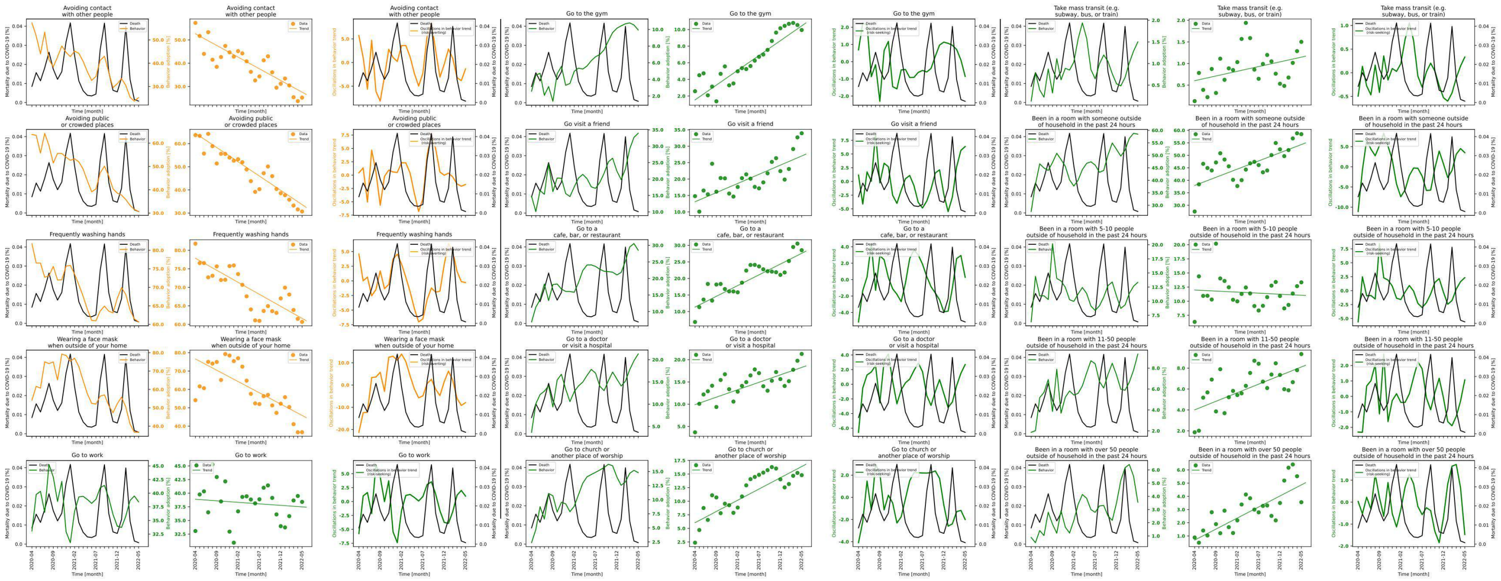

**Figure S7.25 shows behavior and mortality on 1st, 4th and 7th column, behavior trends in 2nd, 5th and 8th column, and behavior oscillatory components juxtaposed with mortality rates on 3rd, 6th and 9th columns for Mississippi.**

| Behaviors | Slopes | Y-intercepts | Start %($S$) | End %($E$) | $\|E-S\|$ | $\frac{\|E-S\|*100}{S}$ |
|---|---|---|---|---|---|---|
| Avoiding contact with other people | -1.002 | 52.623 | 57.297 | 25.241 | 32.056 | 55.947 |
| Avoiding public or crowded places | -1.232 | 64.469 | 63.658 | 30.679 | 32.979 | 51.807 |
| Frequently washing hands | -0.649 | 77.847 | 81.777 | 60.676 | 21.101 | 25.803 |
| Wearing a face mask when outside of your home | -1.232 | 76.608 | 54.114 | 36.68 | 17.434 | 32.217 |
| Go to work | -0.056 | 38.88 | 33.059 | 38.405 | 5.346 | 16.171 |
| Go to the gym | 0.356 | 1.534 | 2.576 | 9.936 | 7.36 | 285.714 |
| Go visit a friend | 0.558 | 13.086 | 14.792 | 33.948 | 19.156 | 129.502 |
| Go to a cafe, bar, or restaurant | 0.649 | 11.36 | 6.854 | 28.522 | 21.668 | 316.137 |
| Go to a doctor or visit a hospital | 0.334 | 9.909 | 3.681 | 21.293 | 17.612 | 478.457 |
| Go to church or another place of worship | 0.413 | 6.047 | 2.372 | 14.794 | 12.422 | 523.693 |
| Take mass transit (e.g. subway, bus, or train) | 0.022 | 0.603 | 0.131 | 1.501 | 1.37 | 1045.802 |
| Been in a room with someone outside of household in the past 24 hours | 0.655 | 37.934 | 27.51 | 58.461 | 30.951 | 112.508 |
| Been in a room with 5-10 people outside of household in the past 24 hours | -0.037 | 11.979 | 6.335 | 13.34 | 7.005 | 110.576 |
| Been in a room with 11-50 people outside of household in the past 24 hours | 0.164 | 4.007 | 1.862 | 9.343 | 7.481 | 401.772 |
| Been in a room with over 50 people outside of household in the past 24 hours | 0.167 | 0.687 | 0.885 | 3.555 | 2.67 | 301.695 |

**Table S7.25 shows the slopes, y-intercept, adherence % at the beginning of the study period, and at the end, the absolute difference between the beginning and the end, and the absolute difference as a percentage of the adherence % at the beginning of the study period of each behavior's trend in Mississippi.**

#### State name: Missouri

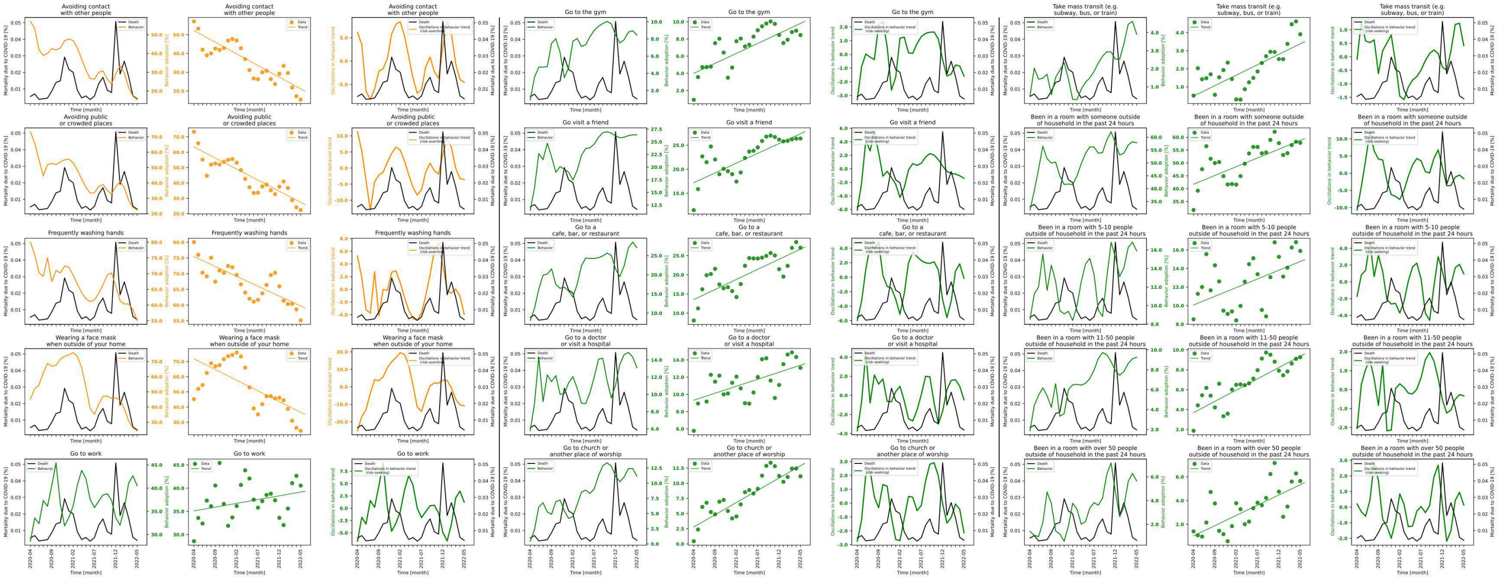

**Figure S7.26 shows behavior and mortality on 1st, 4th and 7th column, behavior trends in 2nd, 5th and 8th column, and behavior oscillatory components juxtaposed with mortality rates on 3rd, 6th and 9th columns for Missouri.**

| Behaviors | Slopes | Y-intercepts | Start %($S$) | End %($E$) | $\lvert E-S \rvert$ | $\frac{\lvert E-S \rvert * 100}{S}$ |
|---|---|---|---|---|---|---|
| Avoiding contact with other people | -1.242 | 52.214 | 57.102 | 15.402 | 41.7 | 73.027 |
| Avoiding public or crowded places | -1.444 | 63.469 | 73.298 | 22.388 | 50.91 | 69.456 |
| Frequently washing hands | -0.629 | 75.43 | 80.015 | 55.125 | 24.89 | 31.107 |
| Wearing a face mask when outside of your home | -1.409 | 71.788 | 45.23 | 24.356 | 20.874 | 46.151 |
| Go to work | 0.163 | 35.102 | 28.579 | 40.554 | 11.975 | 41.901 |
| Go to the gym | 0.232 | 3.998 | 0.917 | 8.453 | 7.536 | 821.81 |
| Go visit a friend | 0.408 | 17.206 | 11.503 | 26.399 | 14.896 | 129.497 |
| Go to a cafe, bar, or restaurant | 0.526 | 13.546 | 8.092 | 27.07 | 18.978 | 234.528 |
| Go to a doctor or visit a hospital | 0.163 | 9.32 | 5.772 | 13.084 | 7.312 | 126.681 |
| Go to church or another place of worship | 0.403 | 2.797 | 0.455 | 11.144 | 10.689 | 2349.231 |
| Take mass transit (e.g. subway, bus, or train) | 0.116 | 0.487 | 0.516 | 3.908 | 3.392 | 657.364 |
| Been in a room with someone outside of household in the past 24 hours | 0.682 | 41.72 | 31.697 | 57.834 | 26.137 | 82.459 |
| Been in a room with 5-10 people outside of household in the past 24 hours | 0.187 | 10.068 | 8.526 | 15.89 | 7.364 | 86.371 |
| Been in a room with 11-50 people outside of household in the past 24 hours | 0.227 | 3.685 | 1.865 | 9.258 | 7.393 | 396.408 |
| Been in a room with over 50 people outside of household in the past 24 hours | 0.172 | 1.029 | 1.406 | 5.648 | 4.242 | 301.707 |

**Table S7.26 shows the slopes, y-intercept, adherence % at the beginning of the study period, and at the end, the absolute difference between the beginning and the end, and the absolute difference as a percentage of the adherence % at the beginning of the study period of each behavior's trend in Missouri.**

### State name: Montana

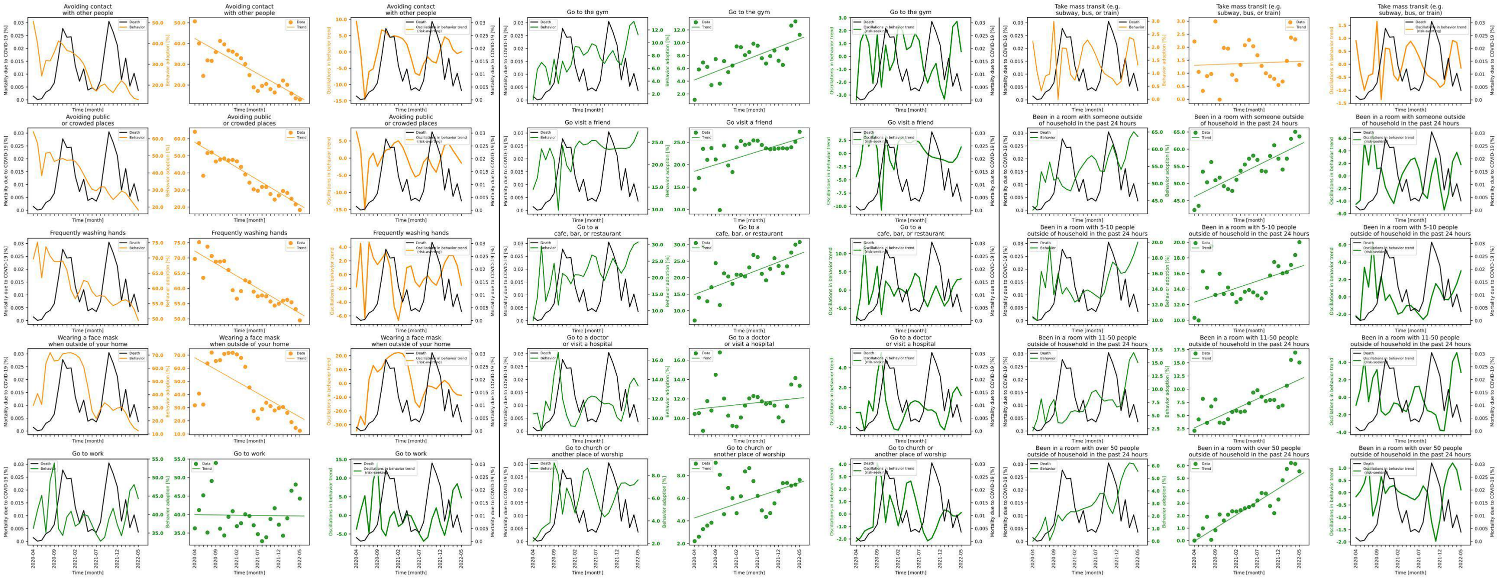

**Figure S7.27 shows behavior and mortality on 1st, 4th and 7th column, behavior trends in 2nd, 5th and 8th column, and behavior oscillatory components juxtaposed with mortality rates on 3rd, 6th and 9th columns for Montana.**

| Behaviors | Slopes | Y-intercepts | Start %($S$) | End %($E$) | $\|E-S\|$ | $\frac{\|E-S\|*100}{S}$ |
|---|---|---|---|---|---|---|
| Avoiding contact with other people | -1.132 | 42.413 | 50.66 | 13.03 | 37.63 | 74.28 |
| Avoiding public or crowded places | -1.468 | 57.875 | 64.07 | 18.278 | 45.792 | 71.472 |
| Frequently washing hands | -0.817 | 72.252 | 69.664 | 49.495 | 20.169 | 28.952 |
| Wearing a face mask when outside of your home | -1.804 | 68.011 | 31.815 | 12.452 | 19.363 | 60.861 |
| Go to work | -0.014 | 39.971 | 36.285 | 44.32 | 8.035 | 22.144 |
| Go to the gym | 0.257 | 4.202 | 1.088 | 11.245 | 10.157 | 933.548 |
| Go visit a friend | 0.292 | 18.549 | 14.462 | 27.362 | 12.9 | 89.199 |
| Go to a cafe, bar, or restaurant | 0.492 | 14.913 | 7.045 | 30.776 | 23.731 | 336.849 |
| Go to a doctor or visit a hospital | 0.046 | 10.922 | 10.45 | 13.351 | 2.901 | 27.761 |
| Go to church or another place of worship | 0.124 | 4.262 | 2.21 | 7.612 | 5.402 | 244.434 |
| Take mass transit (e.g. subway, bus, or train) | 0.006 | 1.301 | 2.218 | 1.319 | 0.899 | 40.532 |
| Been in a room with someone outside of household in the past 24 hours | 0.605 | 46.177 | 42.28 | 63.704 | 21.424 | 50.672 |
| Been in a room with 5-10 people outside of household in the past 24 hours | 0.181 | 12.317 | 10.299 | 20.016 | 9.717 | 94.349 |
| Been in a room with 11-50 people outside of household in the past 24 hours | 0.367 | 2.655 | 2.106 | 15.066 | 12.96 | 615.385 |
| Been in a room with over 50 people outside of household in the past 24 hours | 0.211 | -0.047 | 0.0 | 5.553 | 5.553 | inf |

**Table S7.27 shows the slopes, y-intercept, adherence % at the beginning of the study period, and at the end, the absolute difference between the beginning and the end, and the absolute difference as a percentage of the adherence % at the beginning of the study period of each behavior's trend in Montana.**

### State name: Nebraska

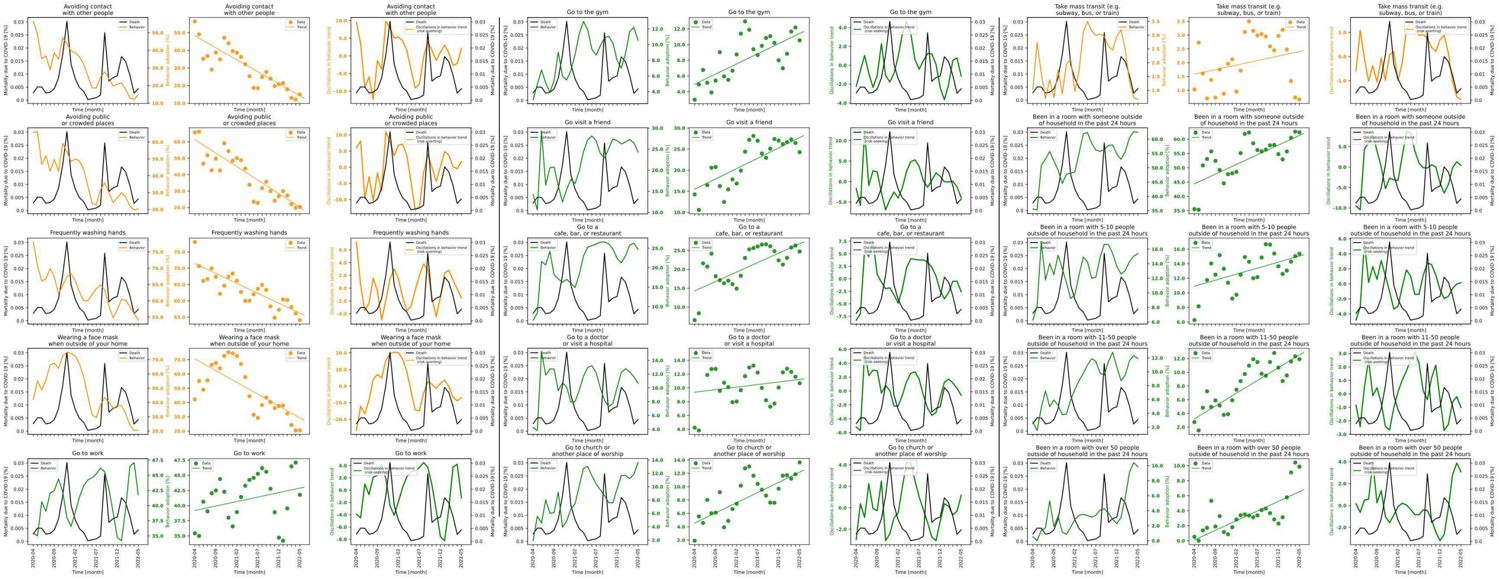

**Figure S7.28 shows behavior and mortality on 1st, 4th and 7th column, behavior trends in 2nd, 5th and 8th column, and behavior oscillatory components juxtaposed with mortality rates on 3rd, 6th and 9th columns for Nebraska.**

| Behaviors | Slopes | Y-intercepts | Start %$(S)$ | End %$(E)$ | $\|E-S\|$ | $\frac{\|E-S\|*100}{S}$ |
|---|---|---|---|---|---|---|
| Avoiding contact with other people | -1.359 | 48.225 | 56.433 | 14.972 | 41.461 | 73.469 |
| Avoiding public or crowded places | -1.645 | 61.339 | 65.71 | 20.331 | 45.379 | 69.06 |
| Frequently washing hands | -0.608 | 71.427 | 78.061 | 54.081 | 23.98 | 30.72 |
| Wearing a face mask when outside of your home | -1.65 | 70.556 | 42.172 | 20.568 | 21.604 | 51.228 |
| Go to work | 0.147 | 39.199 | 35.424 | 41.782 | 6.358 | 17.948 |
| Go to the gym | 0.258 | 4.956 | 2.973 | 10.539 | 7.566 | 254.49 |
| Go visit a friend | 0.49 | 15.483 | 14.254 | 24.277 | 10.023 | 70.317 |
| Go to a cafe, bar, or restaurant | 0.5 | 14.288 | 6.519 | 24.715 | 18.196 | 279.123 |
| Go to a doctor or visit a hospital | 0.074 | 9.369 | 4.229 | 10.672 | 6.443 | 152.353 |
| Go to church or another place of worship | 0.305 | 4.556 | 1.909 | 13.642 | 11.733 | 614.615 |
| Take mass transit (e.g. subway, bus, or train) | 0.033 | 1.587 | 1.03 | 0.665 | 0.365 | 35.437 |
| Been in a room with someone outside of household in the past 24 hours | 0.677 | 44.405 | 35.556 | 62.41 | 26.854 | 75.526 |
| Been in a room with 5-10 people outside of household in the past 24 hours | 0.165 | 10.92 | 6.258 | 15.37 | 9.112 | 145.606 |
| Been in a room with 11-50 people outside of household in the past 24 hours | 0.373 | 3.216 | 2.748 | 11.912 | 9.164 | 333.479 |
| Been in a room with over 50 people outside of household in the past 24 hours | 0.257 | 0.112 | 0.531 | 9.868 | 9.337 | 1758.38 |

**Table S7.28 shows the slopes, y-intercept, adherence % at the beginning of the study period, and at the end, the absolute difference between the beginning and the end, and the absolute difference as a percentage of the adherence % at the beginning of the study period of each behavior's trend in Nebraska.**

### State name: Nevada

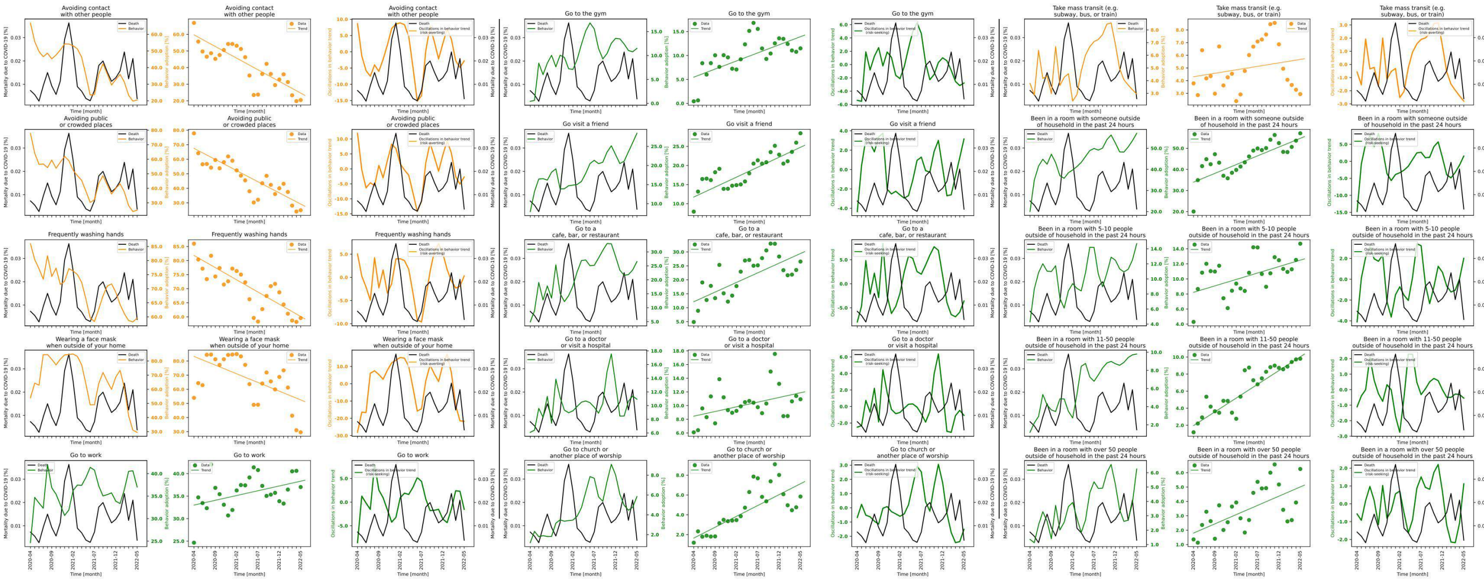

**Figure S7.29 shows behavior and mortality on 1st, 4th and 7th column, behavior trends in 2nd, 5th and 8th column, and behavior oscillatory components juxtaposed with mortality rates on 3rd, 6th and 9th columns for Nevada.**

| Behaviors | Slopes | Y-intercepts | Start %($S$) | End %($E$) | $\|E-S\|$ | $\frac{\|E-S\|*100}{S}$ |
|---|---|---|---|---|---|---|
| Avoiding contact with other people | -1.402 | 59.707 | 66.914 | 20.446 | 46.468 | 69.444 |
| Avoiding public or crowded places | -1.534 | 67.511 | 77.87 | 24.934 | 52.936 | 67.98 |
| Frequently washing hands | -0.868 | 81.822 | 85.997 | 59.561 | 26.436 | 30.741 |
| Wearing a face mask when outside of your home | -1.23 | 83.242 | 53.92 | 29.568 | 24.352 | 45.163 |
| Go to work | 0.212 | 33.014 | 24.66 | 37.02 | 12.36 | 50.122 |
| Go to the gym | 0.338 | 5.478 | 0.434 | 11.496 | 11.062 | 2548.848 |
| Go visit a friend | 0.521 | 11.735 | 7.914 | 28.447 | 20.533 | 259.452 |
| Go to a cafe, bar, or restaurant | 0.691 | 12.143 | 4.885 | 26.539 | 21.654 | 443.275 |
| Go to a doctor or visit a hospital | 0.137 | 8.449 | 6.091 | 10.927 | 4.836 | 79.396 |
| Go to church or another place of worship | 0.219 | 1.65 | 1.188 | 5.842 | 4.654 | 391.751 |
| Take mass transit (e.g. subway, bus, or train) | 0.055 | 4.304 | 3.782 | 2.935 | 0.847 | 22.396 |
| Been in a room with someone outside of household in the past 24 hours | 0.831 | 33.965 | 20.045 | 57.109 | 37.064 | 184.904 |
| Been in a room with 5-10 people outside of household in the past 24 hours | 0.171 | 8.221 | 4.291 | 14.681 | 10.39 | 242.135 |
| Been in a room with 11-50 people outside of household in the past 24 hours | 0.322 | 1.964 | 1.178 | 9.798 | 8.62 | 731.749 |
| Been in a room with over 50 people outside of household in the past 24 hours | 0.128 | 1.799 | 1.357 | 6.258 | 4.901 | 361.164 |

**Table S7.29 shows the slopes, y-intercept, adherence % at the beginning of the study period, and at the end, the absolute difference between the beginning and the end, and the absolute difference as a percentage of the adherence % at the beginning of the study period of each behavior's trend in Nevada.**

### State name: New Hampshire

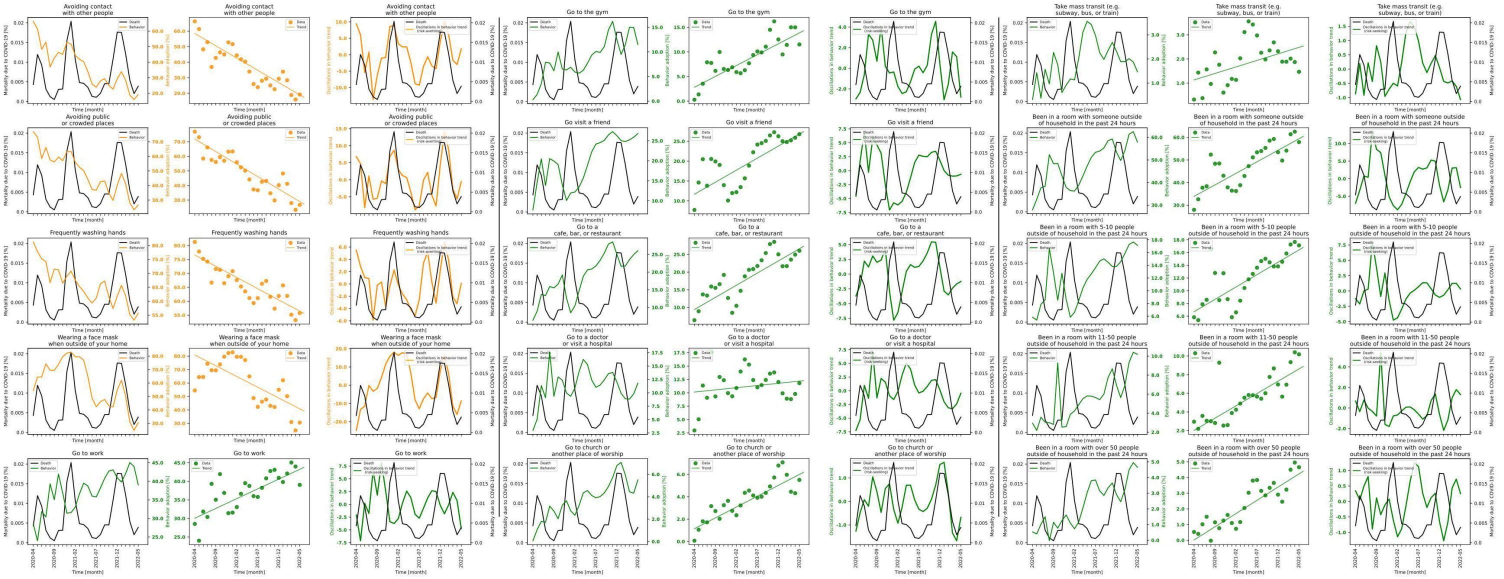

**Figure S7.30 shows behavior and mortality on 1st, 4th and 7th column, behavior trends in 2nd, 5th and 8th column, and behavior oscillatory components juxtaposed with mortality rates on 3rd, 6th and 9th columns for New Hampshire.**

| Behaviors | Slopes | Y-intercepts | Start %($S$) | End %($E$) | $\lvert E-S\rvert$ | $\frac{\lvert E-S\rvert*100}{S}$ |
|---|---|---|---|---|---|---|
| Avoiding contact with other people | -1.578 | 58.429 | 66.258 | 19.193 | 47.065 | 71.033 |
| Avoiding public or crowded places | -1.716 | 71.846 | 76.925 | 26.706 | 50.219 | 65.283 |
| Frequently washing hands | -0.805 | 76.579 | 81.276 | 55.741 | 25.535 | 31.418 |
| Wearing a face mask when outside of your home | -1.602 | 80.854 | 54.49 | 30.591 | 23.899 | 43.859 |
| Go to work | 0.527 | 30.063 | 28.522 | 39.036 | 10.514 | 36.863 |
| Go to the gym | 0.442 | 2.813 | 0.286 | 11.517 | 11.231 | 3926.923 |
| Go visit a friend | 0.606 | 11.671 | 7.712 | 26.8 | 19.088 | 247.51 |
| Go to a cafe, bar, or restaurant | 0.688 | 9.402 | 6.377 | 26.047 | 19.67 | 308.452 |
| Go to a doctor or visit a hospital | 0.082 | 10.139 | 2.964 | 11.81 | 8.846 | 298.448 |
| Go to church or another place of worship | 0.194 | 1.146 | 0.075 | 5.507 | 5.432 | 7242.667 |
| Take mass transit (e.g. subway, bus, or train) | 0.054 | 1.129 | 0.317 | 1.467 | 1.15 | 362.776 |
| Been in a room with someone outside of household in the past 24 hours | 1.02 | 33.887 | 27.937 | 57.889 | 29.952 | 107.213 |
| Been in a room with 5-10 people outside of household in the past 24 hours | 0.388 | 6.708 | 5.84 | 17.132 | 11.292 | 193.356 |
| Been in a room with 11-50 people outside of household in the past 24 hours | 0.264 | 2.018 | 2.95 | 10.183 | 7.233 | 245.186 |
| Been in a room with over 50 people outside of household in the past 24 hours | 0.169 | 0.018 | 0.528 | 4.656 | 4.128 | 781.818 |

**Table S7.30 shows the slopes, y-intercept, adherence % at the beginning of the study period, and at the end, the absolute difference between the beginning and the end, and the absolute difference as a percentage of the adherence % at the beginning of the study period of each behavior's trend in New Hampshire.**

### State name: New Jersey

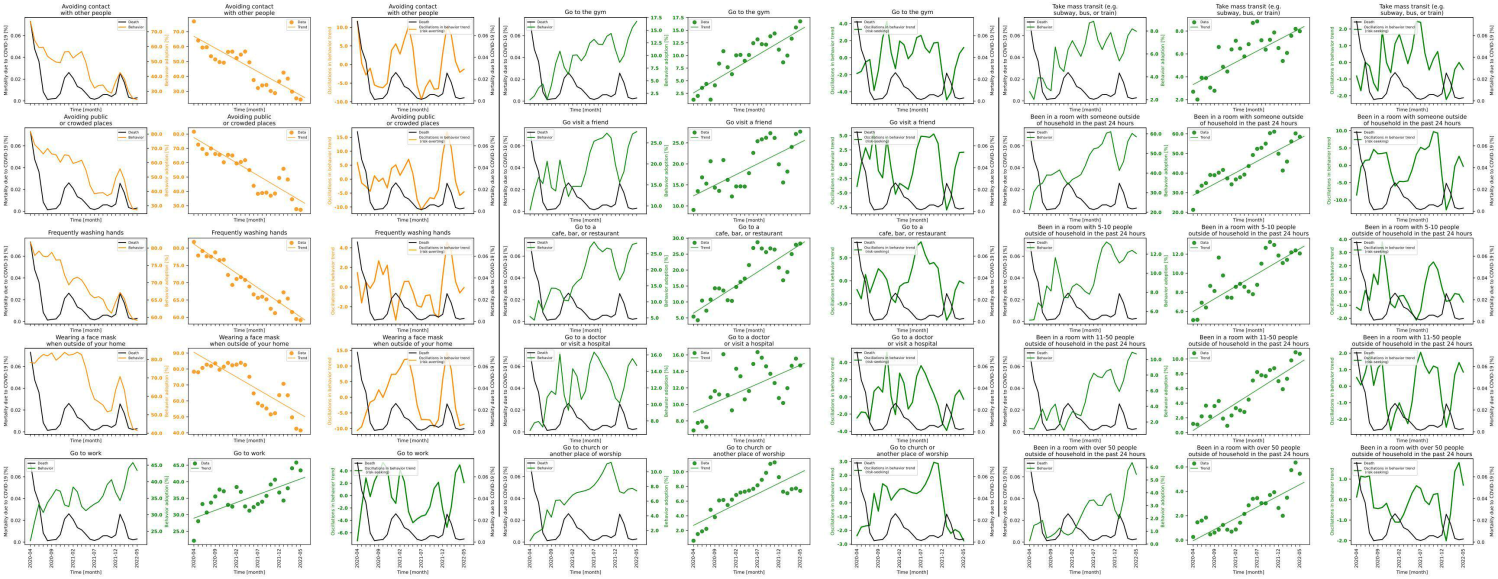

**Figure S7.31 shows behavior and mortality on 1st, 4th and 7th column, behavior trends in 2nd, 5th and 8th column, and behavior oscillatory components juxtaposed with mortality rates on 3rd, 6th and 9th columns for New Jersey.**

| Behaviors | Slopes | Y-intercepts | Start %$(S)$ | End %$(E)$ | $\lvert E-S\rvert$ | $\frac{\lvert E-S\rvert*100}{S}$ |
|---|---|---|---|---|---|---|
| Avoiding contact with other people | -1.582 | 66.892 | 76.9 | 24.493 | 52.407 | 68.15 |
| Avoiding public or crowded places | -1.763 | 77.55 | 81.659 | 27.142 | 54.517 | 66.762 |
| Frequently washing hands | -0.843 | 81.148 | 81.764 | 59.185 | 22.579 | 27.615 |
| Wearing a face mask when outside of your home | -1.553 | 90.496 | 78.332 | 41.525 | 36.807 | 46.988 |
| Go to work | 0.476 | 28.963 | 22.146 | 43.432 | 21.286 | 96.117 |
| Go to the gym | 0.499 | 2.596 | 1.242 | 16.752 | 15.51 | 1248.792 |
| Go visit a friend | 0.507 | 12.395 | 9.038 | 27.682 | 18.644 | 206.285 |
| Go to a cafe, bar, or restaurant | 0.859 | 6.488 | 5.408 | 28.307 | 22.899 | 423.428 |
| Go to a doctor or visit a hospital | 0.228 | 9.066 | 6.845 | 14.726 | 7.881 | 115.135 |
| Go to church or another place of worship | 0.287 | 2.701 | 0.601 | 7.402 | 6.801 | 1131.614 |
| Take mass transit (e.g. subway, bus, or train) | 0.195 | 3.325 | 2.697 | 7.979 | 5.282 | 195.847 |
| Been in a room with someone outside of household in the past 24 hours | 1.157 | 28.674 | 21.266 | 58.479 | 37.213 | 174.988 |
| Been in a room with 5-10 people outside of household in the past 24 hours | 0.263 | 5.988 | 5.076 | 12.068 | 6.992 | 137.746 |
| Been in a room with 11-50 people outside of household in the past 24 hours | 0.368 | 0.291 | 1.18 | 10.701 | 9.521 | 806.864 |
| Been in a room with over 50 people outside of household in the past 24 hours | 0.184 | -0.05 | 0.259 | 5.442 | 5.183 | 2001.158 |

**Table S7.31 shows the slopes, y-intercept, adherence % at the beginning of the study period, and at the end, the absolute difference between the beginning and the end, and the absolute difference as a percentage of the adherence % at the beginning of the study period of each behavior's trend in New Jersey.**

### State name: New Mexico

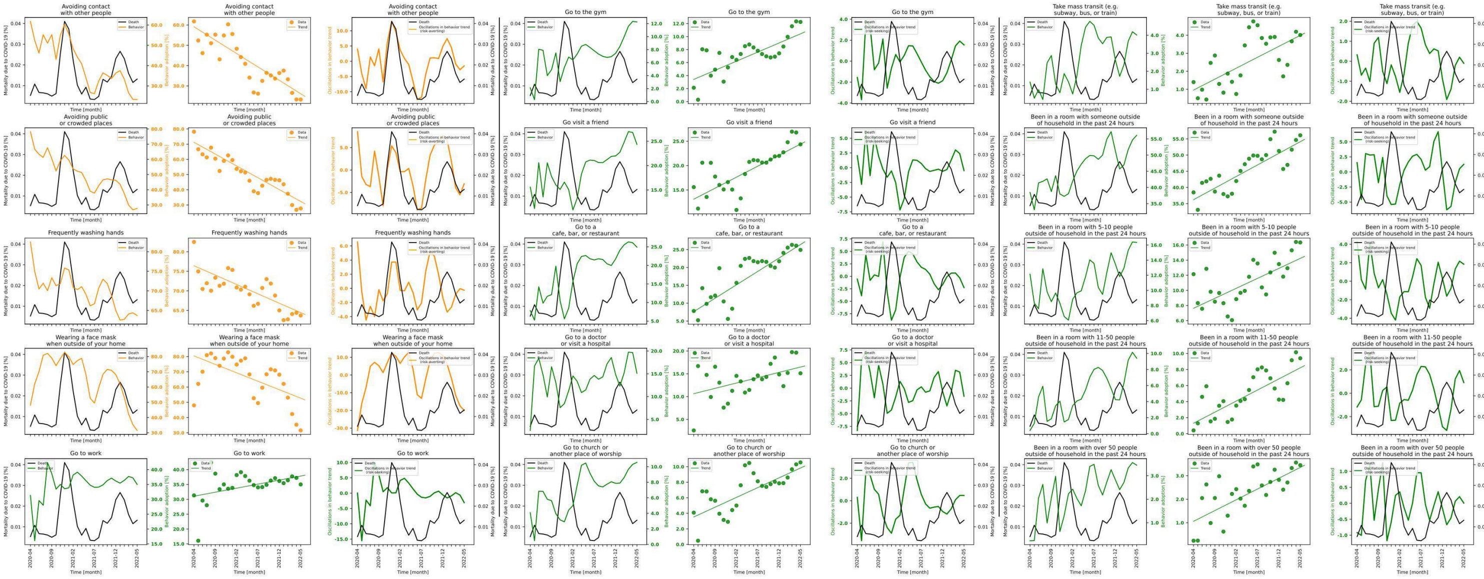

**Figure S7.32 shows behavior and mortality on 1st, 4th and 7th column, behavior trends in 2nd, 5th and 8th column, and behavior oscillatory components juxtaposed with mortality rates on 3rd, 6th and 9th columns for New Mexico.**

| Behaviors | Slopes | Y-intercepts | Start %($S$) | End %($E$) | $\lvert E-S \rvert$ | $\frac{\lvert E-S \rvert * 100}{S}$ |
|---|---|---|---|---|---|---|
| Avoiding contact with other people | -1.319 | 59.12 | 61.786 | 23.301 | 38.485 | 62.288 |
| Avoiding public or crowded places | -1.557 | 71.241 | 78.23 | 27.68 | 50.55 | 64.617 |
| Frequently washing hands | -0.479 | 76.416 | 82.498 | 63.705 | 18.793 | 22.78 |
| Wearing a face mask when outside of your home | -1.108 | 80.42 | 47.995 | 31.551 | 16.444 | 34.262 |
| Go to work | 0.272 | 31.043 | 31.328 | 35.013 | 3.685 | 11.763 |
| Go to the gym | 0.28 | 3.413 | 2.131 | 12.252 | 10.121 | 474.941 |
| Go visit a friend | 0.452 | 13.105 | 15.583 | 24.359 | 8.776 | 56.318 |
| Go to a cafe, bar, or restaurant | 0.752 | 7.629 | 7.819 | 24.92 | 17.101 | 218.711 |
| Go to a doctor or visit a hospital | 0.234 | 10.639 | 2.577 | 15.115 | 12.538 | 486.535 |
| Go to church or another place of worship | 0.252 | 3.53 | 4.086 | 10.555 | 6.469 | 158.321 |
| Take mass transit (e.g. subway, bus, or train) | 0.122 | 0.954 | 1.398 | 4.015 | 2.617 | 187.196 |
| Been in a room with someone outside of household in the past 24 hours | 0.717 | 36.219 | 38.474 | 56.118 | 17.644 | 45.86 |
| Been in a room with 5-10 people outside of household in the past 24 hours | 0.264 | 7.635 | 12.146 | 16.34 | 4.194 | 34.53 |
| Been in a room with 11-50 people outside of household in the past 24 hours | 0.276 | 1.304 | 0.408 | 9.403 | 8.995 | 2204.657 |
| Been in a room with over 50 people outside of household in the past 24 hours | 0.092 | 1.061 | 0.234 | 3.453 | 3.219 | 1375.641 |

**Table S7.32 shows the slopes, y-intercept, adherence % at the beginning of the study period, and at the end, the absolute difference between the beginning and the end, and the absolute difference as a percentage of the adherence % at the beginning of the study period of each behavior's trend in New Mexico.**

### State name: New York

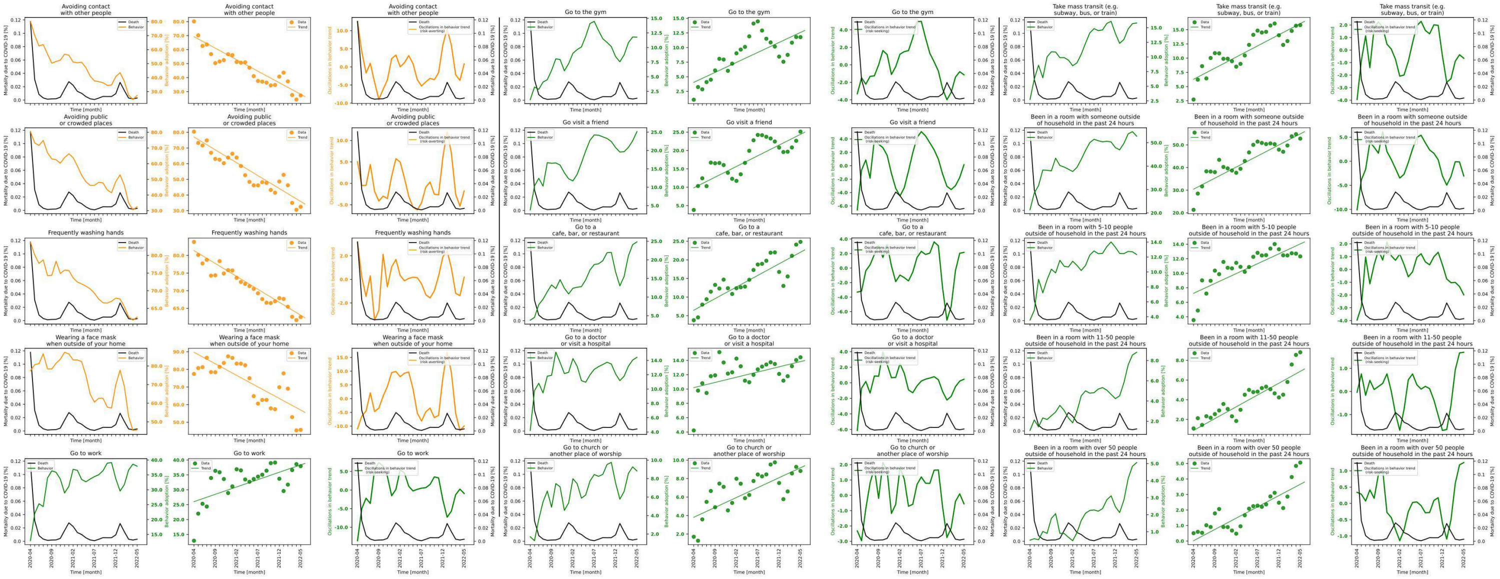

**Figure S7.33 shows behavior and mortality on 1st, 4th and 7th column, behavior trends in 2nd, 5th and 8th column, and behavior oscillatory components juxtaposed with mortality rates on 3rd, 6th and 9th columns for New York.**

| Behaviors | Slopes | Y-intercepts | Start %($S$) | End %($E$) | $\|E-S\|$ | $\frac{\|E-S\|*100}{S}$ |
|---|---|---|---|---|---|---|
| Avoiding contact with other people | -1.646 | 69.243 | 80.168 | 27.295 | 52.873 | 65.953 |
| Avoiding public or crowded places | -1.645 | 77.013 | 80.405 | 32.473 | 47.932 | 59.613 |
| Frequently washing hands | -0.733 | 81.377 | 83.817 | 62.504 | 21.313 | 25.428 |
| Wearing a face mask when outside of your home | -1.311 | 89.708 | 77.421 | 45.67 | 31.751 | 41.011 |
| Go to work | 0.494 | 26.01 | 12.894 | 37.915 | 25.021 | 194.051 |
| Go to the gym | 0.342 | 4.041 | 1.071 | 11.78 | 10.709 | 999.907 |
| Go visit a friend | 0.583 | 9.83 | 3.812 | 25.144 | 21.332 | 559.601 |
| Go to a cafe, bar, or restaurant | 0.623 | 6.475 | 3.786 | 24.834 | 21.048 | 555.943 |
| Go to a doctor or visit a hospital | 0.145 | 10.197 | 4.217 | 14.432 | 10.215 | 242.234 |
| Go to church or another place of worship | 0.214 | 3.812 | 1.704 | 8.826 | 7.122 | 417.958 |
| Take mass transit (e.g. subway, bus, or train) | 0.392 | 6.281 | 2.725 | 15.807 | 13.082 | 480.073 |
| Been in a room with someone outside of household in the past 24 hours | 0.978 | 30.431 | 21.32 | 52.782 | 31.462 | 147.57 |
| Been in a room with 5-10 people outside of household in the past 24 hours | 0.272 | 7.239 | 3.548 | 12.325 | 8.777 | 247.379 |
| Been in a room with 11-50 people outside of household in the past 24 hours | 0.24 | 0.888 | 1.092 | 8.815 | 7.723 | 707.234 |
| Been in a room with over 50 people outside of household in the past 24 hours | 0.146 | 0.021 | 0.506 | 5.059 | 4.553 | 899.802 |

**Table S7.33 shows the slopes, y-intercept, adherence % at the beginning of the study period, and at the end, the absolute difference between the beginning and the end, and the absolute difference as a percentage of the adherence % at the beginning of the study period of each behavior's trend in New York.**

### State name: North Carolina

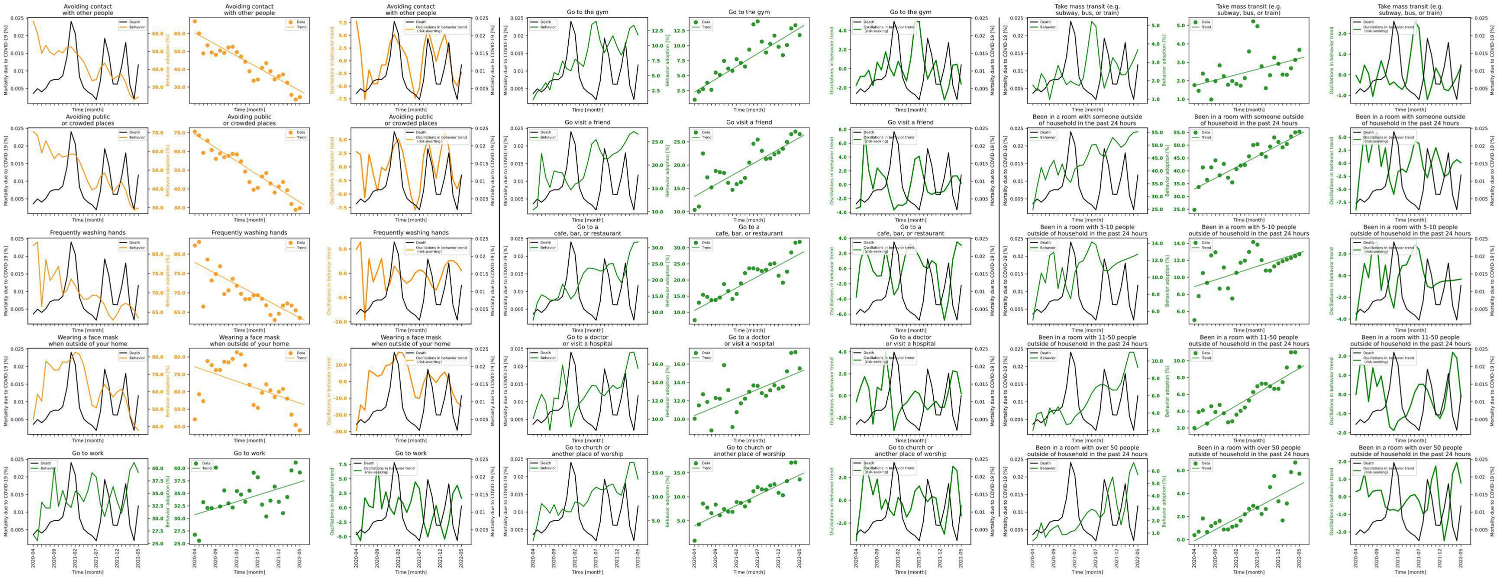

**Figure S7.34 shows behavior and mortality on 1st, 4th and 7th column, behavior trends in 2nd, 5th and 8th column, and behavior oscillatory components juxtaposed with mortality rates on 3rd, 6th and 9th columns for North Carolina.**

| Behaviors | Slopes | Y-intercepts | Start %($S$) | End %($E$) | $\|E-S\|$ | $\frac{\|E-S\|*100}{S}$ |
|---|---|---|---|---|---|---|
| Avoiding contact with other people | -1.3 | 60.432 | 66.877 | 24.335 | 42.542 | 63.612 |
| Avoiding public or crowded places | -1.454 | 69.228 | 70.538 | 29.754 | 40.784 | 57.818 |
| Frequently washing hands | -0.572 | 77.838 | 82.226 | 63.523 | 18.703 | 22.746 |
| Wearing a face mask when outside of your home | -0.837 | 74.352 | 44.162 | 37.813 | 6.349 | 14.377 |
| Go to work | 0.261 | 30.748 | 26.752 | 39.164 | 12.412 | 46.397 |
| Go to the gym | 0.427 | 2.272 | 0.983 | 11.766 | 10.783 | 1096.948 |
| Go visit a friend | 0.504 | 13.307 | 10.375 | 26.64 | 16.265 | 156.771 |
| Go to a cafe, bar, or restaurant | 0.699 | 10.587 | 7.528 | 31.818 | 24.29 | 322.662 |
| Go to a doctor or visit a hospital | 0.19 | 10.398 | 10.048 | 15.532 | 5.484 | 54.578 |
| Go to church or another place of worship | 0.423 | 3.989 | 0.865 | 13.634 | 12.769 | 1476.185 |
| Take mass transit (e.g. subway, bus, or train) | 0.059 | 1.74 | 1.763 | 3.672 | 1.909 | 108.281 |
| Been in a room with someone outside of household in the past 24 hours | 0.856 | 32.887 | 24.79 | 55.268 | 30.478 | 122.945 |
| Been in a room with 5-10 people outside of household in the past 24 hours | 0.159 | 8.901 | 4.961 | 12.719 | 7.758 | 156.38 |
| Been in a room with 11-50 people outside of household in the past 24 hours | 0.296 | 1.705 | 1.997 | 9.273 | 7.276 | 364.347 |
| Been in a room with over 50 people outside of household in the past 24 hours | 0.193 | -0.098 | 0.386 | 5.713 | 5.327 | 1380.052 |

**Table S7.34 shows the slopes, y-intercept, adherence % at the beginning of the study period, and at the end, the absolute difference between the beginning and the end, and the absolute difference as a percentage of the adherence % at the beginning of the study period of each behavior's trend in North Carolina.**

#### State name: North Dakota

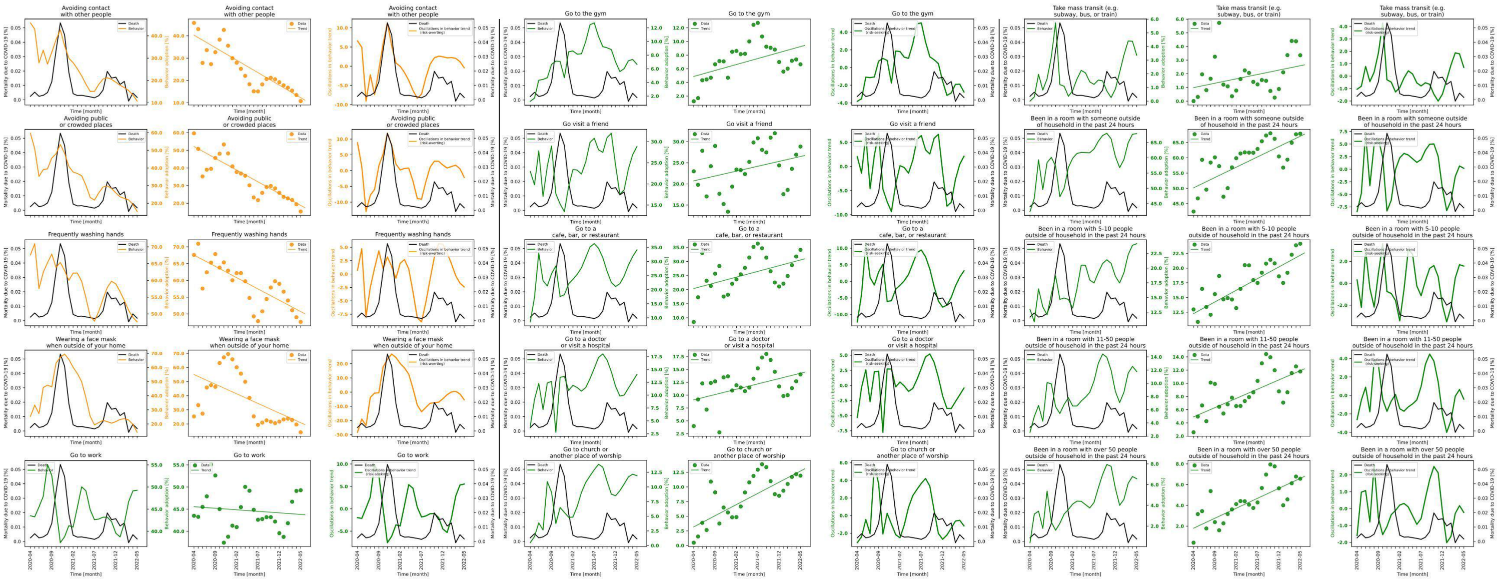

**Figure S7.35 shows behavior and mortality on 1st, 4th and 7th column, behavior trends in 2nd, 5th and 8th column, and behavior oscillatory components juxtaposed with mortality rates on 3rd, 6th and 9th columns for North Dakota.**

| Behaviors | Slopes | Y-intercepts | Start %($S$) | End %($E$) | $\|E-S\|$ | $\frac{\|E-S\|*100}{S}$ |
|---|---|---|---|---|---|---|
| Avoiding contact with other people | -1.12 | 40.297 | 45.767 | 10.766 | 35.001 | 76.477 |
| Avoiding public or crowded places | -1.335 | 52.181 | 59.672 | 15.315 | 44.357 | 74.335 |
| Frequently washing hands | -0.674 | 67.559 | 67.588 | 47.619 | 19.969 | 29.545 |
| Wearing a face mask when outside of your home | -1.364 | 54.979 | 25.317 | 14.1 | 11.217 | 44.306 |
| Go to work | -0.069 | 45.55 | 43.505 | 49.274 | 5.769 | 13.261 |
| Go to the gym | 0.175 | 4.876 | 1.235 | 6.65 | 5.415 | 438.462 |
| Go visit a friend | 0.231 | 20.72 | 22.977 | 28.844 | 5.867 | 25.534 |
| Go to a cafe, bar, or restaurant | 0.406 | 20.398 | 8.523 | 34.12 | 25.597 | 300.329 |
| Go to a doctor or visit a hospital | 0.207 | 9.082 | 3.983 | 14.035 | 10.052 | 252.373 |
| Go to church or another place of worship | 0.381 | 3.182 | 0.483 | 11.925 | 11.442 | 2368.944 |
| Take mass transit (e.g. subway, bus, or train) | 0.064 | 0.989 | 0.0 | 3.356 | 3.356 | inf |
| Been in a room with someone outside of household in the past 24 hours | 0.675 | 50.177 | 42.433 | 67.919 | 25.486 | 60.062 |
| Been in a room with 5-10 people outside of household in the past 24 hours | 0.397 | 12.26 | 12.972 | 24.155 | 11.183 | 86.209 |
| Been in a room with 11-50 people outside of household in the past 24 hours | 0.28 | 4.913 | 2.577 | 11.791 | 9.214 | 357.548 |
| Been in a room with over 50 people outside of household in the past 24 hours | 0.192 | 1.8 | 0.402 | 6.571 | 6.169 | 1534.577 |

**Table S7.35 shows the slopes, y-intercept, adherence % at the beginning of the study period, and at the end, the absolute difference between the beginning and the end, and the absolute difference as a percentage of the adherence % at the beginning of the study period of each behavior's trend in North Dakota.**

### State name: Ohio

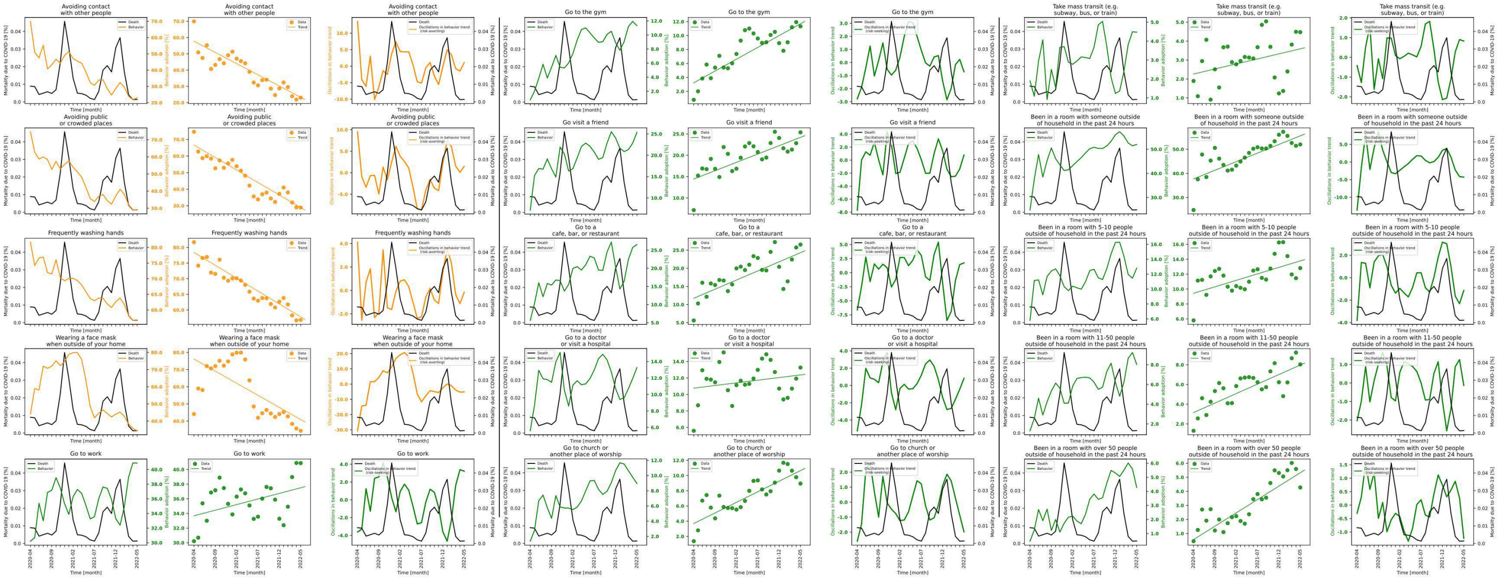

**Figure S7.36 shows behavior and mortality on 1st, 4th and 7th column, behavior trends in 2nd, 5th and 8th column, and behavior oscillatory components juxtaposed with mortality rates on 3rd, 6th and 9th columns for Ohio.**

| Behaviors | Slopes | Y-intercepts | Start %($S$) | End %($E$) | $\|E-S\|$ | $\frac{\|E-S\|*100}{S}$ |
|---|---|---|---|---|---|---|
| Avoiding contact with other people | -1.376 | 57.736 | 69.932 | 23.037 | 46.895 | 67.058 |
| Avoiding public or crowded places | -1.507 | 66.644 | 74.653 | 28.945 | 45.708 | 61.227 |
| Frequently washing hands | -0.825 | 78.402 | 81.696 | 56.81 | 24.886 | 30.462 |
| Wearing a face mask when outside of your home | -1.411 | 76.157 | 44.109 | 34.345 | 9.764 | 22.136 |
| Go to work | 0.153 | 33.681 | 30.214 | 40.875 | 10.661 | 35.285 |
| Go to the gym | 0.339 | 3.227 | 0.791 | 11.321 | 10.53 | 1331.226 |
| Go visit a friend | 0.39 | 14.446 | 7.146 | 25.376 | 18.23 | 255.108 |
| Go to a cafe, bar, or restaurant | 0.503 | 11.737 | 5.613 | 26.544 | 20.931 | 372.902 |
| Go to a doctor or visit a hospital | 0.064 | 10.77 | 5.587 | 13.268 | 7.681 | 137.48 |
| Go to church or another place of worship | 0.278 | 3.691 | 1.367 | 8.944 | 7.577 | 554.279 |
| Take mass transit (e.g. subway, bus, or train) | 0.053 | 2.262 | 1.896 | 4.473 | 2.577 | 135.918 |
| Been in a room with someone outside of household in the past 24 hours | 0.714 | 37.721 | 24.818 | 51.995 | 27.177 | 109.505 |
| Been in a room with 5-10 people outside of household in the past 24 hours | 0.174 | 9.43 | 5.796 | 12.813 | 7.017 | 121.066 |
| Been in a room with 11-50 people outside of household in the past 24 hours | 0.193 | 3.15 | 1.305 | 8.036 | 6.731 | 515.785 |
| Been in a room with over 50 people outside of household in the past 24 hours | 0.19 | 0.568 | 0.455 | 4.276 | 3.821 | 839.78 |

**Table S7.36 shows the slopes, y-intercept, adherence % at the beginning of the study period, and at the end, the absolute difference between the beginning and the end, and the absolute difference as a percentage of the adherence % at the beginning of the study period of each behavior's trend in Ohio.**

### State name: Oklahoma

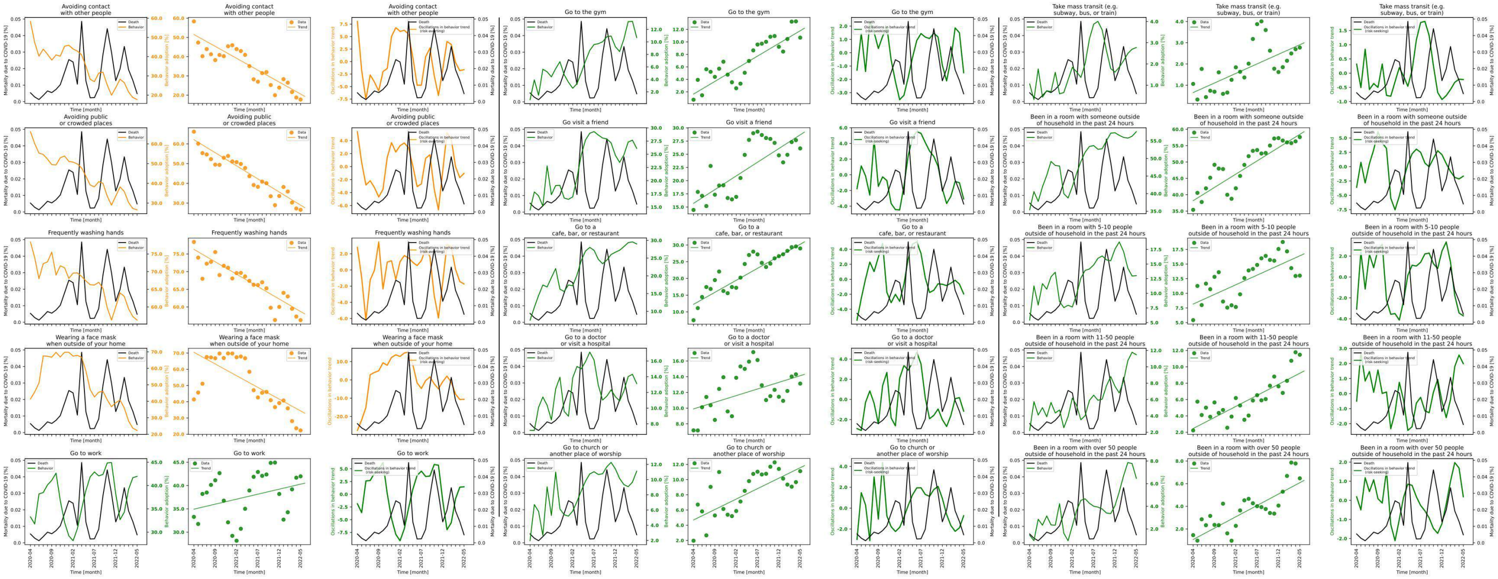

**Figure S7.37 shows behavior and mortality on 1st, 4th and 7th column, behavior trends in 2nd, 5th and 8th column, and behavior oscillatory components juxtaposed with mortality rates on 3rd, 6th and 9th columns for Oklahoma.**

| Behaviors | Slopes | Y-intercepts | Start %($S$) | End %($E$) | $\|E-S\|$ | $\frac{\|E-S\|*100}{S}$ |
|---|---|---|---|---|---|---|
| Avoiding contact with other people | -1.239 | 51.54 | 58.344 | 17.702 | 40.642 | 69.659 |
| Avoiding public or crowded places | -1.326 | 62.132 | 66.179 | 26.599 | 39.58 | 59.807 |
| Frequently washing hands | -0.704 | 76.304 | 78.4 | 56.279 | 22.121 | 28.216 |
| Wearing a face mask when outside of your home | -1.445 | 70.481 | 41.414 | 22.325 | 19.089 | 46.093 |
| Go to work | 0.215 | 34.934 | 33.281 | 41.98 | 8.699 | 26.138 |
| Go to the gym | 0.406 | 1.644 | 0.742 | 10.685 | 9.943 | 1340.027 |
| Go visit a friend | 0.519 | 15.7 | 14.446 | 26.107 | 11.661 | 80.721 |
| Go to a cafe, bar, or restaurant | 0.719 | 12.249 | 7.502 | 28.976 | 21.474 | 286.244 |
| Go to a doctor or visit a hospital | 0.169 | 9.905 | 7.174 | 13.122 | 5.948 | 82.911 |
| Go to church or another place of worship | 0.275 | 4.689 | 1.963 | 11.116 | 9.153 | 466.276 |
| Take mass transit (e.g. subway, bus, or train) | 0.09 | 0.653 | 1.061 | 2.774 | 1.713 | 161.451 |
| Been in a room with someone outside of household in the past 24 hours | 0.811 | 38.184 | 35.391 | 57.617 | 22.226 | 62.801 |
| Been in a room with 5-10 people outside of household in the past 24 hours | 0.33 | 8.168 | 5.424 | 13.025 | 7.601 | 140.136 |
| Been in a room with 11-50 people outside of household in the past 24 hours | 0.27 | 2.456 | 2.224 | 11.475 | 9.251 | 415.962 |
| Been in a room with over 50 people outside of household in the past 24 hours | 0.201 | 1.07 | 1.455 | 6.473 | 5.018 | 344.88 |

**Table S7.37 shows the slopes, y-intercept, adherence % at the beginning of the study period, and at the end, the absolute difference between the beginning and the end, and the absolute difference as a percentage of the adherence % at the beginning of the study period of each behavior's trend in Oklahoma.**

### State name: Oregon

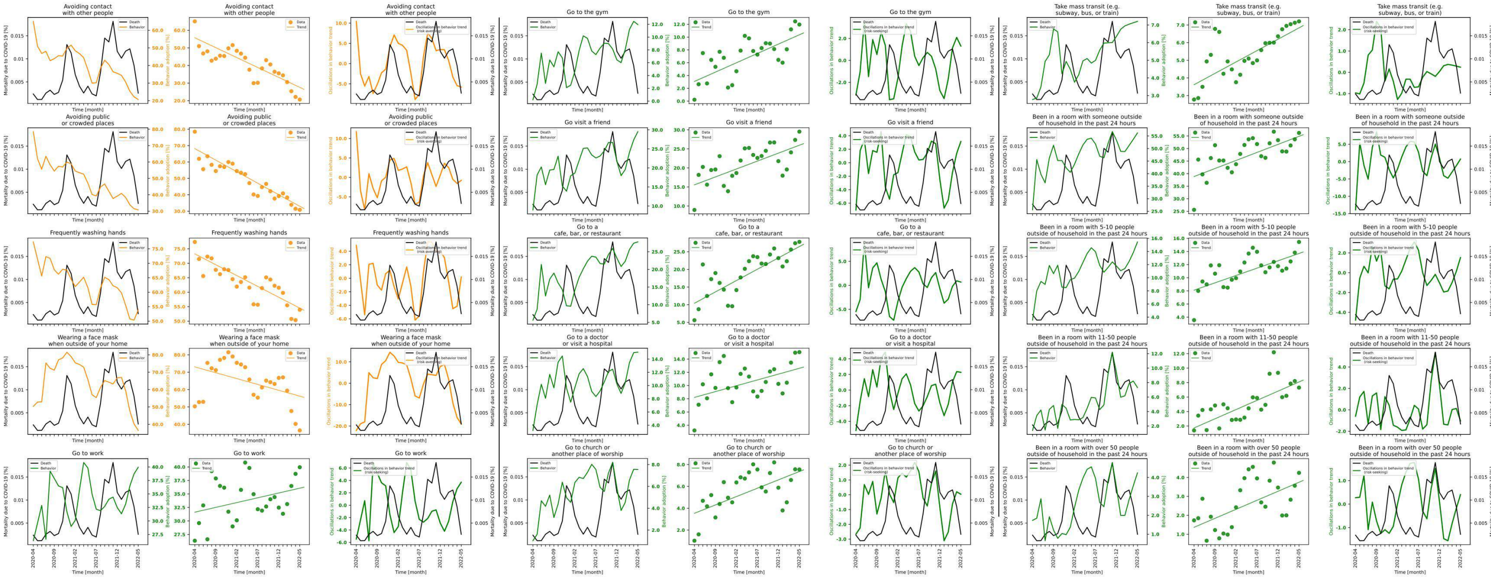

**Figure S7.38 shows behavior and mortality on 1st, 4th and 7th column, behavior trends in 2nd, 5th and 8th column, and behavior oscillatory components juxtaposed with mortality rates on 3rd, 6th and 9th columns for Oregon.**

| Behaviors | Slopes | Y-intercepts | Start %($S$) | End %($E$) | $\lvert E-S \rvert$ | $\frac{\lvert E-S \rvert * 100}{S}$ |
|---|---|---|---|---|---|---|
| Avoiding contact with other people | -1.126 | 55.744 | 65.049 | 20.671 | 44.378 | 68.222 |
| Avoiding public or crowded places | -1.406 | 68.161 | 78.512 | 30.827 | 47.685 | 60.736 |
| Frequently washing hands | -0.75 | 73.241 | 77.365 | 53.93 | 23.435 | 30.291 |
| Wearing a face mask when outside of your home | -0.674 | 73.064 | 50.349 | 36.49 | 13.859 | 27.526 |
| Go to work | 0.178 | 31.626 | 26.343 | 39.946 | 13.603 | 51.638 |
| Go to the gym | 0.291 | 3.073 | 0.219 | 11.96 | 11.741 | 5361.187 |
| Go visit a friend | 0.416 | 15.554 | 8.961 | 29.534 | 20.573 | 229.584 |
| Go to a cafe, bar, or restaurant | 0.642 | 10.418 | 5.616 | 27.792 | 22.176 | 394.872 |
| Go to a doctor or visit a hospital | 0.176 | 8.204 | 3.206 | 15.053 | 11.847 | 369.526 |
| Go to church or another place of worship | 0.153 | 3.537 | 1.026 | 7.55 | 6.524 | 635.867 |
| Take mass transit (e.g. subway, bus, or train) | 0.129 | 3.617 | 2.776 | 7.206 | 4.43 | 159.582 |
| Been in a room with someone outside of household in the past 24 hours | 0.641 | 38.849 | 25.581 | 56.225 | 30.644 | 119.792 |
| Been in a room with 5-10 people outside of household in the past 24 hours | 0.223 | 8.111 | 3.549 | 15.449 | 11.9 | 335.306 |
| Been in a room with 11-50 people outside of household in the past 24 hours | 0.254 | 1.731 | 1.391 | 7.311 | 5.92 | 425.593 |
| Been in a room with over 50 people outside of household in the past 24 hours | 0.095 | 1.365 | 1.732 | 4.244 | 2.512 | 145.035 |

**Table S7.38 shows the slopes, y-intercept, adherence % at the beginning of the study period, and at the end, the absolute difference between the beginning and the end, and the absolute difference as a percentage of the adherence % at the beginning of the study period of each behavior's trend in Oregon.**

### State name: Pennsylvania

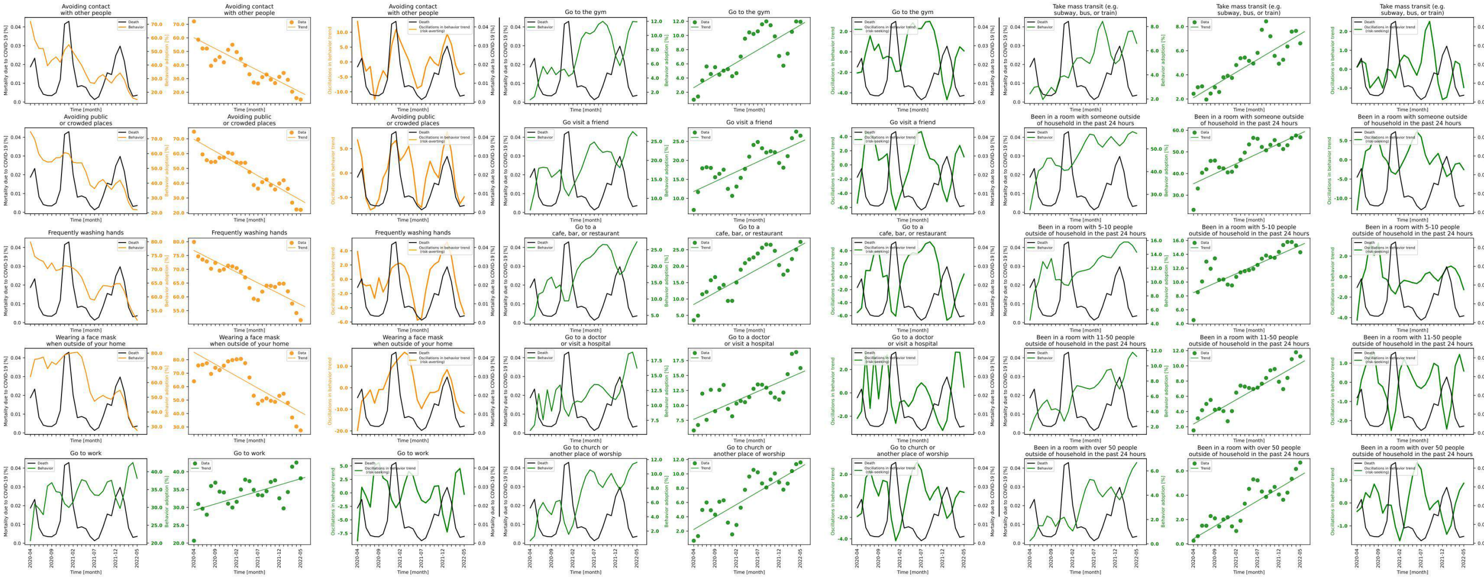

Figure S7.39 shows behavior and mortality on 1st, 4th and 7th column, behavior trends in 2nd, 5th and 8th column, and behavior oscillatory components juxtaposed with mortality rates on 3rd, 6th and 9th columns for Pennsylvania.

| Behaviors | Slopes | Y-intercepts | Start %($S$) | End %($E$) | $\|E-S\|$ | $\frac{\|E-S\|*100}{S}$ |
|---|---|---|---|---|---|---|
| Avoiding contact with other people | -1.606 | 60.084 | 72.118 | 14.638 | 57.48 | 79.703 |
| Avoiding public or crowded places | -1.646 | 69.685 | 74.817 | 21.993 | 52.824 | 70.604 |
| Frequently washing hands | -0.784 | 76.803 | 79.914 | 51.551 | 28.363 | 35.492 |
| Wearing a face mask when outside of your home | -1.782 | 85.529 | 63.969 | 27.459 | 36.51 | 57.075 |
| Go to work | 0.354 | 29.187 | 20.671 | 38.147 | 17.476 | 84.544 |
| Go to the gym | 0.353 | 2.646 | 0.939 | 11.927 | 10.988 | 1170.181 |
| Go visit a friend | 0.521 | 11.818 | 6.958 | 26.507 | 19.549 | 280.957 |
| Go to a cafe, bar, or restaurant | 0.718 | 8.343 | 3.537 | 27.4 | 23.863 | 674.668 |
| Go to a doctor or visit a hospital | 0.31 | 7.678 | 5.814 | 16.243 | 10.429 | 179.377 |
| Go to church or another place of worship | 0.348 | 2.226 | 0.651 | 11.606 | 10.955 | 1682.796 |
| Take mass transit (e.g. subway, bus, or train) | 0.21 | 2.108 | 2.434 | 6.61 | 4.176 | 171.569 |
| Been in a room with someone outside of household in the past 24 hours | 0.925 | 35.294 | 23.225 | 56.801 | 33.576 | 144.568 |
| Been in a room with 5-10 people outside of household in the past 24 hours | 0.273 | 8.477 | 4.505 | 14.304 | 9.799 | 217.514 |
| Been in a room with 11-50 people outside of household in the past 24 hours | 0.319 | 2.36 | 1.516 | 11.243 | 9.727 | 641.623 |
| Been in a room with over 50 people outside of household in the past 24 hours | 0.208 | 0.422 | 0.259 | 6.683 | 6.424 | 2480.309 |

Table S7.39 shows the slopes, y-intercept, adherence % at the beginning of the study period, and at the end, the absolute difference between the beginning and the end, and the absolute difference as a percentage of the adherence % at the beginning of the study period of each behavior's trend in Pennsylvania.

### State name: Rhode Island

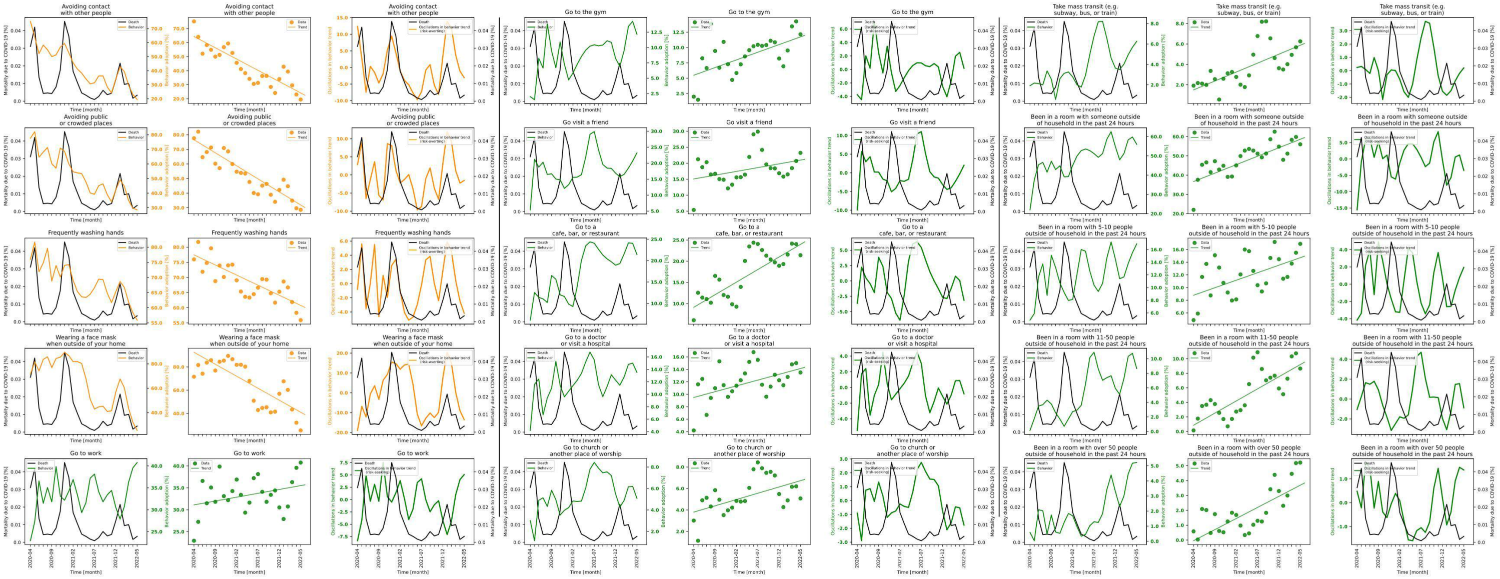

**Figure S7.40 shows behavior and mortality on 1st, 4th and 7th column, behavior trends in 2nd, 5th and 8th column, and behavior oscillatory components juxtaposed with mortality rates on 3rd, 6th and 9th columns for Rhode Island.**

| Behaviors | Slopes | Y-intercepts | Start %($S$) | End %($E$) | $\|E-S\|$ | $\frac{\|E-S\|*100}{S}$ |
|---|---|---|---|---|---|---|
| Avoiding contact with other people | -1.61 | 64.413 | 75.179 | 19.467 | 55.712 | 74.106 |
| Avoiding public or crowded places | -1.786 | 76.551 | 77.614 | 28.581 | 49.033 | 63.175 |
| Frequently washing hands | -0.667 | 77.443 | 75.959 | 55.924 | 20.035 | 26.376 |
| Wearing a face mask when outside of your home | -2.062 | 92.587 | 71.563 | 25.422 | 46.141 | 64.476 |
| Go to work | 0.176 | 31.108 | 22.936 | 40.771 | 17.835 | 77.76 |
| Go to the gym | 0.25 | 5.483 | 1.984 | 12.157 | 10.173 | 512.752 |
| Go visit a friend | 0.236 | 15.059 | 5.353 | 23.22 | 17.867 | 333.775 |
| Go to a cafe, bar, or restaurant | 0.594 | 9.129 | 6.077 | 21.414 | 15.337 | 252.378 |
| Go to a doctor or visit a hospital | 0.186 | 9.49 | 4.19 | 13.503 | 9.313 | 222.267 |
| Go to church or another place of worship | 0.117 | 3.803 | 3.013 | 5.075 | 2.062 | 68.437 |
| Take mass transit (e.g. subway, bus, or train) | 0.175 | 1.505 | 1.919 | 6.259 | 4.34 | 226.159 |
| Been in a room with someone outside of household in the past 24 hours | 0.878 | 36.66 | 21.989 | 56.006 | 34.017 | 154.7 |
| Been in a room with 5-10 people outside of household in the past 24 hours | 0.237 | 8.789 | 4.865 | 16.894 | 12.029 | 247.256 |
| Been in a room with 11-50 people outside of household in the past 24 hours | 0.333 | 0.858 | 0.146 | 8.647 | 8.501 | 5822.603 |
| Been in a room with over 50 people outside of household in the past 24 hours | 0.149 | -0.045 | 0.585 | 5.216 | 4.631 | 791.624 |

**Table S7.40 shows the slopes, y-intercept, adherence % at the beginning of the study period, and at the end, the absolute difference between the beginning and the end, and the absolute difference as a percentage of the adherence % at the beginning of the study period of each behavior's trend in Rhode Island.**

### State name: South Carolina

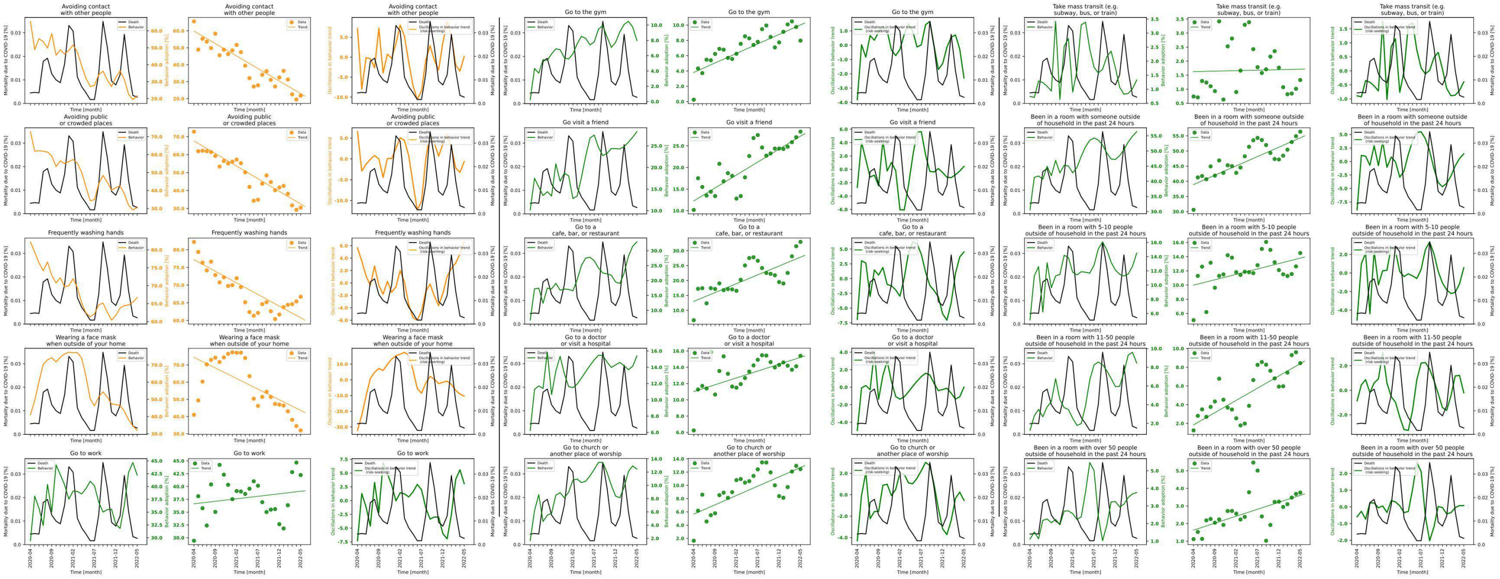

**Figure S7.41 shows behavior and mortality on 1st, 4th and 7th column, behavior trends in 2nd, 5th and 8th column, and behavior oscillatory components juxtaposed with mortality rates on 3rd, 6th and 9th columns for South Carolina.**

| Behaviors | Slopes | Y-intercepts | Start %$(S)$ | End %$(E)$ | $\|E-S\|$ | $\frac{\|E-S\|*100}{S}$ |
|---|---|---|---|---|---|---|
| Avoiding contact with other people | -1.475 | 59.861 | 65.562 | 21.712 | 43.85 | 66.883 |
| Avoiding public or crowded places | -1.405 | 67.628 | 72.772 | 30.491 | 42.281 | 58.101 |
| Frequently washing hands | -0.657 | 77.195 | 82.21 | 66.703 | 15.507 | 18.863 |
| Wearing a face mask when outside of your home | -1.233 | 74.436 | 41.02 | 31.966 | 9.054 | 22.072 |
| Go to work | 0.098 | 36.624 | 29.424 | 42.233 | 12.809 | 43.532 |
| Go to the gym | 0.248 | 3.811 | 0.246 | 7.972 | 7.726 | 3140.65 |
| Go visit a friend | 0.6 | 12.26 | 10.168 | 28.302 | 18.134 | 178.344 |
| Go to a cafe, bar, or restaurant | 0.593 | 13.009 | 6.687 | 32.968 | 26.281 | 393.016 |
| Go to a doctor or visit a hospital | 0.168 | 11.01 | 6.236 | 15.4 | 9.164 | 146.953 |
| Go to church or another place of worship | 0.281 | 5.663 | 1.652 | 12.444 | 10.792 | 653.269 |
| Take mass transit (e.g. subway, bus, or train) | 0.004 | 1.626 | 0.74 | 1.327 | 0.587 | 79.324 |
| Been in a room with someone outside of household in the past 24 hours | 0.617 | 38.914 | 30.581 | 56.372 | 25.791 | 84.337 |
| Been in a room with 5-10 people outside of household in the past 24 hours | 0.152 | 9.99 | 5.064 | 14.511 | 9.447 | 186.552 |
| Been in a room with 11-50 people outside of household in the past 24 hours | 0.262 | 1.847 | 1.249 | 8.441 | 7.192 | 575.821 |
| Been in a room with over 50 people outside of household in the past 24 hours | 0.079 | 1.628 | 1.115 | 3.761 | 2.646 | 237.309 |

**Table S7.41 shows the slopes, y-intercept, adherence % at the beginning of the study period, and at the end, the absolute difference between the beginning and the end, and the absolute difference as a percentage of the adherence % at the beginning of the study period of each behavior's trend in South Carolina.**

### State name: South Dakota

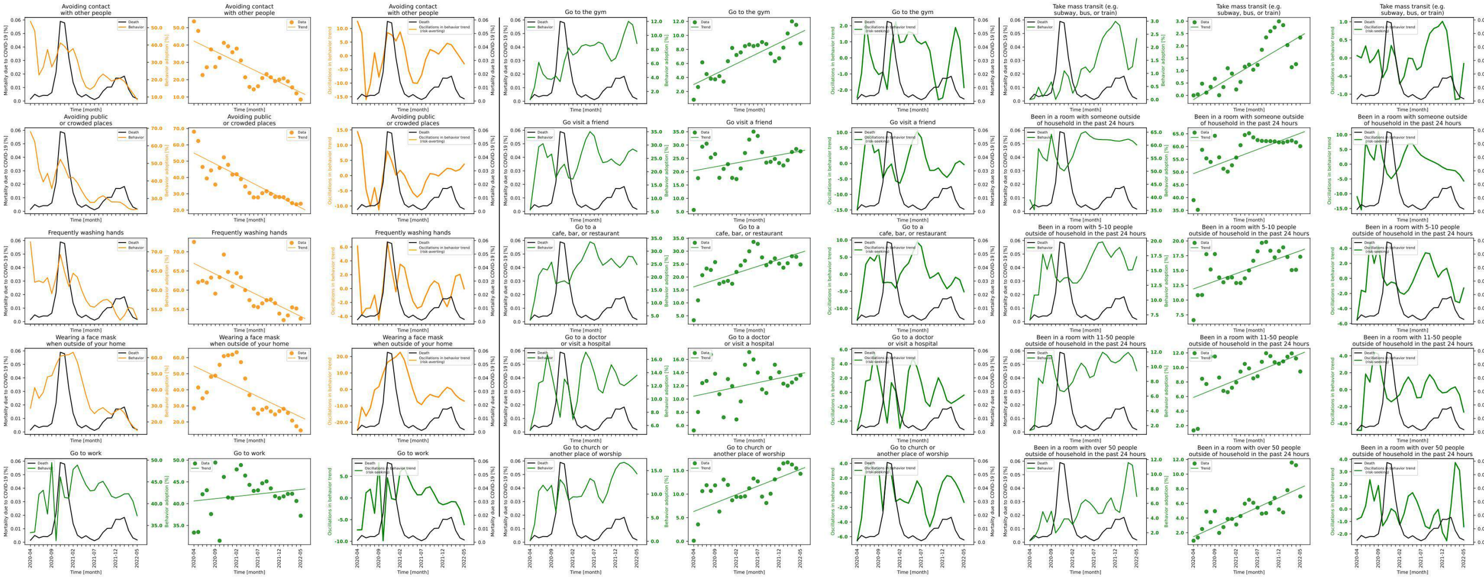

**Figure S7.42 shows behavior and mortality on 1st, 4th and 7th column, behavior trends in 2nd, 5th and 8th column, and behavior oscillatory components juxtaposed with mortality rates on 3rd, 6th and 9th columns for South Dakota.**

| Behaviors | Slopes | Y-intercepts | Start %($S$) | End %($E$) | $\|E-S\|$ | $\frac{\|E-S\|*100}{S}$ |
|---|---|---|---|---|---|---|
| Avoiding contact with other people | -1.183 | 42.237 | 53.649 | 8.554 | 45.095 | 84.056 |
| Avoiding public or crowded places | -1.338 | 54.976 | 68.074 | 23.918 | 44.156 | 64.865 |
| Frequently washing hands | -0.551 | 66.948 | 72.538 | 52.583 | 19.955 | 27.51 |
| Wearing a face mask when outside of your home | -1.267 | 54.728 | 28.522 | 14.7 | 13.822 | 48.461 |
| Go to work | 0.106 | 40.617 | 33.36 | 37.226 | 3.866 | 11.589 |
| Go to the gym | 0.296 | 3.0 | 0.831 | 8.859 | 8.028 | 966.065 |
| Go visit a friend | 0.295 | 20.362 | 5.65 | 27.647 | 21.997 | 389.327 |
| Go to a cafe, bar, or restaurant | 0.529 | 16.209 | 3.286 | 24.836 | 21.55 | 655.813 |
| Go to a doctor or visit a hospital | 0.136 | 10.434 | 5.243 | 13.594 | 8.351 | 159.279 |
| Go to church or another place of worship | 0.358 | 6.314 | 0.119 | 14.316 | 14.197 | 11930.252 |
| Take mass transit (e.g. subway, bus, or train) | 0.102 | -0.167 | 0.0 | 2.337 | 2.337 | inf |
| Been in a room with someone outside of household in the past 24 hours | 0.632 | 49.393 | 38.992 | 60.12 | 21.128 | 54.185 |
| Been in a room with 5-10 people outside of household in the past 24 hours | 0.258 | 11.894 | 6.609 | 17.33 | 10.721 | 162.218 |
| Been in a room with 11-50 people outside of household in the past 24 hours | 0.239 | 5.881 | 1.336 | 9.442 | 8.106 | 606.737 |
| Been in a room with over 50 people outside of household in the past 24 hours | 0.266 | 1.457 | 0.901 | 6.954 | 6.053 | 671.809 |

**Table S7.42 shows the slopes, y-intercept, adherence % at the beginning of the study period, and at the end, the absolute difference between the beginning and the end, and the absolute difference as a percentage of the adherence % at the beginning of the study period of each behavior's trend in South Dakota.**

### State name: Tennessee

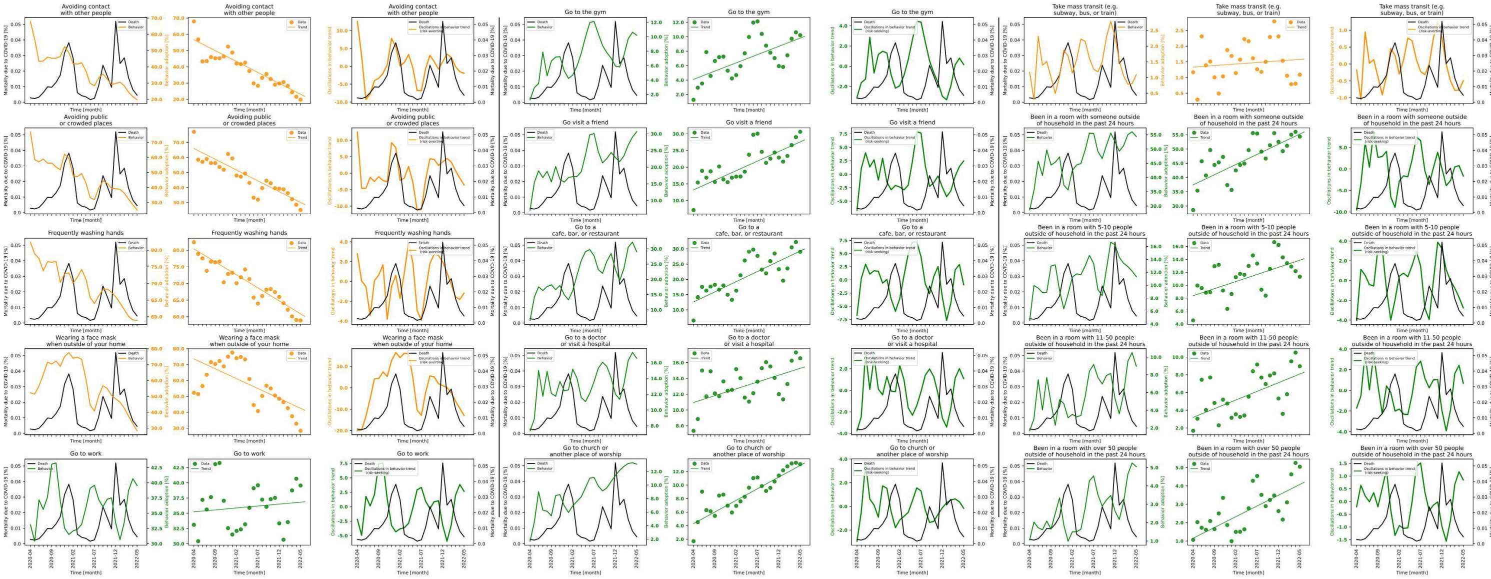

**Figure S7.43 shows behavior and mortality on 1st, 4th and 7th column, behavior trends in 2nd, 5th and 8th column, and behavior oscillatory components juxtaposed with mortality rates on 3rd, 6th and 9th columns for Tennessee.**

| Behaviors | Slopes | Y-intercepts | Start %($S$) | End %($E$) | $\lvert E-S \rvert$ | $\frac{\lvert E-S \rvert * 100}{S}$ |
|---|---|---|---|---|---|---|
| Avoiding contact with other people | -1.344 | 56.695 | 67.955 | 19.835 | 48.12 | 70.812 |
| Avoiding public or crowded places | -1.438 | 65.977 | 77.093 | 25.034 | 52.059 | 67.528 |
| Frequently washing hands | -0.781 | 80.388 | 82.402 | 58.911 | 23.491 | 28.508 |
| Wearing a face mask when outside of your home | -1.245 | 73.608 | 52.279 | 28.336 | 23.943 | 45.799 |
| Go to work | 0.064 | 35.233 | 33.103 | 39.559 | 6.456 | 19.503 |
| Go to the gym | 0.229 | 4.107 | 1.268 | 10.218 | 8.95 | 705.836 |
| Go visit a friend | 0.576 | 13.197 | 7.108 | 30.63 | 23.522 | 330.923 |
| Go to a cafe, bar, or restaurant | 0.669 | 12.564 | 6.518 | 29.032 | 22.514 | 345.413 |
| Go to a doctor or visit a hospital | 0.173 | 10.954 | 7.363 | 16.549 | 9.186 | 124.759 |
| Go to church or another place of worship | 0.344 | 4.302 | 1.708 | 13.084 | 11.376 | 666.042 |
| Take mass transit (e.g. subway, bus, or train) | 0.01 | 1.334 | 1.17 | 1.092 | 0.078 | 6.667 |
| Been in a room with someone outside of household in the past 24 hours | 0.714 | 37.416 | 28.587 | 54.359 | 25.772 | 90.153 |
| Been in a room with 5-10 people outside of household in the past 24 hours | 0.22 | 8.373 | 4.56 | 11.317 | 6.757 | 148.18 |
| Been in a room with 11-50 people outside of household in the past 24 hours | 0.196 | 3.198 | 1.681 | 8.961 | 7.28 | 433.076 |
| Been in a room with over 50 people outside of household in the past 24 hours | 0.117 | 1.153 | 1.062 | 5.044 | 3.982 | 374.953 |

**Table S7.43 shows the slopes, y-intercept, adherence % at the beginning of the study period, and at the end, the absolute difference between the beginning and the end, and the absolute difference as a percentage of the adherence % at the beginning of the study period of each behavior's trend in Tennessee.**

### State name: Texas

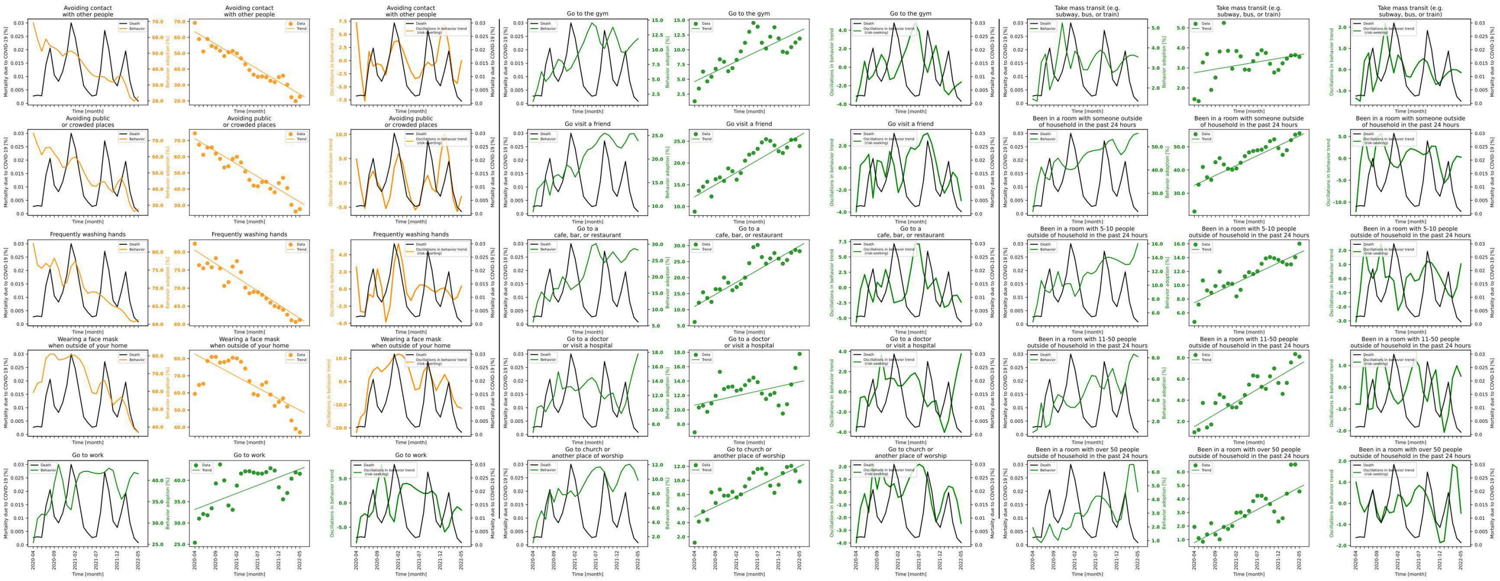

**Figure S7.44 shows behavior and mortality on 1st, 4th and 7th column, behavior trends in 2nd, 5th and 8th column, and behavior oscillatory components juxtaposed with mortality rates on 3rd, 6th and 9th columns for Texas.**

| Behaviors | Slopes | Y-intercepts | Start %$(S)$ | End %$(E)$ | $\|E-S\|$ | $\frac{\|E-S\|*100}{S}$ |
|---|---|---|---|---|---|---|
| Avoiding contact with other people | -1.575 | 63.534 | 69.141 | 22.543 | 46.598 | 67.396 |
| Avoiding public or crowded places | -1.564 | 71.138 | 74.49 | 27.652 | 46.838 | 62.878 |
| Frequently washing hands | -0.755 | 80.493 | 82.305 | 61.19 | 21.115 | 25.655 |
| Wearing a face mask when outside of your home | -1.313 | 82.653 | 59.43 | 37.007 | 22.423 | 37.73 |
| Go to work | 0.378 | 33.187 | 25.39 | 41.557 | 16.167 | 63.675 |
| Go to the gym | 0.341 | 4.626 | 1.278 | 11.887 | 10.609 | 830.125 |
| Go visit a friend | 0.563 | 12.217 | 8.816 | 23.892 | 15.076 | 171.007 |
| Go to a cafe, bar, or restaurant | 0.754 | 11.029 | 6.161 | 28.255 | 22.094 | 358.611 |
| Go to a doctor or visit a hospital | 0.13 | 10.646 | 6.866 | 17.773 | 10.907 | 158.855 |
| Go to church or another place of worship | 0.286 | 4.857 | 1.167 | 9.841 | 8.674 | 743.273 |
| Take mass transit (e.g. subway, bus, or train) | 0.036 | 2.759 | 1.438 | 3.546 | 2.108 | 146.592 |
| Been in a room with someone outside of household in the past 24 hours | 0.851 | 33.428 | 22.14 | 55.801 | 33.661 | 152.037 |
| Been in a room with 5-10 people outside of household in the past 24 hours | 0.288 | 7.474 | 4.689 | 16.014 | 11.325 | 241.523 |
| Been in a room with 11-50 people outside of household in the past 24 hours | 0.234 | 1.517 | 0.976 | 8.105 | 7.129 | 730.43 |
| Been in a room with over 50 people outside of household in the past 24 hours | 0.163 | 0.776 | 1.942 | 4.558 | 2.616 | 134.706 |

**Table S7.44 shows the slopes, y-intercept, adherence % at the beginning of the study period, and at the end, the absolute difference between the beginning and the end, and the absolute difference as a percentage of the adherence % at the beginning of the study period of each behavior's trend in Texas.**

### State name: Utah

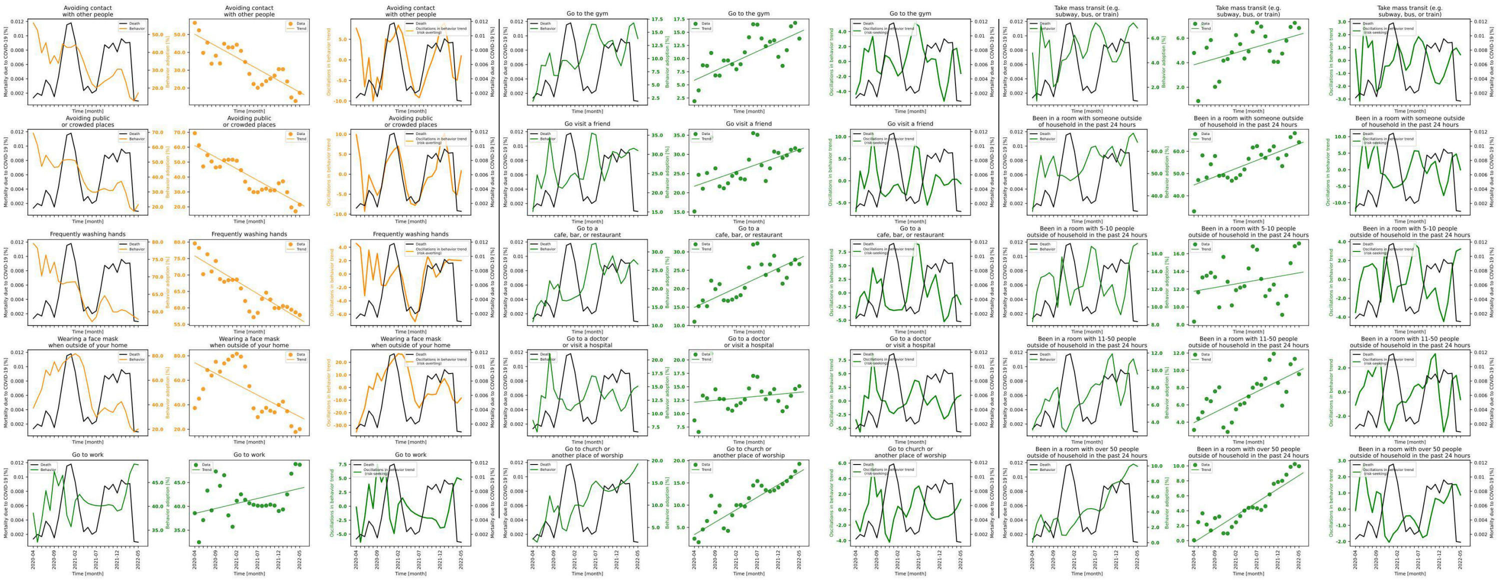

**Figure S7.45 shows behavior and mortality on 1st, 4th and 7th column, behavior trends in 2nd, 5th and 8th column, and behavior oscillatory components juxtaposed with mortality rates on 3rd, 6th and 9th columns for Utah.**

| Behaviors | Slopes | Y-intercepts | Start %($S$) | End %($E$) | $\|E-S\|$ | $\frac{\|E-S\|*100}{S}$ |
|---|---|---|---|---|---|---|
| Avoiding contact with other people | -1.3 | 50.154 | 56.539 | 17.3 | 39.239 | 69.402 |
| Avoiding public or crowded places | -1.555 | 61.046 | 69.407 | 21.464 | 47.943 | 69.075 |
| Frequently washing hands | -0.768 | 75.781 | 79.575 | 57.844 | 21.731 | 27.309 |
| Wearing a face mask when outside of your home | -1.762 | 74.276 | 37.297 | 20.109 | 17.188 | 46.084 |
| Go to work | 0.21 | 38.504 | 38.568 | 48.727 | 10.159 | 26.34 |
| Go to the gym | 0.366 | 5.873 | 1.886 | 13.789 | 11.903 | 631.124 |
| Go visit a friend | 0.381 | 21.715 | 15.139 | 30.991 | 15.852 | 104.71 |
| Go to a cafe, bar, or restaurant | 0.532 | 14.934 | 11.039 | 26.68 | 15.641 | 141.689 |
| Go to a doctor or visit a hospital | 0.074 | 12.111 | 8.738 | 15.116 | 6.378 | 72.992 |
| Go to church or another place of worship | 0.56 | 3.408 | 2.478 | 19.271 | 16.793 | 677.684 |
| Take mass transit (e.g. subway, bus, or train) | 0.098 | 3.832 | 4.793 | 6.826 | 2.033 | 42.416 |
| Been in a room with someone outside of household in the past 24 hours | 0.739 | 44.894 | 33.049 | 64.071 | 31.022 | 93.867 |
| Been in a room with 5-10 people outside of household in the past 24 hours | 0.083 | 11.769 | 8.342 | 17.15 | 8.808 | 105.586 |
| Been in a room with 11-50 people outside of household in the past 24 hours | 0.24 | 3.939 | 3.099 | 9.556 | 6.457 | 208.358 |
| Been in a room with over 50 people outside of household in the past 24 hours | 0.357 | -0.209 | 0.061 | 9.953 | 9.892 | 16216.393 |

**Table S7.45 shows the slopes, y-intercept, adherence % at the beginning of the study period, and at the end, the absolute difference between the beginning and the end, and the absolute difference as a percentage of the adherence % at the beginning of the study period of each behavior's trend in Utah.**

### State name: Vermont

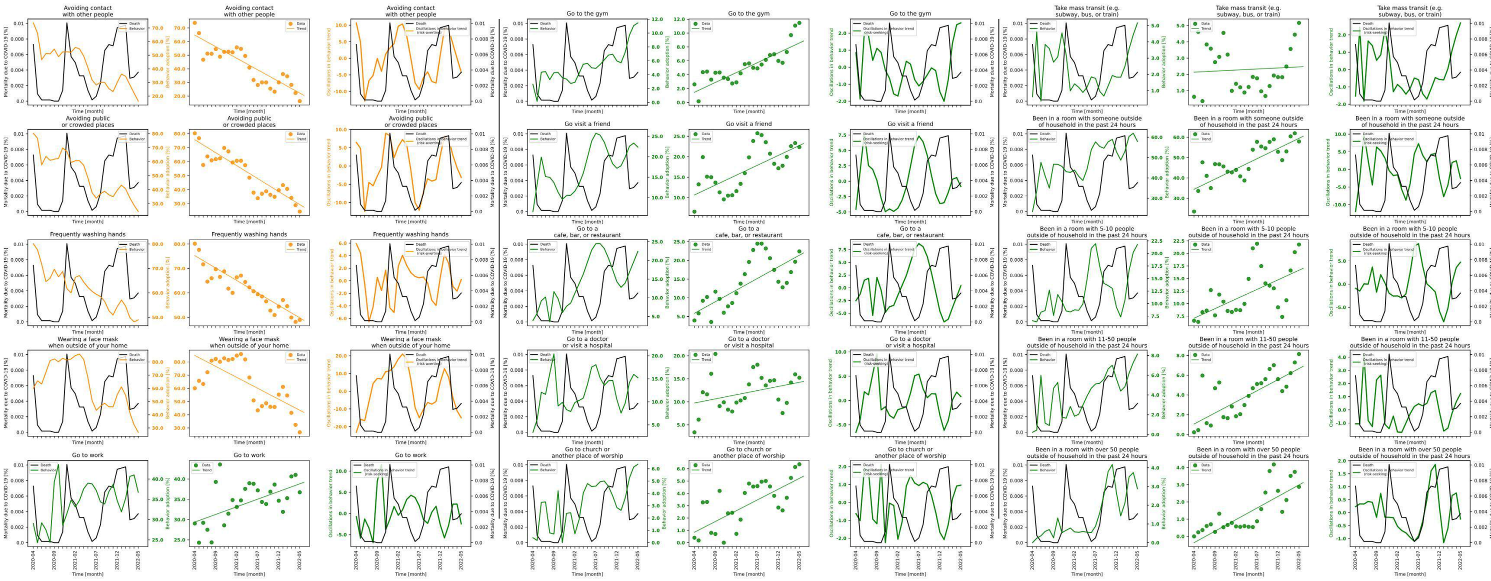

**Figure S7.46 shows behavior and mortality on 1st, 4th and 7th column, behavior trends in 2nd, 5th and 8th column, and behavior oscillatory components juxtaposed with mortality rates on 3rd, 6th and 9th columns for Vermont.**

| Behaviors | Slopes | Y-intercepts | Start %($S$) | End %($E$) | $\|E-S\|$ | $\frac{\|E-S\|*100}{S}$ |
|---|---|---|---|---|---|---|
| Avoiding contact with other people | -1.701 | 64.751 | 73.729 | 16.31 | 57.419 | 77.878 |
| Avoiding public or crowded places | -1.85 | 75.642 | 80.264 | 24.433 | 55.831 | 69.559 |
| Frequently washing hands | -1.017 | 75.23 | 80.117 | 48.974 | 31.143 | 38.872 |
| Wearing a face mask when outside of your home | -1.66 | 84.918 | 60.078 | 26.737 | 33.341 | 55.496 |
| Go to work | 0.377 | 29.391 | 29.037 | 36.735 | 7.698 | 26.511 |
| Go to the gym | 0.284 | 1.467 | 2.607 | 11.463 | 8.856 | 339.701 |
| Go visit a friend | 0.483 | 10.662 | 6.578 | 22.347 | 15.769 | 239.723 |
| Go to a cafe, bar, or restaurant | 0.621 | 5.858 | 3.934 | 22.417 | 18.483 | 469.827 |
| Go to a doctor or visit a hospital | 0.18 | 9.745 | 3.381 | 15.245 | 11.864 | 350.902 |
| Go to church or another place of worship | 0.175 | 0.847 | 0.4 | 6.362 | 5.962 | 1490.5 |
| Take mass transit (e.g. subway, bus, or train) | 0.013 | 2.137 | 0.621 | 5.165 | 4.544 | 731.723 |
| Been in a room with someone outside of household in the past 24 hours | 1.004 | 34.415 | 23.481 | 57.943 | 34.462 | 146.765 |
| Been in a room with 5-10 people outside of household in the past 24 hours | 0.382 | 7.091 | 6.59 | 21.775 | 15.185 | 230.425 |
| Been in a room with 11-50 people outside of household in the past 24 hours | 0.225 | 1.056 | 0.221 | 8.141 | 7.92 | 3583.71 |
| Been in a room with over 50 people outside of household in the past 24 hours | 0.134 | -0.367 | 0.0 | 2.877 | 2.877 | inf |

**Table S7.46 shows the slopes, y-intercept, adherence % at the beginning of the study period, and at the end, the absolute difference between the beginning and the end, and the absolute difference as a percentage of the adherence % at the beginning of the study period of each behavior's trend in Vermont.**

### State name: Virginia

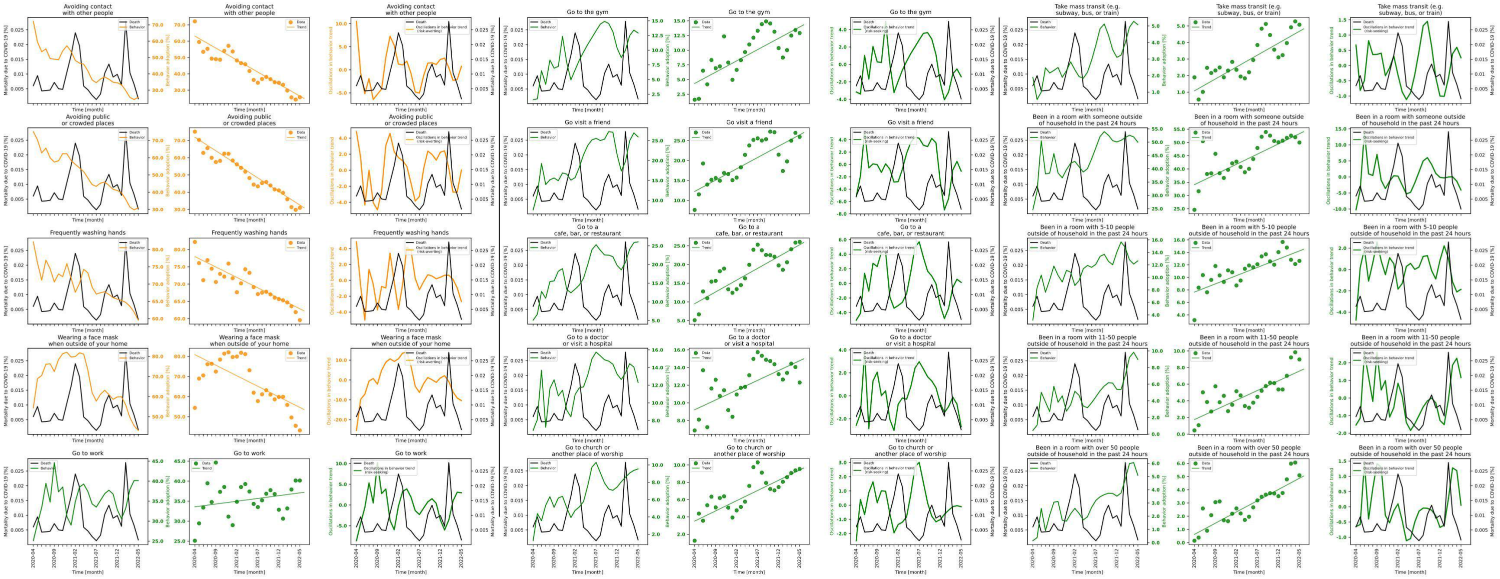

**Figure S7.47 shows behavior and mortality on 1st, 4th and 7th column, behavior trends in 2nd, 5th and 8th column, and behavior oscillatory components juxtaposed with mortality rates on 3rd, 6th and 9th columns for Virginia.**

| Behaviors | Slopes | Y-intercepts | Start %($S$) | End %($E$) | $\|E-S\|$ | $\frac{\|E-S\|*100}{S}$ |
|---|---|---|---|---|---|---|
| Avoiding contact with other people | -1.467 | 63.088 | 72.118 | 25.699 | 46.419 | 64.365 |
| Avoiding public or crowded places | -1.575 | 71.878 | 75.106 | 30.942 | 44.164 | 58.802 |
| Frequently washing hands | -0.604 | 77.96 | 82.187 | 59.576 | 22.611 | 27.512 |
| Wearing a face mask when outside of your home | -1.05 | 80.849 | 54.394 | 43.188 | 11.206 | 20.602 |
| Go to work | 0.14 | 33.577 | 25.099 | 40.154 | 15.055 | 59.982 |
| Go to the gym | 0.384 | 4.339 | 1.571 | 12.918 | 11.347 | 722.279 |
| Go visit a friend | 0.568 | 12.242 | 7.593 | 25.93 | 18.337 | 241.499 |
| Go to a cafe, bar, or restaurant | 0.631 | 9.515 | 5.101 | 26.055 | 20.954 | 410.782 |
| Go to a doctor or visit a hospital | 0.222 | 9.218 | 6.874 | 12.305 | 5.431 | 79.008 |
| Go to church or another place of worship | 0.236 | 3.518 | 1.254 | 9.54 | 8.286 | 660.766 |
| Take mass transit (e.g. subway, bus, or train) | 0.144 | 1.077 | 1.894 | 5.079 | 3.185 | 168.163 |
| Been in a room with someone outside of household in the past 24 hours | 0.769 | 34.074 | 24.584 | 49.984 | 25.4 | 103.319 |
| Been in a room with 5-10 people outside of household in the past 24 hours | 0.264 | 7.65 | 3.165 | 12.639 | 9.474 | 299.336 |
| Been in a room with 11-50 people outside of household in the past 24 hours | 0.233 | 1.745 | 0.444 | 8.973 | 8.529 | 1920.946 |
| Been in a room with over 50 people outside of household in the past 24 hours | 0.17 | 0.625 | 0.144 | 5.09 | 4.946 | 3434.722 |

**Table S7.47 shows the slopes, y-intercept, adherence % at the beginning of the study period, and at the end, the absolute difference between the beginning and the end, and the absolute difference as a percentage of the adherence % at the beginning of the study period of each behavior's trend in Virginia.**

### State name: Washington

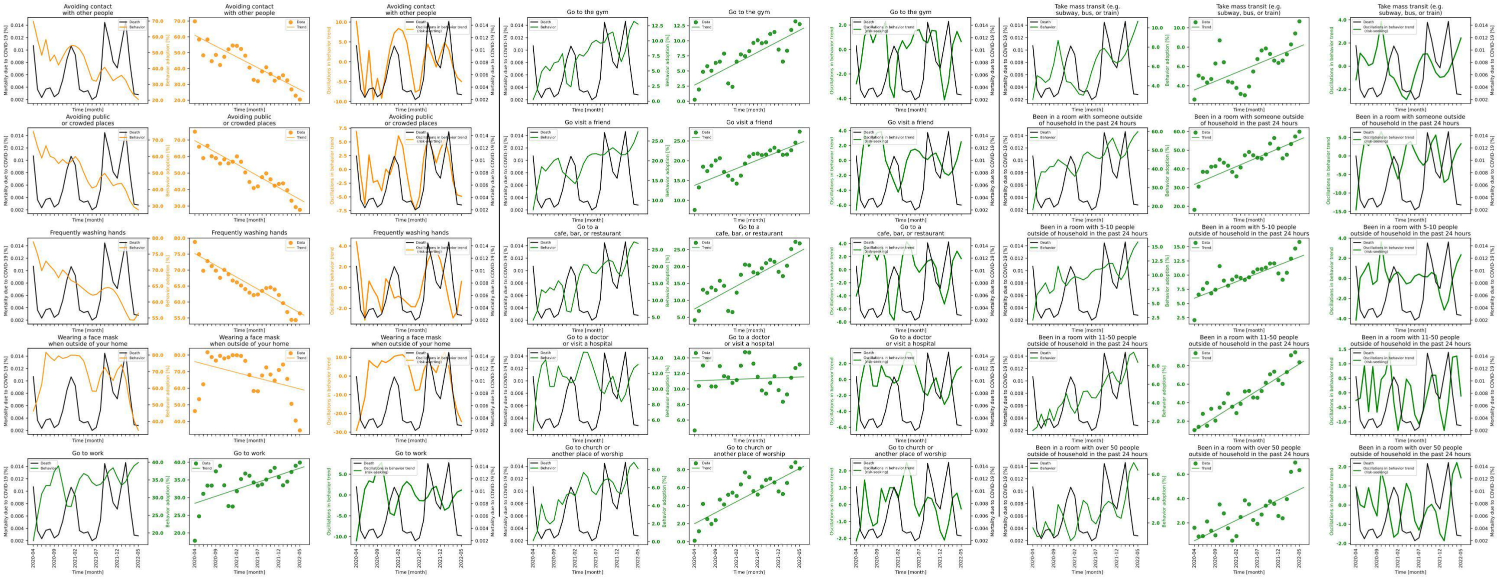

**Figure S7.48 shows behavior and mortality on 1st, 4th and 7th column, behavior trends in 2nd, 5th and 8th column, and behavior oscillatory components juxtaposed with mortality rates on 3rd, 6th and 9th columns for Washington.**

| Behaviors | Slopes | Y-intercepts | Start %$(S)$ | End %$(E)$ | $\|E-S\|$ | $\frac{\|E-S\|*100}{S}$ |
|---|---|---|---|---|---|---|
| Avoiding contact with other people | -1.367 | 60.977 | 69.782 | 20.457 | 49.325 | 70.684 |
| Avoiding public or crowded places | -1.424 | 69.615 | 75.064 | 27.713 | 47.351 | 63.081 |
| Frequently washing hands | -0.742 | 75.144 | 78.791 | 56.438 | 22.353 | 28.37 |
| Wearing a face mask when outside of your home | -0.657 | 75.992 | 46.242 | 34.605 | 11.637 | 25.165 |
| Go to work | 0.397 | 28.394 | 17.802 | 39.906 | 22.104 | 124.166 |
| Go to the gym | 0.363 | 2.675 | 0.244 | 12.763 | 12.519 | 5130.738 |
| Go visit a friend | 0.426 | 13.787 | 7.57 | 27.351 | 19.781 | 261.308 |
| Go to a cafe, bar, or restaurant | 0.679 | 7.472 | 4.127 | 26.852 | 22.725 | 550.642 |
| Go to a doctor or visit a hospital | 0.02 | 11.049 | 4.663 | 13.149 | 8.486 | 181.986 |
| Go to church or another place of worship | 0.245 | 1.995 | 0.103 | 8.12 | 8.017 | 7783.495 |
| Take mass transit (e.g. subway, bus, or train) | 0.179 | 3.57 | 2.574 | 10.662 | 8.088 | 314.219 |
| Been in a room with someone outside of household in the past 24 hours | 0.959 | 31.701 | 18.12 | 59.916 | 41.796 | 230.662 |
| Been in a room with 5-10 people outside of household in the past 24 hours | 0.291 | 5.928 | 2.067 | 15.843 | 13.776 | 666.473 |
| Been in a room with 11-50 people outside of household in the past 24 hours | 0.288 | 0.99 | 1.019 | 8.375 | 7.356 | 721.884 |
| Been in a room with over 50 people outside of household in the past 24 hours | 0.169 | 0.531 | 1.596 | 6.336 | 4.74 | 296.992 |

**Table S7.48 shows the slopes, y-intercept, adherence % at the beginning of the study period, and at the end, the absolute difference between the beginning and the end, and the absolute difference as a percentage of the adherence % at the beginning of the study period of each behavior's trend in Washington.**

### State name: West Virginia

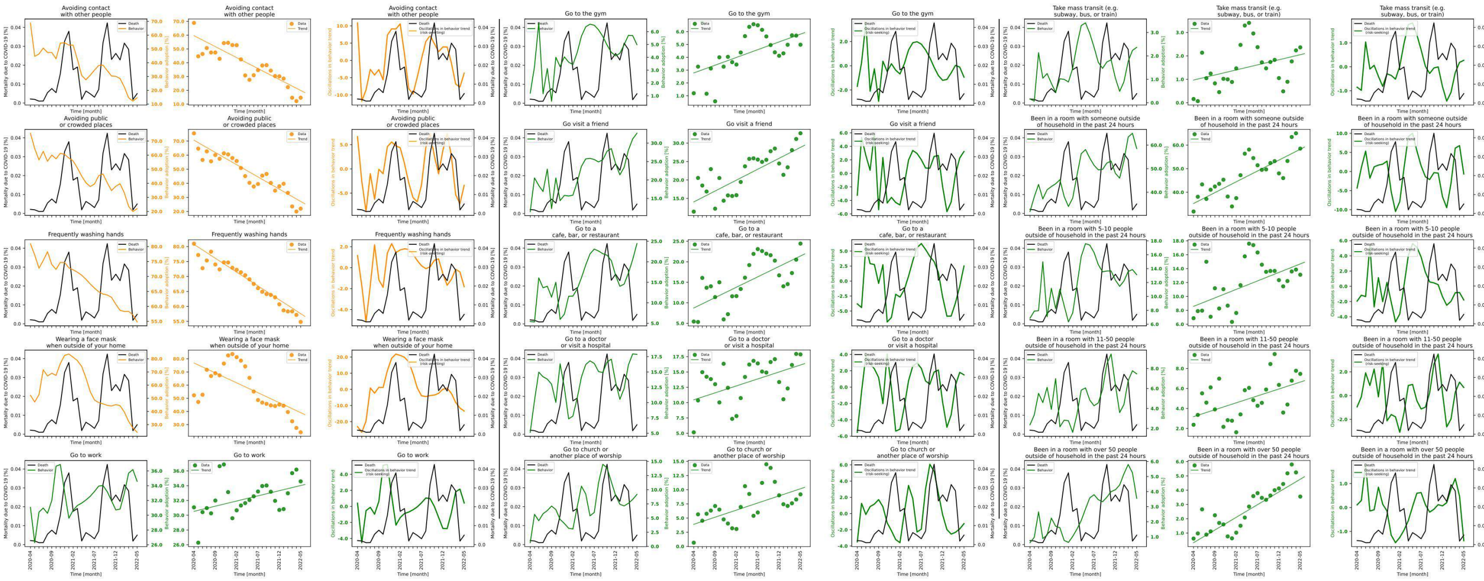

**Figure S7.49 shows behavior and mortality on 1st, 4th and 7th column, behavior trends in 2nd, 5th and 8th column, and behavior oscillatory components juxtaposed with mortality rates on 3rd, 6th and 9th columns for West Virginia.**

| Behaviors | Slopes | Y-intercepts | Start %($S$) | End %($E$) | $\|E-S\|$ | $\frac{\|E-S\|*100}{S}$ |
|---|---|---|---|---|---|---|
| Avoiding contact with other people | -1.591 | 59.617 | 68.888 | 14.593 | 54.295 | 78.816 |
| Avoiding public or crowded places | -1.739 | 70.606 | 75.528 | 22.023 | 53.505 | 70.841 |
| Frequently washing hands | -0.926 | 80.724 | 80.958 | 54.823 | 26.135 | 32.282 |
| Wearing a face mask when outside of your home | -1.508 | 76.815 | 52.266 | 24.187 | 28.079 | 53.723 |
| Go to work | 0.14 | 30.527 | 31.068 | 34.587 | 3.519 | 11.327 |
| Go to the gym | 0.121 | 2.782 | 1.202 | 4.984 | 3.782 | 314.642 |
| Go visit a friend | 0.591 | 14.052 | 11.416 | 32.671 | 21.255 | 186.186 |
| Go to a cafe, bar, or restaurant | 0.507 | 8.73 | 5.43 | 24.429 | 18.999 | 349.89 |
| Go to a doctor or visit a hospital | 0.227 | 10.458 | 5.15 | 17.872 | 12.722 | 247.029 |
| Go to church or another place of worship | 0.251 | 3.899 | 0.658 | 9.182 | 8.524 | 1295.441 |
| Take mass transit (e.g. subway, bus, or train) | 0.043 | 0.969 | 0.16 | 2.364 | 2.204 | 1377.5 |
| Been in a room with someone outside of household in the past 24 hours | 0.921 | 35.221 | 31.814 | 58.52 | 26.706 | 83.944 |
| Been in a room with 5-10 people outside of household in the past 24 hours | 0.244 | 8.571 | 6.865 | 13.122 | 6.257 | 91.143 |
| Been in a room with 11-50 people outside of household in the past 24 hours | 0.138 | 3.185 | 2.373 | 7.461 | 5.088 | 214.412 |
| Been in a room with over 50 people outside of household in the past 24 hours | 0.178 | 0.323 | 0.611 | 3.553 | 2.942 | 481.506 |

**Table S7.49 shows the slopes, y-intercept, adherence % at the beginning of the study period, and at the end, the absolute difference between the beginning and the end, and the absolute difference as a percentage of the adherence % at the beginning of the study period of each behavior's trend in West Virginia.**

### State name: Wisconsin

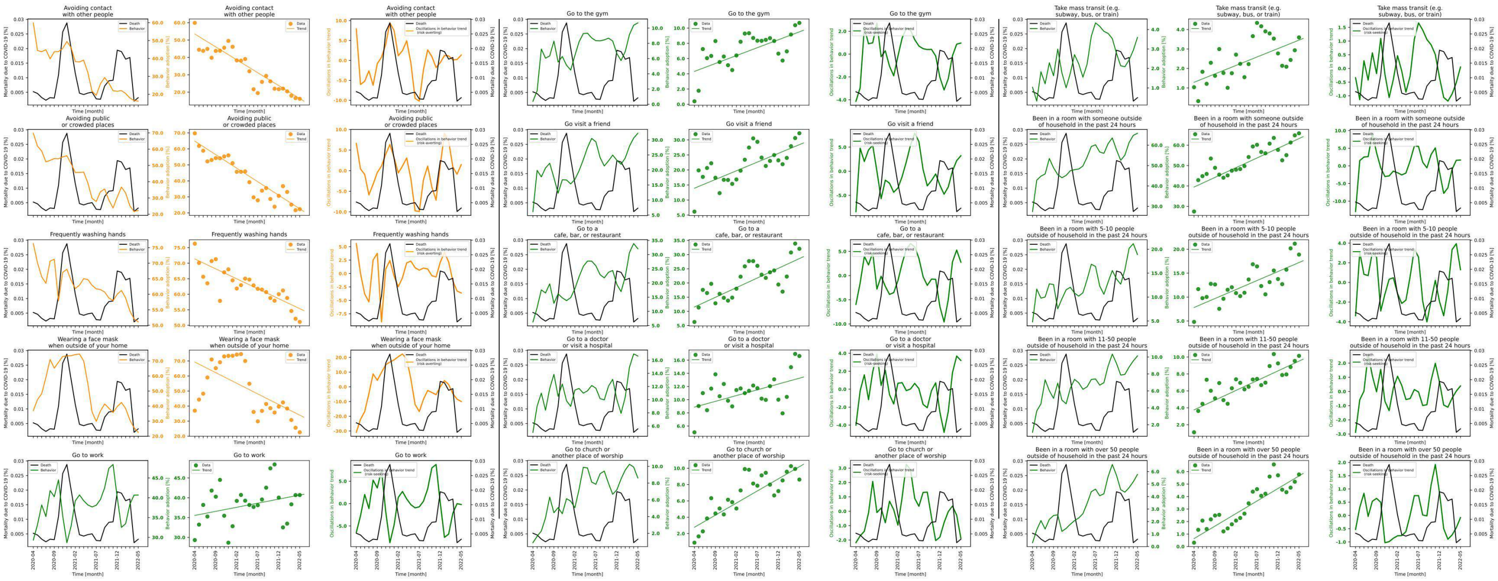

**Figure S7.50 shows behavior and mortality on 1st, 4th and 7th column, behavior trends in 2nd, 5th and 8th column, and behavior oscillatory components juxtaposed with mortality rates on 3rd, 6th and 9th columns for Wisconsin.**

| Behaviors | Slopes | Y-intercepts | Start %($S$) | End %($E$) | $\|E-S\|$ | $\frac{\|E-S\|*100}{S}$ |
|---|---|---|---|---|---|---|
| Avoiding contact with other people | -1.485 | 53.462 | 59.888 | 16.186 | 43.702 | 72.973 |
| Avoiding public or crowded places | -1.691 | 64.815 | 69.743 | 22.322 | 47.421 | 67.994 |
| Frequently washing hands | -0.64 | 71.368 | 76.287 | 51.099 | 25.188 | 33.017 |
| Wearing a face mask when outside of your home | -1.411 | 69.244 | 36.951 | 22.573 | 14.378 | 38.911 |
| Go to work | 0.206 | 35.545 | 29.323 | 40.686 | 11.363 | 38.751 |
| Go to the gym | 0.208 | 4.317 | 0.391 | 10.682 | 10.291 | 2631.969 |
| Go visit a friend | 0.579 | 13.935 | 6.135 | 32.213 | 26.078 | 425.069 |
| Go to a cafe, bar, or restaurant | 0.682 | 11.541 | 6.259 | 32.057 | 25.798 | 412.174 |
| Go to a doctor or visit a hospital | 0.175 | 8.874 | 5.045 | 16.633 | 11.588 | 229.693 |
| Go to church or another place of worship | 0.297 | 2.771 | 0.883 | 8.613 | 7.73 | 875.425 |
| Take mass transit (e.g. subway, bus, or train) | 0.087 | 1.288 | 1.035 | 3.594 | 2.559 | 247.246 |
| Been in a room with someone outside of household in the past 24 hours | 0.951 | 39.465 | 27.476 | 65.83 | 38.354 | 139.591 |
| Been in a room with 5-10 people outside of household in the past 24 hours | 0.376 | 7.821 | 4.81 | 18.889 | 14.079 | 292.703 |
| Been in a room with 11-50 people outside of household in the past 24 hours | 0.23 | 3.754 | 1.108 | 10.136 | 9.028 | 814.801 |
| Been in a room with over 50 people outside of household in the past 24 hours | 0.201 | 0.635 | 0.307 | 5.771 | 5.464 | 1779.805 |

**Table S7.50 shows the slopes, y-intercept, adherence % at the beginning of the study period, and at the end, the absolute difference between the beginning and the end, and the absolute difference as a percentage of the adherence % at the beginning of the study period of each behavior's trend in Wisconsin.**

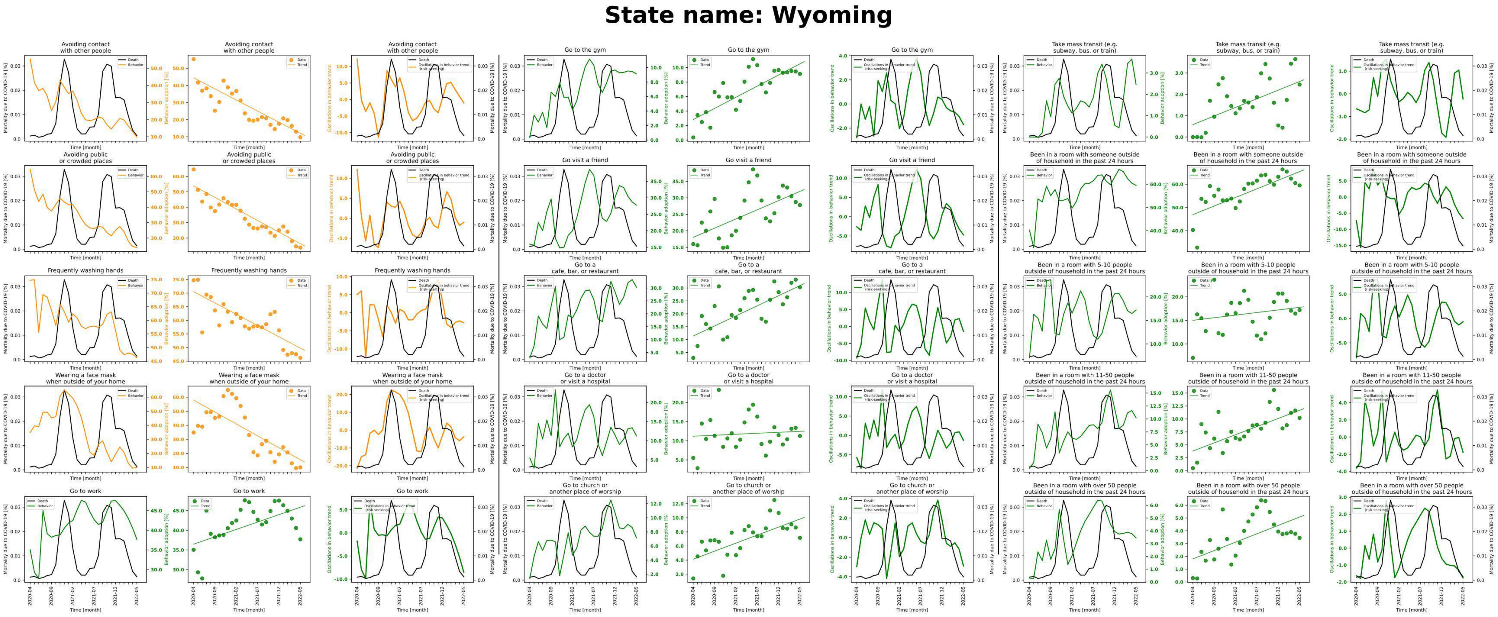

**Figure S7.51 shows behavior and mortality on 1st, 4th and 7th column, behavior trends in 2nd, 5th and 8th column, and behavior oscillatory components juxtaposed with mortality rates on 3rd, 6th and 9th columns for Wyoming.**

| Behaviors | Slopes | Y-intercepts | Start %($S$) | End %($E$) | $\|E-S\|$ | $\frac{\|E-S\|*100}{S}$ |
|---|---|---|---|---|---|---|
| Avoiding contact with other people | -1.295 | 44.422 | 55.331 | 9.726 | 45.605 | 82.422 |
| Avoiding public or crowded places | -1.5 | 53.827 | 64.539 | 13.79 | 50.749 | 78.633 |
| Frequently washing hands | -0.829 | 70.524 | 74.717 | 46.204 | 28.513 | 38.161 |
| Wearing a face mask when outside of your home | -1.698 | 57.898 | 34.844 | 9.997 | 24.847 | 71.309 |
| Go to work | 0.376 | 36.472 | 35.034 | 37.703 | 2.669 | 7.618 |
| Go to the gym | 0.307 | 2.829 | 0.357 | 9.117 | 8.76 | 2453.782 |
| Go visit a friend | 0.557 | 18.029 | 15.969 | 27.797 | 11.828 | 74.069 |
| Go to a cafe, bar, or restaurant | 0.782 | 11.382 | 2.873 | 30.28 | 27.407 | 953.951 |
| Go to a doctor or visit a hospital | 0.051 | 11.202 | 5.514 | 11.269 | 5.755 | 104.371 |
| Go to church or another place of worship | 0.228 | 4.127 | 1.388 | 7.156 | 5.768 | 415.562 |
| Take mass transit (e.g. subway, bus, or train) | 0.081 | 0.581 | 0.0 | 2.456 | 2.456 | inf |
| Been in a room with someone outside of household in the past 24 hours | 0.74 | 46.944 | 40.314 | 59.492 | 19.178 | 47.572 |
| Been in a room with 5-10 people outside of household in the past 24 hours | 0.11 | 15.017 | 7.049 | 17.211 | 10.162 | 144.162 |
| Been in a room with 11-50 people outside of household in the past 24 hours | 0.314 | 3.813 | 0.497 | 10.205 | 9.708 | 1953.32 |
| Been in a room with over 50 people outside of household in the past 24 hours | 0.131 | 1.802 | 0.303 | 3.444 | 3.141 | 1036.634 |

**Table S7.51 shows the slopes, y-intercept, adherence % at the beginning of the study period, and at the end, the absolute difference between the beginning and the end, and the absolute difference as a percentage of the adherence % at the beginning of the study period of each behavior's trend in Wyoming.**

### State name: National

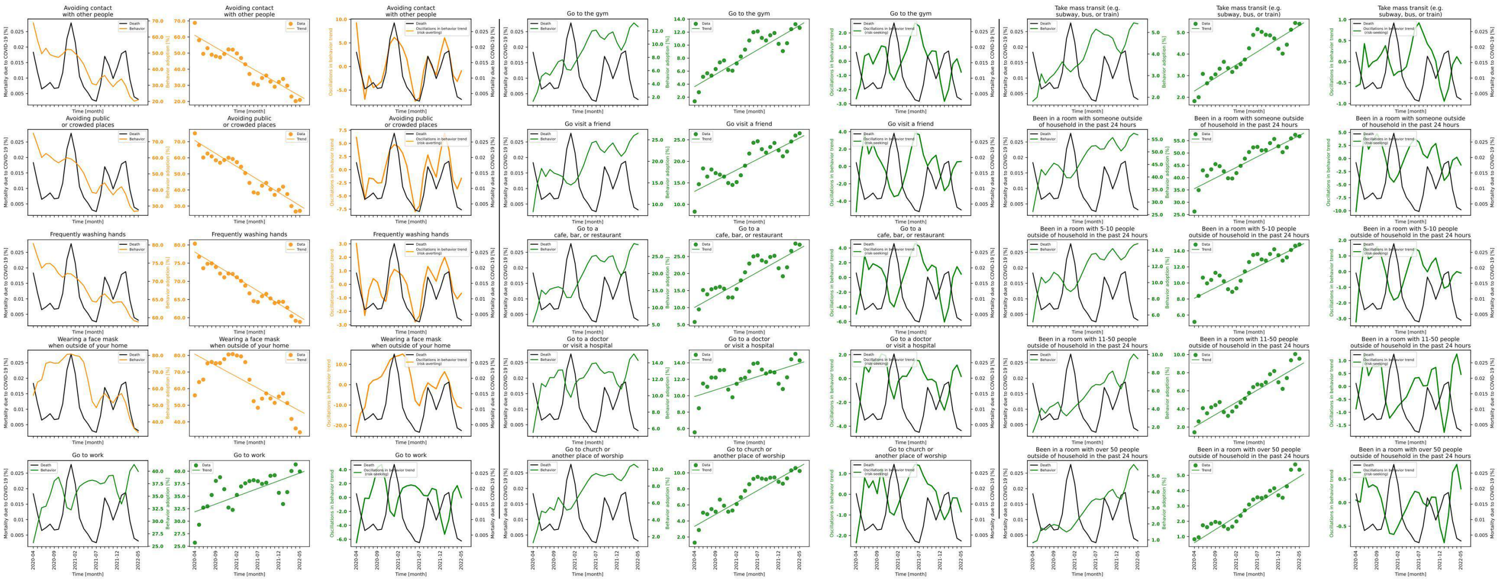

**Figure S7.52 shows behavior and mortality on 1st, 4th and 7th column, behavior trends in 2nd, 5th and 8th column, and behavior oscillatory components juxtaposed with mortality rates on 3rd, 6th and 9th columns for National.**

| Behaviors | Slopes | Y-intercepts | Start %($S$) | End %($E$) | $\|E-S\|$ | $\frac{\|E-S\|*100}{S}$ |
|---|---|---|---|---|---|---|
| Avoiding contact with other people | -1.506 | 61.032 | 68.768 | 20.944 | 47.824 | 69.544 |
| Avoiding public or crowded places | -1.617 | 70.703 | 75.226 | 27.023 | 48.203 | 64.078 |
| Frequently washing hands | -0.713 | 78.058 | 80.356 | 58.845 | 21.511 | 26.77 |
| Wearing a face mask when outside of your home | -1.364 | 80.716 | 55.964 | 33.738 | 22.226 | 39.715 |
| Go to work | 0.31 | 31.884 | 25.708 | 39.944 | 14.236 | 55.376 |
| Go to the gym | 0.376 | 3.627 | 1.363 | 12.648 | 11.285 | 827.953 |
| Go visit a friend | 0.497 | 13.027 | 8.313 | 26.517 | 18.204 | 218.982 |
| Go to a cafe, bar, or restaurant | 0.69 | 10.063 | 5.779 | 28.468 | 22.689 | 392.611 |
| Go to a doctor or visit a hospital | 0.162 | 9.899 | 5.545 | 14.29 | 8.745 | 157.71 |
| Go to church or another place of worship | 0.297 | 3.336 | 1.29 | 10.229 | 8.939 | 692.946 |
| Take mass transit (e.g. subway, bus, or train) | 0.121 | 2.29 | 1.819 | 5.4 | 3.581 | 196.866 |
| Been in a room with someone outside of household in the past 24 hours | 0.864 | 35.535 | 26.257 | 56.766 | 30.509 | 116.194 |
| Been in a room with 5-10 people outside of household in the past 24 hours | 0.261 | 8.097 | 5.15 | 14.795 | 9.645 | 187.282 |
| Been in a room with 11-50 people outside of household in the past 24 hours | 0.271 | 2.015 | 1.42 | 9.543 | 8.123 | 572.042 |
| Been in a room with over 50 people outside of household in the past 24 hours | 0.172 | 0.601 | 0.826 | 5.364 | 4.538 | 549.395 |

**Table S7.52 shows the slopes, y-intercept, adherence % at the beginning of the study period, and at the end, the absolute difference between the beginning and the end, and the absolute difference as a percentage of the adherence % at the beginning of the study period of each behavior's trend in National.**

### 8 State-level correlations between oscillations in behavior trends and disease severity metrics

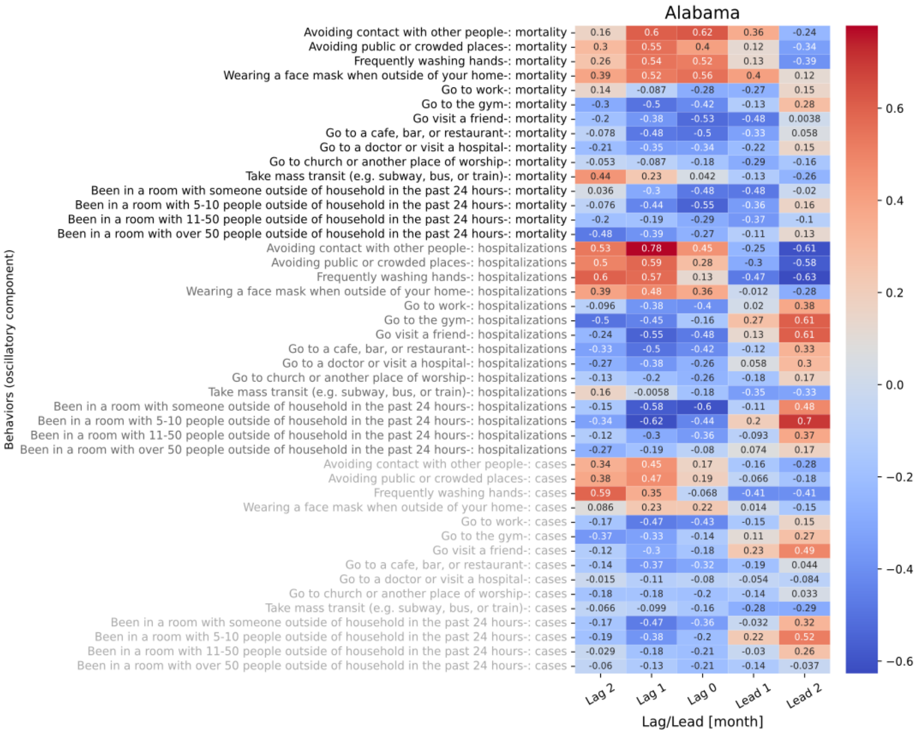

**Figure S8.1 shows the 5 lagged correlations of oscillations in behavior trend with mortality (black), hospitalizations (gray), and cases (light gray) for Alabama.**

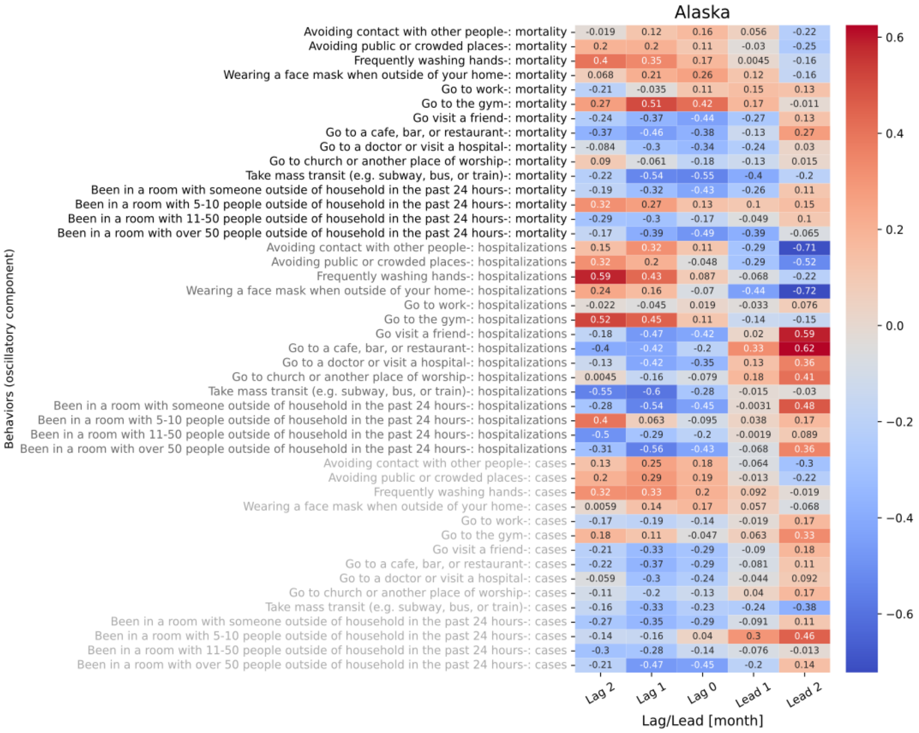

**Figure S8.2 shows the 5 lagged correlations of oscillations in behavior trend with mortality (black), hospitalizations (gray), and cases (light gray) for Alaska.**

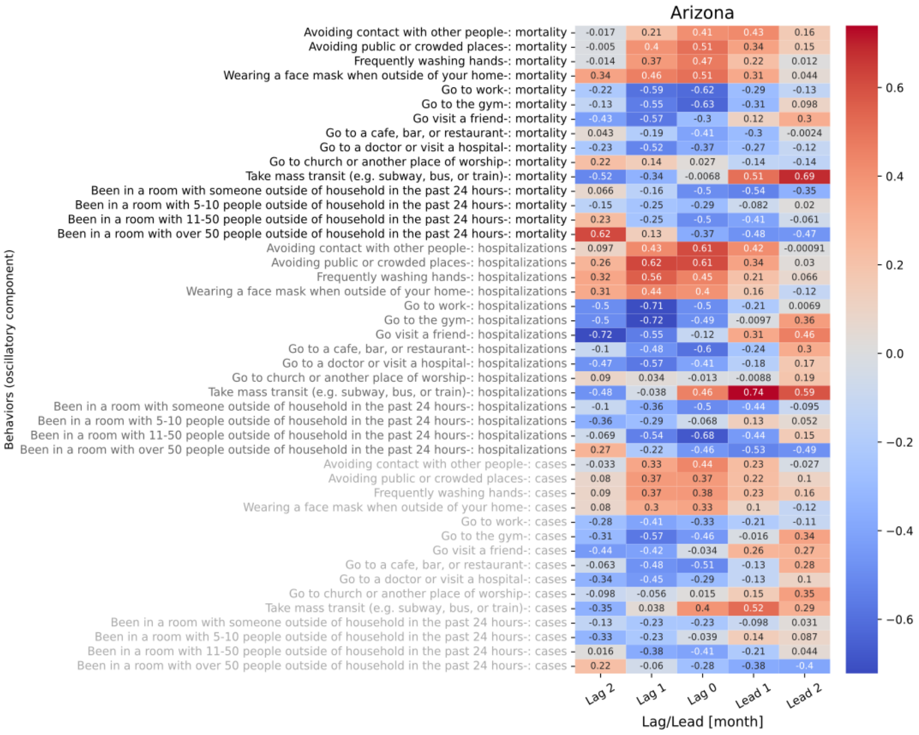

**Figure S8.3 shows the 5 lagged correlations of oscillations in behavior trend with mortality (black), hospitalizations (gray), and cases (light gray) for Arizona.**

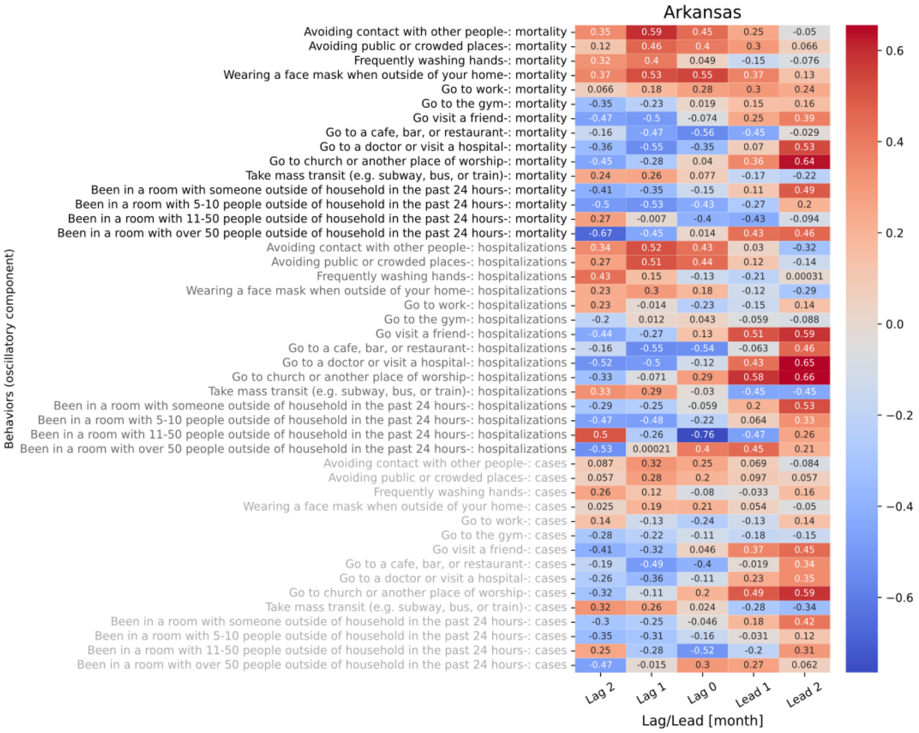

**Figure S8.4 shows the 5 lagged correlations of oscillations in behavior trend with mortality (black), hospitalizations (gray), and cases (light gray) for Arkansas.**

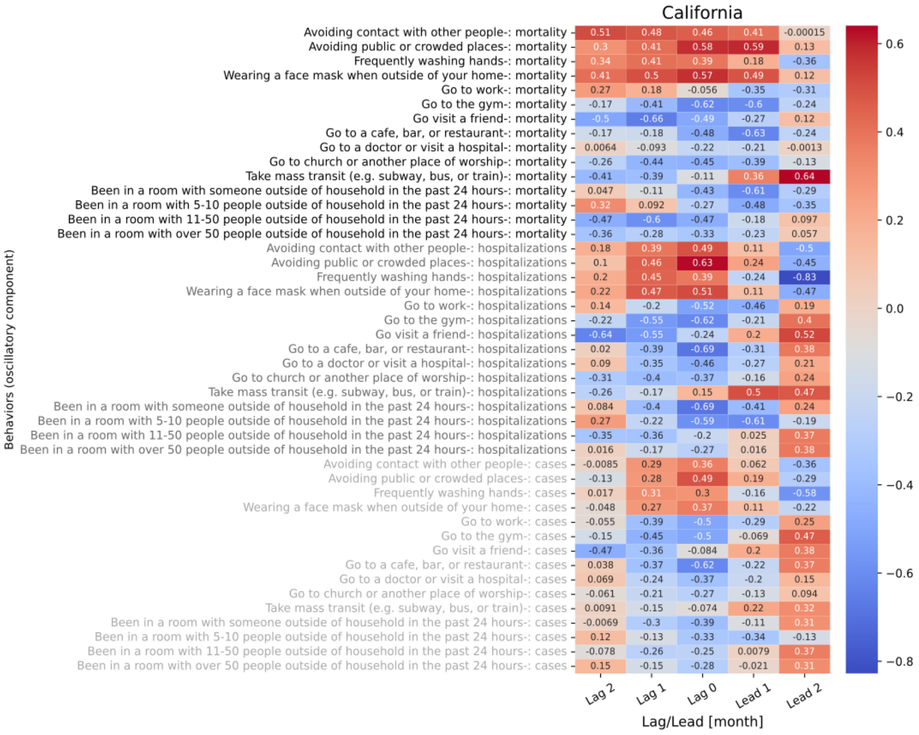

**Figure S8.5 shows the 5 lagged correlations of oscillations in behavior trend with mortality (black), hospitalizations (gray), and cases (light gray) for California.**

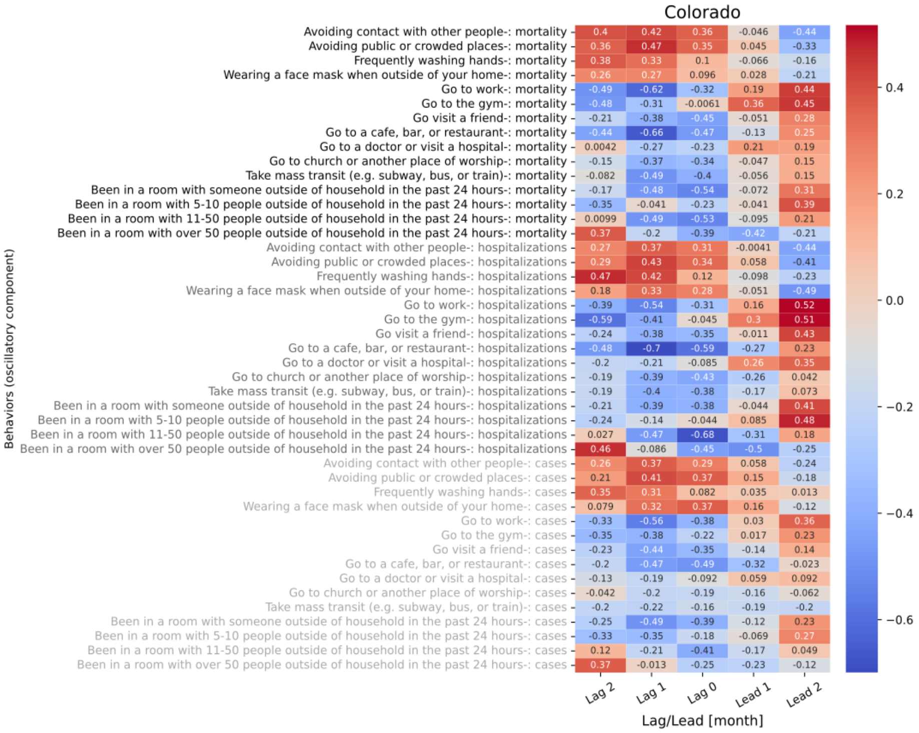

**Figure S8.6 shows the 5 lagged correlations of oscillations in behavior trend with mortality (black), hospitalizations (gray), and cases (light gray) for Colorado.**

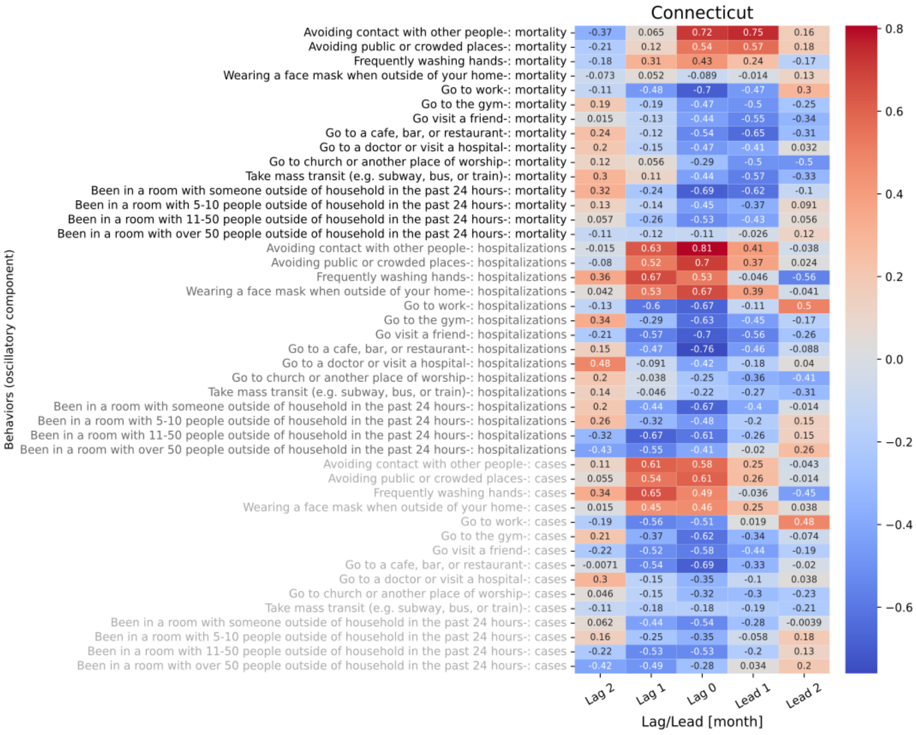

**Figure S8.7 shows the 5 lagged correlations of oscillations in behavior trend with mortality (black), hospitalizations (gray), and cases (light gray) for Connecticut.**

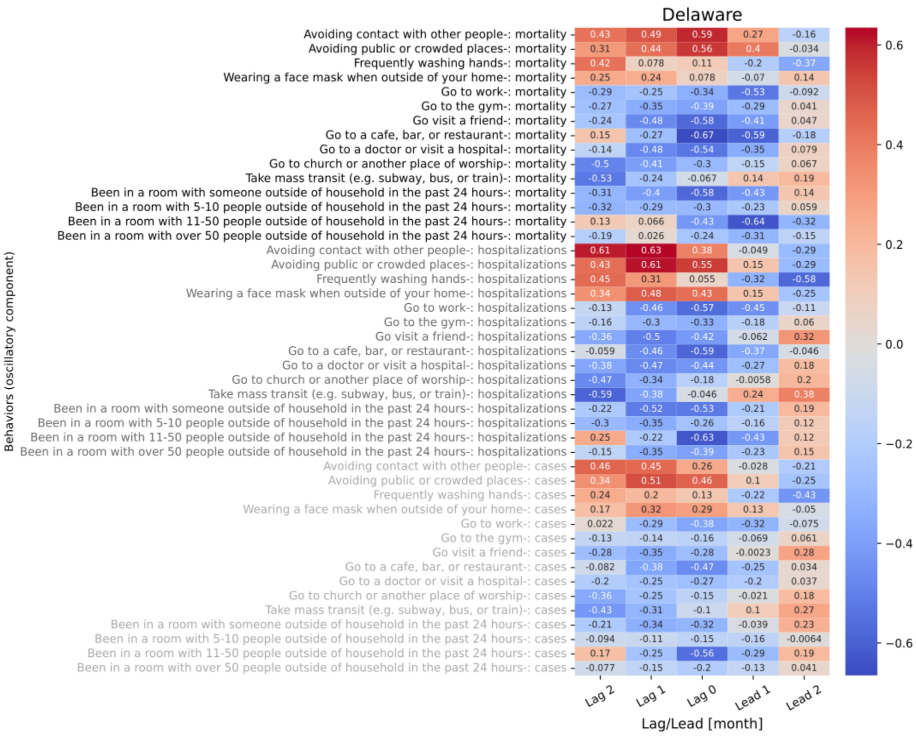

**Figure S8.8 shows the 5 lagged correlations of oscillations in behavior trend with mortality (black), hospitalizations (gray), and cases (light gray) for Delaware.**

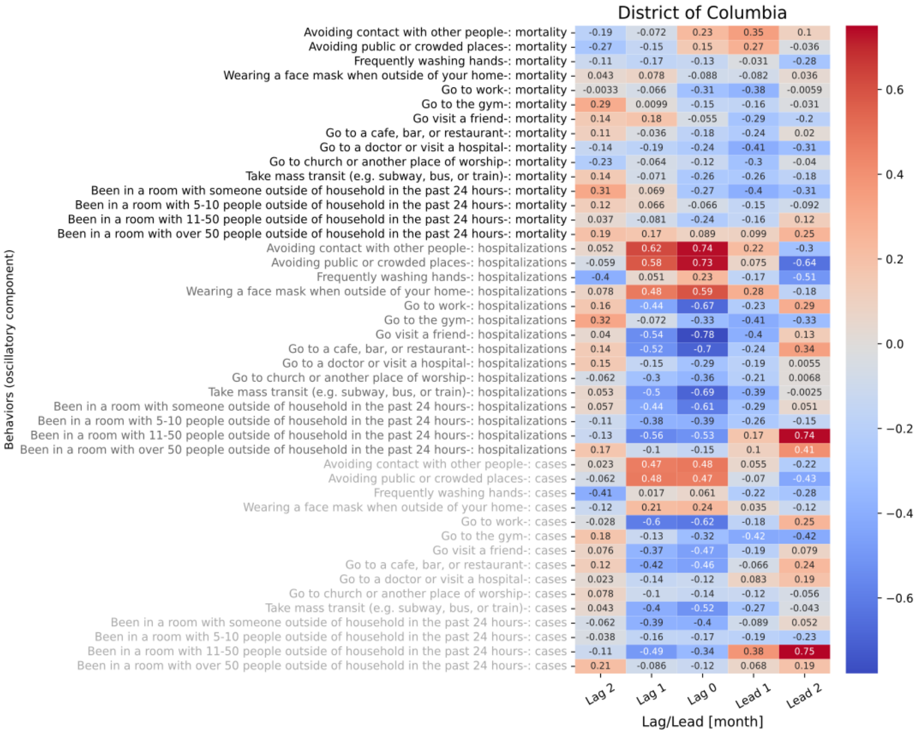

**Figure S8.9 shows the 5 lagged correlations of oscillations in behavior trend with mortality (black), hospitalizations (gray), and cases (light gray) for District of Columbia.**

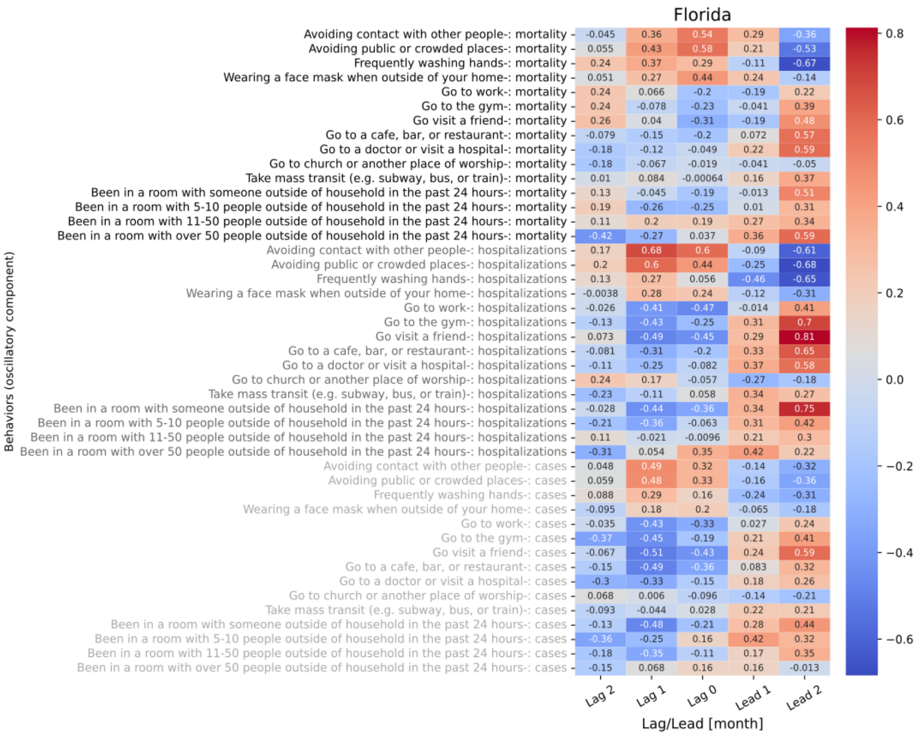

**Figure S8.10 shows the 5 lagged correlations of oscillations in behavior trend with mortality (black), hospitalizations (gray), and cases (light gray) for Florida.**

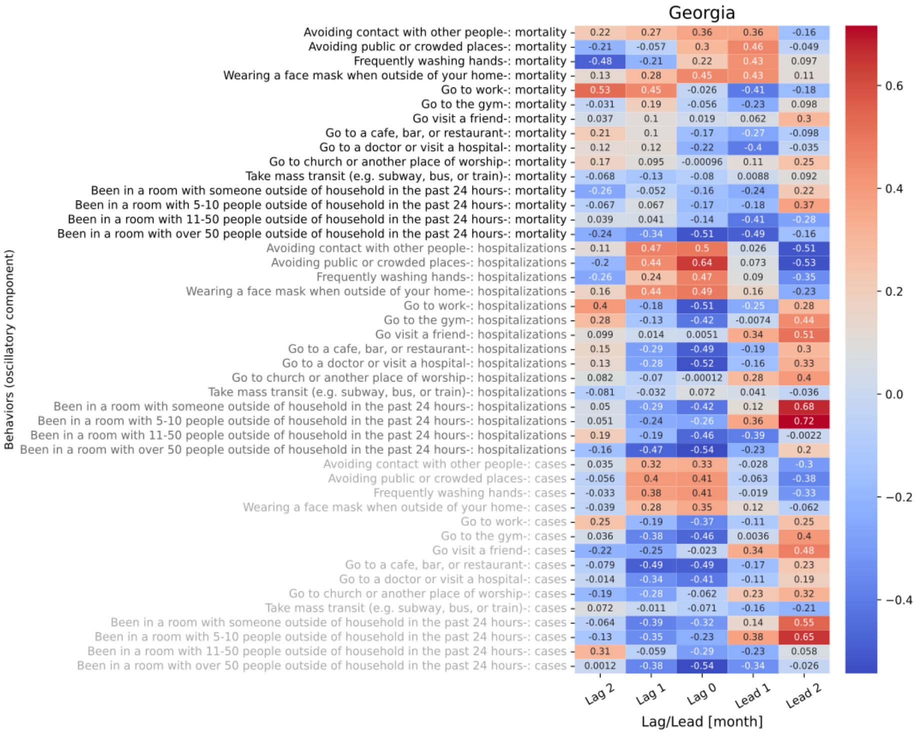

**Figure S8.11 shows the 5 lagged correlations of oscillations in behavior trend with mortality (black), hospitalizations (gray), and cases (light gray) for Georgia.**

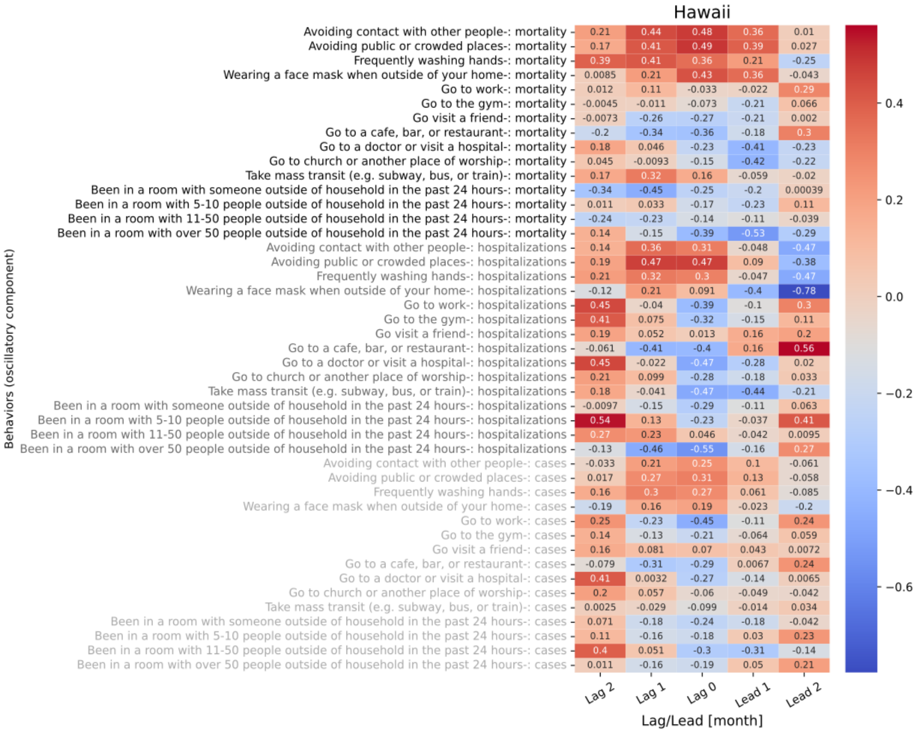

**Figure S8.12 shows the 5 lagged correlations of oscillations in behavior trend with mortality (black), hospitalizations (gray), and cases (light gray) for Hawaii.**

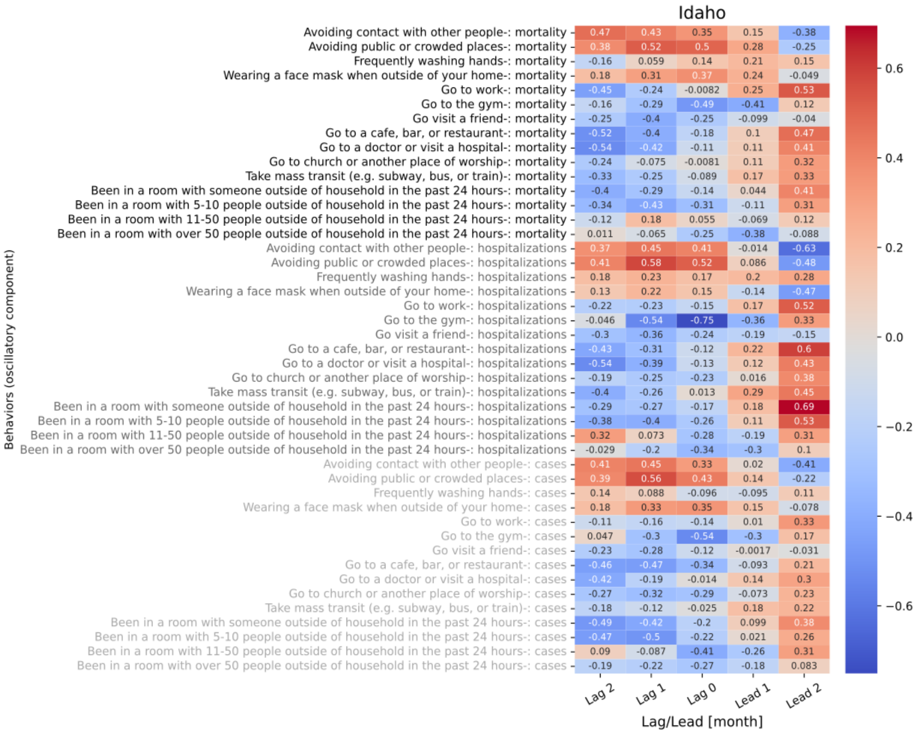

**Figure S8.13 shows the 5 lagged correlations of oscillations in behavior trend with mortality (black), hospitalizations (gray), and cases (light gray) for Idaho.**

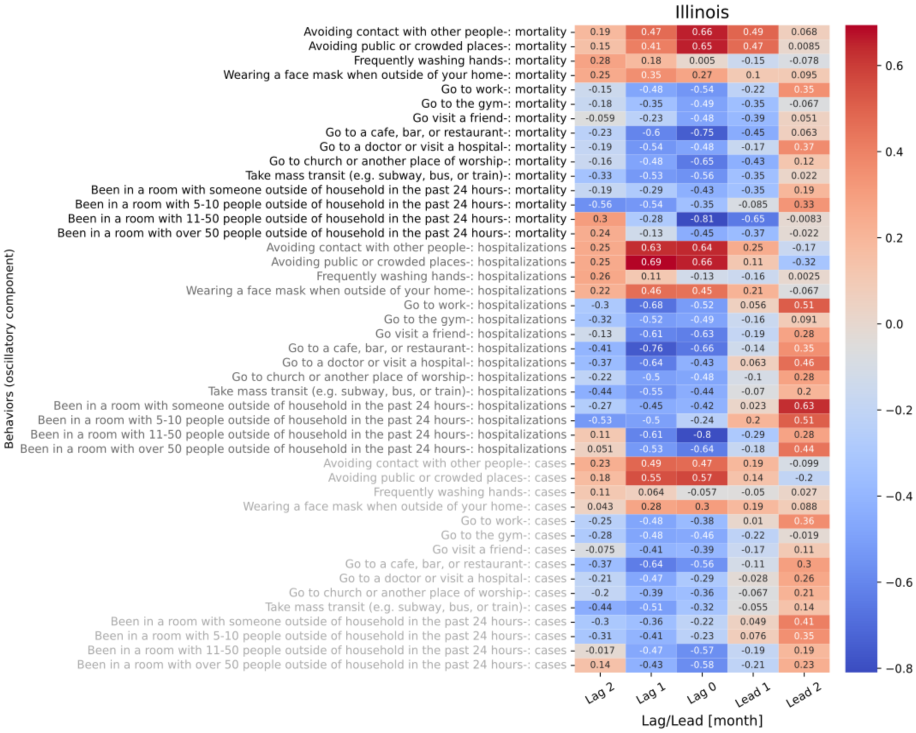

**Figure S8.14 shows the 5 lagged correlations of oscillations in behavior trend with mortality (black), hospitalizations (gray), and cases (light gray) for Illinois.**

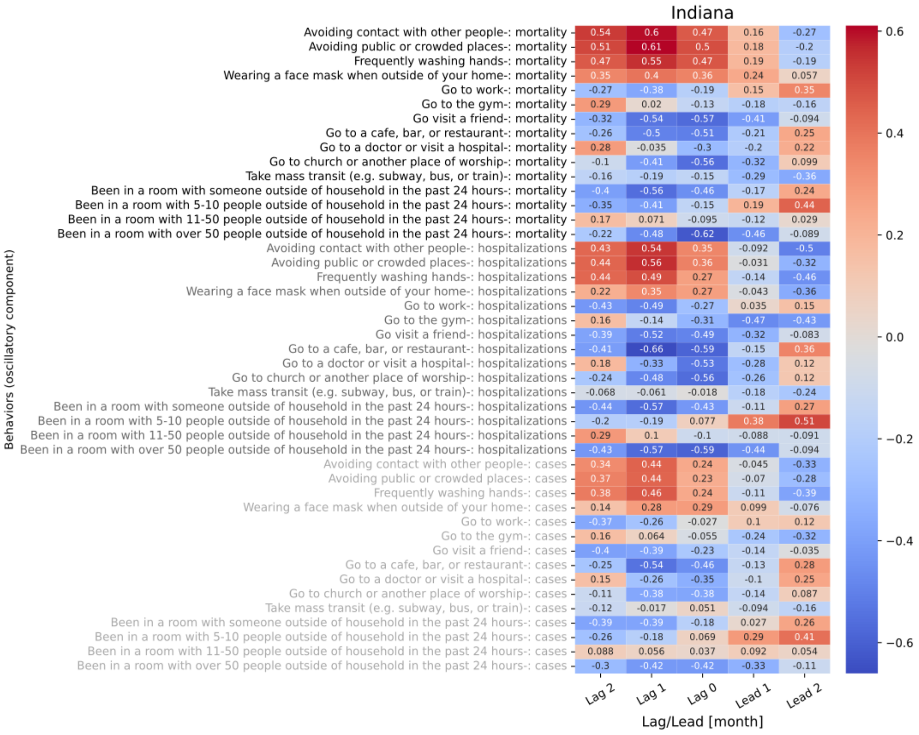

**Figure S8.15 shows the 5 lagged correlations of oscillations in behavior trend with mortality (black), hospitalizations (gray), and cases (light gray) for Indiana.**

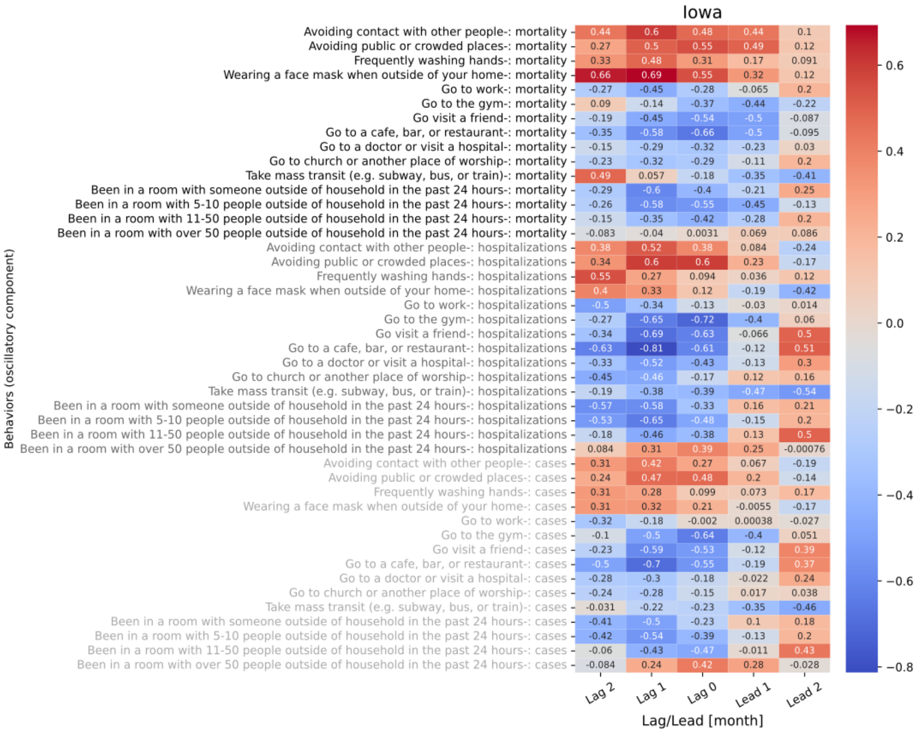

**Figure S8.16 shows the 5 lagged correlations of oscillations in behavior trend with mortality (black), hospitalizations (gray), and cases (light gray) for Iowa.**

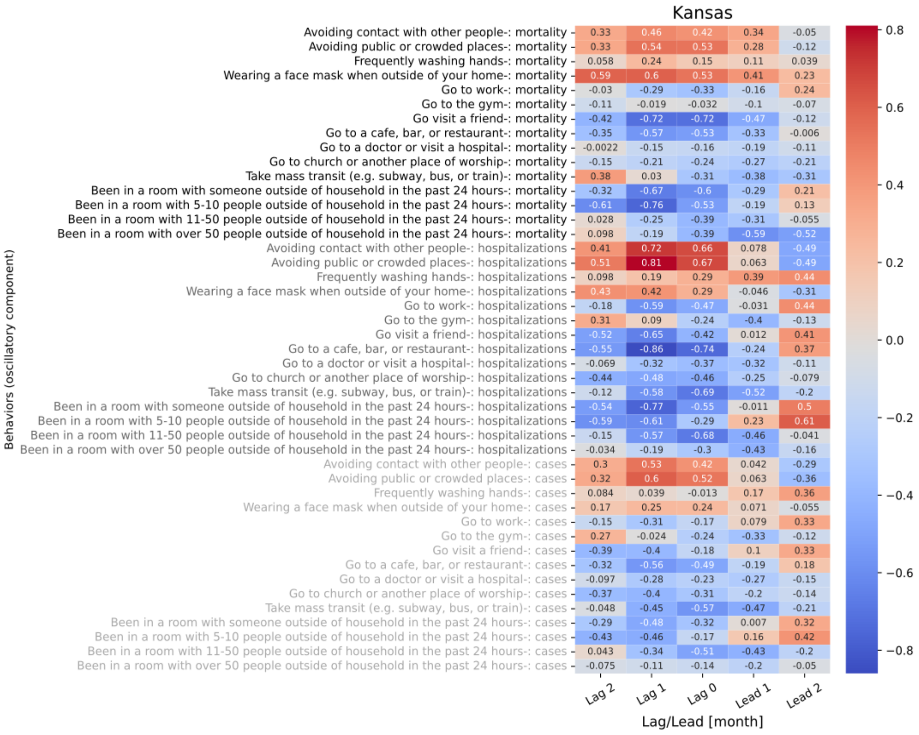

**Figure S8.17 shows the 5 lagged correlations of oscillations in behavior trend with mortality (black), hospitalizations (gray), and cases (light gray) for Kansas.**

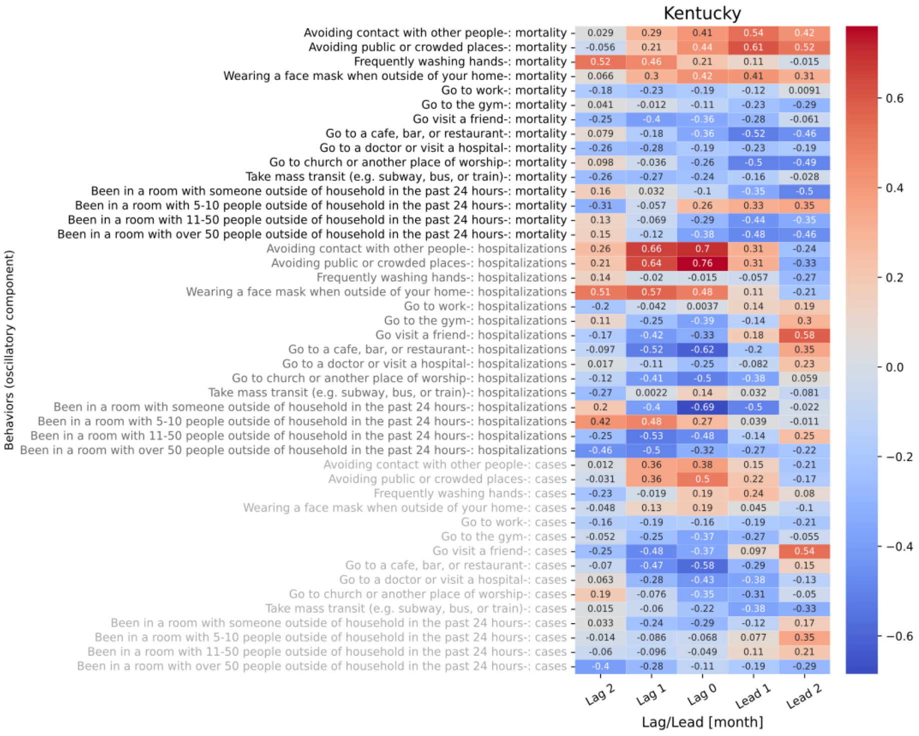

**Figure S8.18 shows the 5 lagged correlations of oscillations in behavior trend with mortality (black), hospitalizations (gray), and cases (light gray) for Kentucky.**

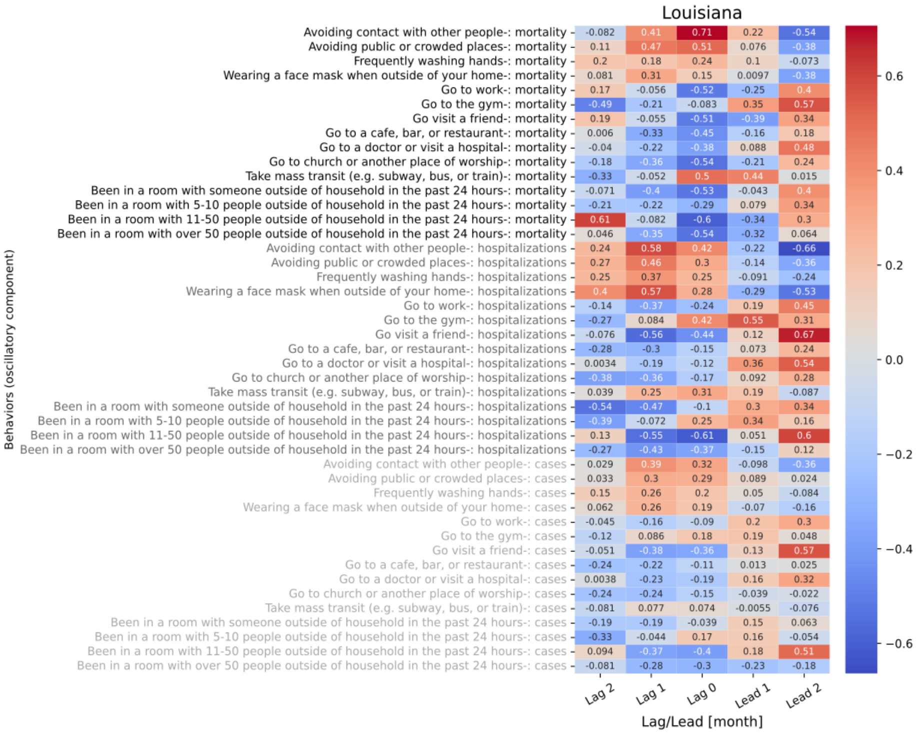

**Figure S8.19 shows the 5 lagged correlations of oscillations in behavior trend with mortality (black), hospitalizations (gray), and cases (light gray) for Louisiana.**

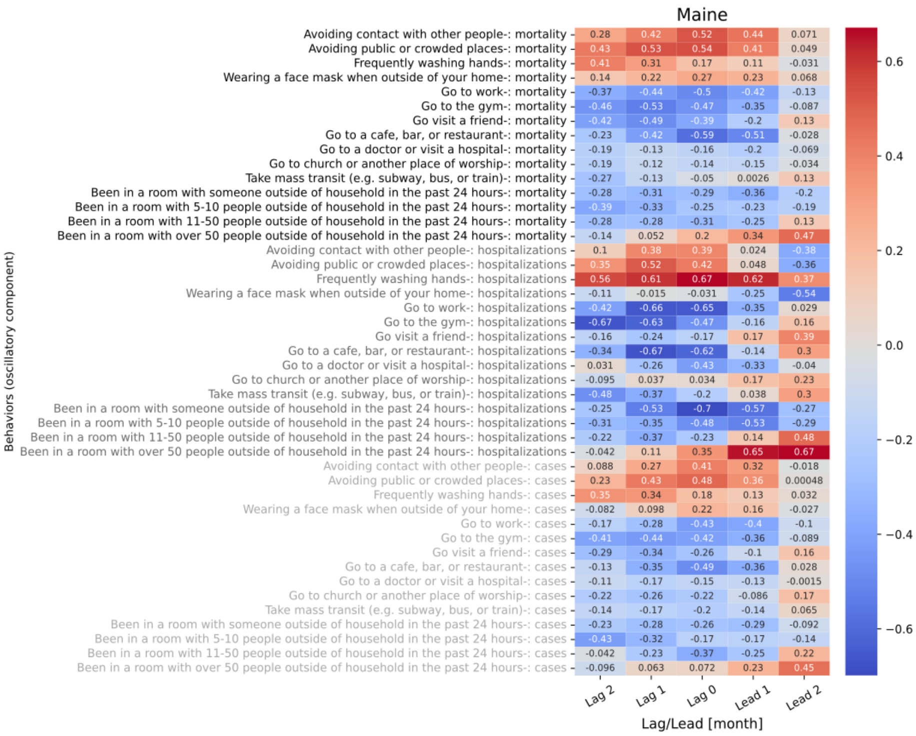

**Figure S8.20 shows the 5 lagged correlations of oscillations in behavior trend with mortality (black), hospitalizations (gray), and cases (light gray) for Maine.**

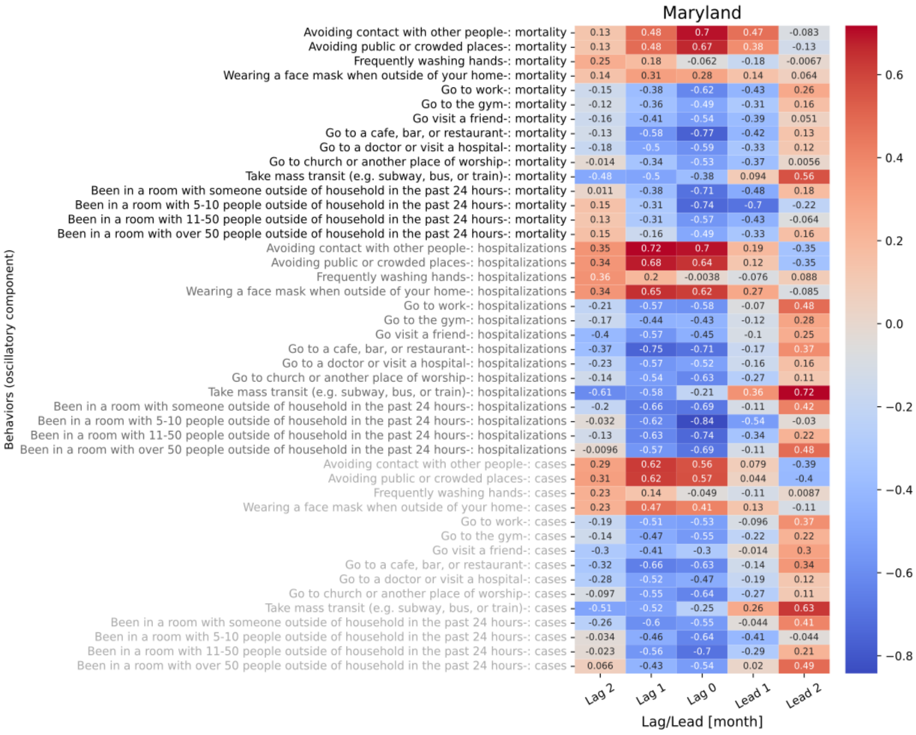

**Figure S8.21 shows the 5 lagged correlations of oscillations in behavior trend with mortality (black), hospitalizations (gray), and cases (light gray) for Maryland.**

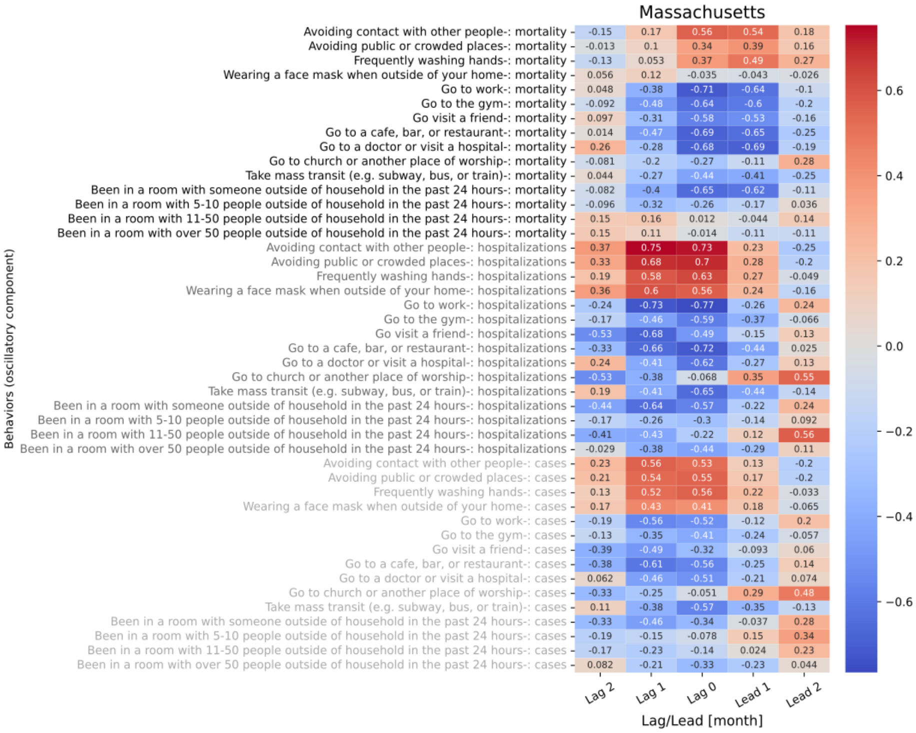

**Figure S8.22 shows the 5 lagged correlations of oscillations in behavior trend with mortality (black), hospitalizations (gray), and cases (light gray) for Massachusetts.**

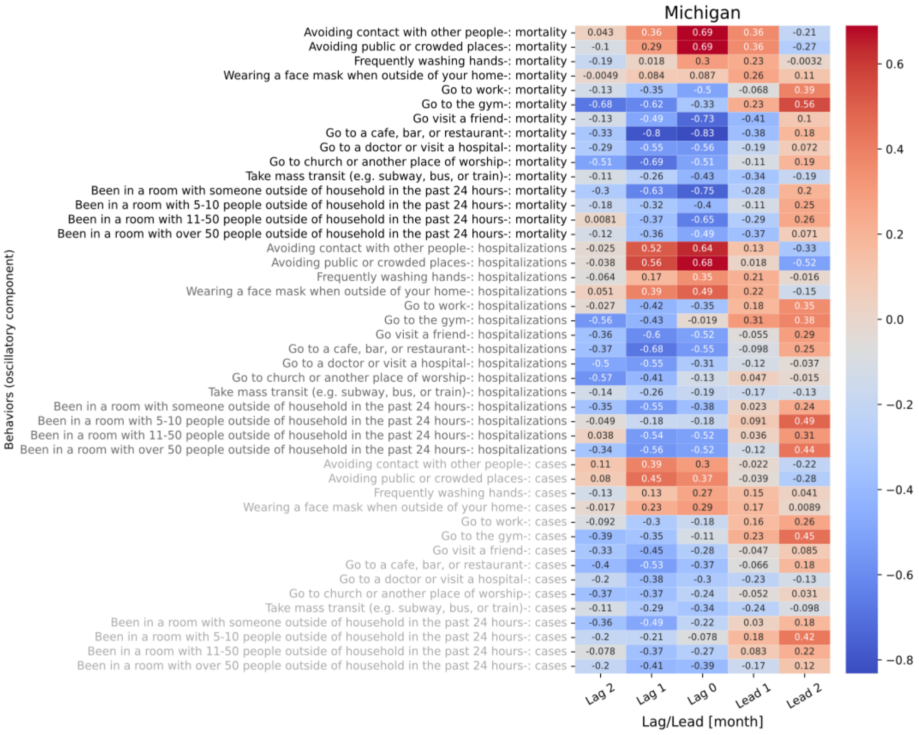

**Figure S8.23 shows the 5 lagged correlations of oscillations in behavior trend with mortality (black), hospitalizations (gray), and cases (light gray) for Michigan.**

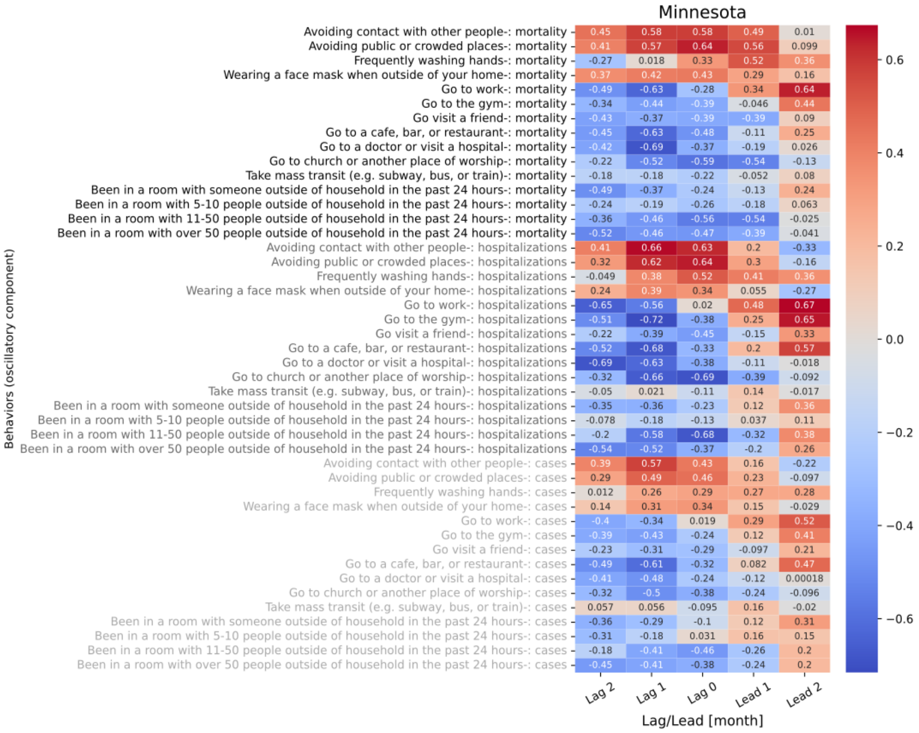

**Figure S8.24 shows the 5 lagged correlations of oscillations in behavior trend with mortality (black), hospitalizations (gray), and cases (light gray) for Minnesota.**

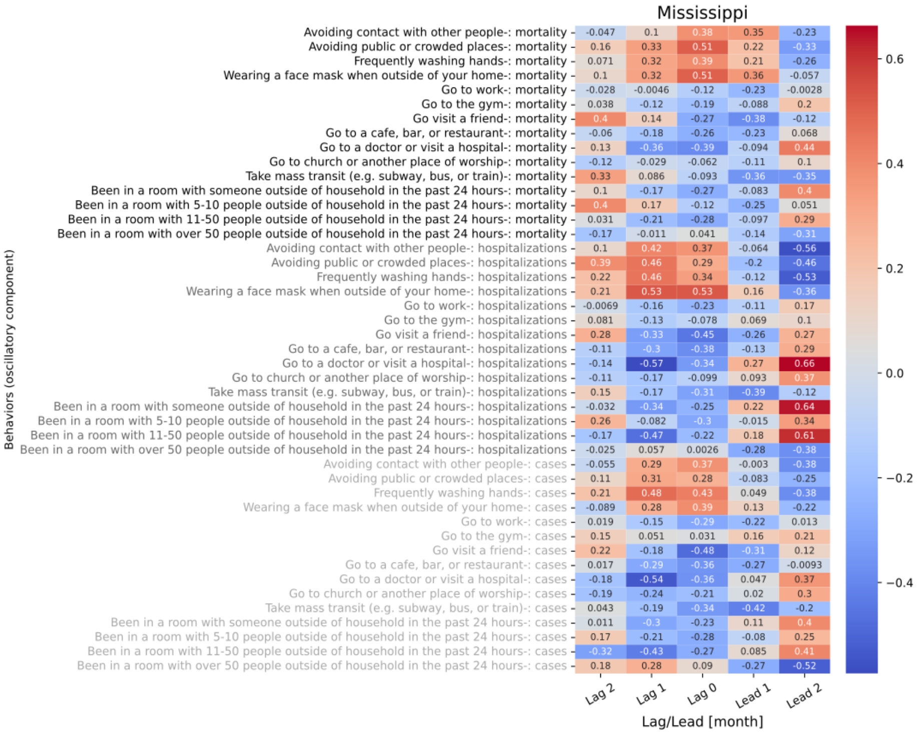

**Figure S8.25 shows the 5 lagged correlations of oscillations in behavior trend with mortality (black), hospitalizations (gray), and cases (light gray) for Mississippi.**

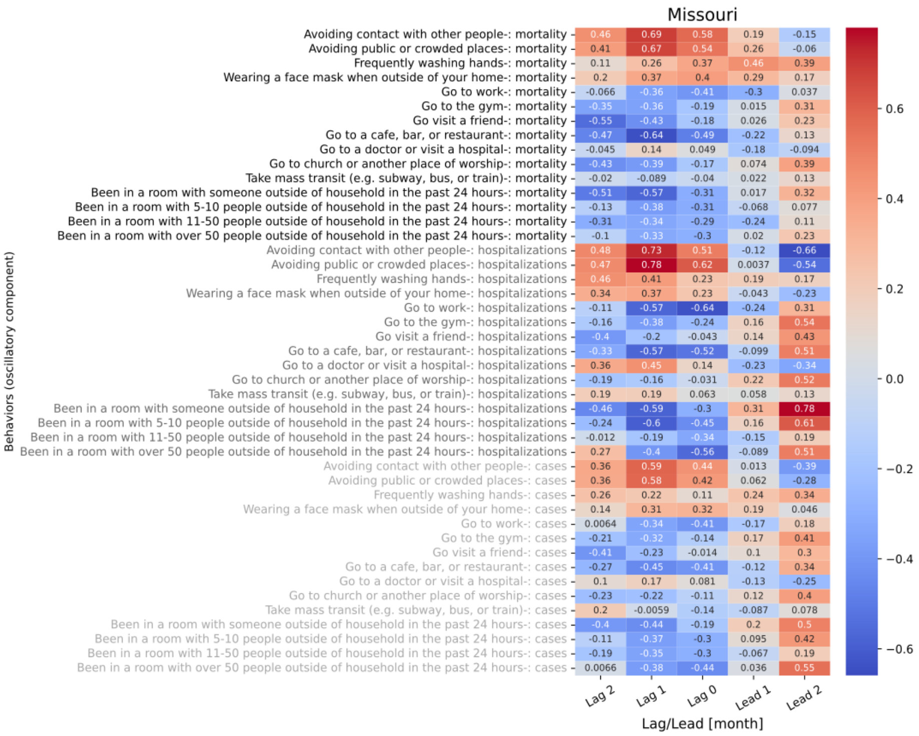

**Figure S8.26 shows the 5 lagged correlations of oscillations in behavior trend with mortality (black), hospitalizations (gray), and cases (light gray) for Missouri.**

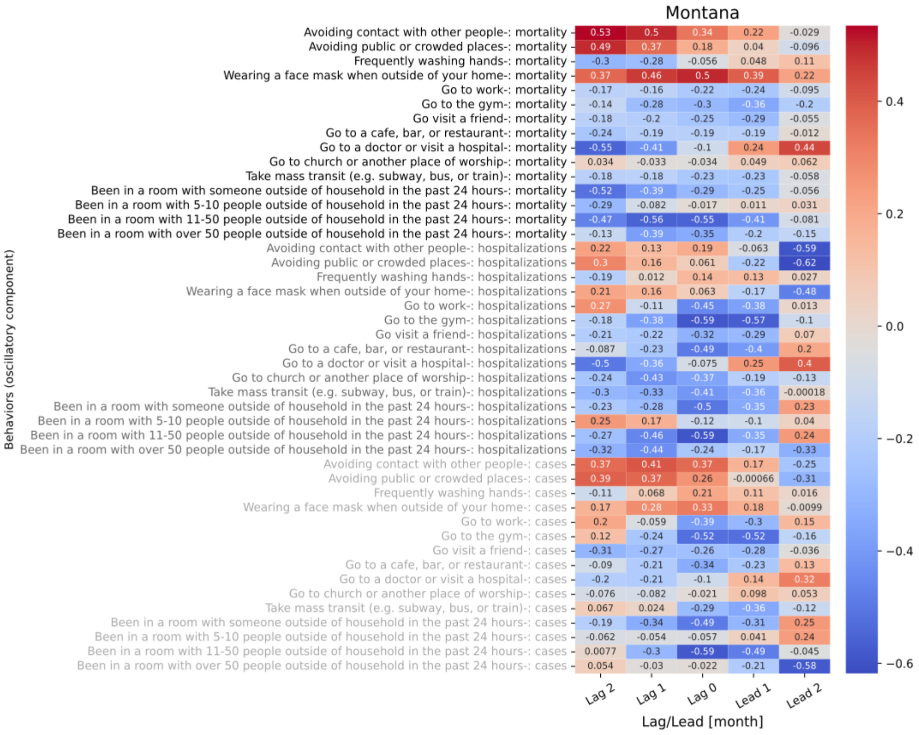

Figure S8.27 shows the 5 lagged correlations of oscillations in behavior trend with mortality (black), hospitalizations (gray), and cases (light gray) for Montana.

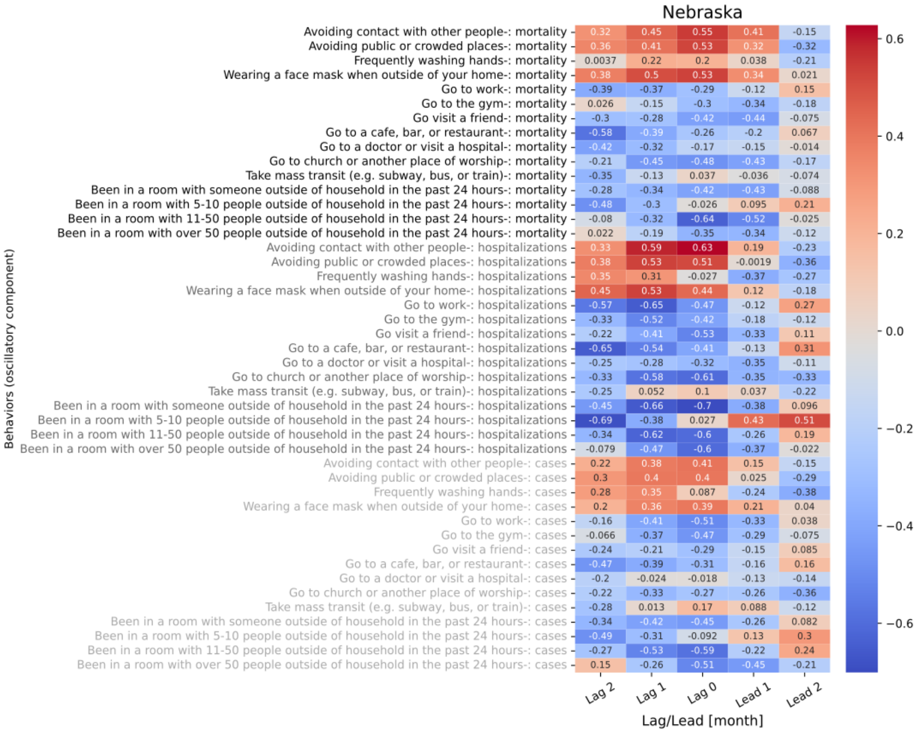

Figure S8.28 shows the 5 lagged correlations of oscillations in behavior trend with mortality (black), hospitalizations (gray), and cases (light gray) for Nebraska.

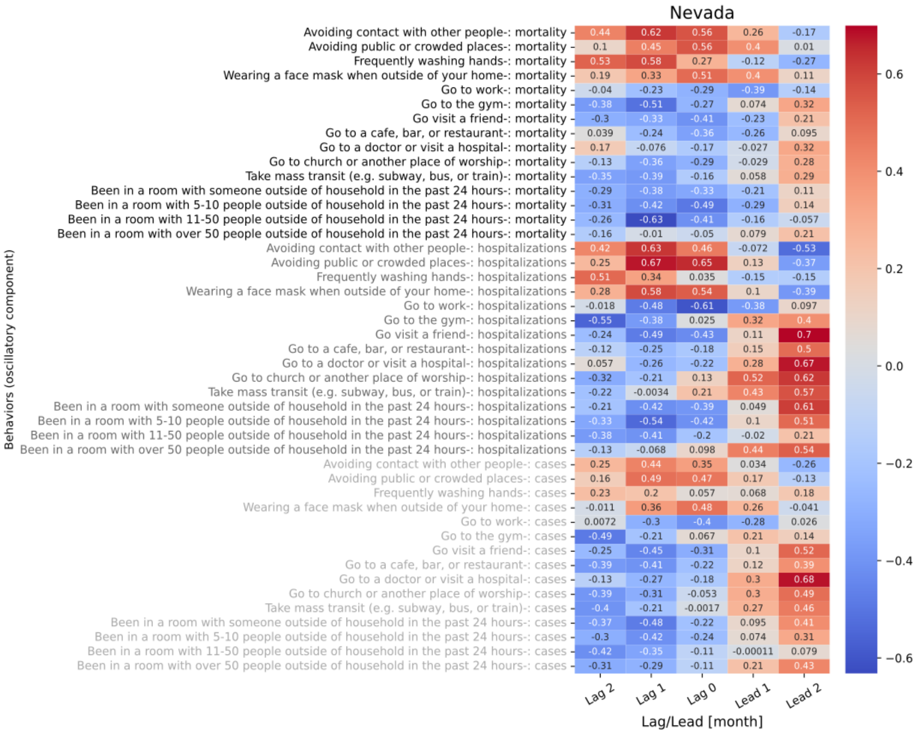

Figure S8.29 shows the 5 lagged correlations of oscillations in behavior trend with mortality (black), hospitalizations (gray), and cases (light gray) for Nevada.

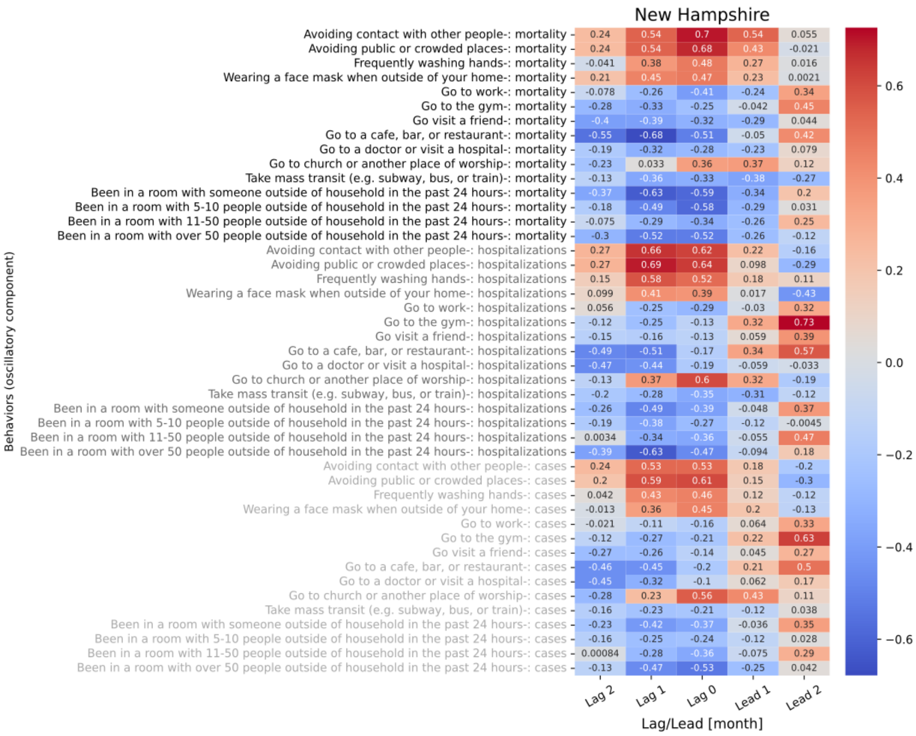

Figure S8.30 shows the 5 lagged correlations of oscillations in behavior trend with mortality (black), hospitalizations (gray), and cases (light gray) for New Hampshire.

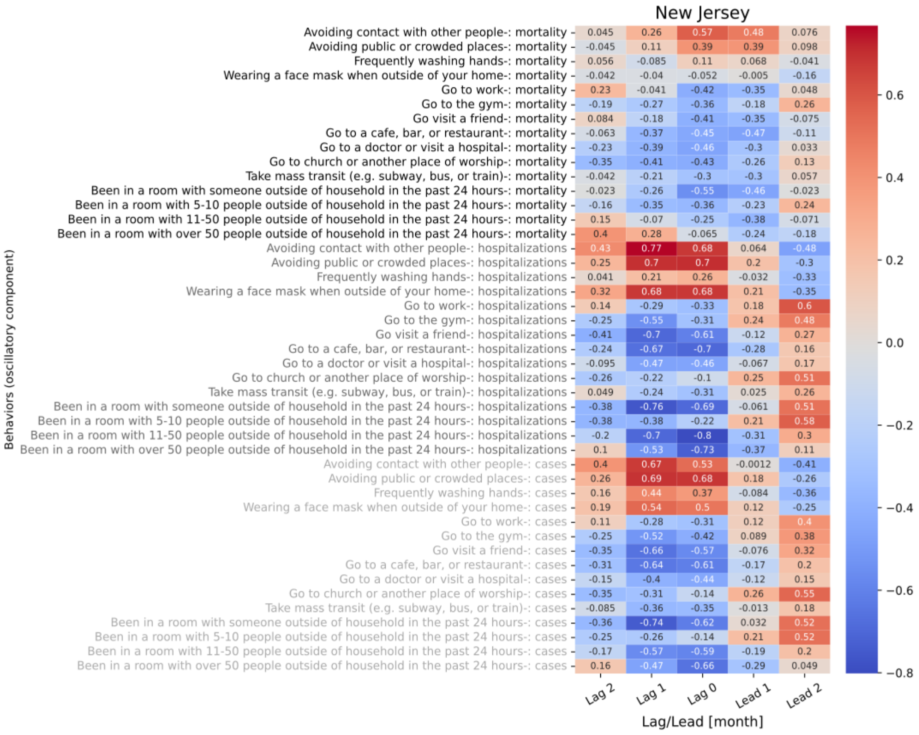

**Figure S8.31 shows the 5 lagged correlations of oscillations in behavior trend with mortality (black), hospitalizations (gray), and cases (light gray) for New Jersey.**

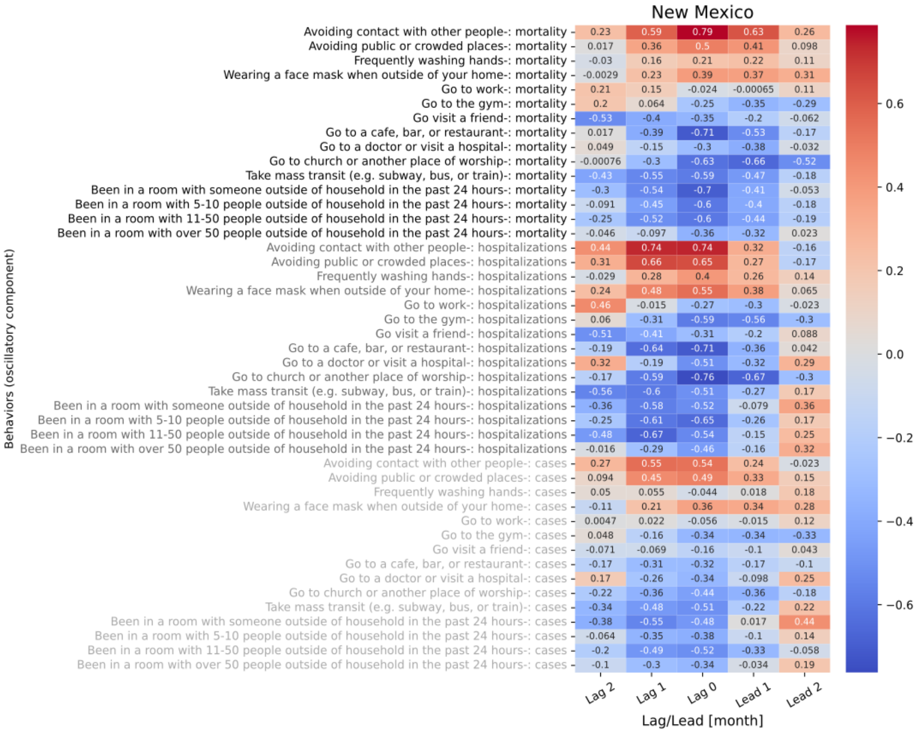

**Figure S8.32 shows the 5 lagged correlations of oscillations in behavior trend with mortality (black), hospitalizations (gray), and cases (light gray) for New Mexico.**

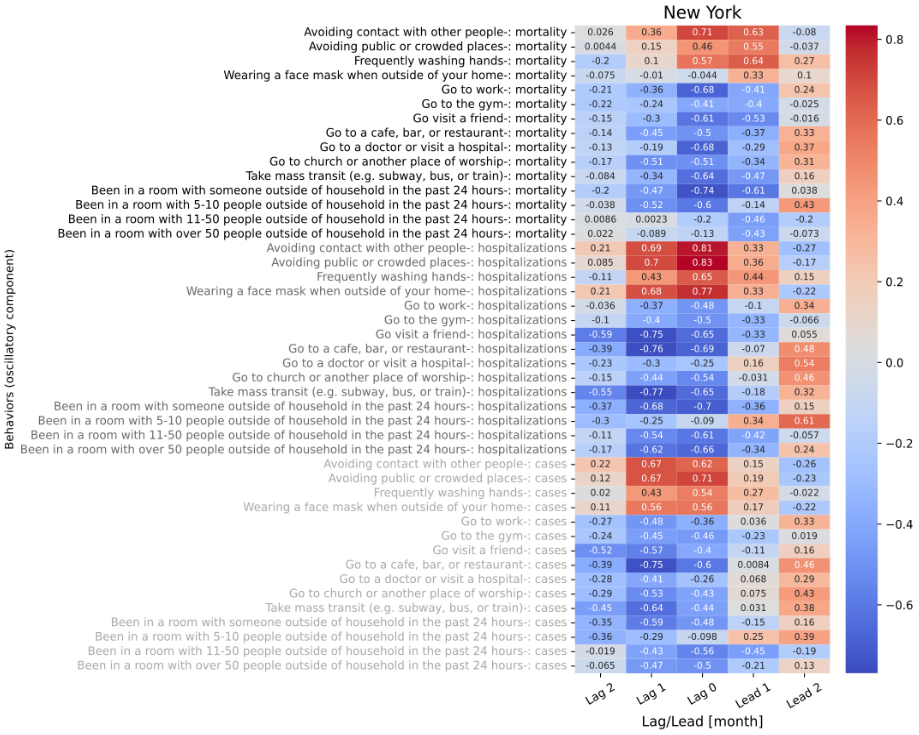

**Figure S8.33 shows the 5 lagged correlations of oscillations in behavior trend with mortality (black), hospitalizations (gray), and cases (light gray) for New York.**

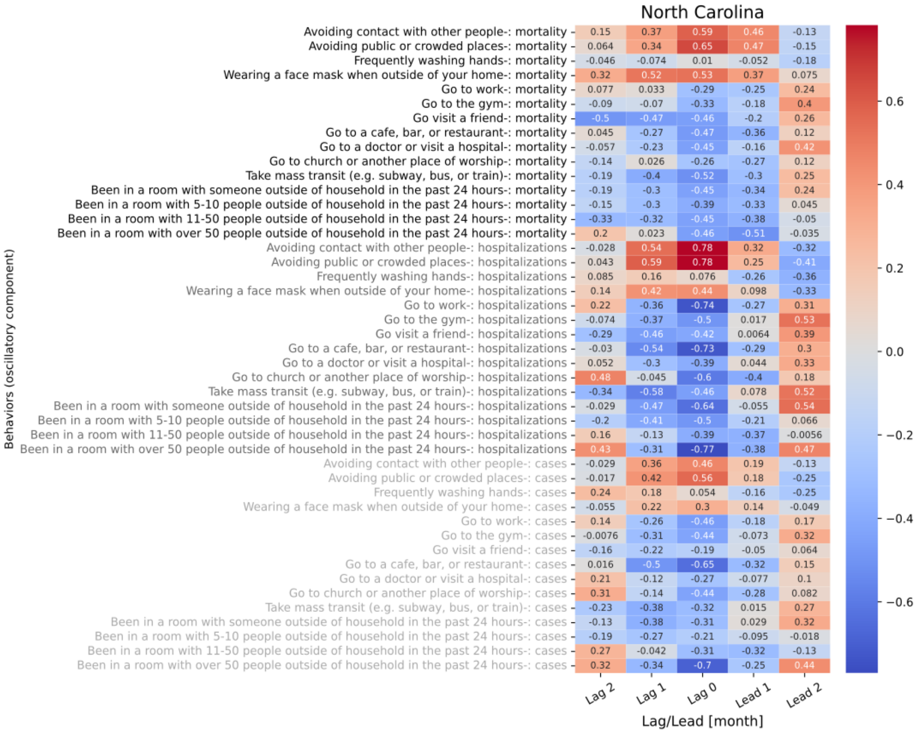

**Figure S8.34 shows the 5 lagged correlations of oscillations in behavior trend with mortality (black), hospitalizations (gray), and cases (light gray) for North Carolina.**

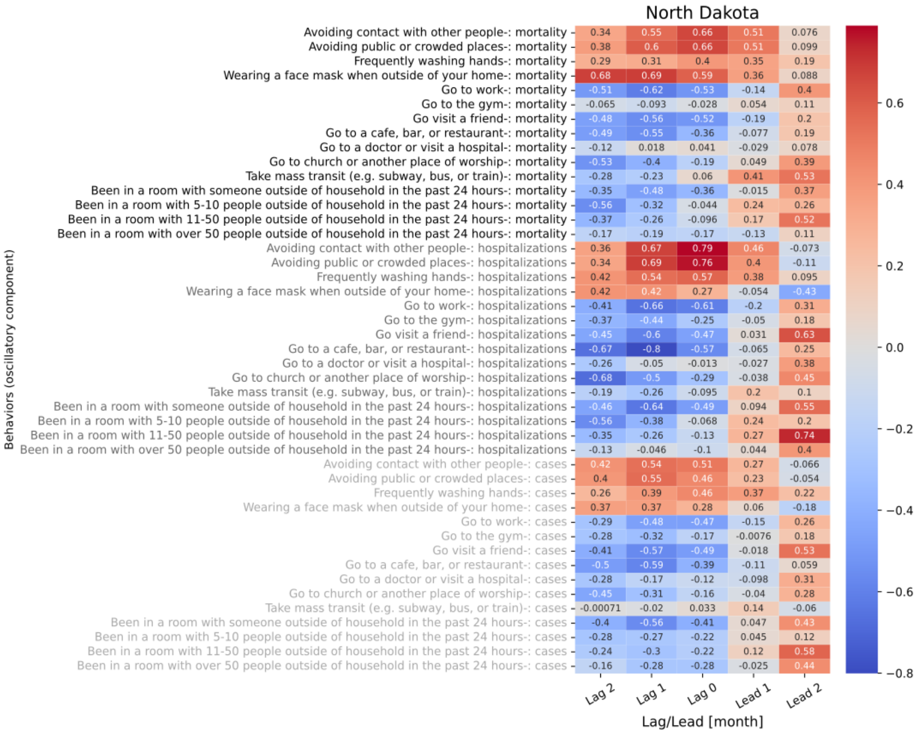

Figure S8.35 shows the 5 lagged correlations of oscillations in behavior trend with mortality (black), hospitalizations (gray), and cases (light gray) for North Dakota.

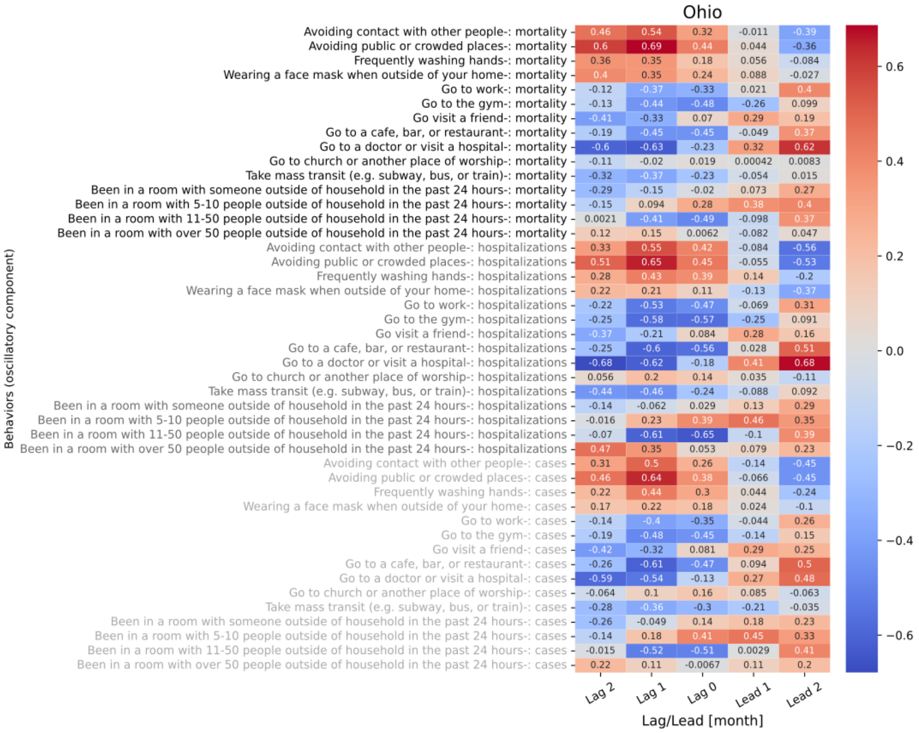

Figure S8.36 shows the 5 lagged correlations of oscillations in behavior trend with mortality (black), hospitalizations (gray), and cases (light gray) for Ohio.

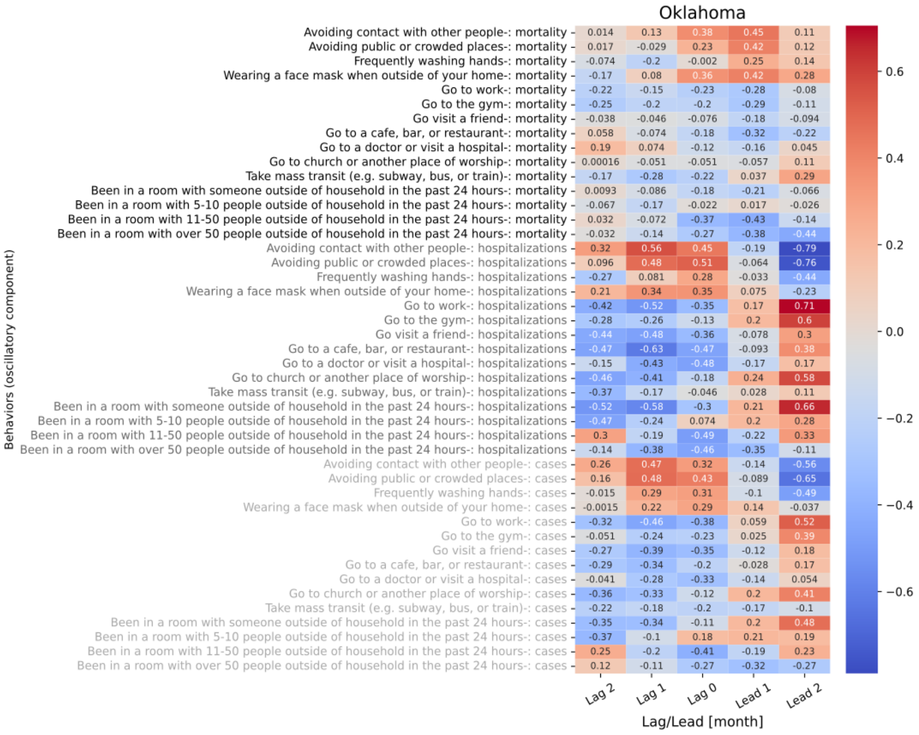

**Figure S8.37 shows the 5 lagged correlations of oscillations in behavior trend with mortality (black), hospitalizations (gray), and cases (light gray) for Oklahoma.**

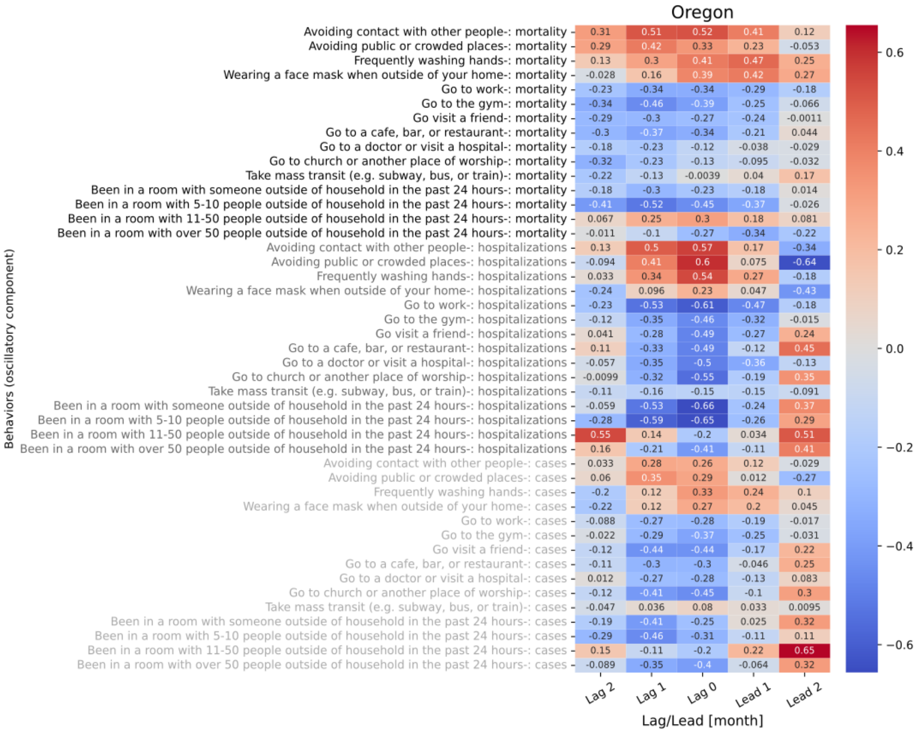

**Figure S8.38 shows the 5 lagged correlations of oscillations in behavior trend with mortality (black), hospitalizations (gray), and cases (light gray) for Oregon.**

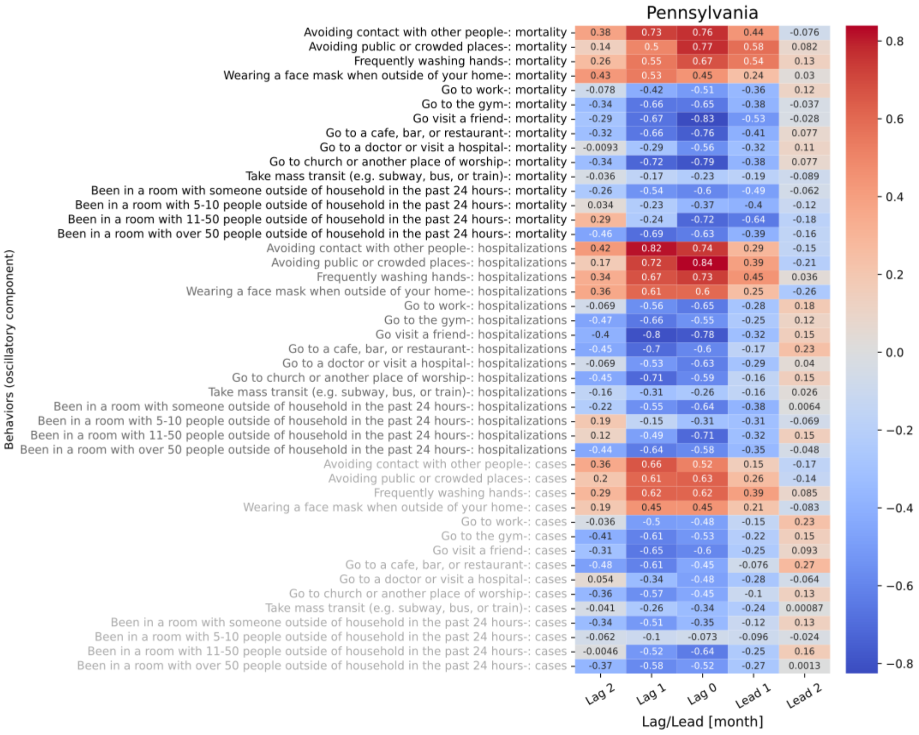

**Figure S8.39 shows the 5 lagged correlations of oscillations in behavior trend with mortality (black), hospitalizations (gray), and cases (light gray) for Pennsylvania.**

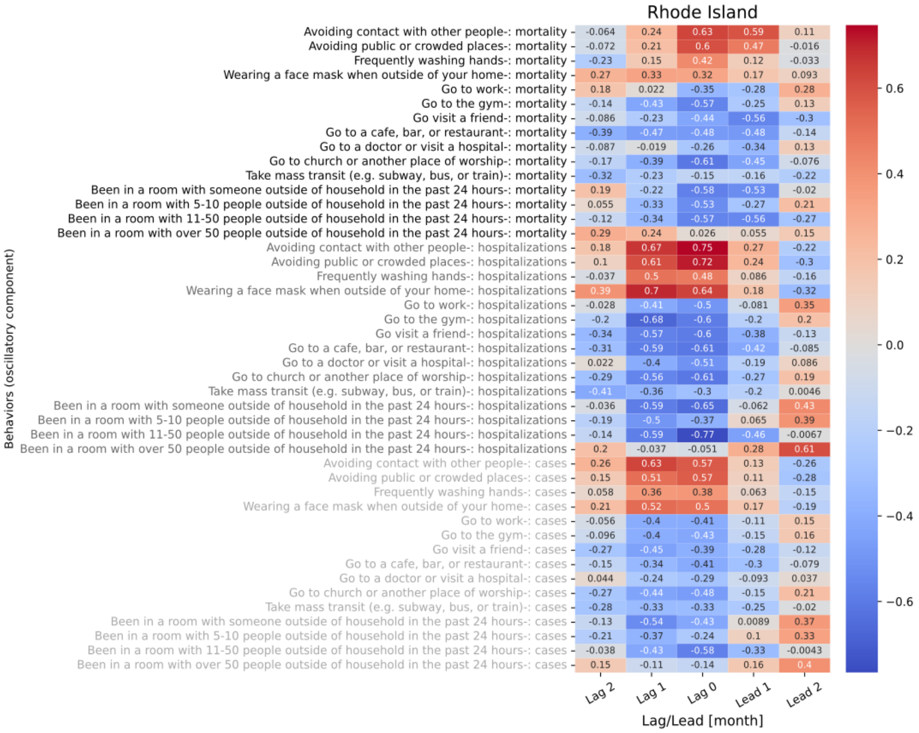

**Figure S8.40 shows the 5 lagged correlations of oscillations in behavior trend with mortality (black), hospitalizations (gray), and cases (light gray) for Rhode Island.**

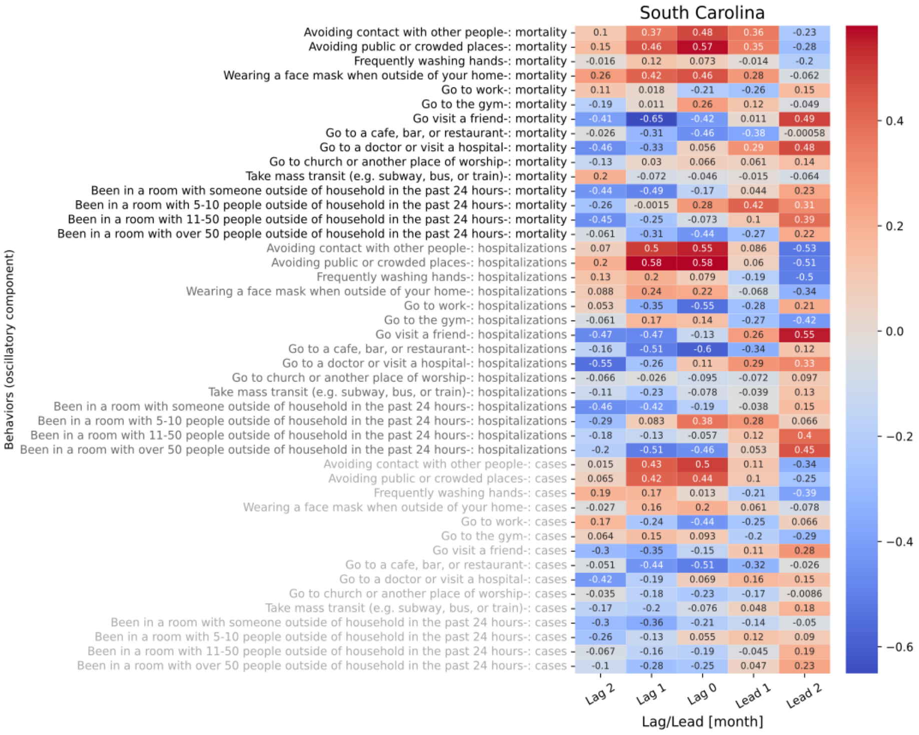

**Figure S8.41 shows the 5 lagged correlations of oscillations in behavior trend with mortality (black), hospitalizations (gray), and cases (light gray) for South Carolina.**

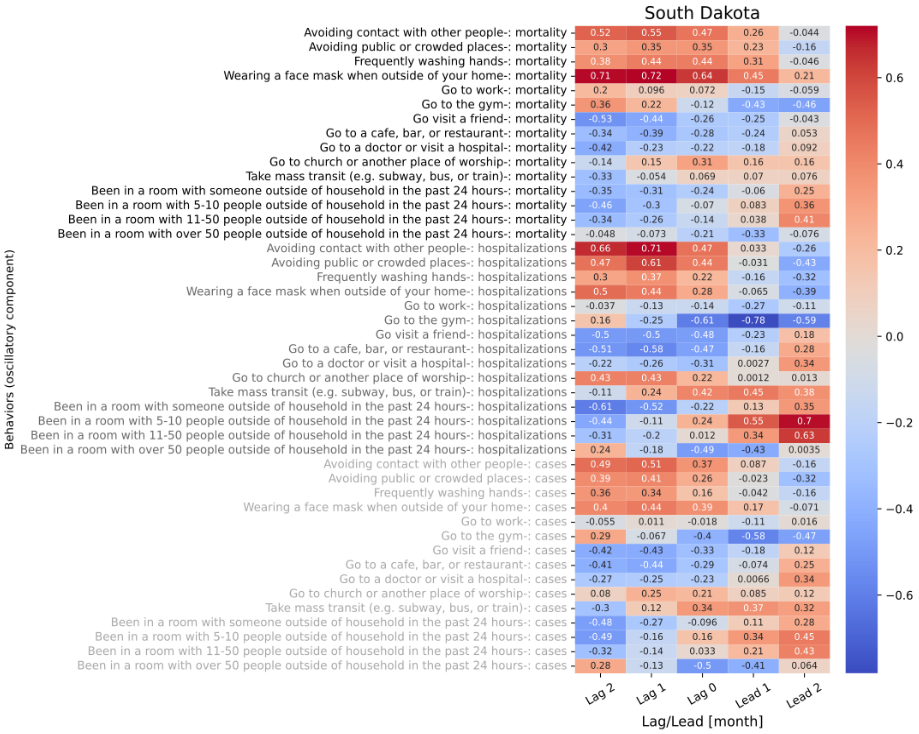

**Figure S8.42 shows the 5 lagged correlations of oscillations in behavior trend with mortality (black), hospitalizations (gray), and cases (light gray) for South Dakota.**

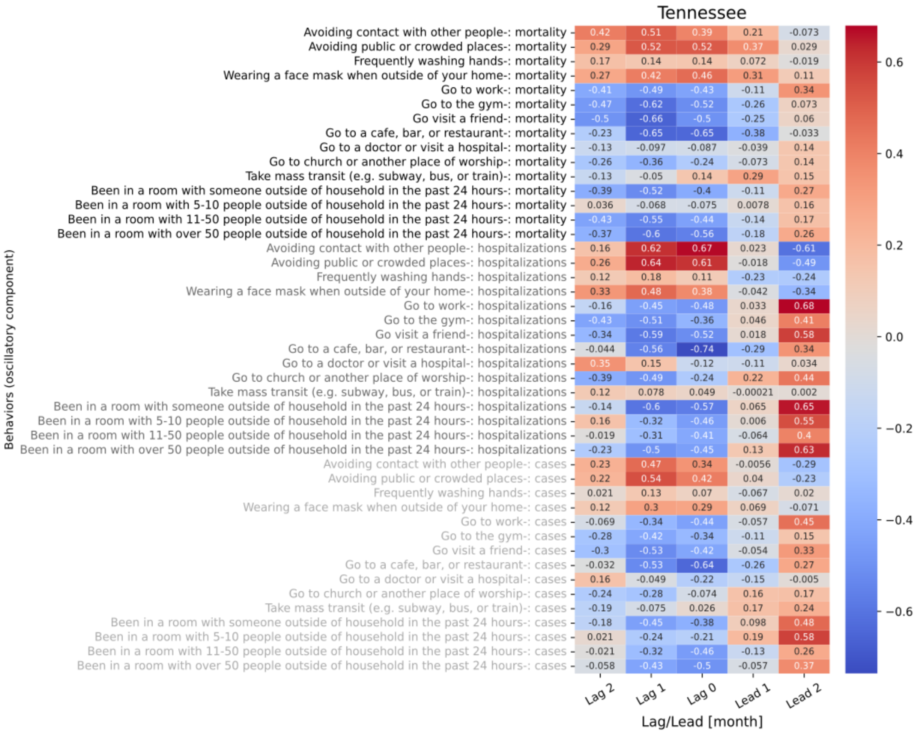

**Figure S8.43 shows the 5 lagged correlations of oscillations in behavior trend with mortality (black), hospitalizations (gray), and cases (light gray) for Tennessee.**

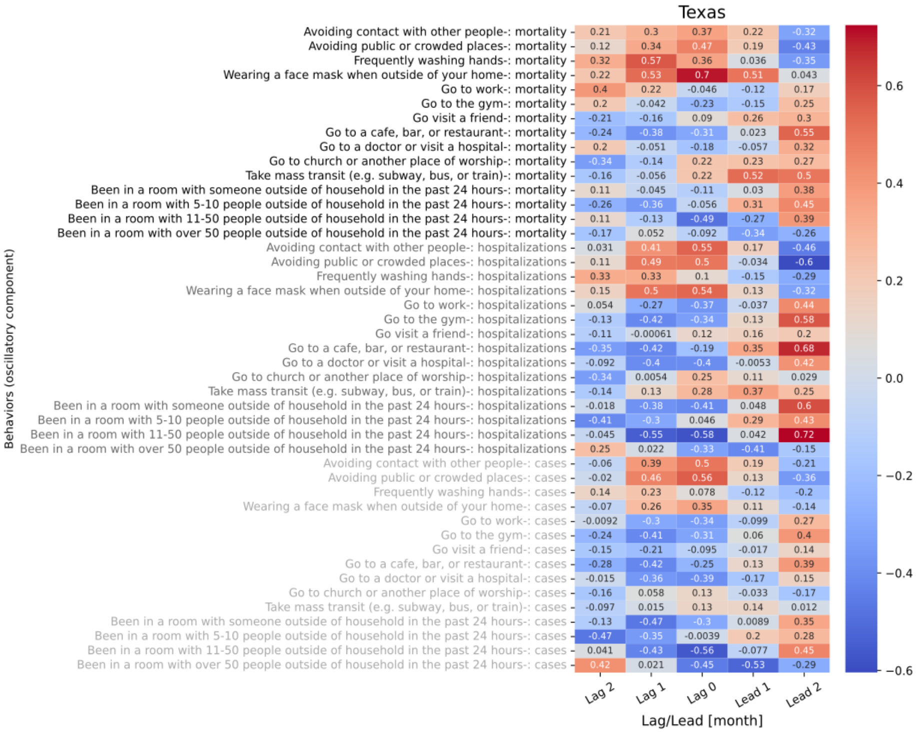

**Figure S8.44 shows the 5 lagged correlations of oscillations in behavior trend with mortality (black), hospitalizations (gray), and cases (light gray) for Texas.**

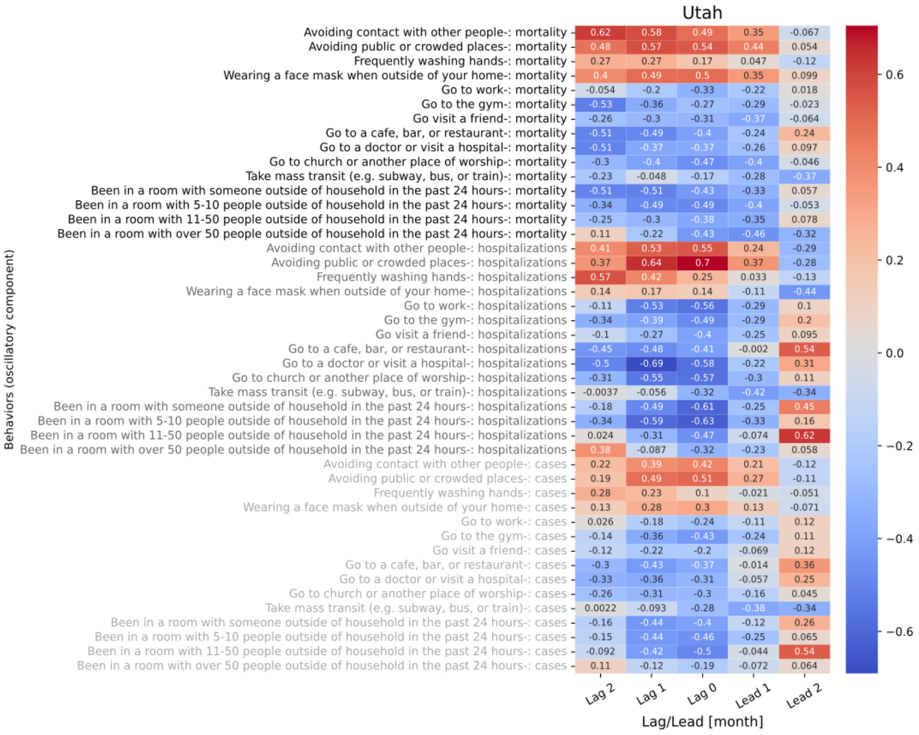

**Figure S8.45 shows the 5 lagged correlations of oscillations in behavior trend with mortality (black), hospitalizations (gray), and cases (light gray) for Utah.**

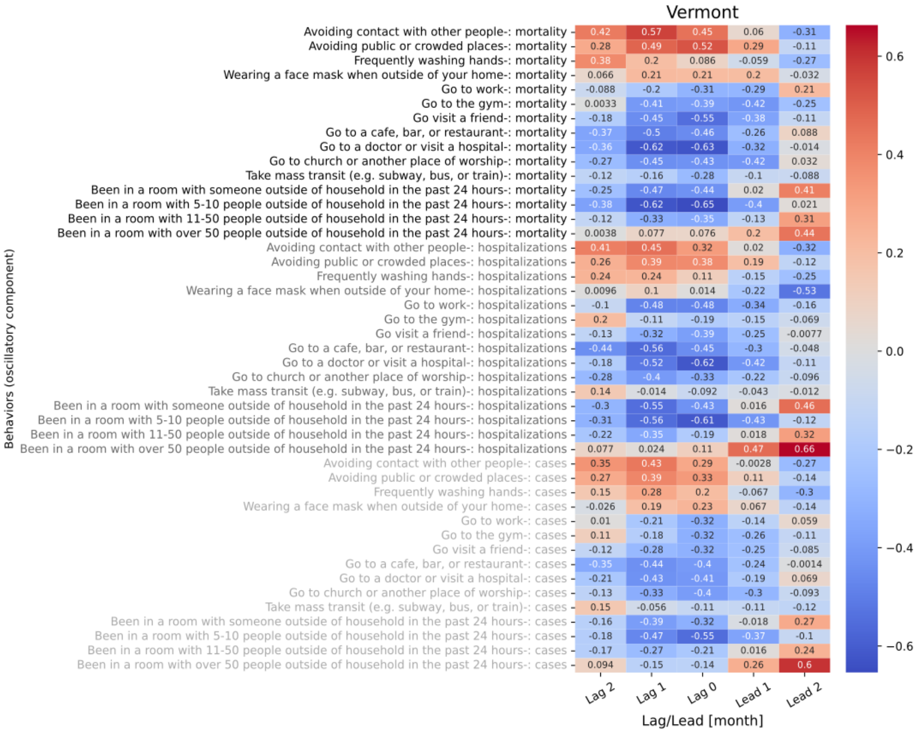

**Figure S8.46 shows the 5 lagged correlations of oscillations in behavior trend with mortality (black), hospitalizations (gray), and cases (light gray) for Vermont.**

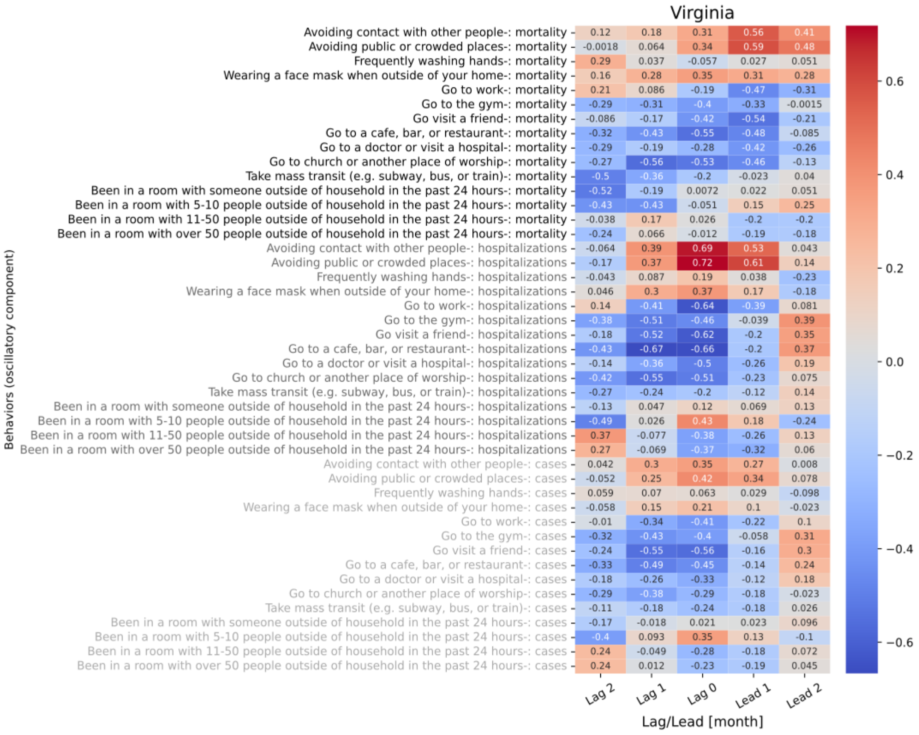

Figure S8.47 shows the 5 lagged correlations of oscillations in behavior trend with mortality (black), hospitalizations (gray), and cases (light gray) for Virginia.

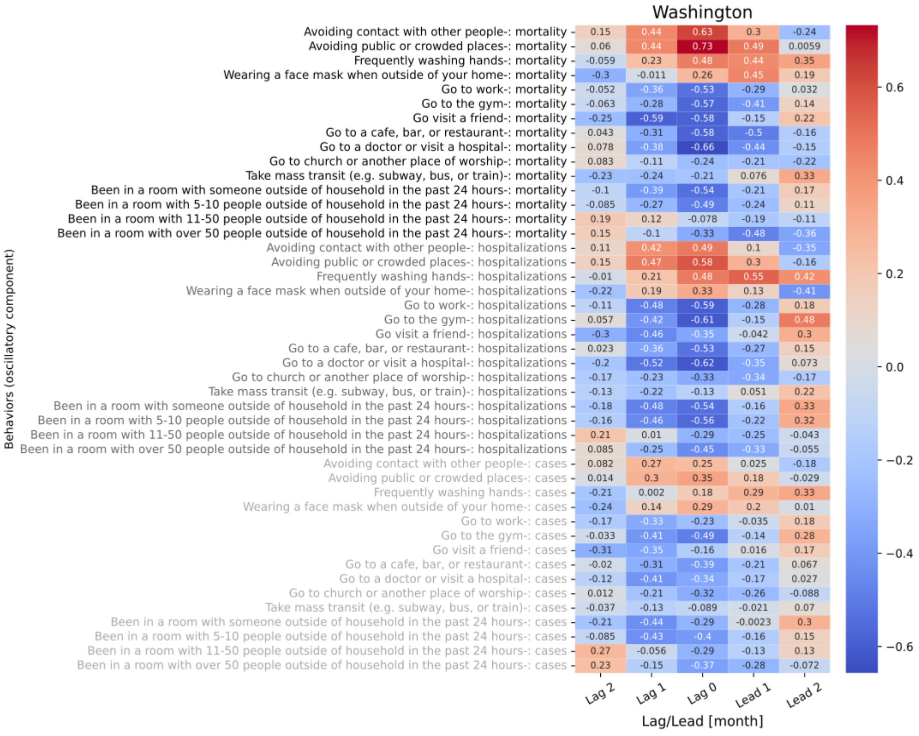

Figure S8.48 shows the 5 lagged correlations of oscillations in behavior trend with mortality (black), hospitalizations (gray), and cases (light gray) for Washington.

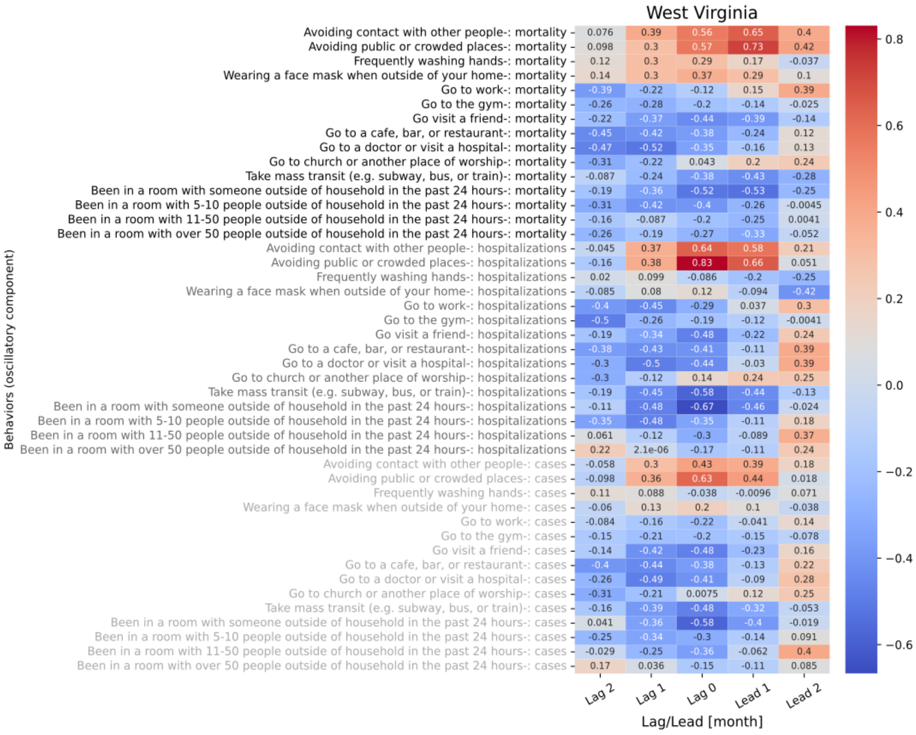

**Figure S8.49 shows the 5 lagged correlations of oscillations in behavior trend with mortality (black), hospitalizations (gray), and cases (light gray) for West Virginia.**

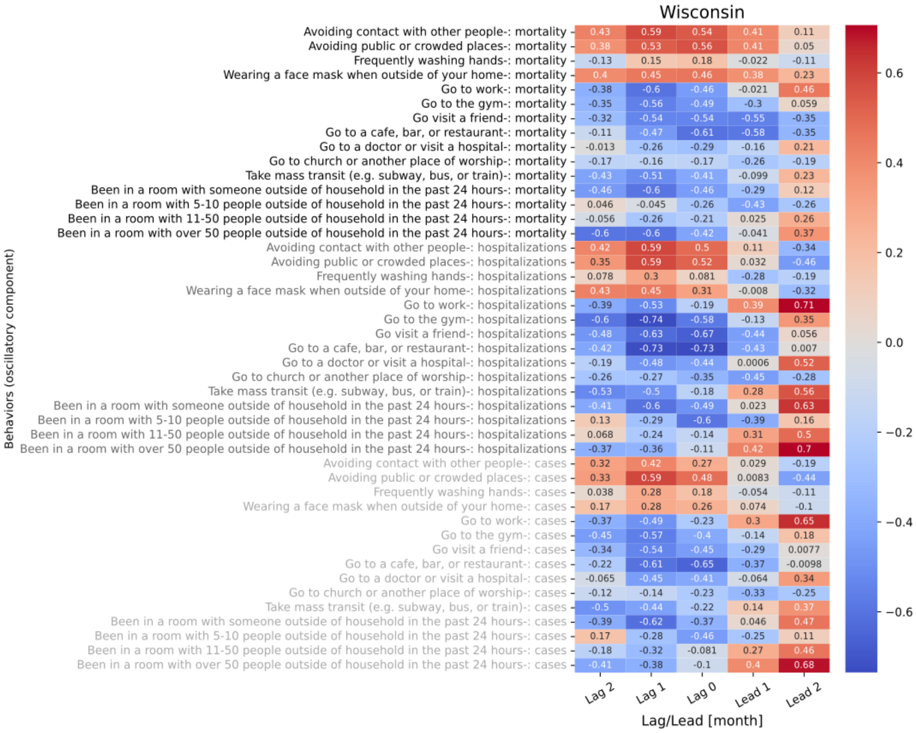

**Figure S8.50 shows the 5 lagged correlations of oscillations in behavior trend with mortality (black), hospitalizations (gray), and cases (light gray) for Wisconsin.**

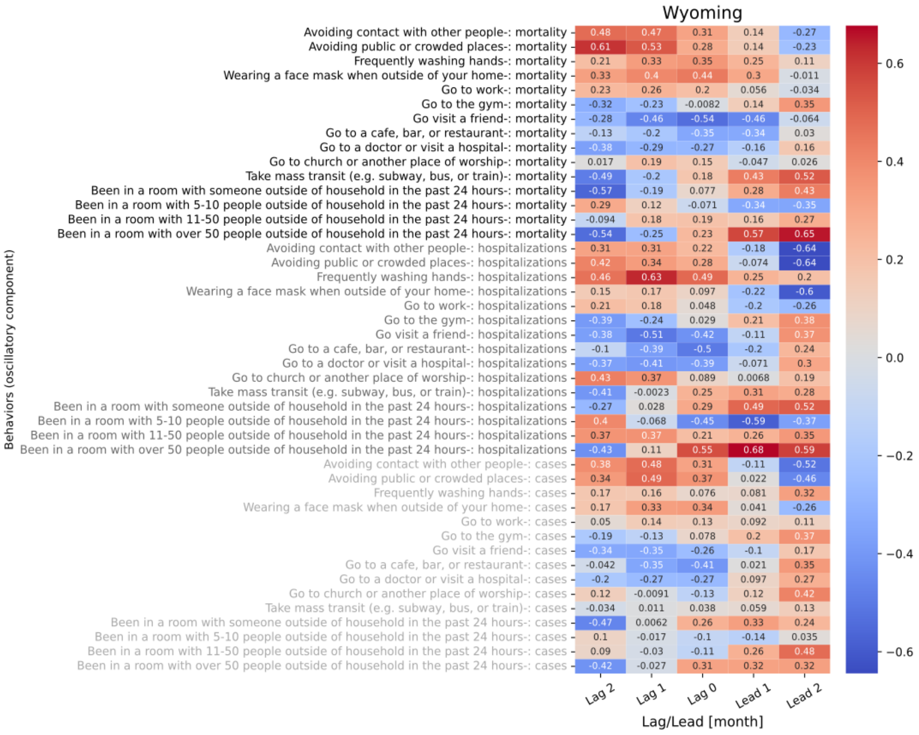

**Figure S8.51 shows the 5 lagged correlations of oscillations in behavior trend with mortality (black), hospitalizations (gray), and cases (light gray) for Wyoming.**

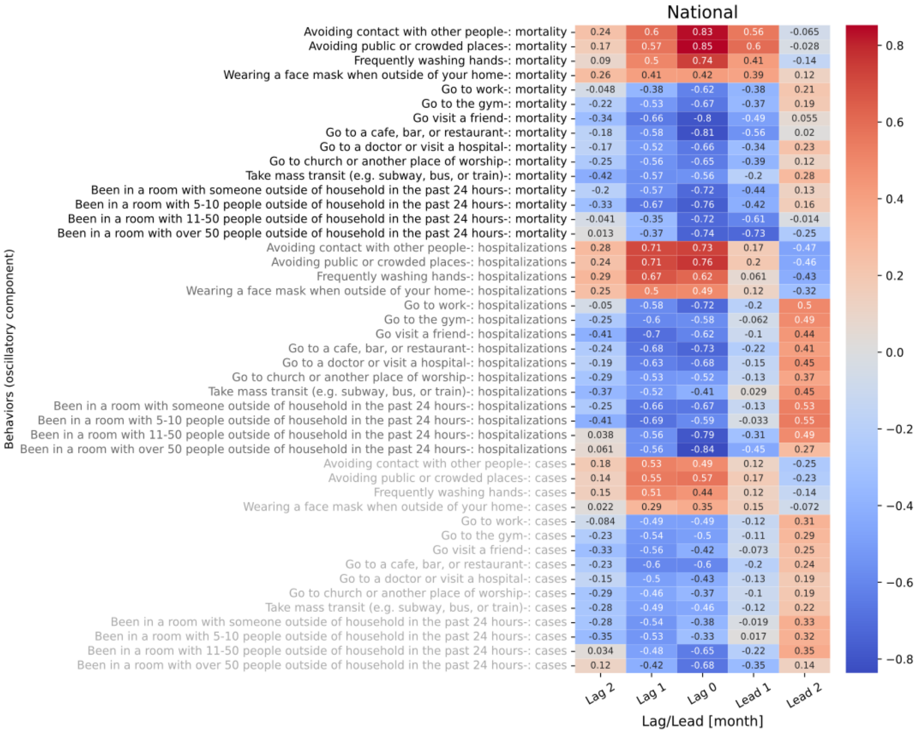

**Figure S8.52 shows the 5 lagged correlations of oscillations in behavior trend with mortality (black), hospitalizations (gray), and cases (light gray) for National.**

### 9 Correlations between oscillations in state-level behavior trends and national-level severity metrics

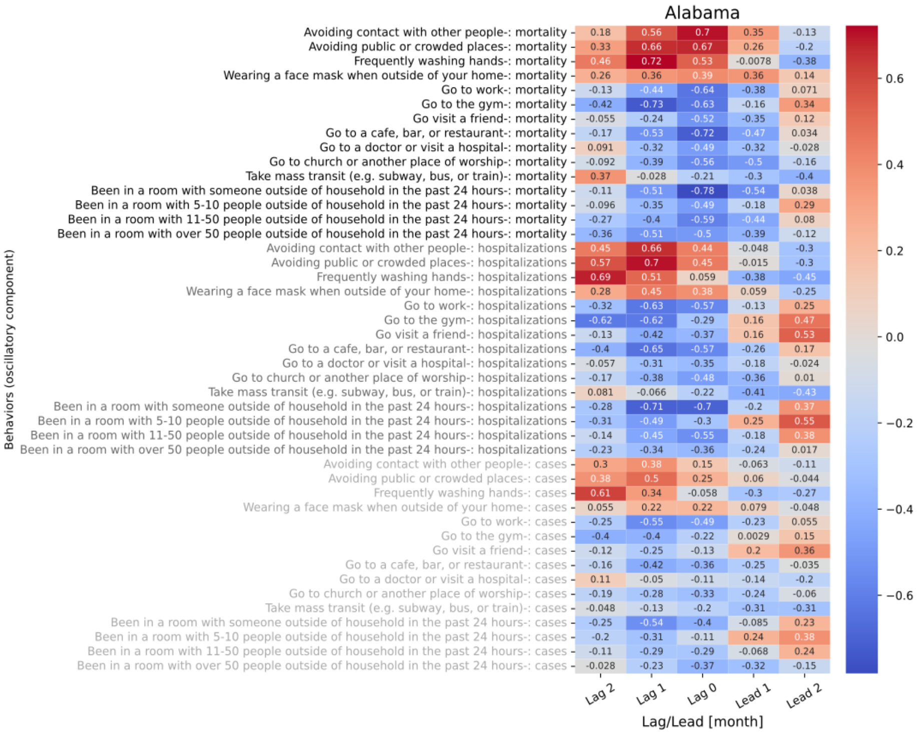

**Figure S9.1 shows the 5 lagged correlations of oscillations in behavior trend for Alabama with national level mortality (black), hospitalizations (gray), and cases (light gray).**

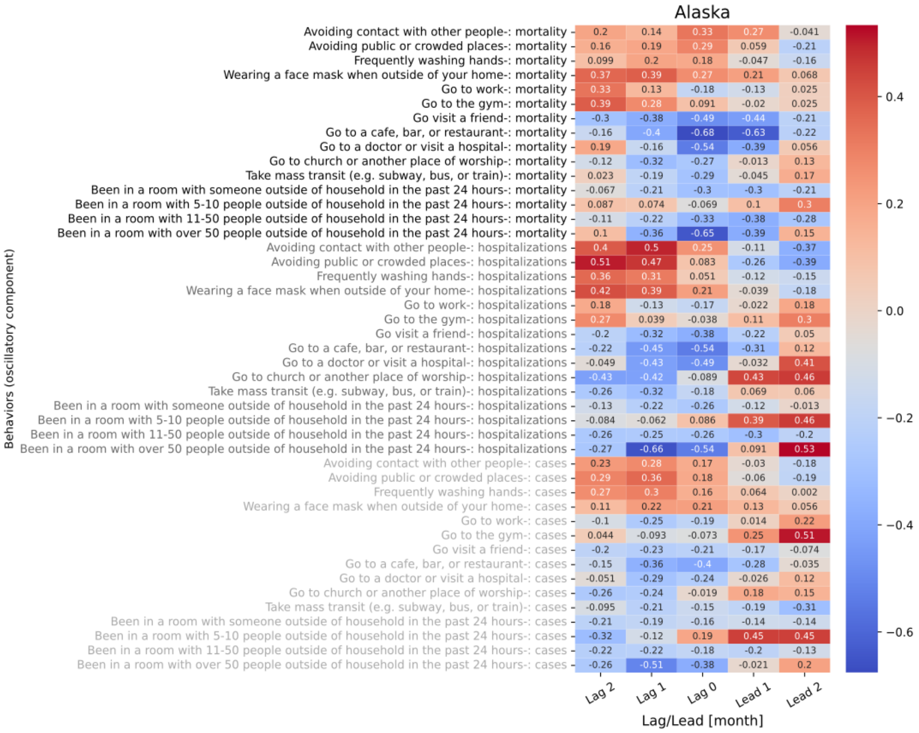

**Figure S9.2 shows the 5 lagged correlations of oscillations in behavior trend for Alaska with national level mortality (black), hospitalizations (gray), and cases (light gray).**

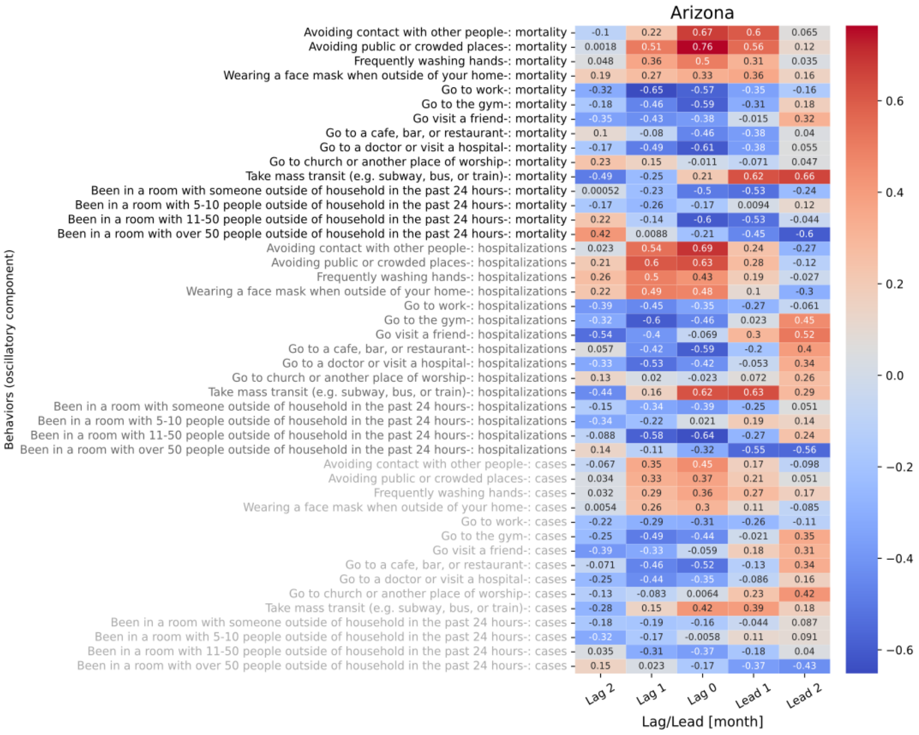

**Figure S9.3 shows the 5 lagged correlations of oscillations in behavior trend for Arizona with national level mortality (black), hospitalizations (gray), and cases (light gray).**

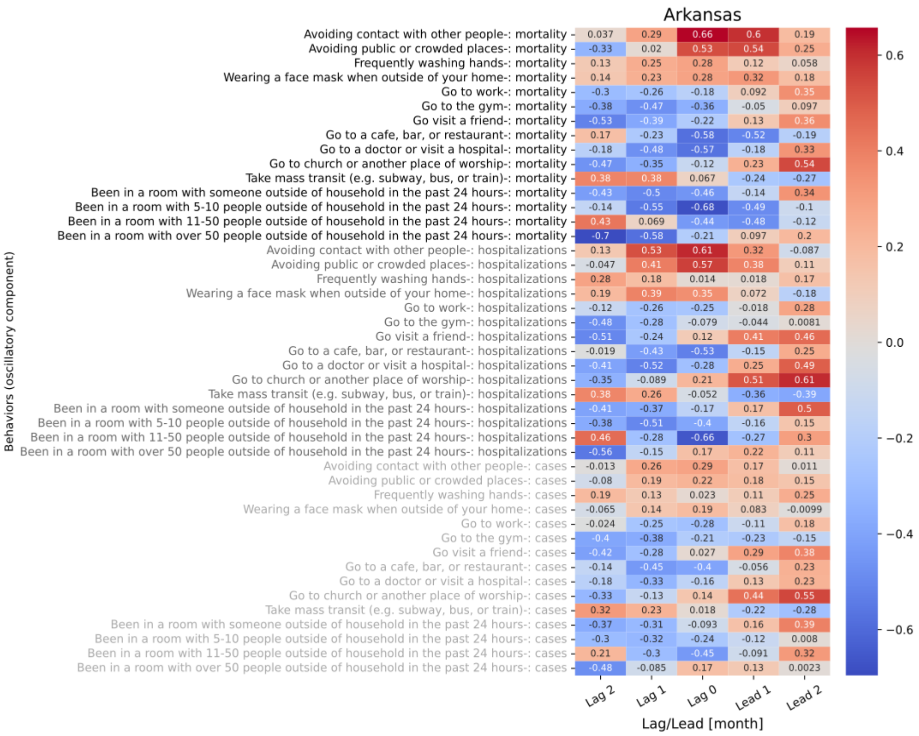

**Figure S9.4 shows the 5 lagged correlations of oscillations in behavior trend for Arkansas with national level mortality (black), hospitalizations (gray), and cases (light gray).**

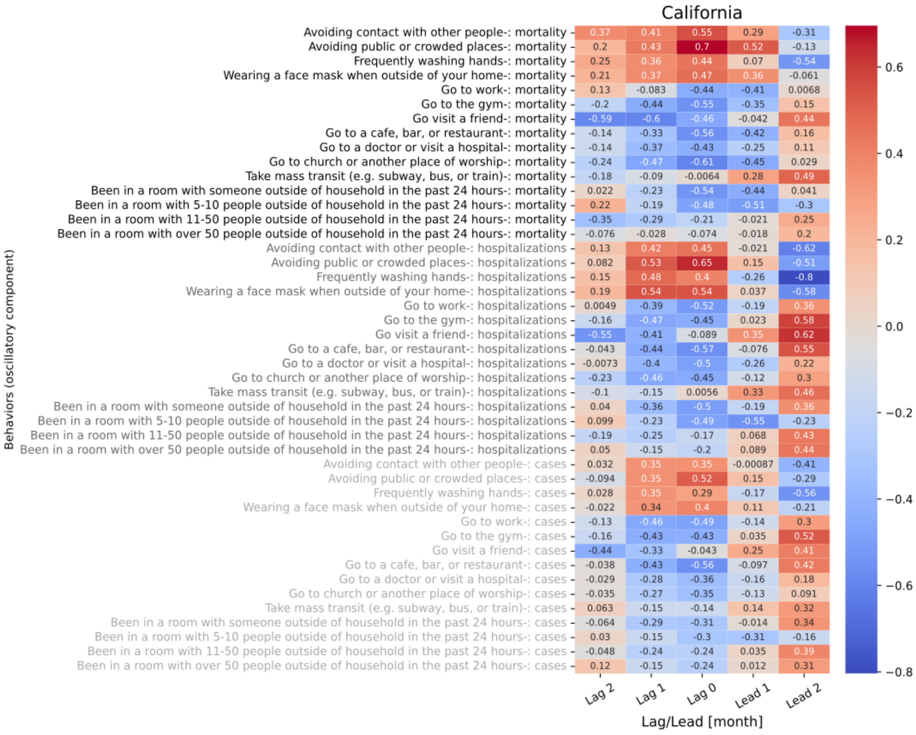

**Figure S9.5 shows the 5 lagged correlations of oscillations in behavior trend for California with national level mortality (black), hospitalizations (gray), and cases (light gray).**

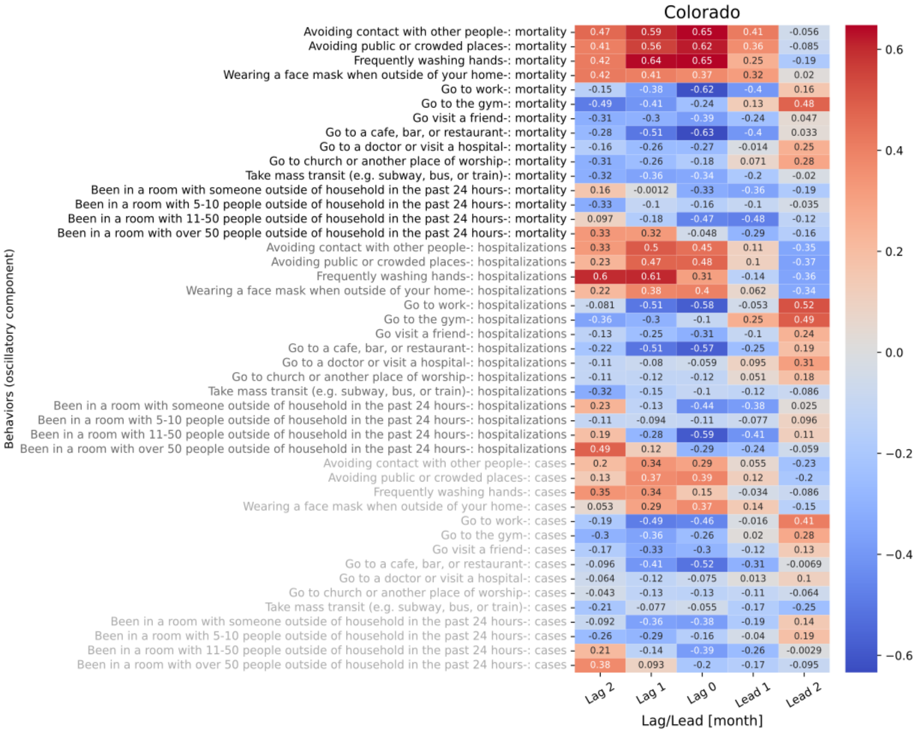

**Figure S9.6 shows the 5 lagged correlations of oscillations in behavior trend for Colorado with national level mortality (black), hospitalizations (gray), and cases (light gray).**

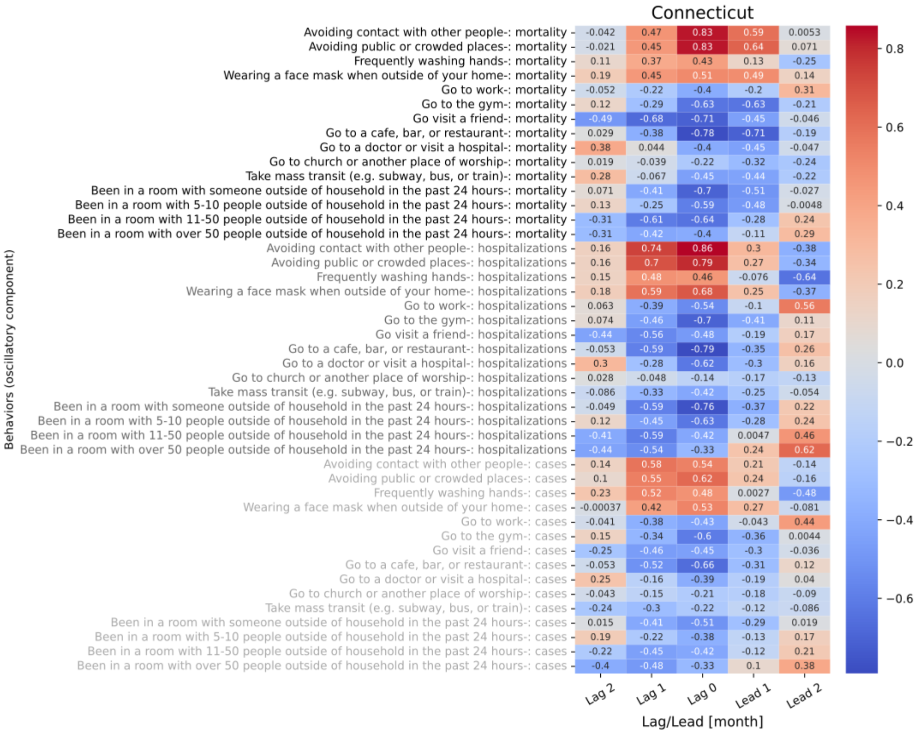

**Figure S9.7 shows the 5 lagged correlations of oscillations in behavior trend for Connecticut with national level mortality (black), hospitalizations (gray), and cases (light gray).**

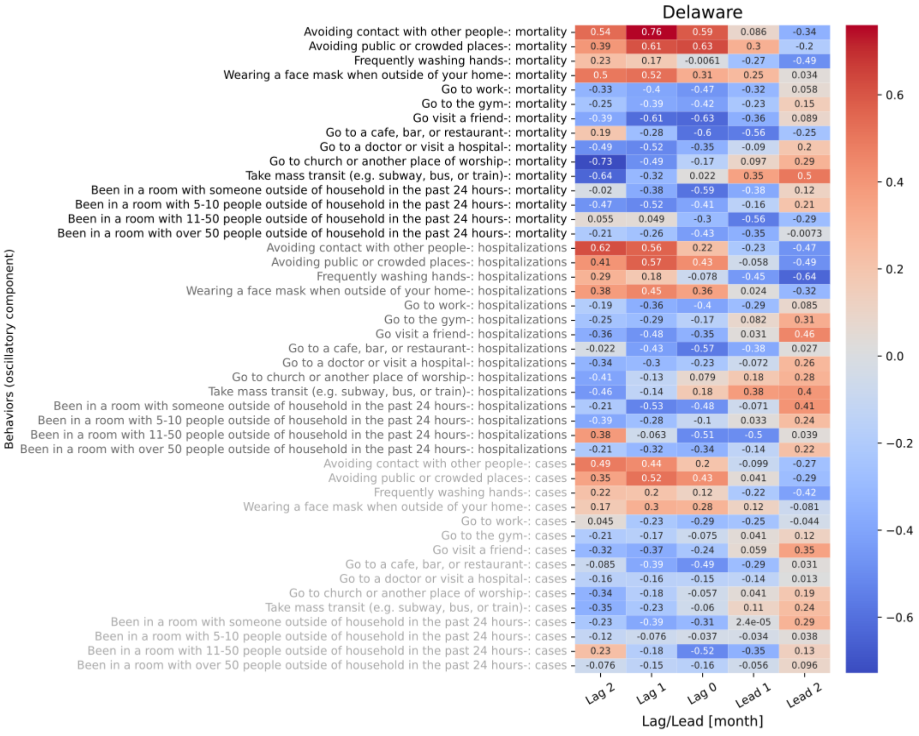

**Figure S9.8 shows the 5 lagged correlations of oscillations in behavior trend for Delaware with national level mortality (black), hospitalizations (gray), and cases (light gray).**

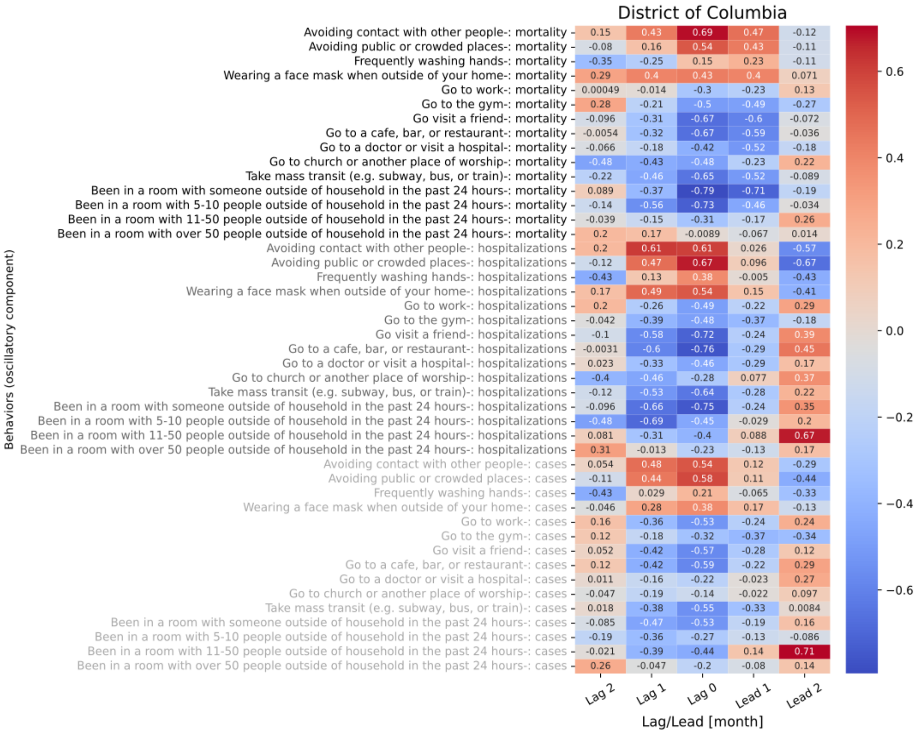

Figure S9.9 shows the 5 lagged correlations of oscillations in behavior trend for District of Columbia with national level mortality (black), hospitalizations (gray), and cases (light gray).

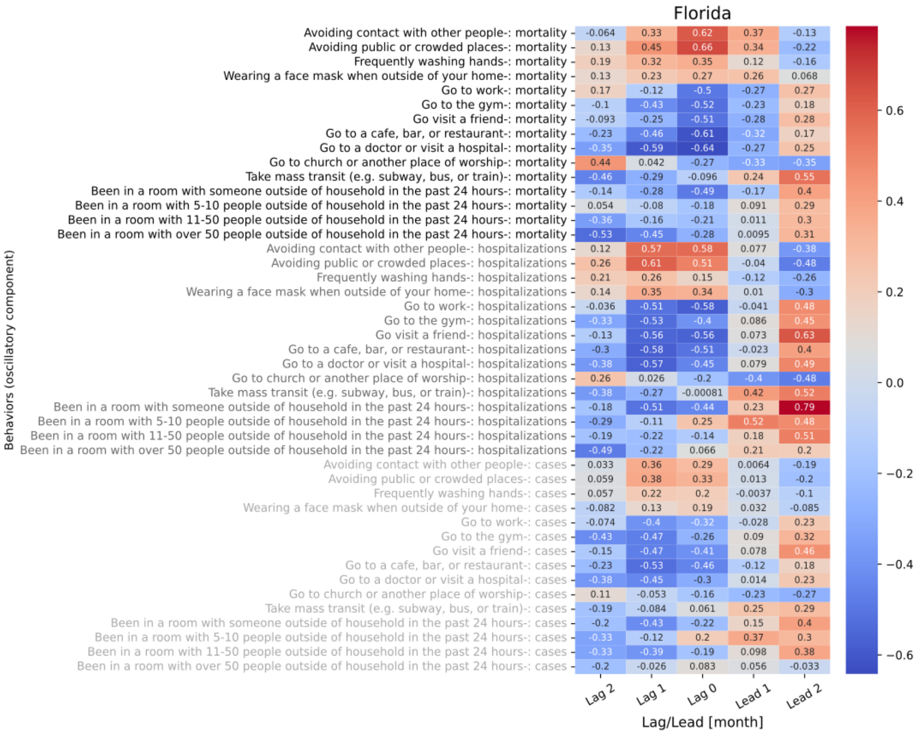

Figure S9.10 shows the 5 lagged correlations of oscillations in behavior trend for Florida with national level mortality (black), hospitalizations (gray), and cases (light gray).

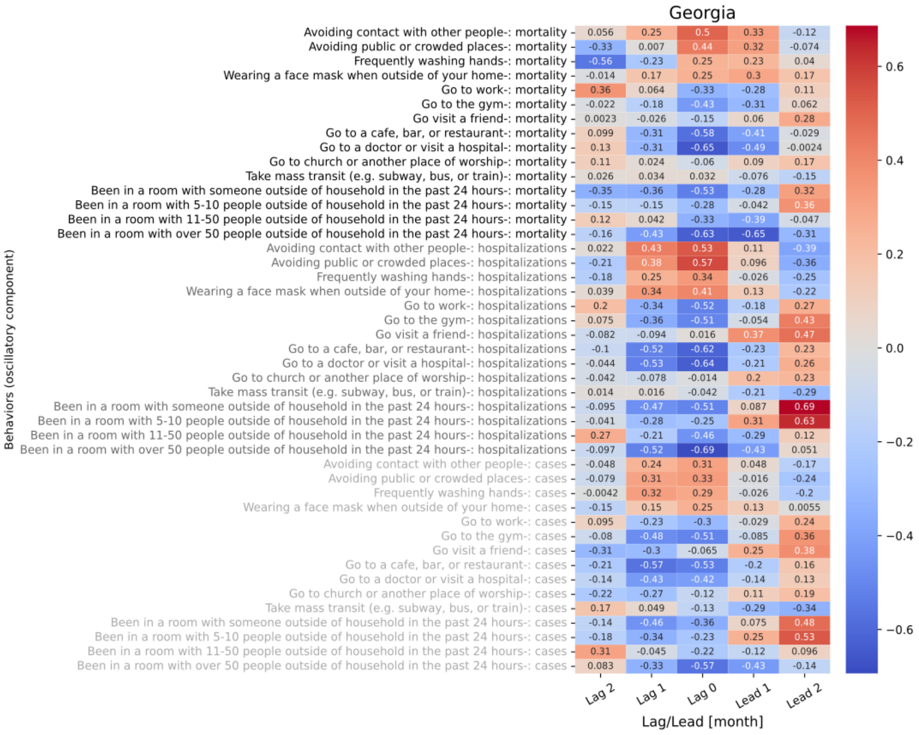

**Figure S9.11 shows the 5 lagged correlations of oscillations in behavior trend for Georgia with national level mortality (black), hospitalizations (gray), and cases (light gray).**

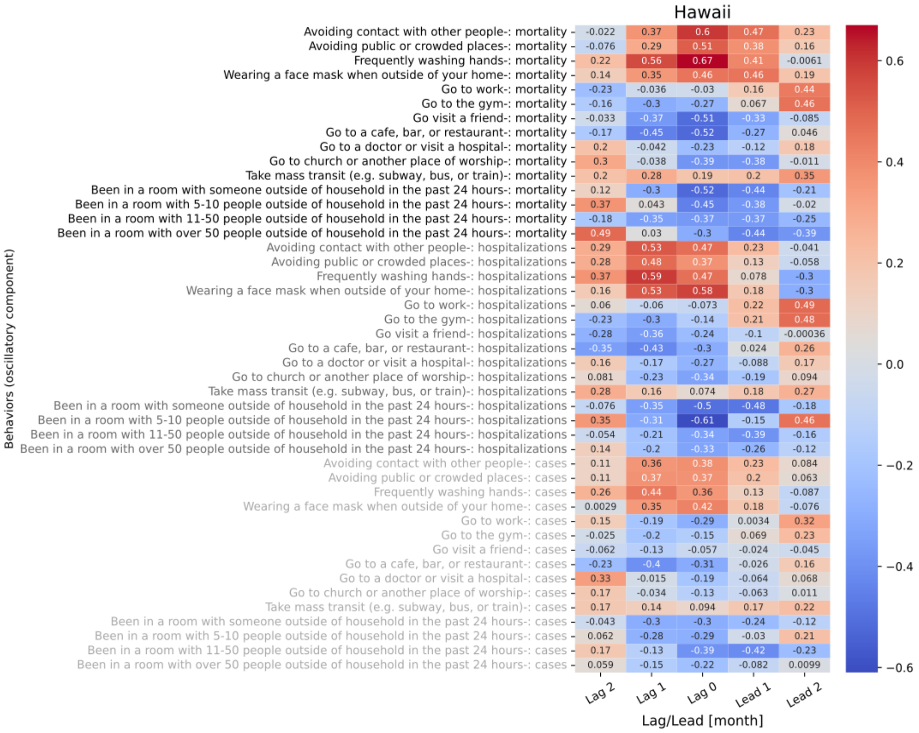

**Figure S9.12 shows the 5 lagged correlations of oscillations in behavior trend for Hawaii with national level mortality (black), hospitalizations (gray), and cases (light gray).**

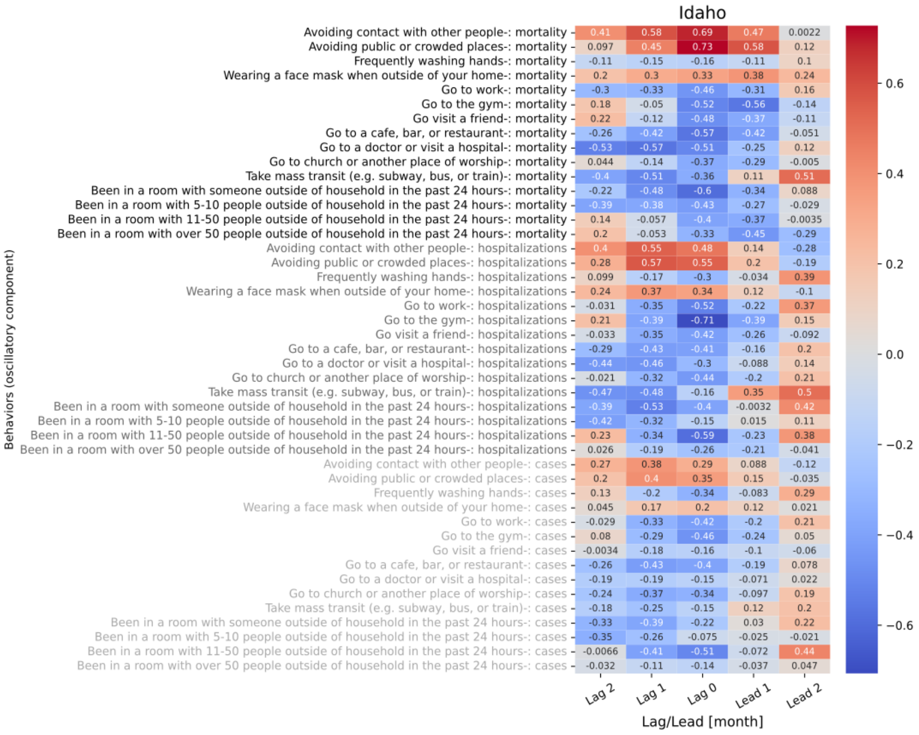

**Figure S9.13 shows the 5 lagged correlations of oscillations in behavior trend for Idaho with national level mortality (black), hospitalizations (gray), and cases (light gray).**

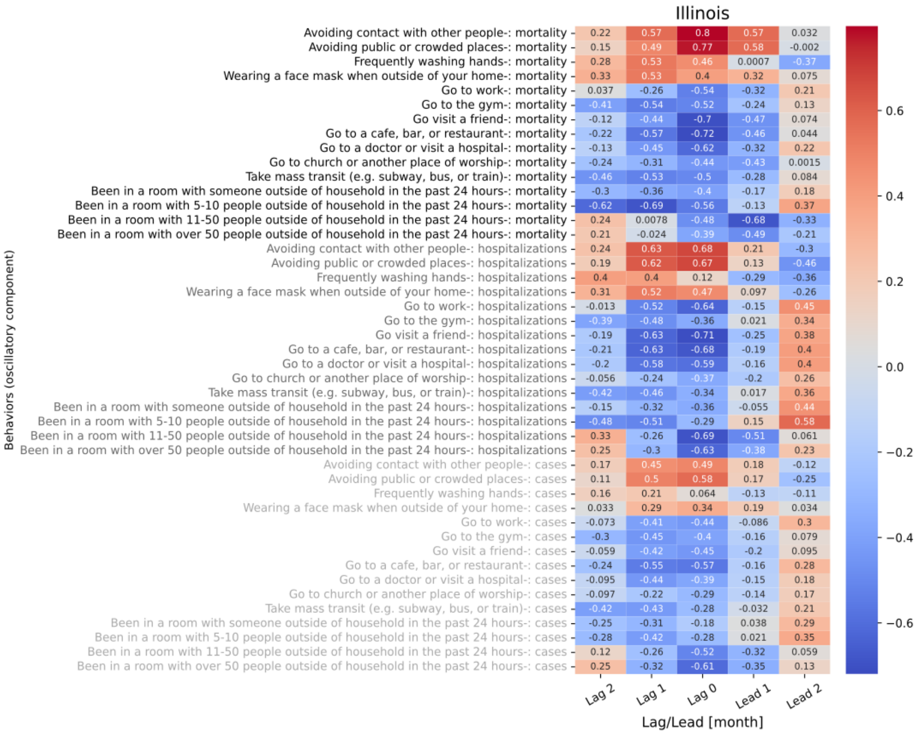

**Figure S9.14 shows the 5 lagged correlations of oscillations in behavior trend for Illinois with national level mortality (black), hospitalizations (gray), and cases (light gray).**

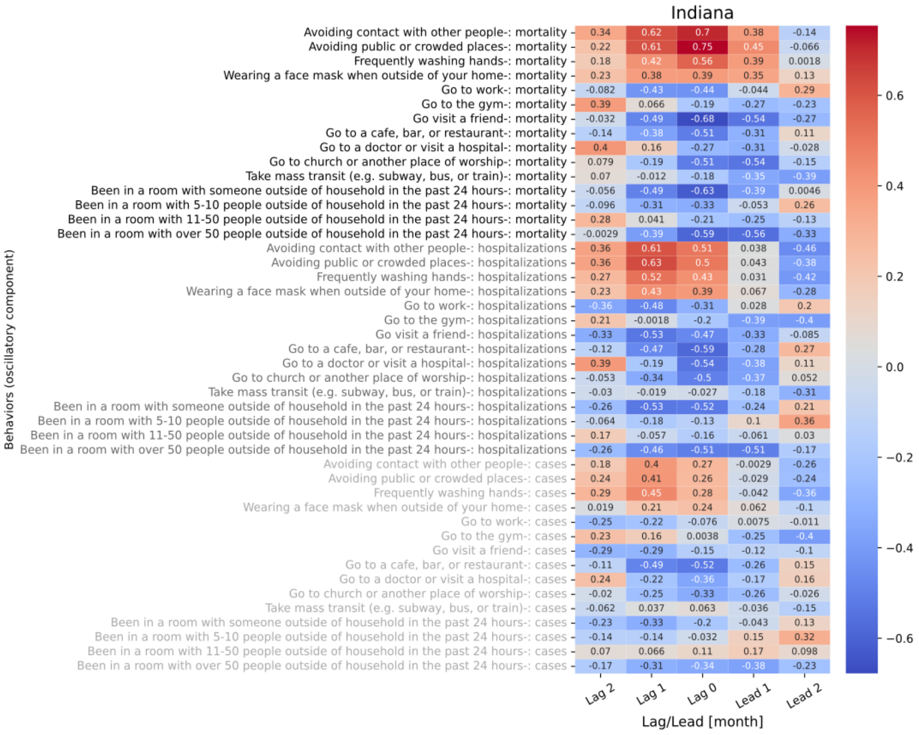

**Figure S9.15 shows the 5 lagged correlations of oscillations in behavior trend for Indiana with national level mortality (black), hospitalizations (gray), and cases (light gray).**

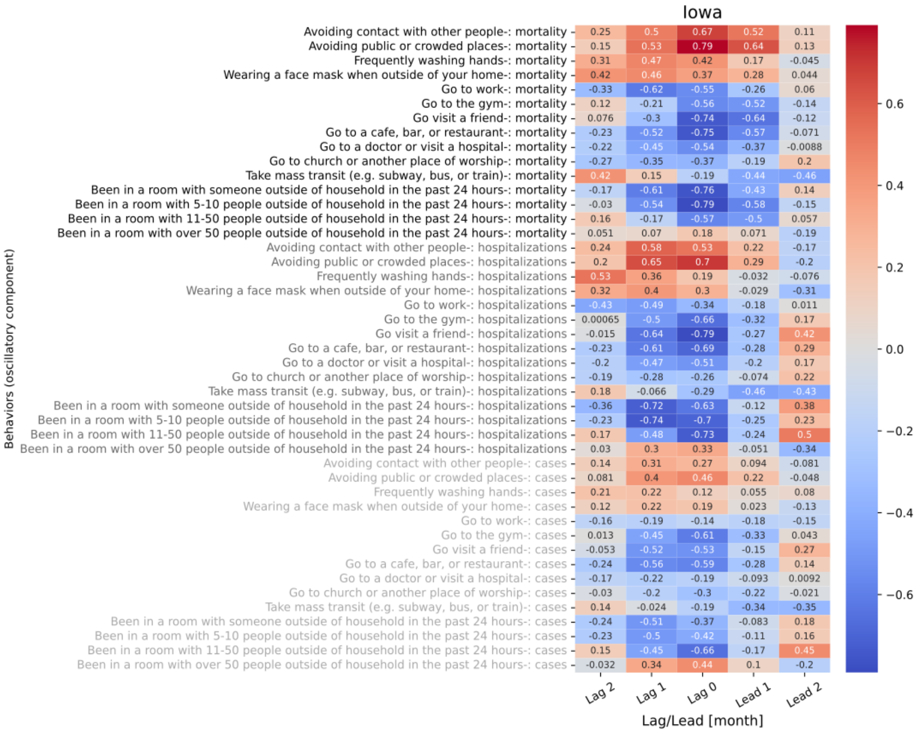

**Figure S9.16 shows the 5 lagged correlations of oscillations in behavior trend for Iowa with national level mortality (black), hospitalizations (gray), and cases (light gray).**

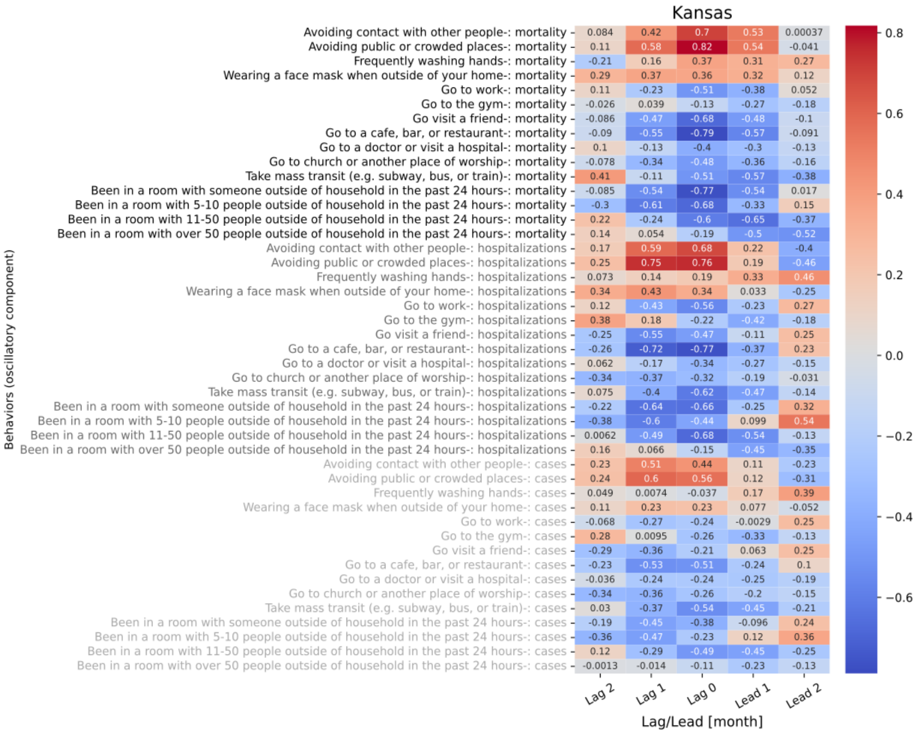

**Figure S9.17 shows the 5 lagged correlations of oscillations in behavior trend for Kansas with national level mortality (black), hospitalizations (gray), and cases (light gray).**

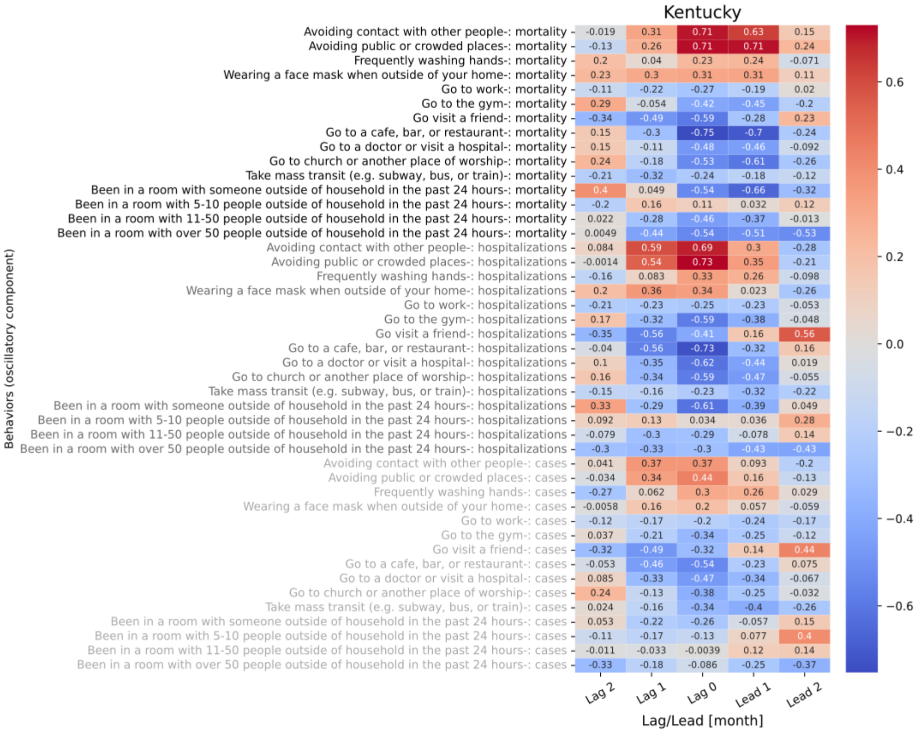

**Figure S9.18 shows the 5 lagged correlations of oscillations in behavior trend for Kentucky with national level mortality (black), hospitalizations (gray), and cases (light gray).**

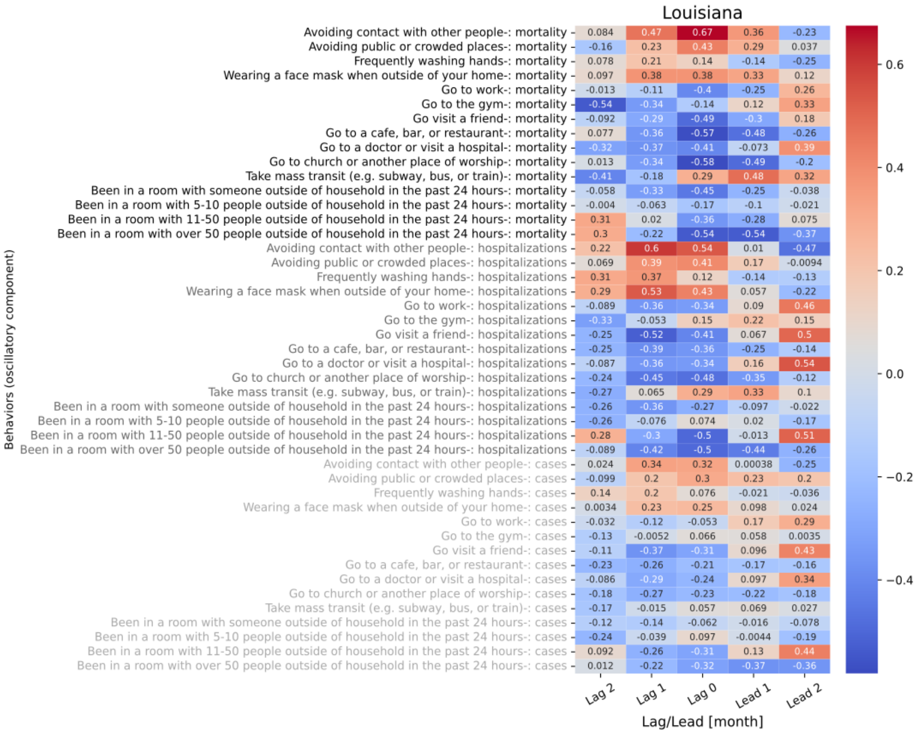

**Figure S9.19 shows the 5 lagged correlations of oscillations in behavior trend for Louisiana with national level mortality (black), hospitalizations (gray), and cases (light gray).**

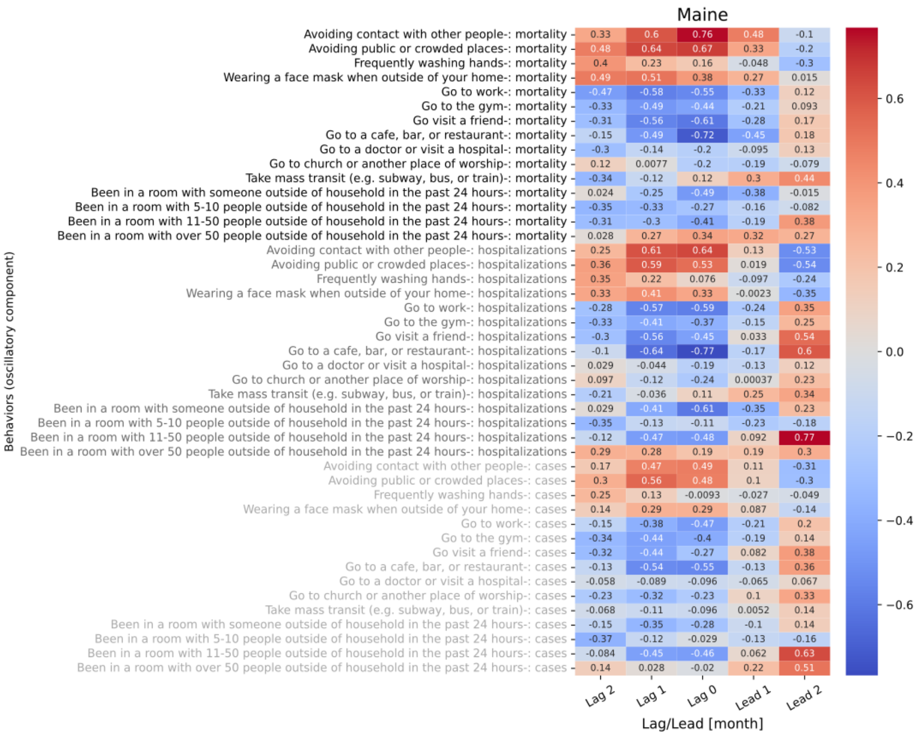

**Figure S9.20 shows the 5 lagged correlations of oscillations in behavior trend for Maine with national level mortality (black), hospitalizations (gray), and cases (light gray).**

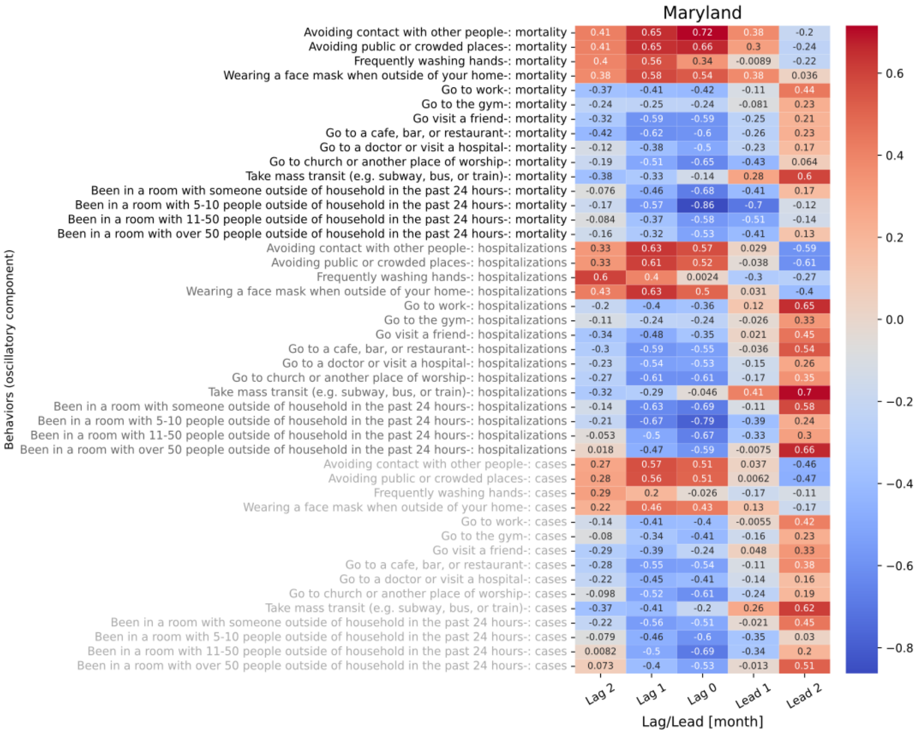

Figure S9.21 shows the 5 lagged correlations of oscillations in behavior trend for Maryland with national level mortality (black), hospitalizations (gray), and cases (light gray).

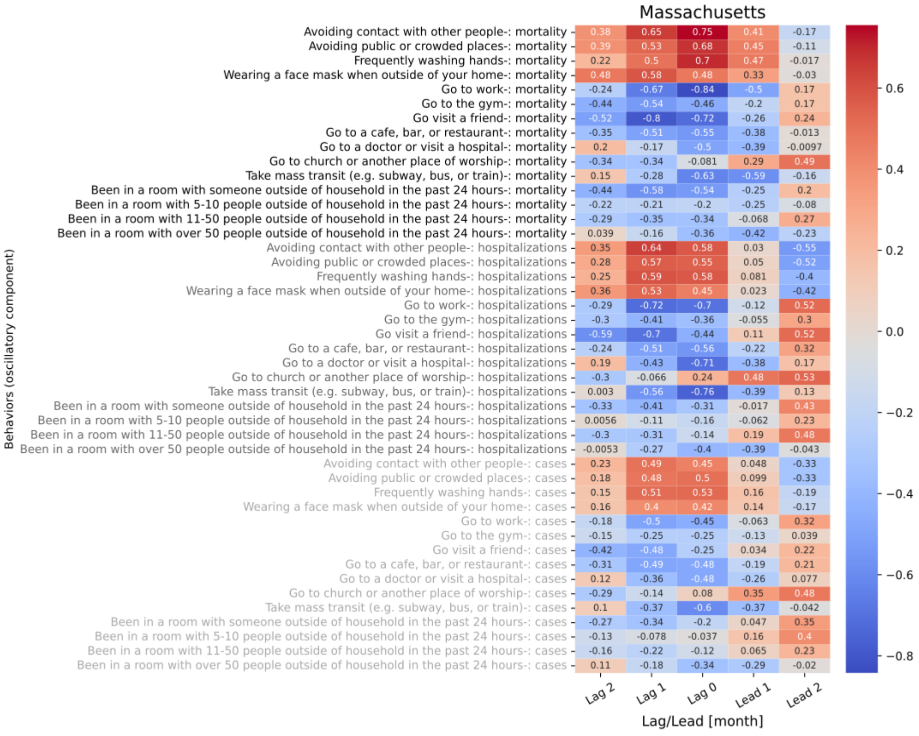

Figure S9.22 shows the 5 lagged correlations of oscillations in behavior trend for Massachusetts with national level mortality (black), hospitalizations (gray), and cases (light gray).

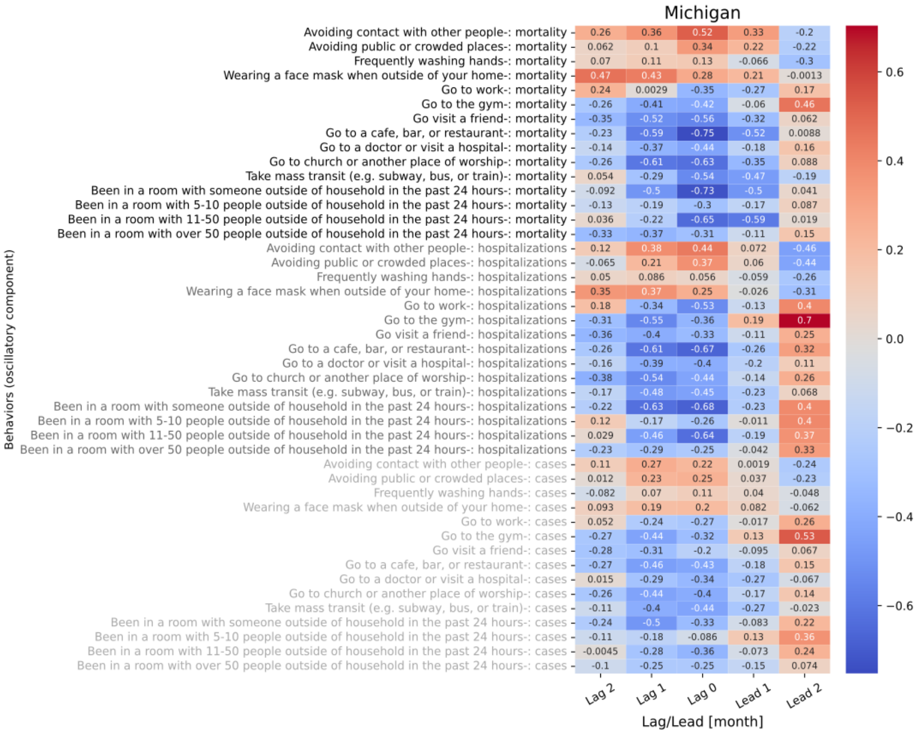

**Figure S9.23 shows the 5 lagged correlations of oscillations in behavior trend for Michigan with national level mortality (black), hospitalizations (gray), and cases (light gray).**

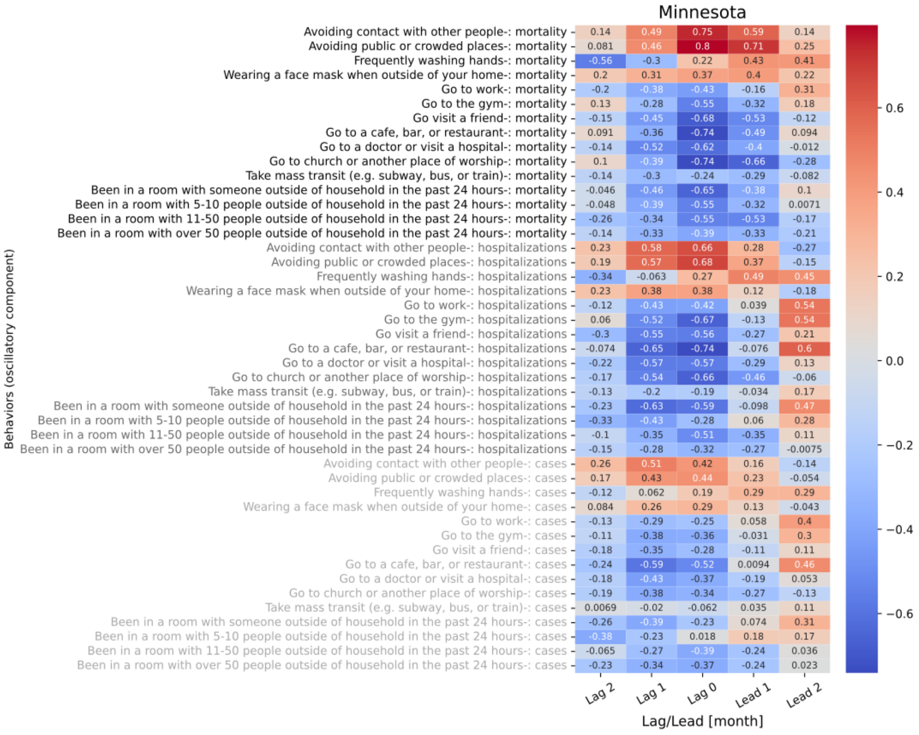

**Figure S9.24 shows the 5 lagged correlations of oscillations in behavior trend for Minnesota with national level mortality (black), hospitalizations (gray), and cases (light gray).**

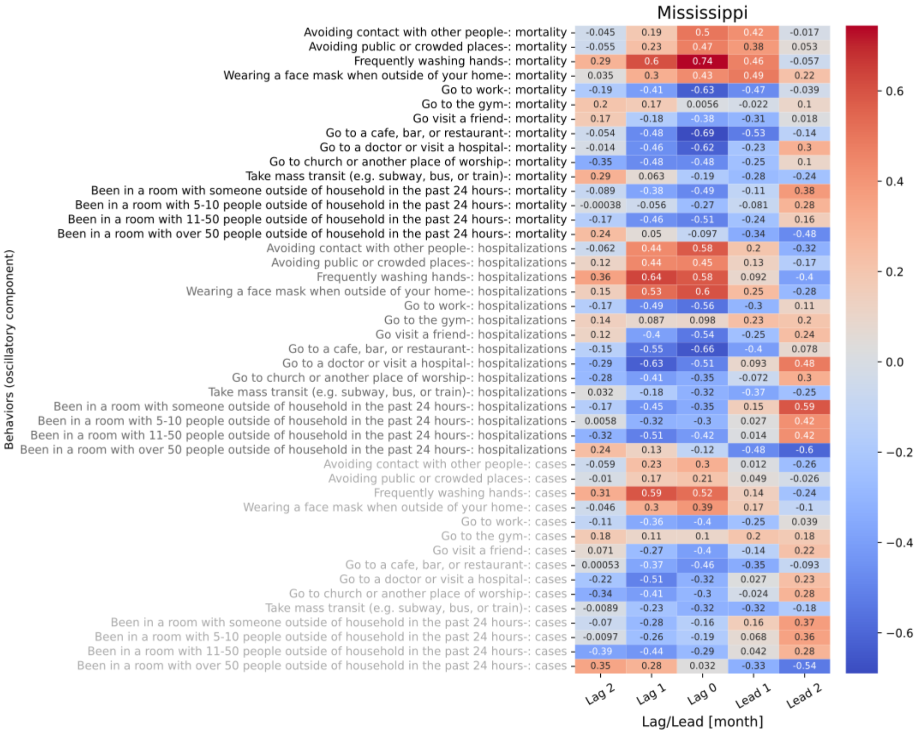

**Figure S9.25 shows the 5 lagged correlations of oscillations in behavior trend for Mississippi with national level mortality (black), hospitalizations (gray), and cases (light gray).**

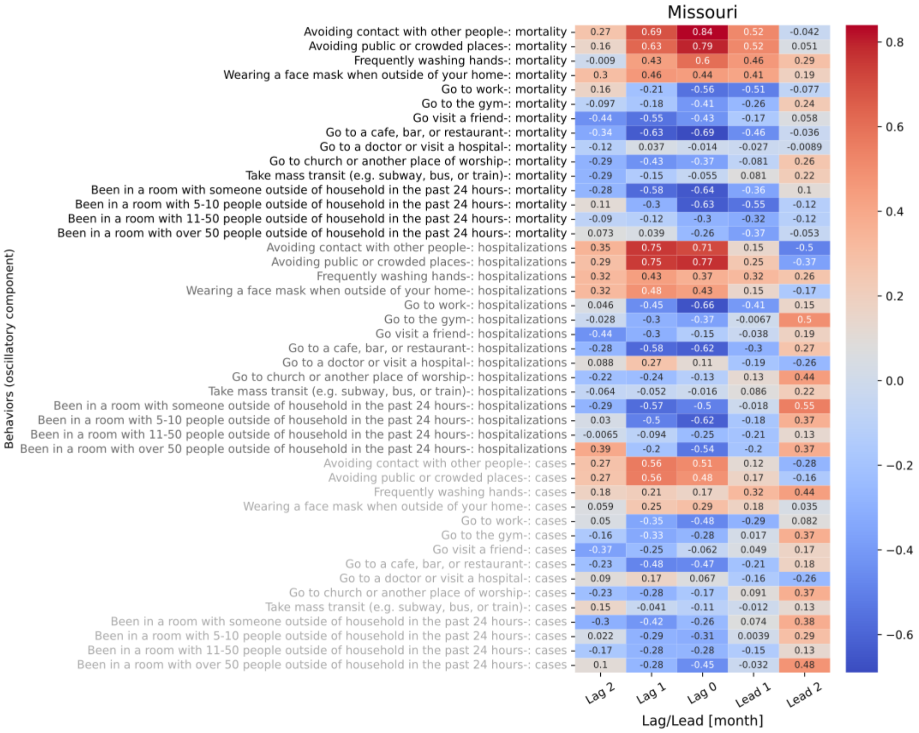

**Figure S9.26 shows the 5 lagged correlations of oscillations in behavior trend for Missouri with national level mortality (black), hospitalizations (gray), and cases (light gray).**

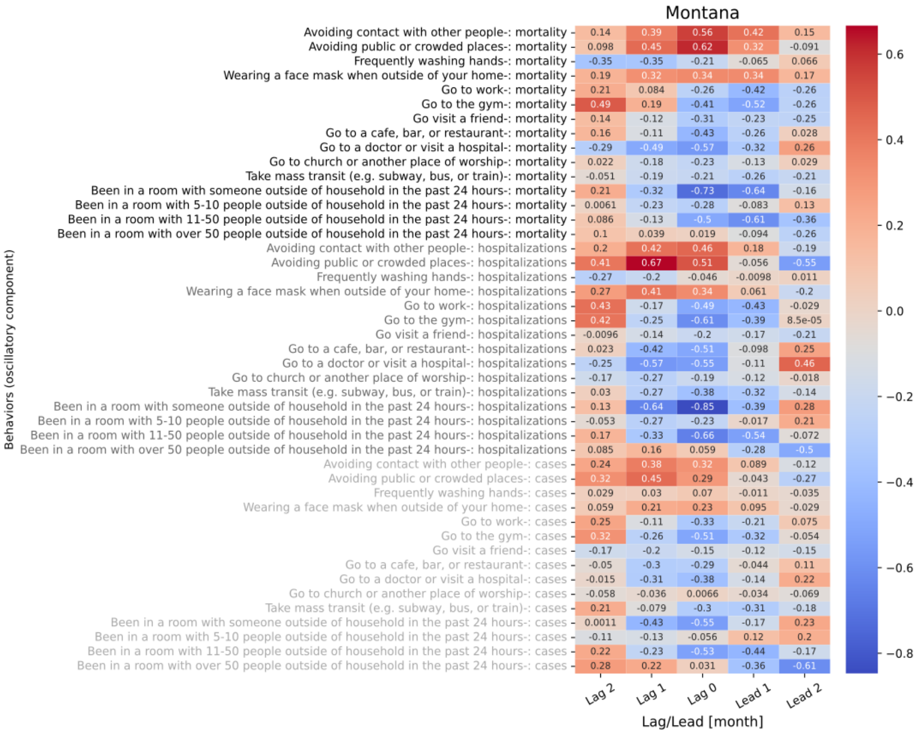

**Figure S9.27 shows the 5 lagged correlations of oscillations in behavior trend for Montana with national level mortality (black), hospitalizations (gray), and cases (light gray).**

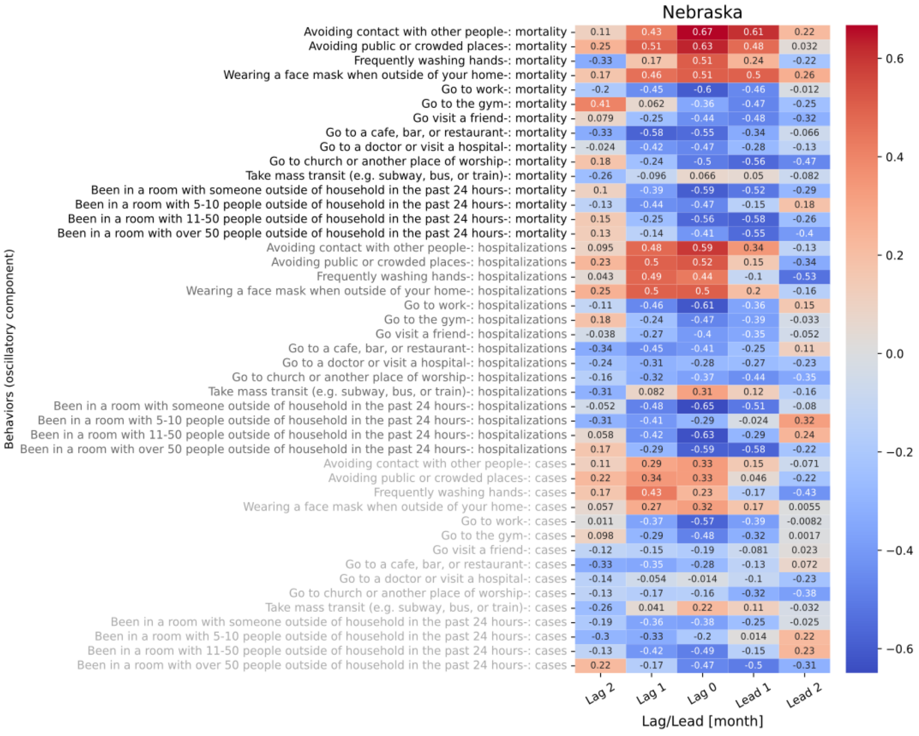

**Figure S9.28 shows the 5 lagged correlations of oscillations in behavior trend for Nebraska with national level mortality (black), hospitalizations (gray), and cases (light gray).**

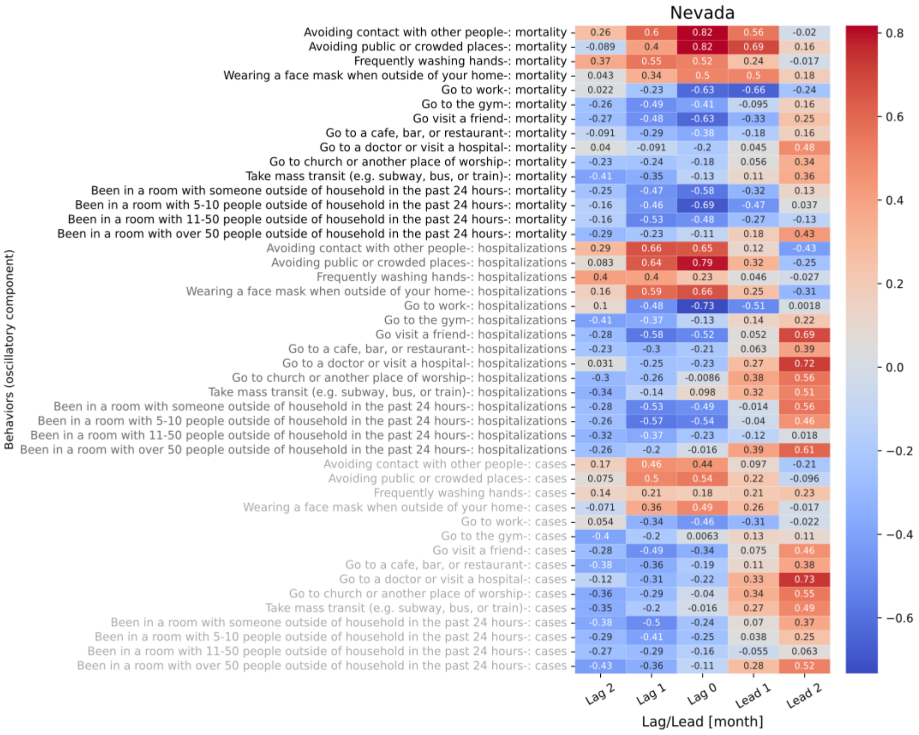

**Figure S9.29 shows the 5 lagged correlations of oscillations in behavior trend for Nevada with national level mortality (black), hospitalizations (gray), and cases (light gray).**

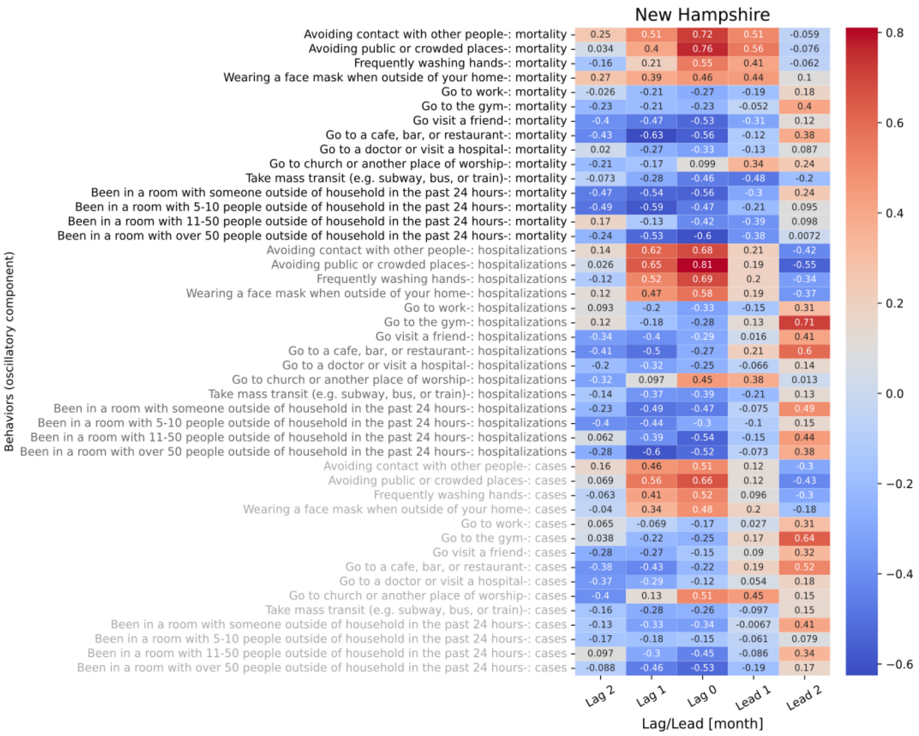

**Figure S9.30 shows the 5 lagged correlations of oscillations in behavior trend for New Hampshire with national level mortality (black), hospitalizations (gray), and cases (light gray).**

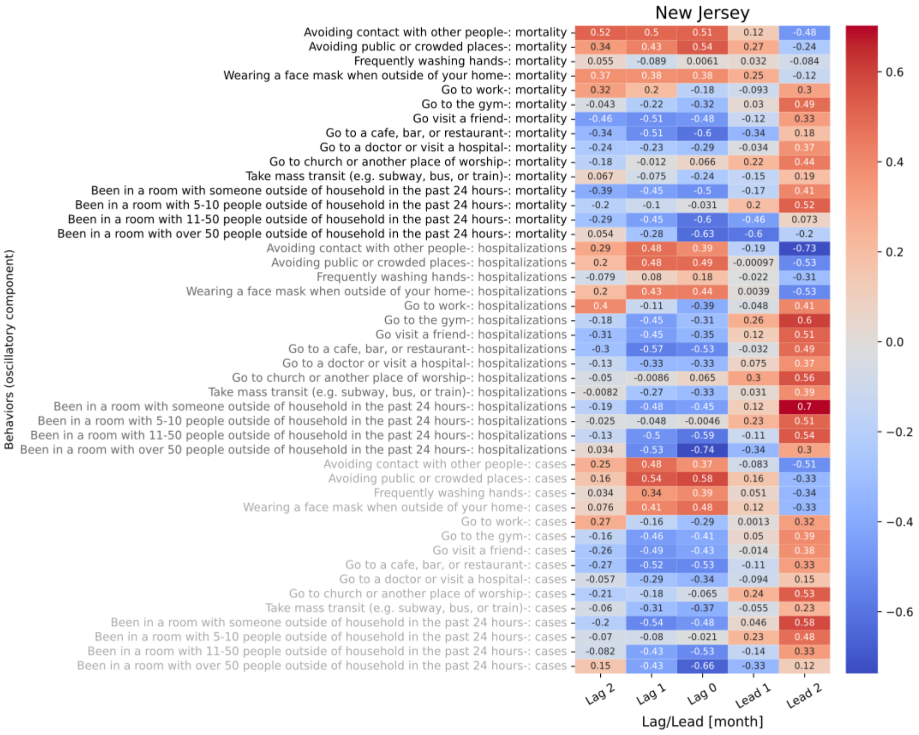

Figure S9.31 shows the 5 lagged correlations of oscillations in behavior trend for New Jersey with national level mortality (black), hospitalizations (gray), and cases (light gray).

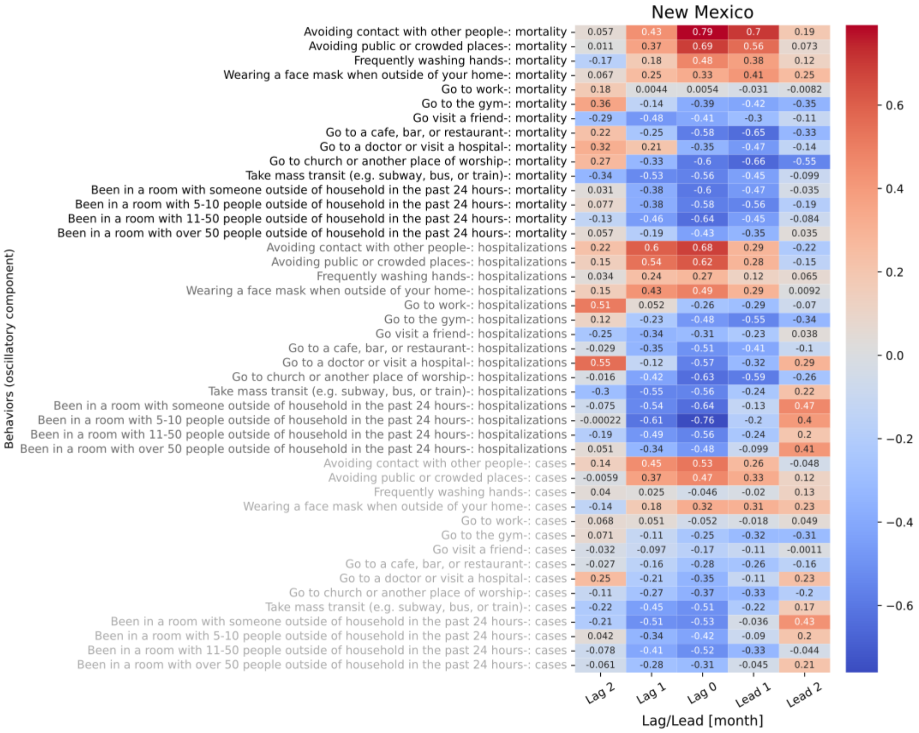

Figure S9.32 shows the 5 lagged correlations of oscillations in behavior trend for New Mexico with national level mortality (black), hospitalizations (gray), and cases (light gray).

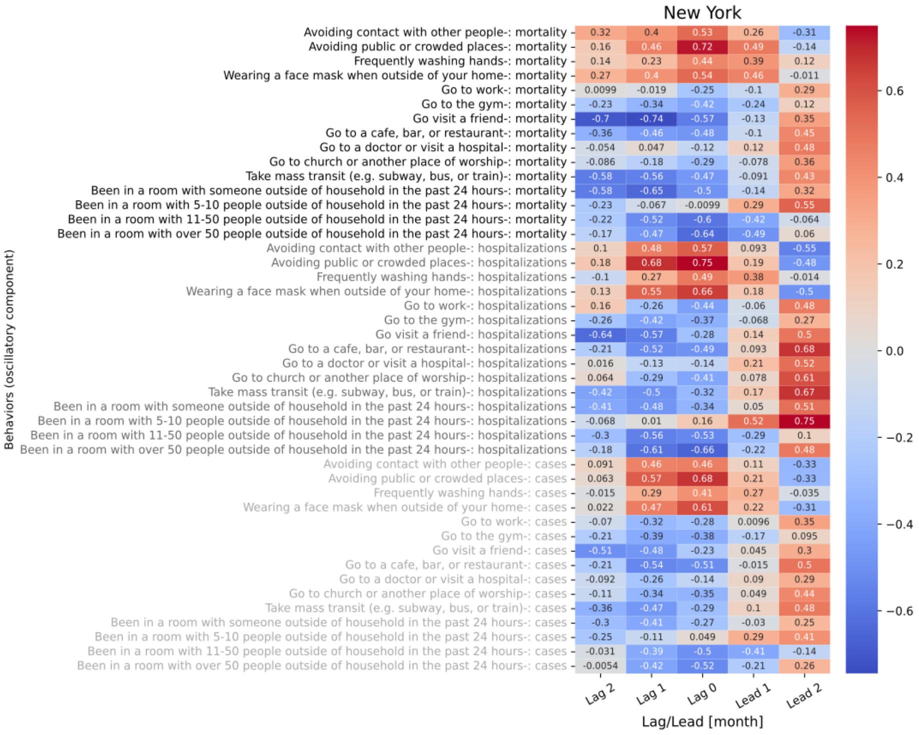

**Figure S9.33 shows the 5 lagged correlations of oscillations in behavior trend for New York with national level mortality (black), hospitalizations (gray), and cases (light gray).**

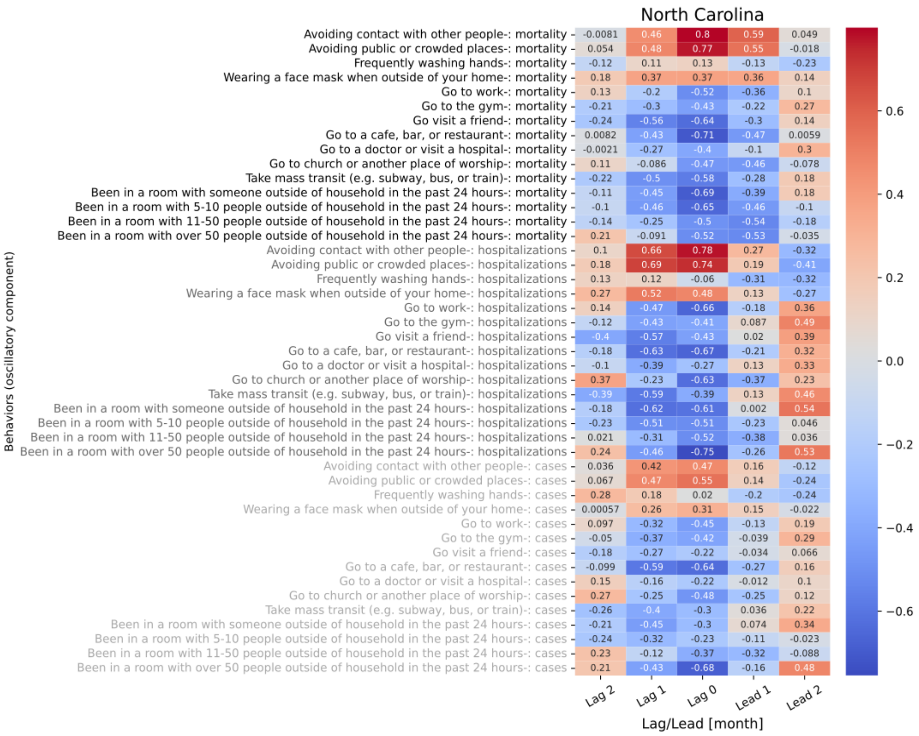

**Figure S9.34 shows the 5 lagged correlations of oscillations in behavior trend for North Carolina with national level mortality (black), hospitalizations (gray), and cases (light gray).**

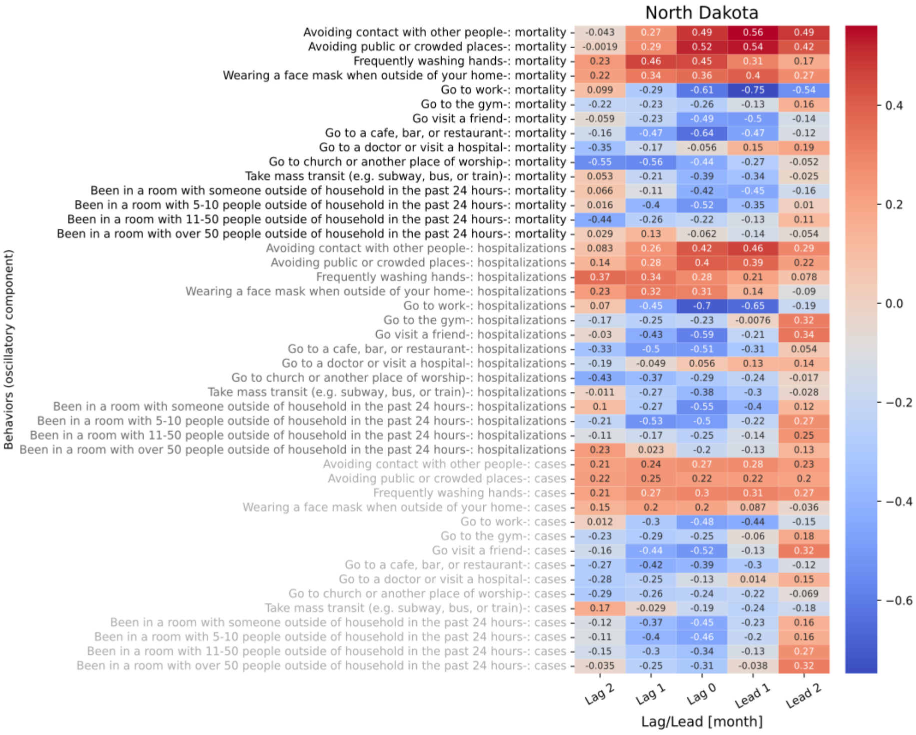

**Figure S9.35 shows the 5 lagged correlations of oscillations in behavior trend for North Dakota with national level mortality (black), hospitalizations (gray), and cases (light gray).**

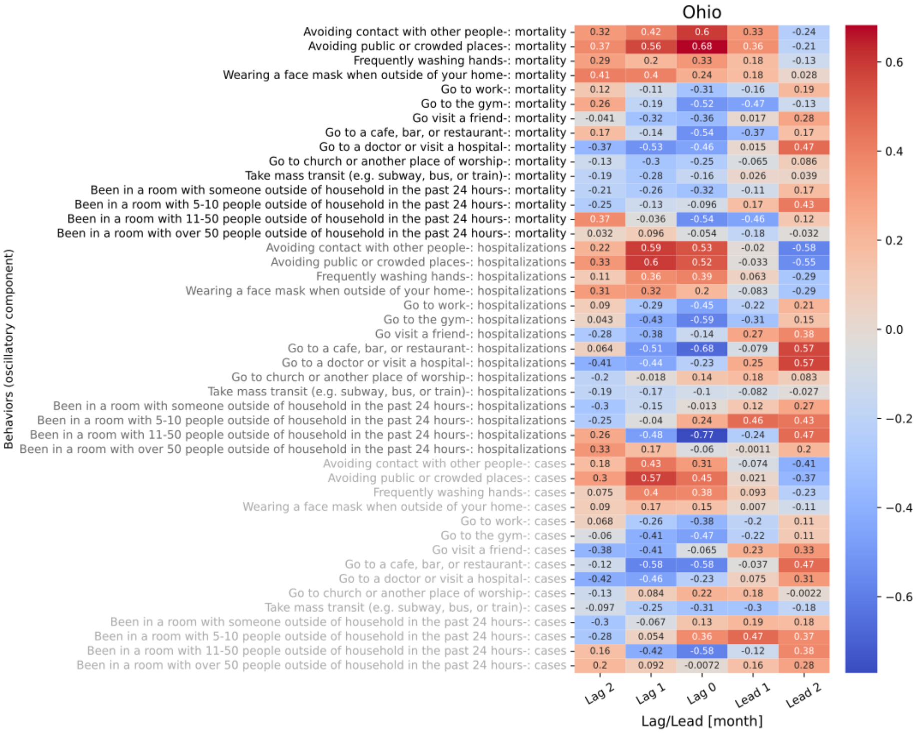

**Figure S9.36 shows the 5 lagged correlations of oscillations in behavior trend for Ohio with national level mortality (black), hospitalizations (gray), and cases (light gray).**

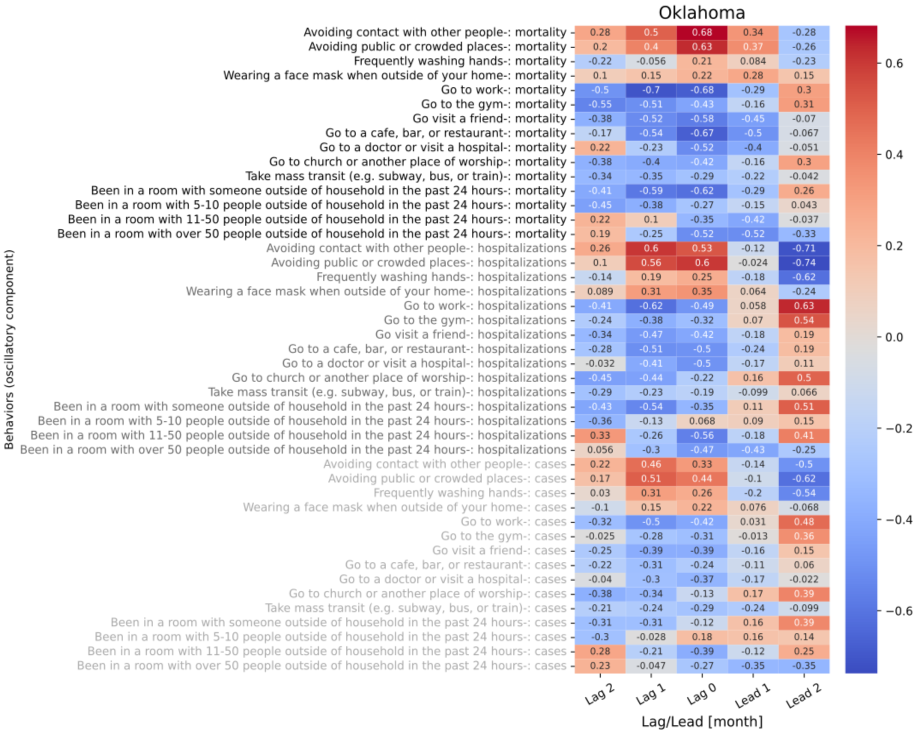

Figure S9.37 shows the 5 lagged correlations of oscillations in behavior trend for Oklahoma with national level mortality (black), hospitalizations (gray), and cases (light gray).

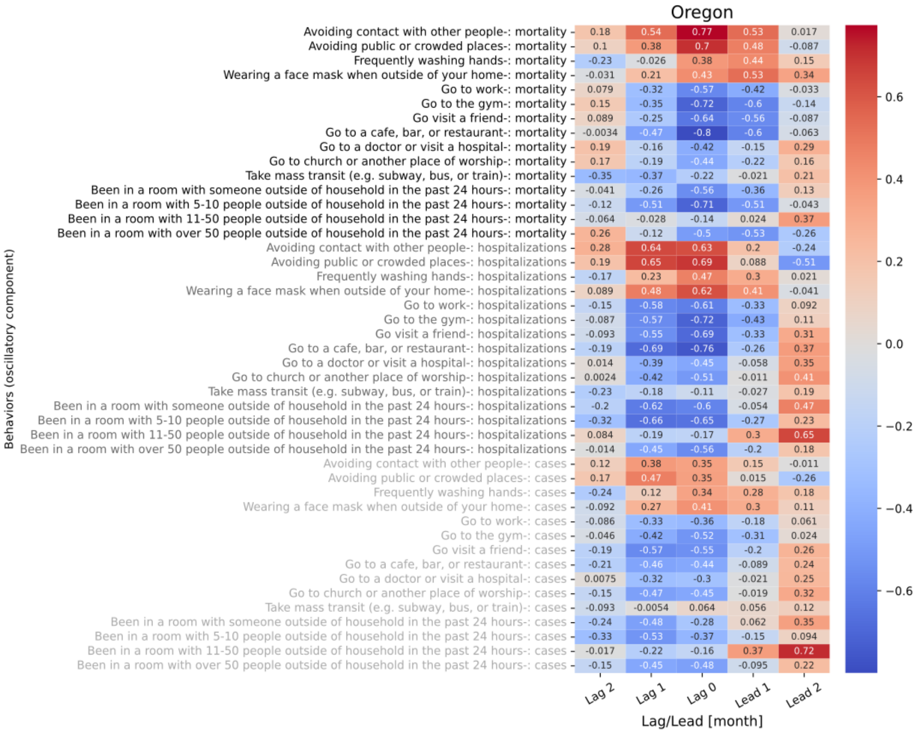

Figure S9.38 shows the 5 lagged correlations of oscillations in behavior trend for Oregon with national level mortality (black), hospitalizations (gray), and cases (light gray).

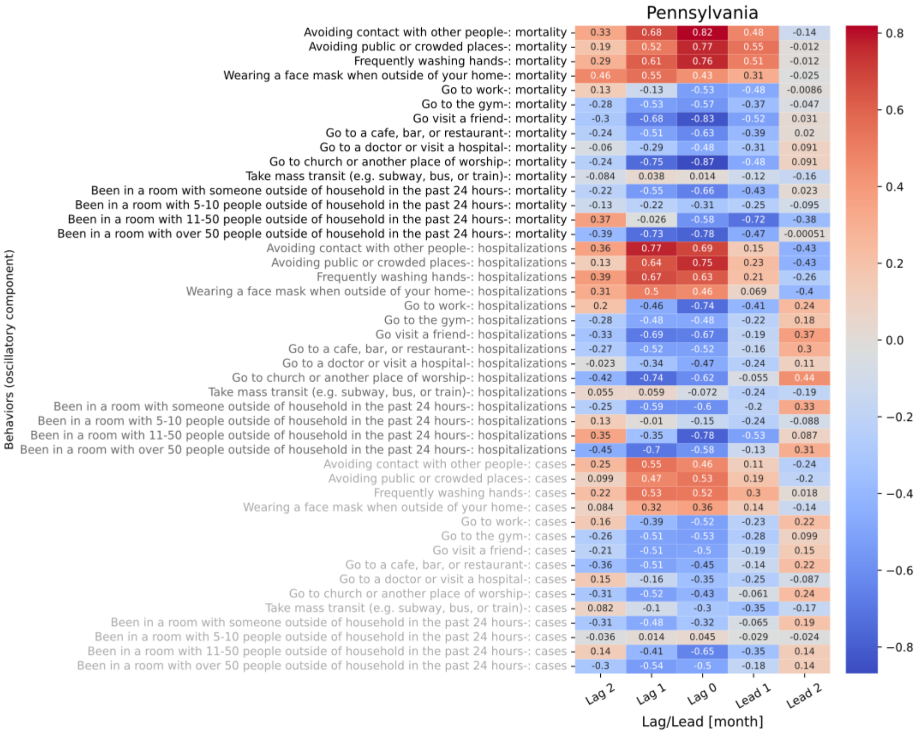

**Figure S9.39 shows the 5 lagged correlations of oscillations in behavior trend for Pennsylvania with national level mortality (black), hospitalizations (gray), and cases (light gray).**

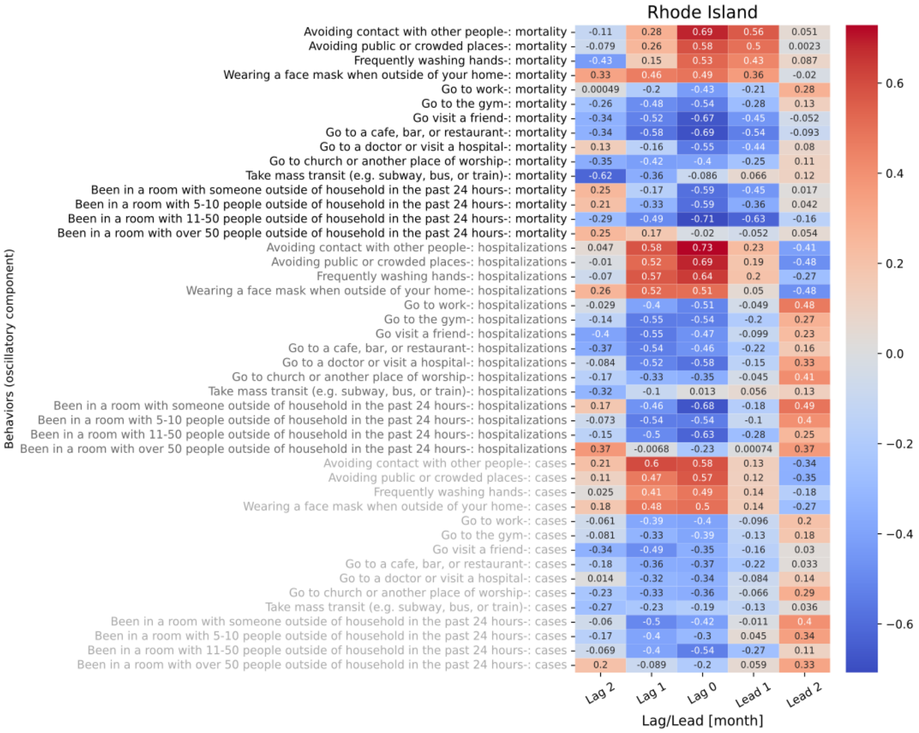

**Figure S9.40 shows the 5 lagged correlations of oscillations in behavior trend for Rhode Island with national level mortality (black), hospitalizations (gray), and cases (light gray).**

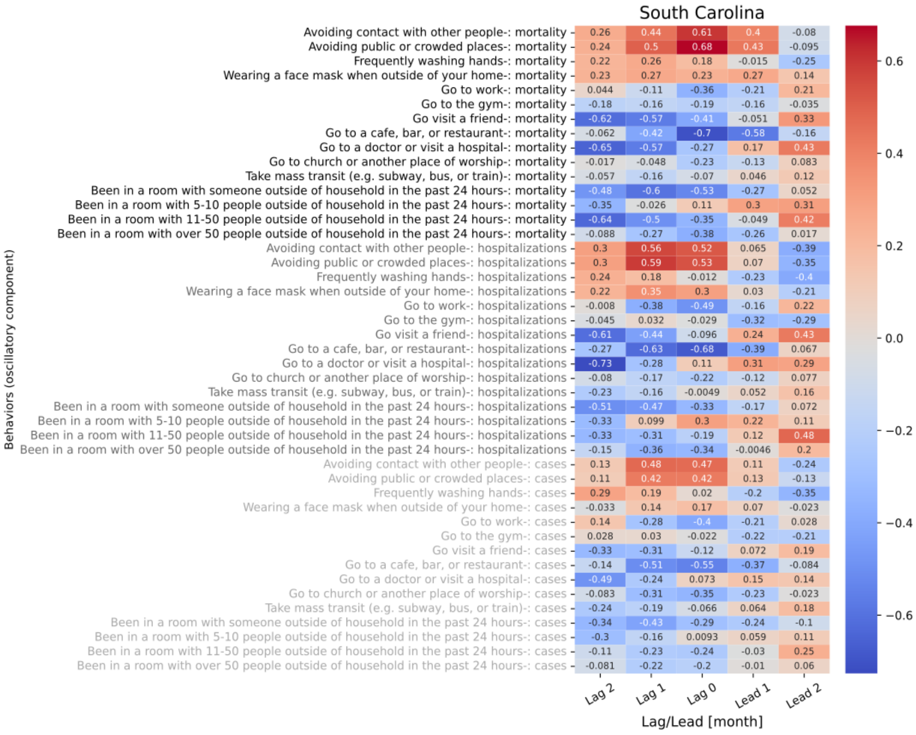

**Figure S9.41 shows the 5 lagged correlations of oscillations in behavior trend for South Carolina with national level mortality (black), hospitalizations (gray), and cases (light gray).**

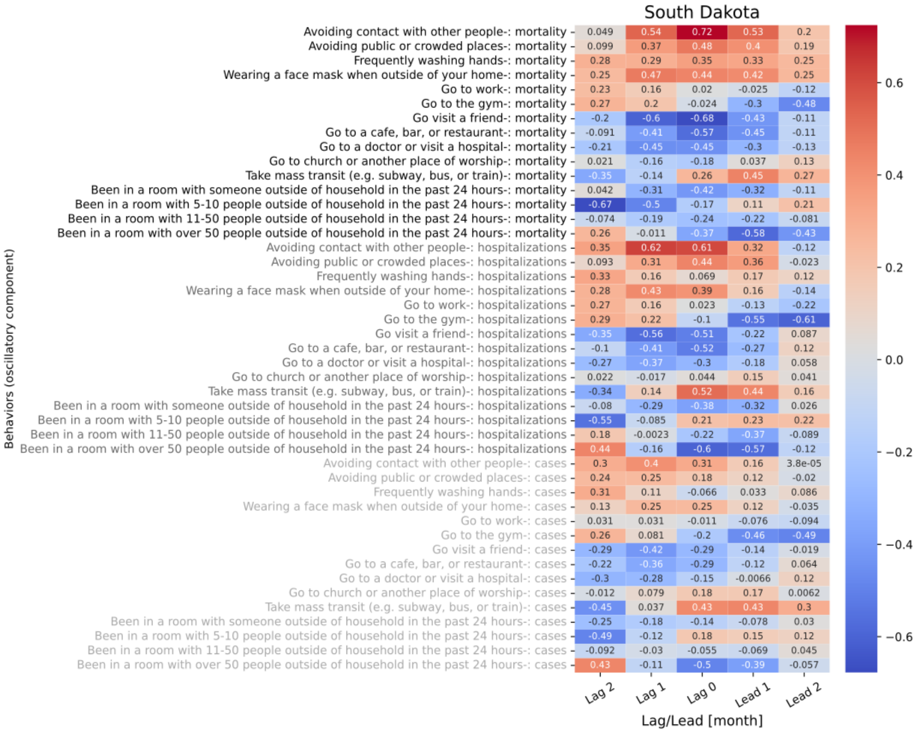

**Figure S9.42 shows the 5 lagged correlations of oscillations in behavior trend for South Dakota with national level mortality (black), hospitalizations (gray), and cases (light gray).**

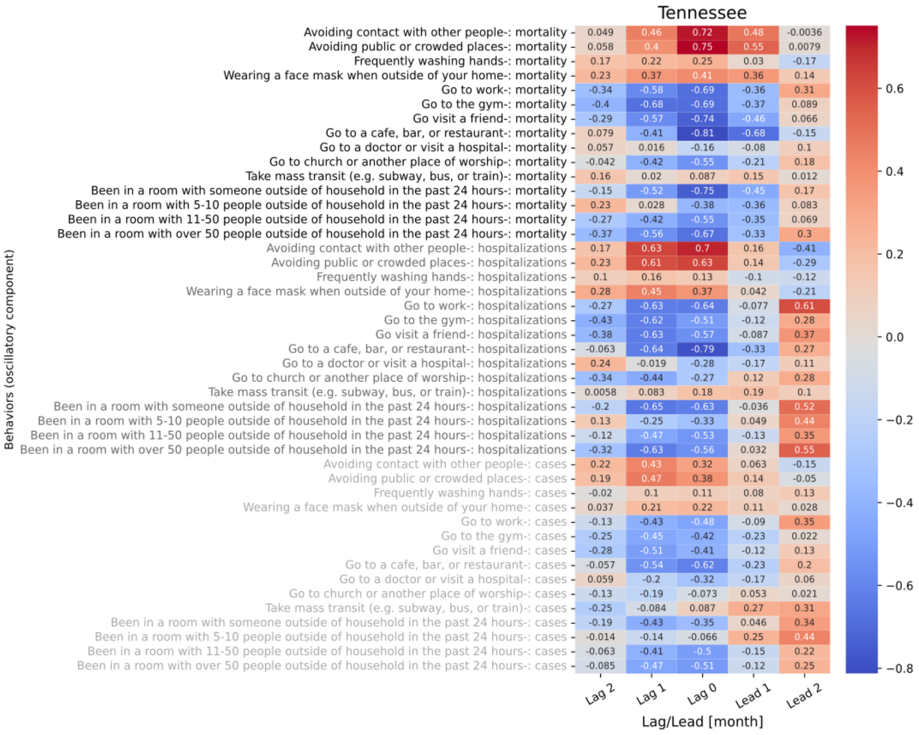

**Figure S9.43 shows the 5 lagged correlations of oscillations in behavior trend for Tennessee with national level mortality (black), hospitalizations (gray), and cases (light gray).**

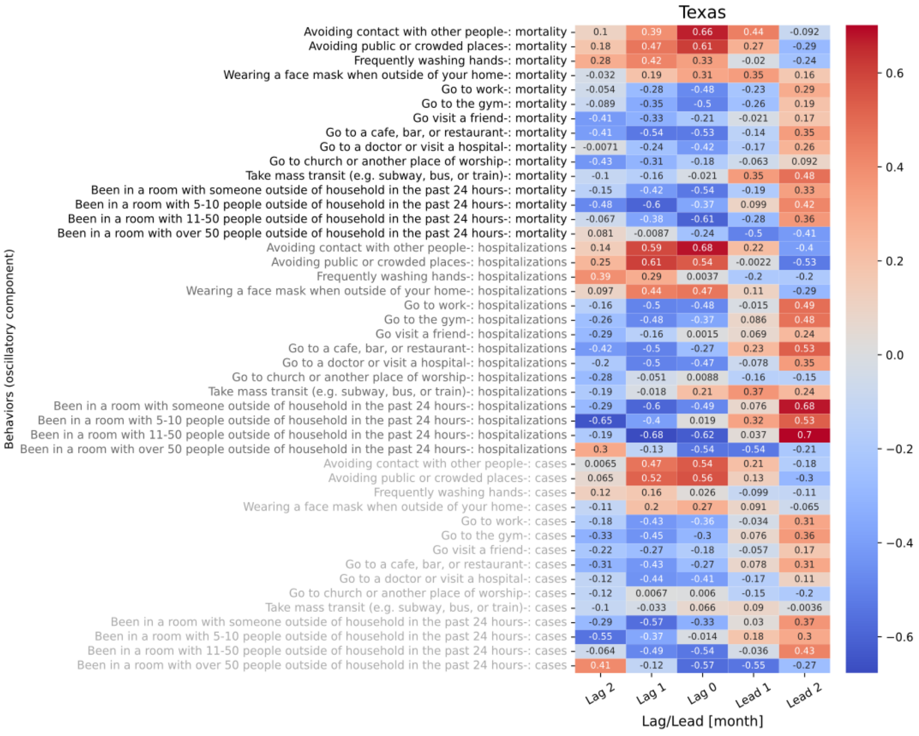

**Figure S9.44 shows the 5 lagged correlations of oscillations in behavior trend for Texas with national level mortality (black), hospitalizations (gray), and cases (light gray).**

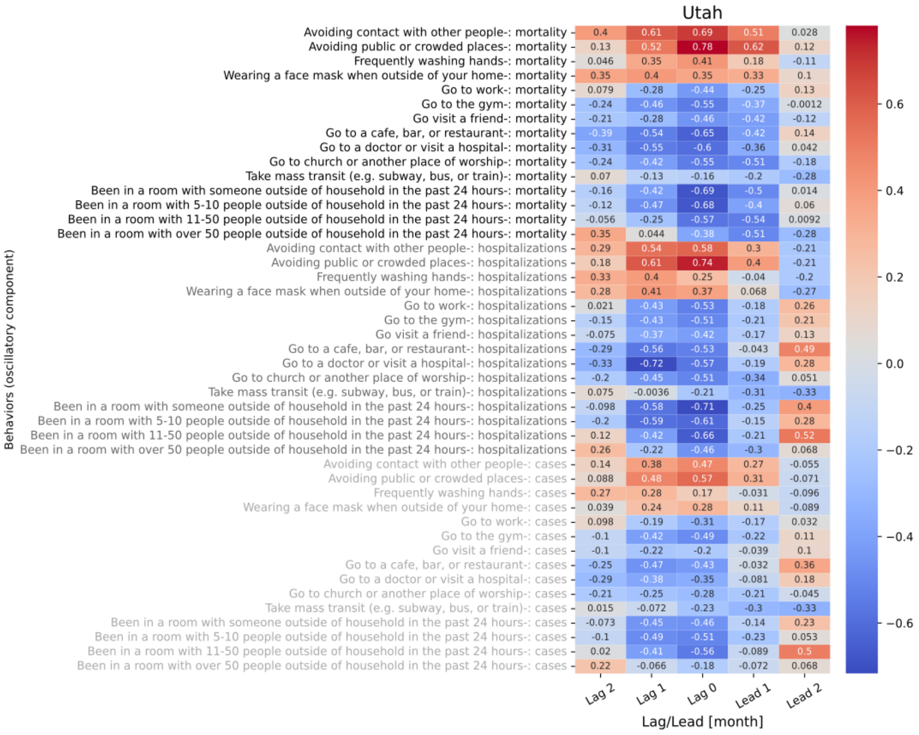

**Figure S9.45 shows the 5 lagged correlations of oscillations in behavior trend for Utah with national level mortality (black), hospitalizations (gray), and cases (light gray).**

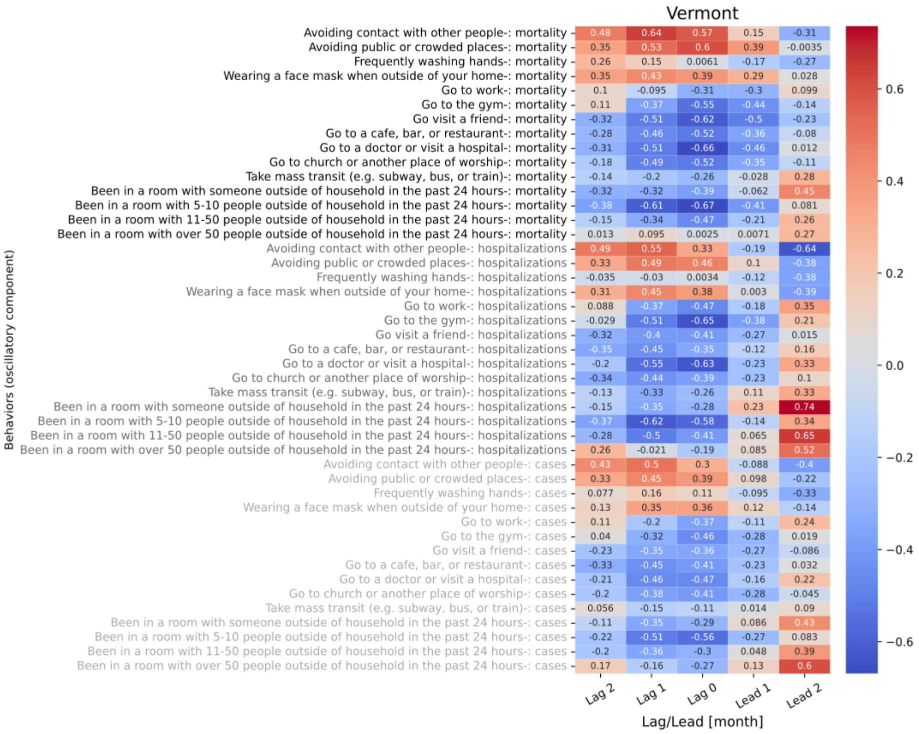

**Figure S9.46 shows the 5 lagged correlations of oscillations in behavior trend for Vermont with national level mortality (black), hospitalizations (gray), and cases (light gray).**

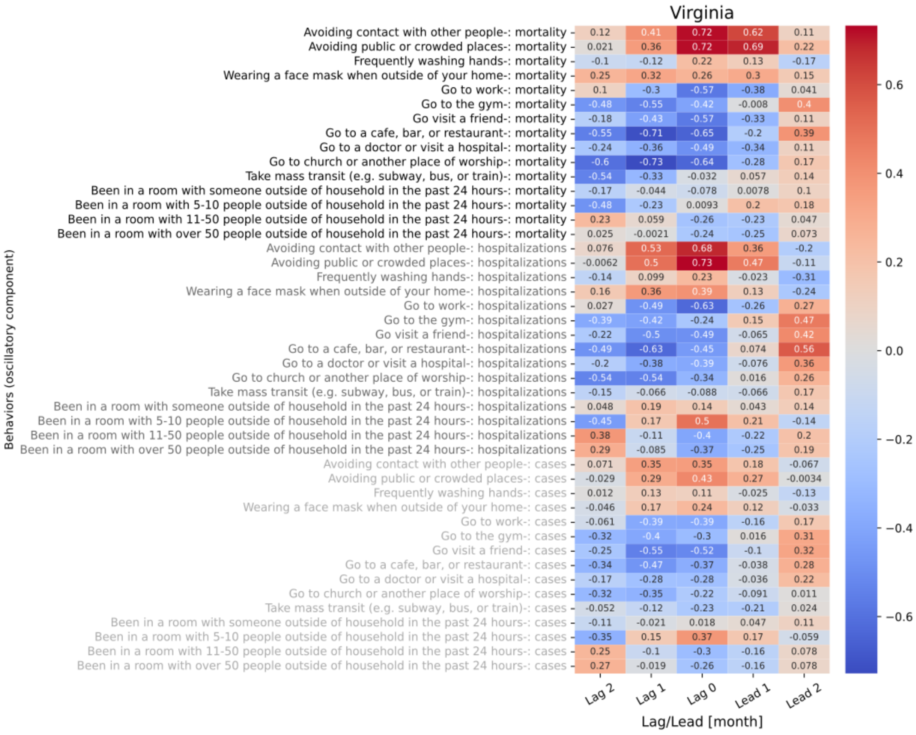

Figure S9.47 shows the 5 lagged correlations of oscillations in behavior trend for Virginia with national level mortality (black), hospitalizations (gray), and cases (light gray).

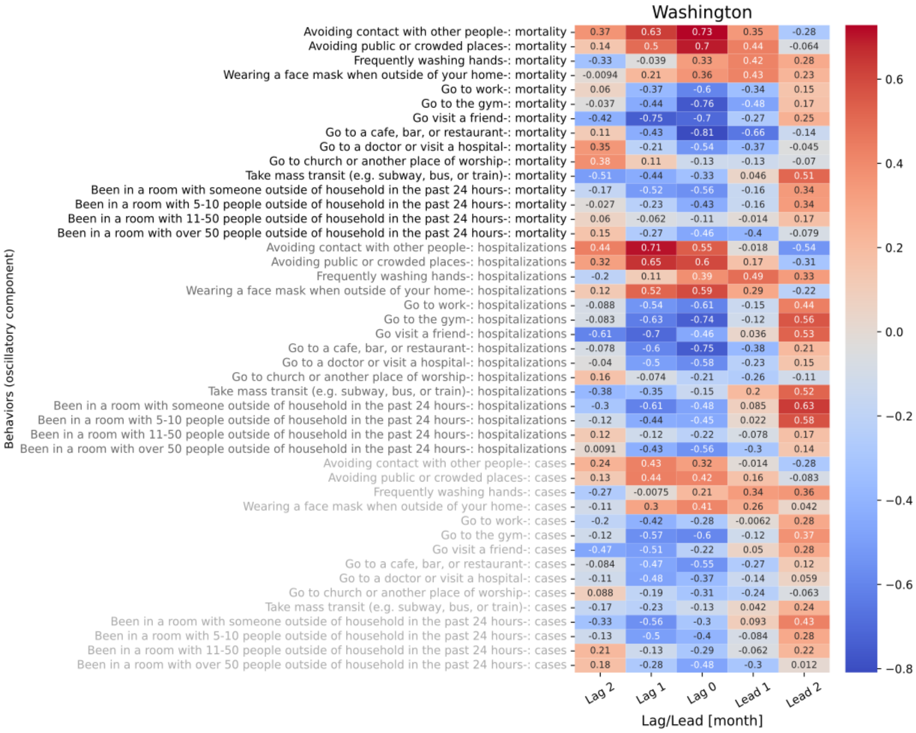

Figure S9.48 shows the 5 lagged correlations of oscillations in behavior trend for Washington with national level mortality (black), hospitalizations (gray), and cases (light gray).

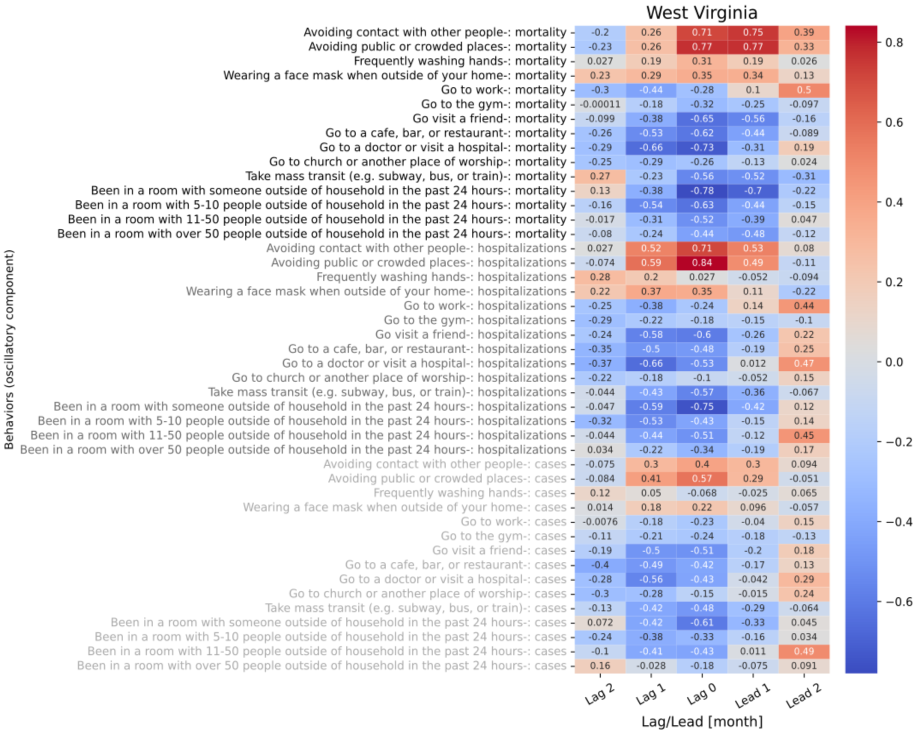

**Figure S9.49 shows the 5 lagged correlations of oscillations in behavior trend for West Virginia with national level mortality (black), hospitalizations (gray), and cases (light gray).**

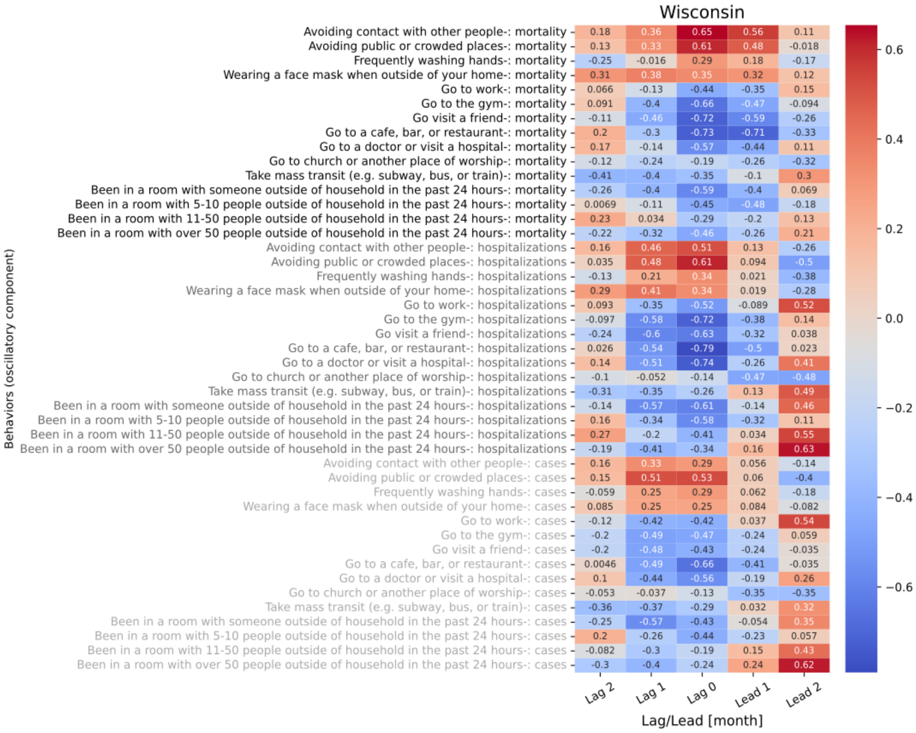

**Figure S9.50 shows the 5 lagged correlations of oscillations in behavior trend for Wisconsin with national level mortality (black), hospitalizations (gray), and cases (light gray).**

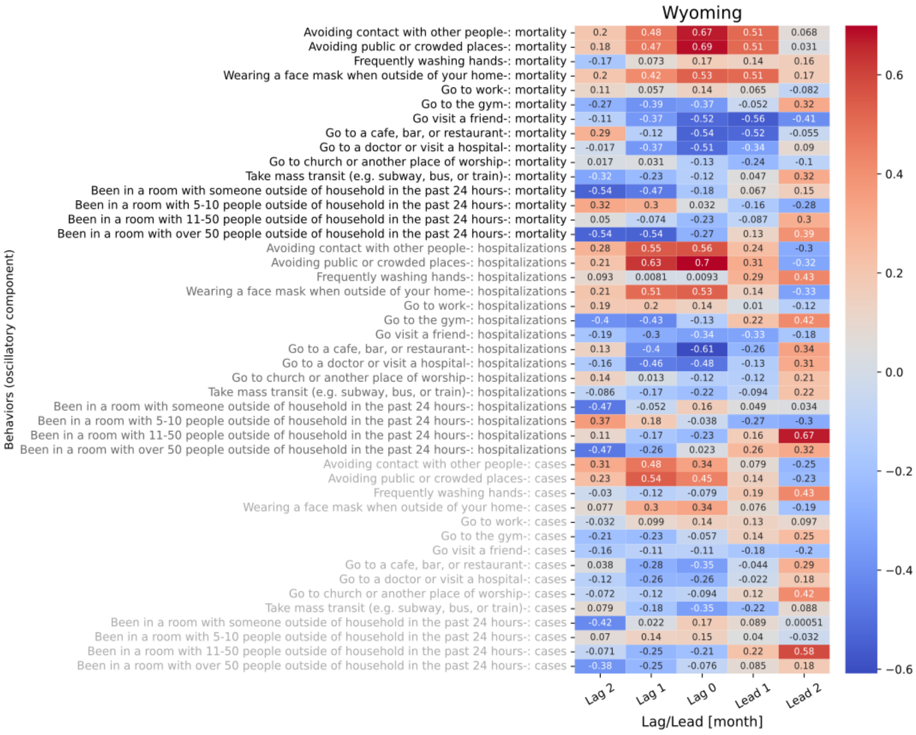

**Figure S9.51 shows the 5 lagged correlations of oscillations in behavior trend for Wyoming with national level mortality (black), hospitalizations (gray), and cases (light gray).**

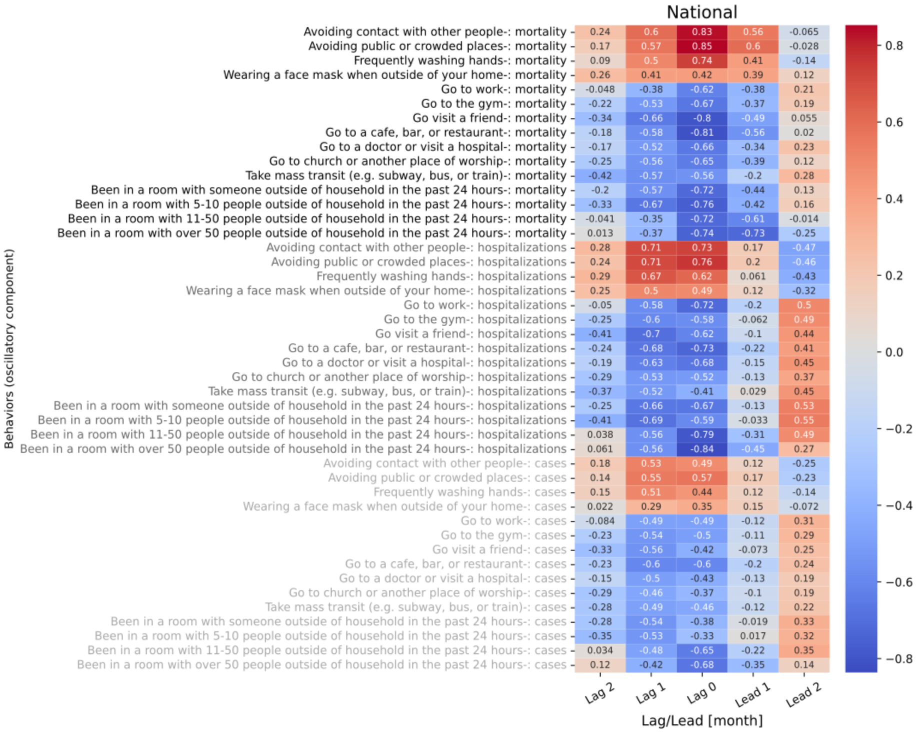

**Figure S9.52 shows the 5 lagged correlations of oscillations in behavior trend for National with national level mortality (black), hospitalizations (gray), and cases (light gray).**
